# How does methamphetamine affect the brain? A systematic review of magnetic resonance imaging studies

**DOI:** 10.1101/2025.02.11.25322099

**Authors:** GXH Liu, M Tayebi, B Bristow, J Wang, Y Lin, G Newburn, P Condron, P McHugh, W Schierding, S Holdsworth, M Scadeng

## Abstract

Methamphetamine (METH) is an addictive psychostimulant that is associated with significant physical and psychological harm. Magnetic resonance imaging (MRI) is a non-invasive and powerful imaging modality that can reveal structural and functional brain changes. We conducted a systematic review to identify and appraise the existing literature examining brain MRI changes associated with METH exposure. A comprehensive literature search was conducted on PubMed, Web of Science, and Scopus. Any study that used MRI to evaluate brain changes in people who used or had been exposed to METH using MRI were included. Data were collected on study design, participant demographics, METH use parameters, MRI acquisition details, and key findings. Quality appraisal was conducted using an *a priori* quality appraisal tool. One hundred and thirty seven studies with 8313 participants were included in our review. Nearly all were conducted in adults/adolescents (121/137, 88%), and most studies were cross-sectional (107/137, 78%). Task-based functional MRI was the most common modality used (48/137, 35%), followed by structural MRI (38/137, 28%). Quality appraisal scores ranged from 40 – 100% (mean 88%). Overall, METH is associated with numerous adverse changes in brain structure and function, including reduced grey matter volume and thickness in frontal and limbic regions and decreased fractional anisotropy in various white matter structures, with associated socio-emotional dysregulation, impaired decision-making and learning processes, and cognitive deficits. Targeted, higher-order, or multi-modal MRI sequences may be useful in further clarifying the impact of METH on the brain and evaluating the use of potential therapeutic agents.

## Introduction

Methamphetamine (METH) is a synthetic psychostimulant that targets the central nervous system and is known to induce intense euphoria in users.(Moszczynska, 2021; Yang et al., 2018) Due to its highly addictive properties and overdose risks, METH use has become increasingly recognised in recent times as a global health concern, and is often coupled alongside the ongoing opioid use epidemic.(Fogger, 2019; Moszczynska, 2021; Paulus and Stewart, 2020) The 2023 World Drug Report published by the United Nations Office on Drugs and Crime reported that the current METH market remains centred within North America and East and South-East Asia, although identified signs of increasing METH production and use in the Middle East and South Asia.(United Nations Office on Drugs and Crime, 2023)

Chronic METH use has been linked to a range of adverse physical and psychological symptoms, including dental problems, weight loss, insomnia, cardiopulmonary disease, cognitive impairment, mood disorders, and increased risk of psychosis.(Ma et al., 2018; McKetin et al., 2008; Salo et al., 2011; Sulaiman et al., 2014) METH use may also predispose towards adverse behavioural outcomes, including violence perpetration and victimisation, limited educational and vocational trajectories, increased involvement with the justice system, and risky sexual behaviour,(Foulds et al., 2020; Guerin et al., 2023; Hittner, 2016; Marshall and Werb, 2010; McKetin et al., 2014; Plüddemann et al., 2008) and often manifests alongside other substance-use disorders and mental health issues which can complicate the delivery of timely and effective care.(Moszczynska, 2021) Due to its harmful effects on the central nervous system, METH has been extensively studied using neuroimaging techniques. In particular, magnetic resonance imaging (MRI) is a non-invasive, non-ionising, and flexible imaging approach that can provide insight into the structural, functional, and neurochemical brain patterns associated with active, short- and long-term abstinent, and treated METH use.(Chen et al., 2019; Garrison and Potenza, 2014; Nierenberg, 2018) Common MRI modalities used to study the effects of METH include structural MRI, diffusion MRI (dMRI), and functional MRI (fMRI), which in turn comprises both resting-state (rs-fMRI) and task-based (task-fMRI) modalities.

Structural MRI (particularly T1-weighted high-resolution scans) has been critical to investigating macrostructural changes in total brain volume, white and grey matter structures, and cortical thickness associated with METH use. In particular, voxel-based morphometry(Suckling and Nestor, 2017) is widely used to evaluate brain volume changes and has demonstrated reduced grey matter volume in numerous brain regions in adult users of METH (MAA), including the bilateral insular cortices, bilateral superior temporal and inferior frontal gyri and left striatal structures.(Hall et al., 2015) In contrast, dMRI can yield detailed information about the microstructural architecture of white matter tracts(Le Bihan, 2014; Le Bihan et al., 1986) by tracking the random Brownian motion of water molecules within tissues, and may reveal areas of tissue disruption and neuronal degeneration.(Soares et al., 2013; White et al., 2008) Previous studies have suggested that METH use is associated with global white matter alterations, including in the frontal lobe,(Lederer et al., 2016) corpus callosum,(Kim et al., 2009) and cingulum,(Ottino-González et al., 2022) and that these changes may be related to compromised fibre and/or myelin integrity.

Alongside anatomical data, MRI can also be used to explore brain function using fMRI. fMRI operates by capturing rapid dynamic changes in blood flow as an indirect measure of neural activity,(Gore, 2003) and can be used to assess brain function at rest (termed rs-fMRI) and during specific tasks (termed task-fMRI). Tasks commonly employed during task-fMRI in METH studies assess a range of neurocognitive skills and processes relevant to substance use, including attention, inhibitory control, impulsivity, and decision-making.(Sabrini et al., 2019) A recent review of 29 papers reported that METH use was associated with impaired cognitive control and impulsivity and that these deficits correlated with neural dysfunction in the anterior cingulate cortex (ACC), prefrontal cortex (PFC), and striatum.(Sabrini et al., 2019)

Previous reviews have attempted to summarise brain MRI changes associated with METH use or exposure. Chen et al.(Chen et al., 2019) identified 12 papers evaluating patients with METH-associated psychosis (MAP) and reported that MAP-related brain alterations predominantly involved the frontal, striatal, and limbic systems. Sabrini et al.(Sabrini et al., 2019) conducted a systematic review examining the relationship between METH use and cognition. They reported that METH use was associated with reduced metabolic activity and structural abnormalities in the PFC and ACC, which may lead to impaired cognitive control and decision-making. Further, Moghaddam et al.(Moghaddam et al., 2021) found that prenatal METH exposure was associated with significant deficits in the frontal lobe, thalamus, striatum, limbic system, and interconnecting white matter tracts, and that these alterations could be identified across structural, functional, and metabolic MRI sequences.

However, previous reviews have limited their analyses to specific patient subgroups with METH use, and few have conducted quality appraisal or a risk of bias assessment. Given the growing popularity of MRI as an imaging technique to characterise the neural impacts of METH, a systematic review that comprehensively summarises and appraises the existing literature across all patient groups would be invaluable for informing current treatment guidelines and guiding future research.

Thus, this review aims to provide a comprehensive overview of studies to-date that have conducted brain MRI in people that have been exposed to METH. Specific aims include 1) qualitatively summarising the neural impact of METH across structural MRI, dMRI, rs-fMRI, and task-fMRI modalities; 2) appraising the quality of the existing literature; and 3) identifying knowledge gaps and current limitations that warrant further investigation and research.

This review article is structured according to MRI modality (i.e. structural, dMRI, rs-fMRI, and task-fMRI). Findings within each modality are further sub-categorised as deemed most appropriate by the review authors to provide an accurate and comprehensive overview of the literature whilst maintaining overall readability. The subcategories for each modality are as follows: participant population for structural and dMRI (i.e. METH use overall, abstinent METH use, and prenatal exposure to METH), MRI analysis method for rs-fMRI (independent component analysis (ICA), seed-based functional connectivity (FC), effective connectivity (EC), and regional homogeneity (ReHo) and Amplitude of Low-Frequency Fluctuations (ALFF)), and cognitive domain assessed by task for task-fMRI (cue reactivity and conditioning; emotional regulation and moral processing; risk management and reward processing; outcome prediction, feedback processing, and learning; Stroop effect; sensorimotor function; and memory).

## Methods

This systematic review was conducted according to the 2020 Preferred Reporting Items for Systematic Reviews and Meta-Analyses (PRISMA) guidelines.(Page et al., 2021) This study was not prospectively registered and a protocol was not prepared.

### Search strategy

A comprehensive literature search was conducted on PubMed, Scopus, and Web of Science on the 11^th^ January 2022 using controlled vocabulary and keywords. In brief, search terms were clustered into five broad categories (METH, MRI, structural MRI, dMRI, and fMRI) and refined using Boolean operators (AND, OR, NOT). Four independent searches were then conducted on each database using a combination of search terms: METH and MRI, METH and structural MRI, METH and dMRI, and METH and fMRI.

No restrictions were placed on publication date or study design. The authors also manually searched the reference lists of relevant articles to identify additional studies that fulfilled the inclusion criteria. De-duplication was conducted according on EndNote using the methods detailed in Bramer et al.(Bramer et al., 2016) Further details on the search strategy are provided in **Supplementary File 1**.

### Study criteria

Included studies comprised those that quantitatively analysed brain changes, as detected by MRI, in people who have been exposed to METH. This included (but was not limited to) previous or current MAA, children or infants who had been prenatally exposed to METH, and healthy volunteers who were administered METH as part of a research study. Studies that examined multiple drugs of interest were included if METH-specific data were reported. Studies published only as conference abstracts, book chapters, reviews, case reports, non-English articles, or animal studies were excluded.

### Study screening

Abstracts were screened by one author (GXHL) and all potentially relevant full texts were independently screened by two authors (GXHL, MT). Discrepancies were settled by discussion with the review team.

### Data extraction

Data extraction was conducted by two authors (GXHL, BB) using a standardised proforma on Microsoft Excel (see **Supplementary File 2**). Information was collected on overall study characteristics and design, participant demographics, clinical parameters of METH and other drug use, MRI acquisition parameters, and a summary of key findings.

### Quality appraisal

All included studies were assessed using an independently designed quality appraisal tool to identify methodological flaws (see **Supplementary File 3**) – this was adapted *a priori* from one used in a review examining dMRI changes following sport-related concussion,(Lees et al., 2021) which was based on the National Institutes of Health Quality Assessment Tool for Observational and Cohort and Cross-Sectional Articles.(National Heart Lung and Blood Institute, 2013) In brief, modifications were made to tailor the tool towards studies examining METH exposure and to generalise it to include all study types and MRI imaging modalities. Key changes included: 1) increased emphasis on clinical parameters of METH and other drug use (e.g. duration of abstinence and use of METH, consideration of polydrug use); 2) focused assessment of confounders adjusted for in statistical analysis; and 3) expansion of MRI acquisition assessment criteria to accommodate for all imaging modalities.

To accommodate for different study types and avoid inappropriate punition, specific questions were classified as “N/A” if they were not applicable for pre-specified study designs or aims (e.g. cross-sectional studies were not penalised for not including follow-up), and final scores were calculated as a percentage of the maximum number of points that could have been scored. The final quality appraisal tool included 11 questions that examined study population and design, drug use parameters, MRI acquisition methods, and approach to statistical analysis, and up to 18 points could be scored.

Quality appraisal was conducted by four authors (GXHL, JW, MT, BB), with each paper being appraised independently by at least two authors. Final scores were decided on by consensus agreements, and discrepancies were resolved by discussion and/or consultation with a third author if necessary.

### Data analysis

Study findings are discussed and summarised qualitatively in the review text, tables, and figures. Although the authors had considered conducting a quantitative meta-analysis, this was ultimately not performed due to significant heterogeneity between study populations, METH and other drug use parameters, and analysis methods, which may have confounded or biased the final analysis findings.

## Results

Overall, our search returned 646 results, of which 402 were removed during de-duplication, and 107 were excluded during abstract and full-text screening. This left 136 records for inclusion in our review. However, one record (Kohno et al. 2016)(Kohno et al., 2016) reported two studies within a single record, and so 137 studies with 8313 participants were included in this review. The PRISMA flow diagram is available in **Figure 1**.

**Figure 1:**
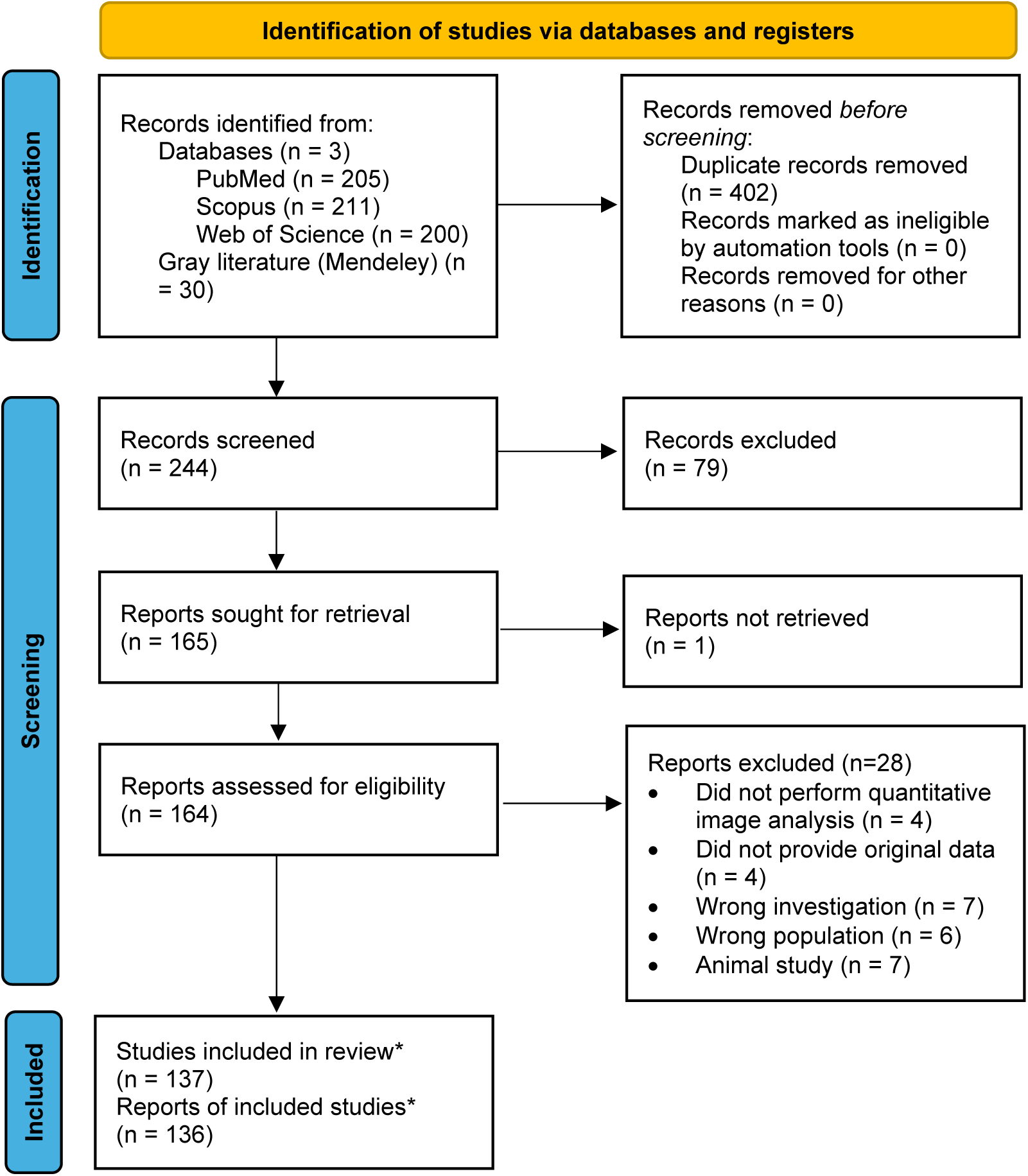
PRISMA flowchart *One study (Kohno et al. 2016)(Kohno et al., 2016) reported 2 studies within 1 record. Therefore, 137 studies with 136 records were included in this review.

Of the 137 studies, nearly all were conducted in adults or adolescents (122/137, 89%); from these, most (116/122, 95%) evaluated previous or current MAA, with the remaining six studies (5%) being done in healthy volunteers that were administered METH as part of the study. 15 (11%) studies assessed children or infants who had been prenatally exposed to METH.

Most studies were cross-sectional analyses without follow-up (100/137, 77%); 14 (10%) studies were observational cohort studies, and 13 (9%) were two-arm randomised controlled trials (RCTs). The most common imaging modality used was task-fMRI (48/137, 35%) followed by structural MRI (38/137, 28%), with 24 studies (18%) using rs-fMRI and 19 (13%) using dMRI. Further details on participant demographics, study design, and MRI modality are available in **Tables 1a-d** and **Supplementary Table 1**.

Overall, 136 (99%) studies underwent quality appraisal; one study (Grodin et al. 2019)(Grodin et al., 2019) could not be assessed as insufficient information was provided in the full-text report. The mean total score was 87.9%, with scores ranging from 40-100% (**Tables 1a-d**). Nearly one-third of studies (39/135, 29%) scored 100%; of these, all compared MAA with historical or ongoing METH exposure against healthy controls (CON), and over one-half (22/39, 56%) used structural MRI. Further details about the absolute number of points scored and the maximum possible score for each study are provided in **Supplementary Table 2**.

### Structural MRI

Structural MRI provides insight into the anatomical form and layout of neural structures.(Symms et al., 2004) More specifically, information can be gathered on whole-brain and tissue-specific volume, density, thickness, and areas of hyperintensity. With regards to METH use, structural MRI studies have identified alterations in a range of cortical grey matter regions and subcortical grey and white matter structures, particularly in frontal, temporal, and limbic regions (**Tables 2a and 2b**, **Figure 2a and 2b**).

**Figure 2a:**
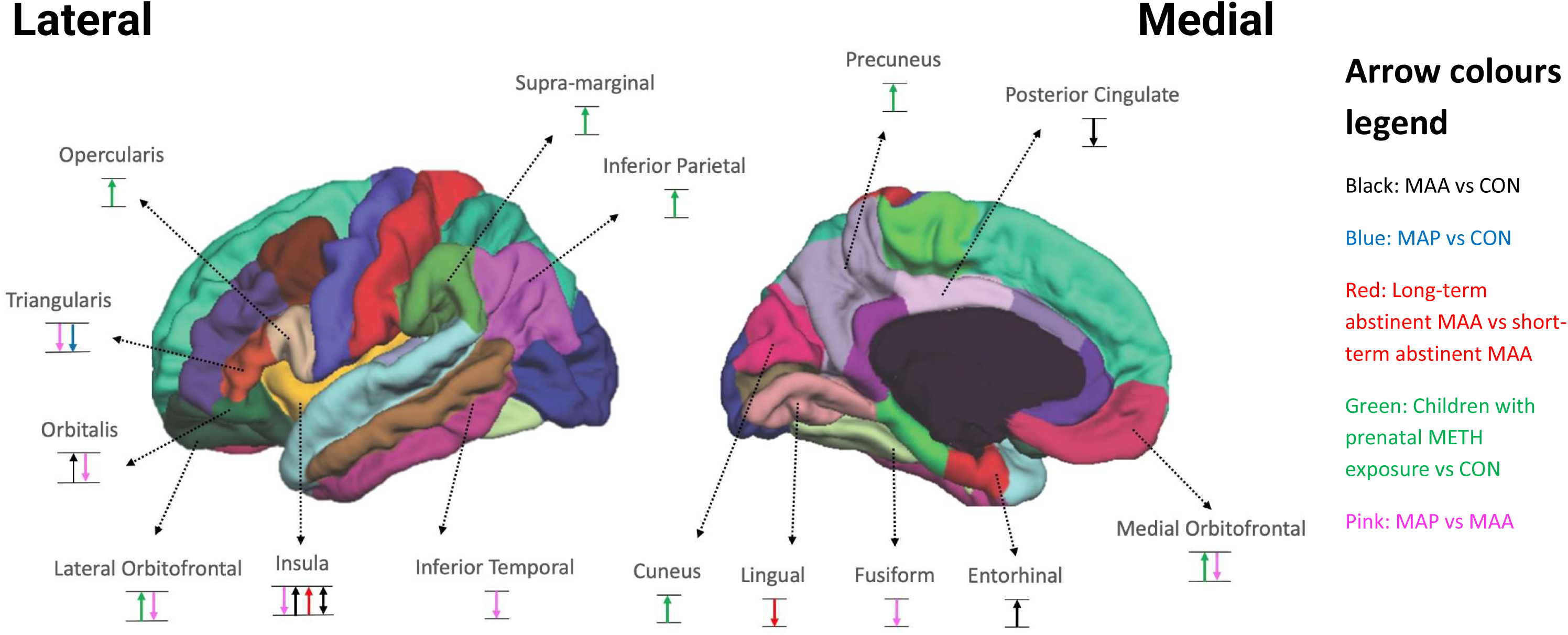
Volumetric changes in cortical structures associated with METH exposure Arrows represent increase (↑), decrease (↓), or no difference (↕) in cortical structure volume between study groups within individual studies as per Table 2a. Each arrow represents an individual study. Arrow colours representative of different study populations as according to the figure legend on the right-hand side. MAA: Adult METH user. MAP: METH-associated psychosis. METH: Methamphetamine. CON: Healthy controls.

**Figure 2b:**
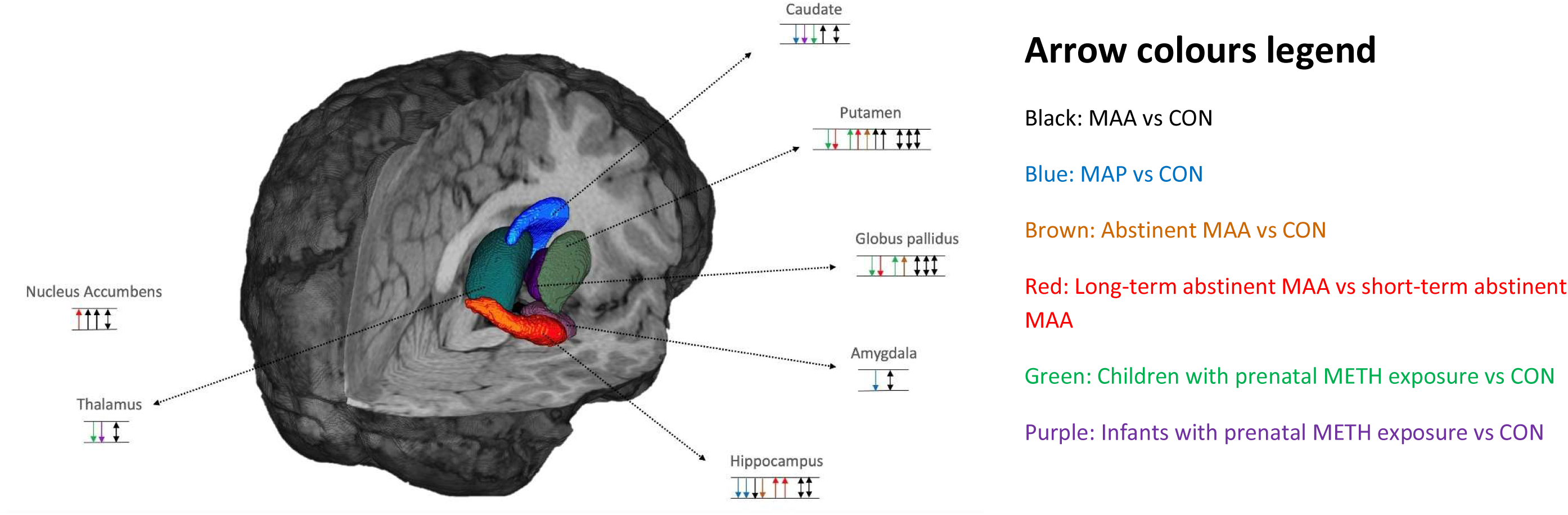
Volumetric changes in subcortical structures associated with METH exposure Arrows represent increase (↑), decrease (↓), or no difference (↕) in subcortical structure volume between study groups within individual studies as per Table 2a. Each arrow represents an individual study. Arrow colours representative of different study populations as according to the figure legend on the right-hand side. MAA: Adult METH user. MAP: METH-associated psychosis. METH: Methamphetamine. CON: Healthy controls.

In our review, 38/137 (28%) papers used structural MRI (**Table 1a**). Both grey and white matter indices were evaluated, including grey and white matter volume and density, grey matter cortical thickness, and white matter hyperintensities.

**Table 1a:**
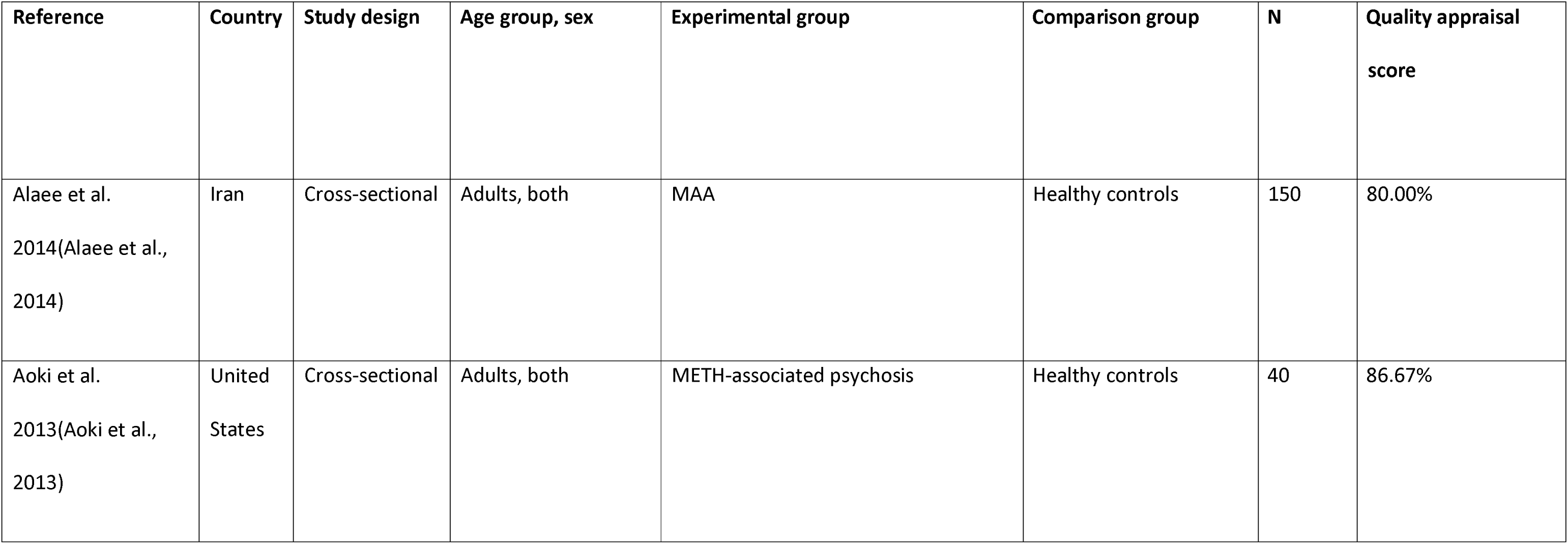
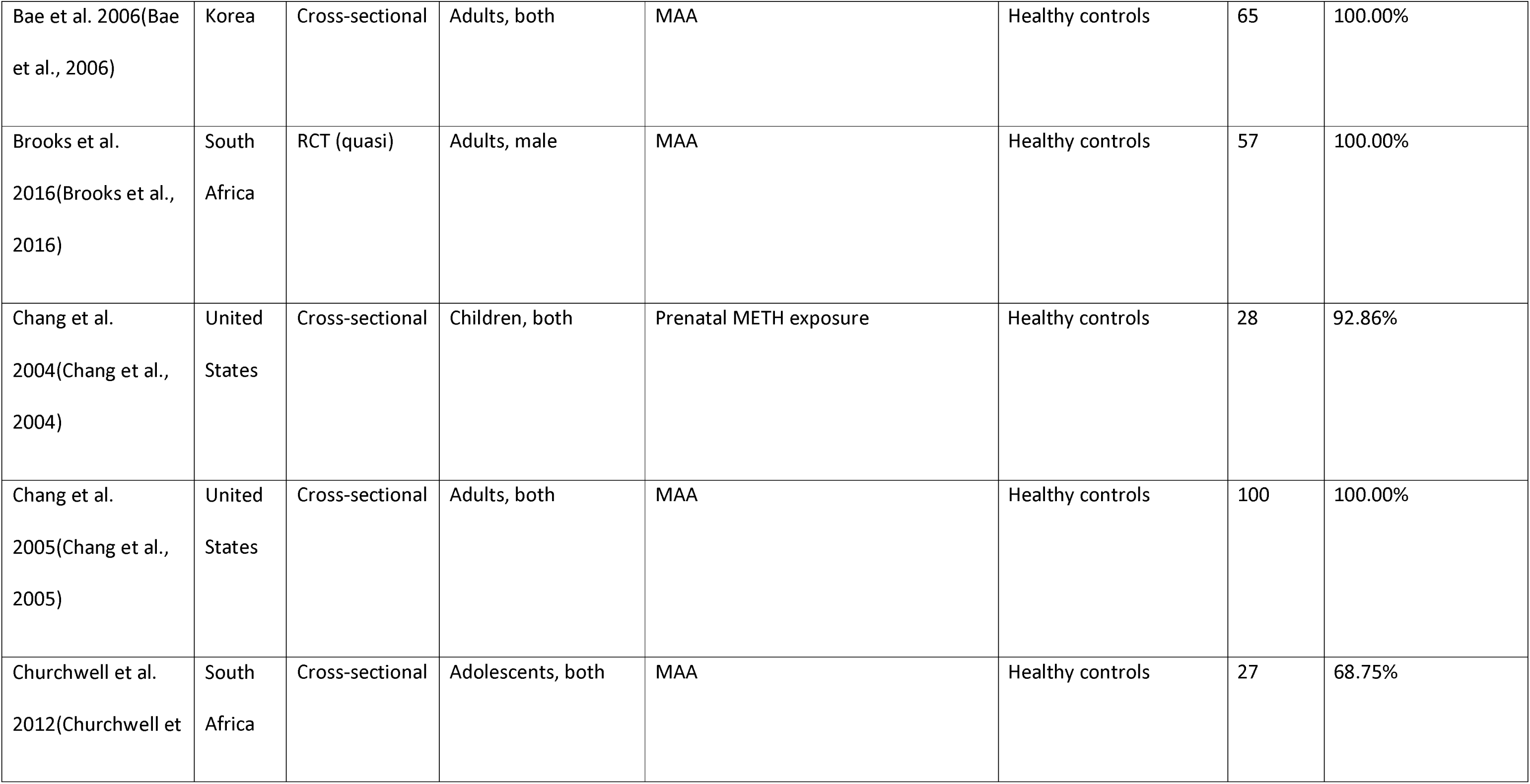
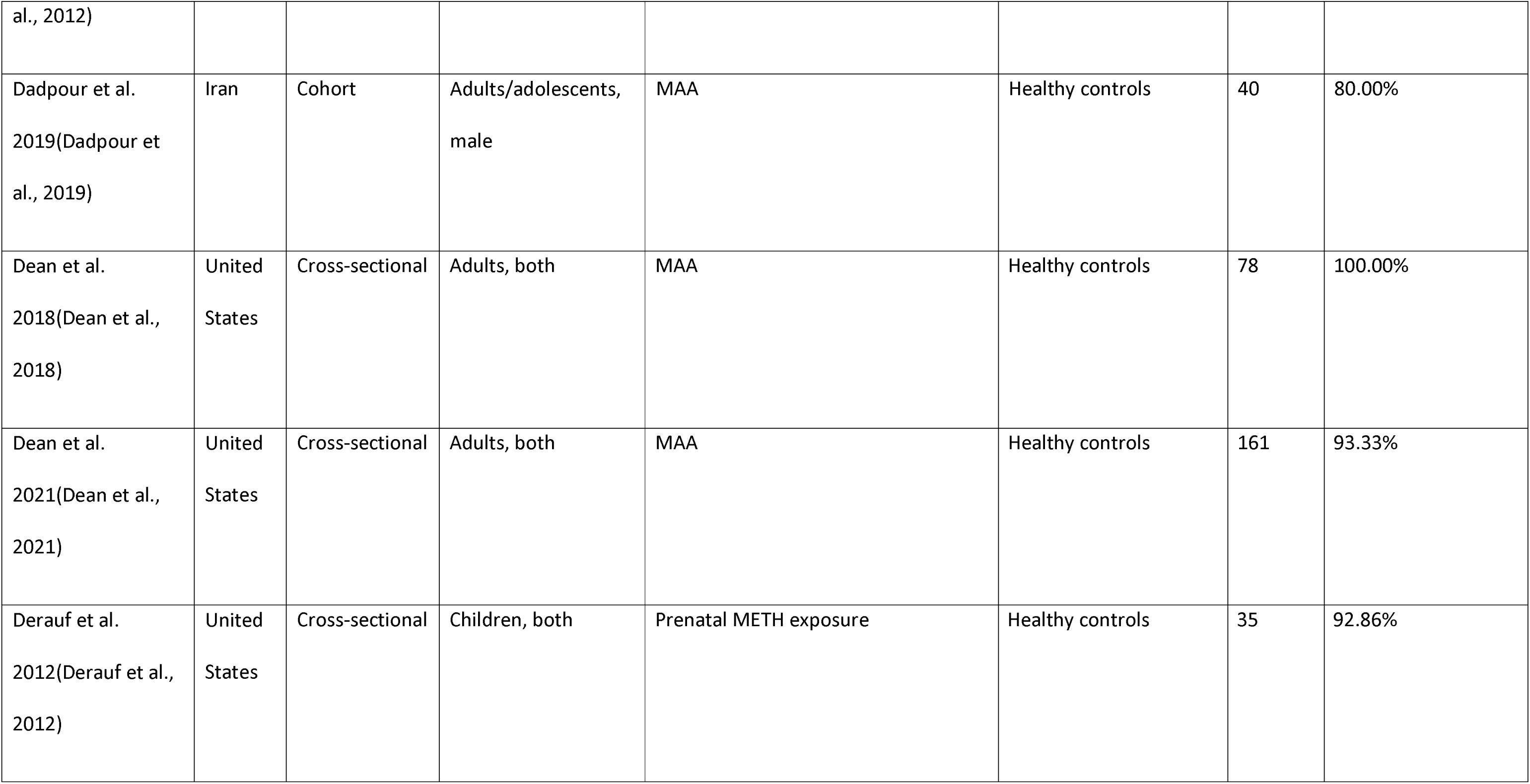
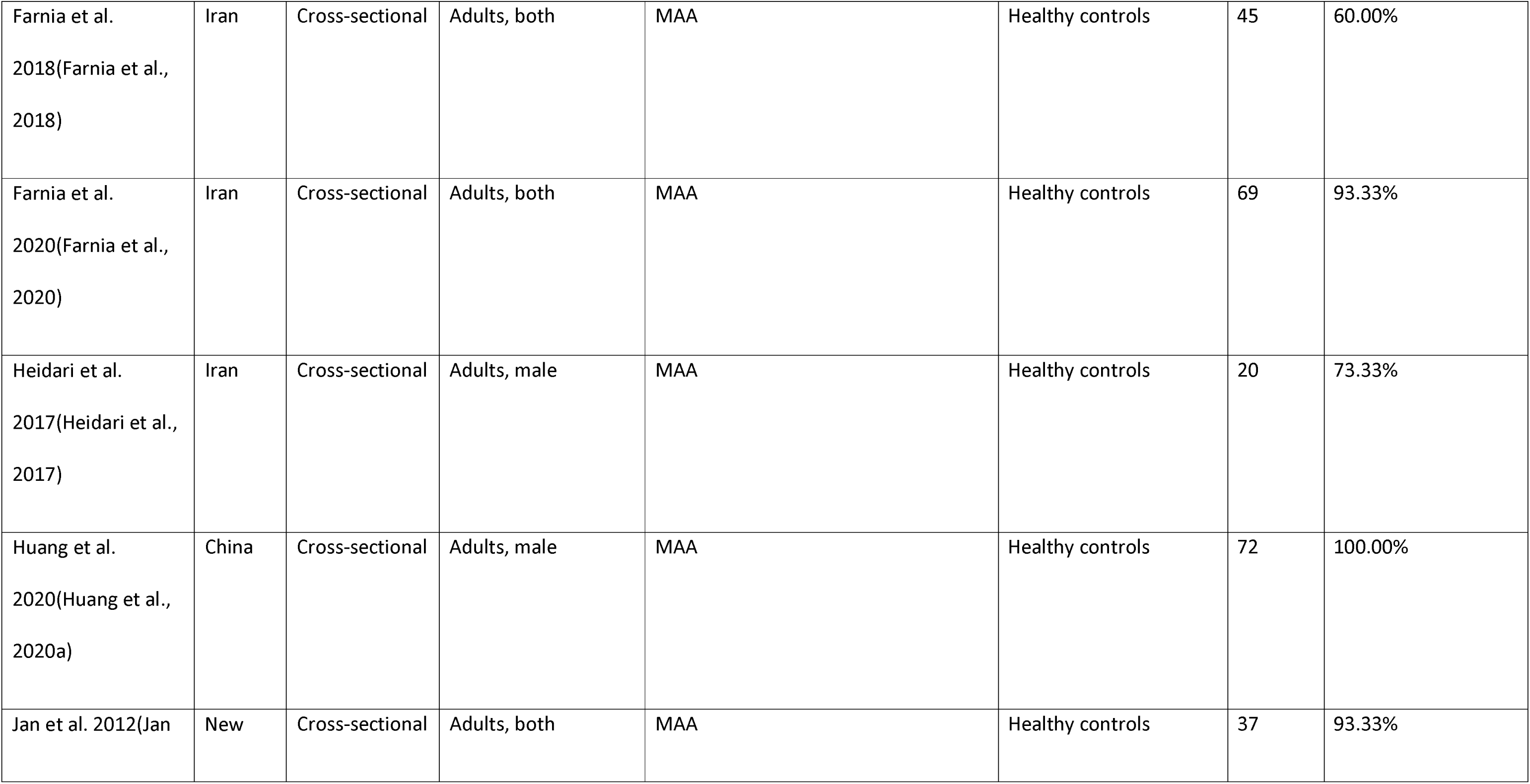
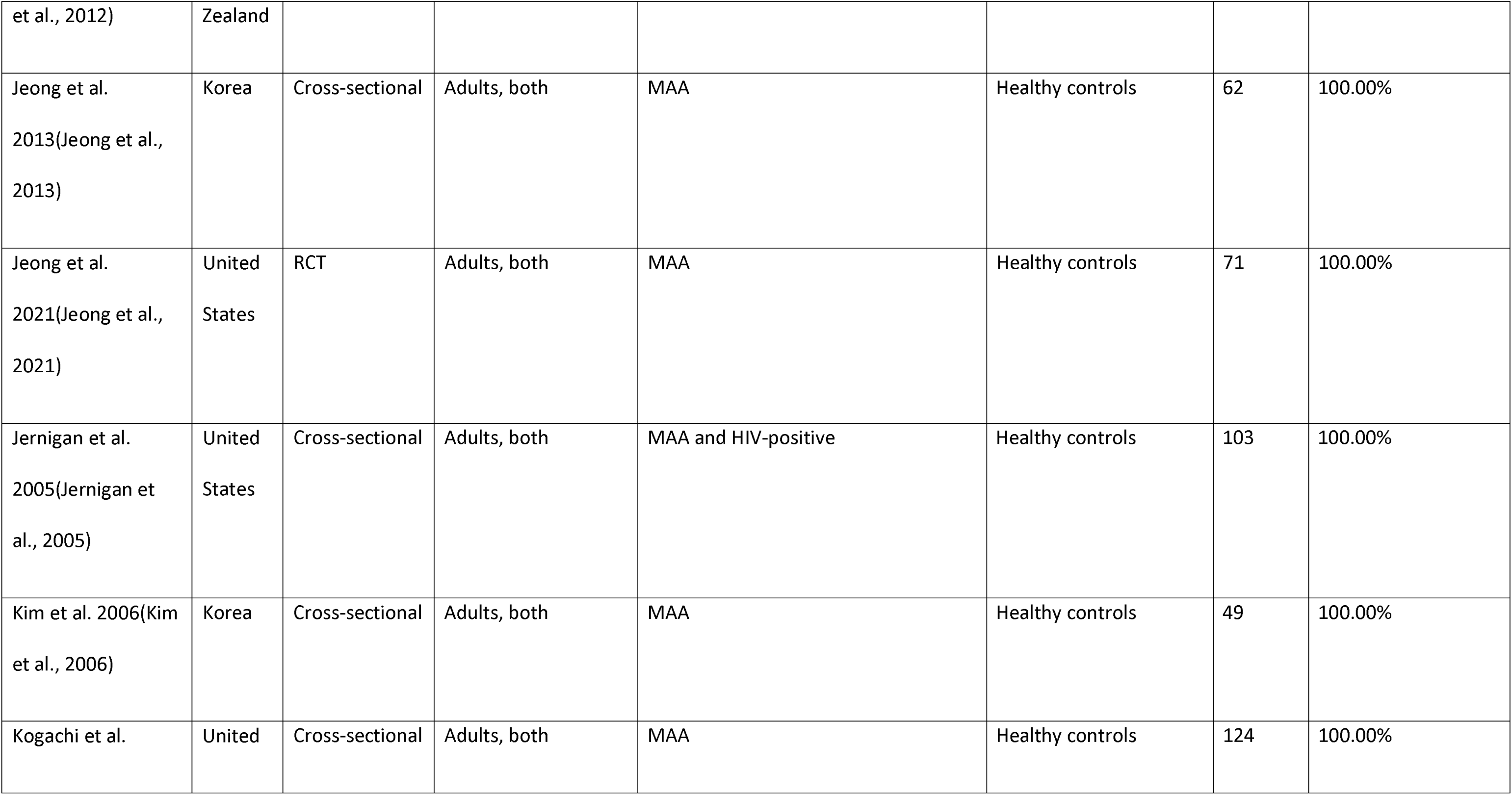
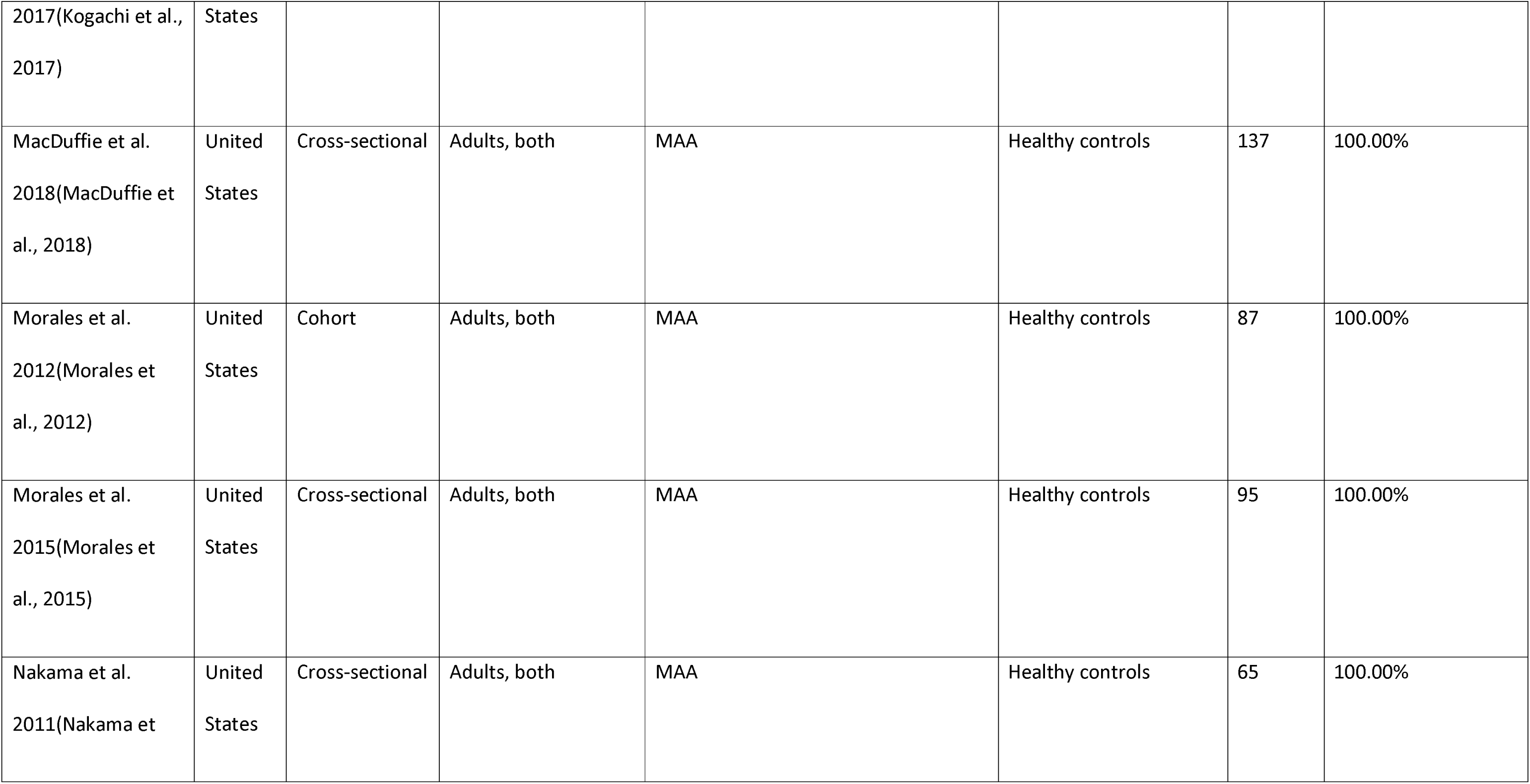
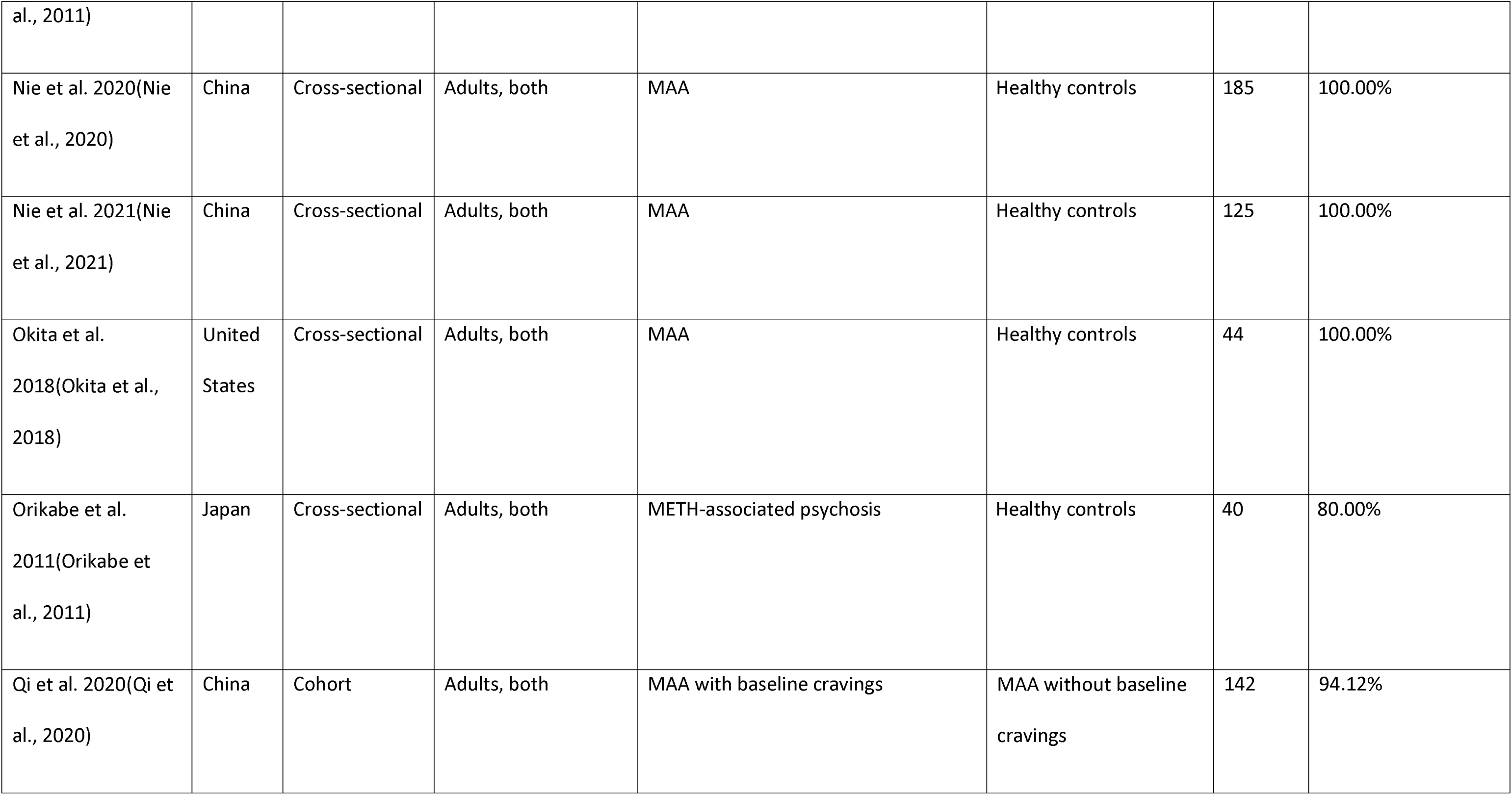
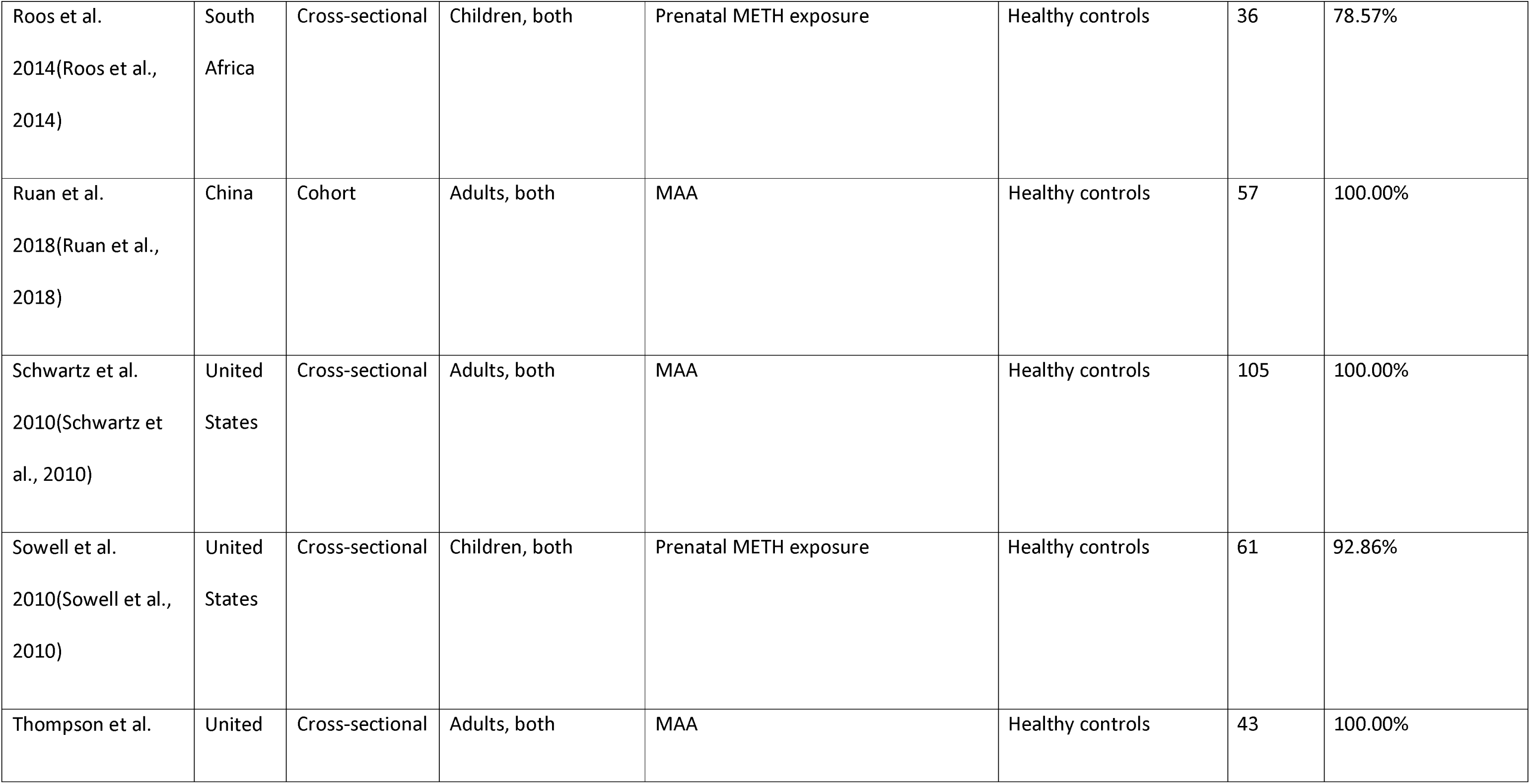
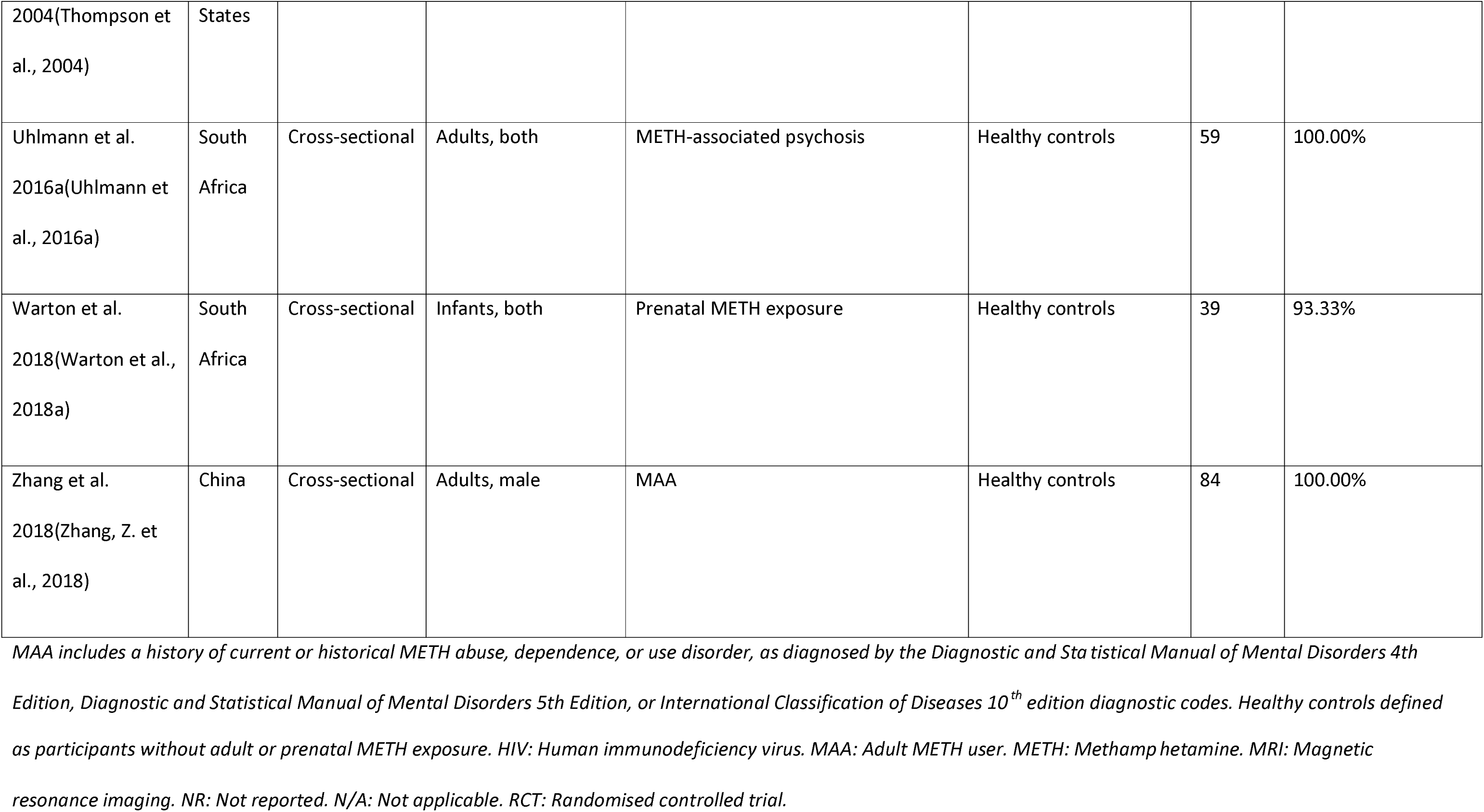
Study demographics and quality appraisal score for structural MRI studies.

| Reference | Country | Study design | Age group, sex | Experimental group | Comparison group | N | Quality appraisal score |
| --- | --- | --- | --- | --- | --- | --- | --- |
| Alaee et al.<br>2014(Alaee et al.,<br>2014) | Iran | Cross-sectional | Adults, both | MAA | Healthy controls | 150 | 80.00% |
| Aoki et al.<br>2013(Aoki et al.,<br>2013) | United States | Cross-sectional | Adults, both | METH-associated psychosis | Healthy controls | 40 | 86.67% |
| Bae et al. 2006(Bae et al., 2006) | Korea | Cross-sectional | Adults, both | MAA | Healthy controls | 65 | 100.00% |
| Brooks et al. 2016(Brooks et al., 2016) | South Africa | RCT (quasi) | Adults, male | MAA | Healthy controls | 57 | 100.00% |
| Chang et al. 2004(Chang et al., 2004) | United States | Cross-sectional | Children, both | Prenatal METH exposure | Healthy controls | 28 | 92.86% |
| Chang et al. 2005(Chang et al., 2005) | United States | Cross-sectional | Adults, both | MAA | Healthy controls | 100 | 100.00% |
| Churchwell et al. 2012(Churchwell et | South Africa | Cross-sectional | Adolescents, both | MAA | Healthy controls | 27 | 68.75% |
| al., 2012) |  |  |  |  |  |  |  |
| Dadpour et al.<br>2019(Dadpour et al., 2019) | Iran | Cohort | Adults/adolescents,<br>male | MAA | Healthy controls | 40 | 80.00% |
| Dean et al.<br>2018(Dean et al., 2018) | United States | Cross-sectional | Adults, both | MAA | Healthy controls | 78 | 100.00% |
| Dean et al.<br>2021(Dean et al., 2021) | United States | Cross-sectional | Adults, both | MAA | Healthy controls | 161 | 93.33% |
| Derauf et al.<br>2012(Derauf et al., 2012) | United States | Cross-sectional | Children, both | Prenatal METH exposure | Healthy controls | 35 | 92.86% |
| Farnia et al.<br>2018(Farnia et al.,<br>2018) | Iran | Cross-sectional | Adults, both | MAA | Healthy controls | 45 | 60.00% |
| Farnia et al.<br>2020(Farnia et al.,<br>2020) | Iran | Cross-sectional | Adults, both | MAA | Healthy controls | 69 | 93.33% |
| Heidari et al.<br>2017(Heidari et al.,<br>2017) | Iran | Cross-sectional | Adults, male | MAA | Healthy controls | 20 | 73.33% |
| Huang et al.<br>2020(Huang et al.,<br>2020a) | China | Cross-sectional | Adults, male | MAA | Healthy controls | 72 | 100.00% |
| Jan et al. 2012(Jan | New | Cross-sectional | Adults, both | MAA | Healthy controls | 37 | 93.33% |
| et al., 2012) | Zealand |  |  |  |  |  |  |
| Jeong et al.<br>2013(Jeong et al.,<br>2013) | Korea | Cross-sectional | Adults, both | MAA | Healthy controls | 62 | 100.00% |
| Jeong et al.<br>2021(Jeong et al.,<br>2021) | United<br>States | RCT | Adults, both | MAA | Healthy controls | 71 | 100.00% |
| Jernigan et al.<br>2005(Jernigan et<br>al., 2005) | United<br>States | Cross-sectional | Adults, both | MAA and HIV-positive | Healthy controls | 103 | 100.00% |
| Kim et al. 2006(Kim<br>et al., 2006) | Korea | Cross-sectional | Adults, both | MAA | Healthy controls | 49 | 100.00% |
| Kogachi et al. | United | Cross-sectional | Adults, both | MAA | Healthy controls | 124 | 100.00% |
| 2017(Kogachi et al., 2017) | States |  |  |  |  |  |  |
| MacDuffie et al. 2018(MacDuffie et al., 2018) | United States | Cross-sectional | Adults, both | MAA | Healthy controls | 137 | 100.00% |
| Morales et al. 2012(Morales et al., 2012) | United States | Cohort | Adults, both | MAA | Healthy controls | 87 | 100.00% |
| Morales et al. 2015(Morales et al., 2015) | United States | Cross-sectional | Adults, both | MAA | Healthy controls | 95 | 100.00% |
| Nakama et al. 2011(Nakama et | United States | Cross-sectional | Adults, both | MAA | Healthy controls | 65 | 100.00% |
| al., 2011) |  |  |  |  |  |  |  |
| Nie et al. 2020(Nie et al., 2020) | China | Cross-sectional | Adults, both | MAA | Healthy controls | 185 | 100.00% |
| Nie et al. 2021(Nie et al., 2021) | China | Cross-sectional | Adults, both | MAA | Healthy controls | 125 | 100.00% |
| Okita et al. 2018(Okita et al., 2018) | United States | Cross-sectional | Adults, both | MAA | Healthy controls | 44 | 100.00% |
| Orikabe et al. 2011(Orikabe et al., 2011) | Japan | Cross-sectional | Adults, both | METH-associated psychosis | Healthy controls | 40 | 80.00% |
| Qi et al. 2020(Qi et al., 2020) | China | Cohort | Adults, both | MAA with baseline cravings | MAA without baseline cravings | 142 | 94.12% |
| Roos et al.<br>2014(Roos et al.,<br>2014) | South<br>Africa | Cross-sectional | Children, both | Prenatal METH exposure | Healthy controls | 36 | 78.57% |
| Ruan et al.<br>2018(Ruan et al.,<br>2018) | China | Cohort | Adults, both | MAA | Healthy controls | 57 | 100.00% |
| Schwartz et al.<br>2010(Schwartz et<br>al., 2010) | United<br>States | Cross-sectional | Adults, both | MAA | Healthy controls | 105 | 100.00% |
| Sowell et al.<br>2010(Sowell et al.,<br>2010) | United<br>States | Cross-sectional | Children, both | Prenatal METH exposure | Healthy controls | 61 | 92.86% |
| Thompson et al. | United | Cross-sectional | Adults, both | MAA | Healthy controls | 43 | 100.00% |
| 2004(Thompson et al., 2004) | States |  |  |  |  |  |  |
| Uhlmann et al. 2016a(Uhlmann et al., 2016a) | South Africa | Cross-sectional | Adults, both | METH-associated psychosis | Healthy controls | 59 | 100.00% |
| Warton et al. 2018(Warton et al., 2018a) | South Africa | Cross-sectional | Infants, both | Prenatal METH exposure | Healthy controls | 39 | 93.33% |
| Zhang et al. 2018(Zhang, Z. et al., 2018) | China | Cross-sectional | Adults, male | MAA | Healthy controls | 84 | 100.00% |

*MAA includes a history of current or historical METH abuse, dependence, or use disorder, as diagnosed by the Diagnostic and Statistical Manual of Mental Disorders 4th*
*Edition, Diagnostic and Statistical Manual of Mental Disorders 5th Edition, or International Classification of Diseases 10<sup>th</sup> edition diagnostic codes. Healthy controls defined*

**Table 1b:**
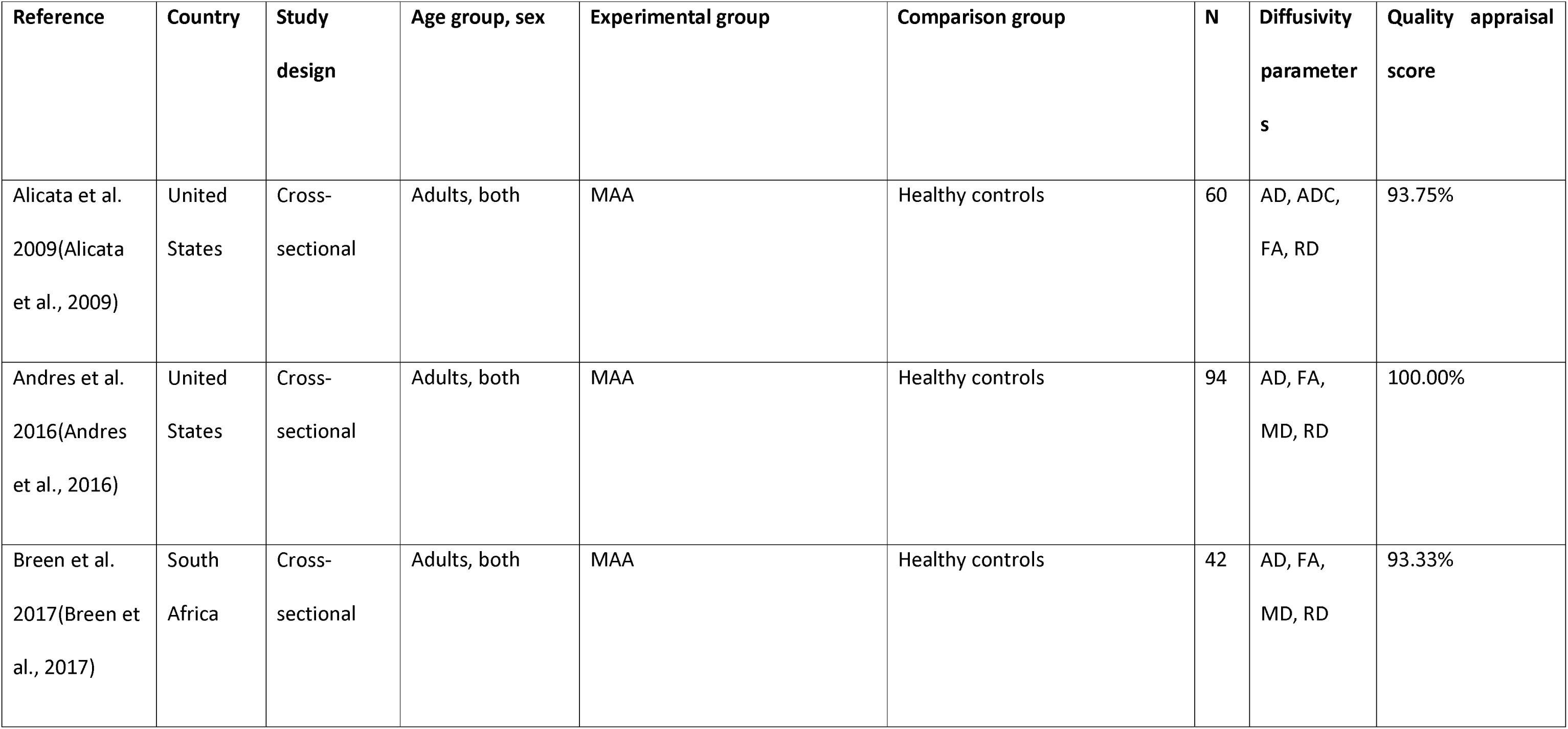
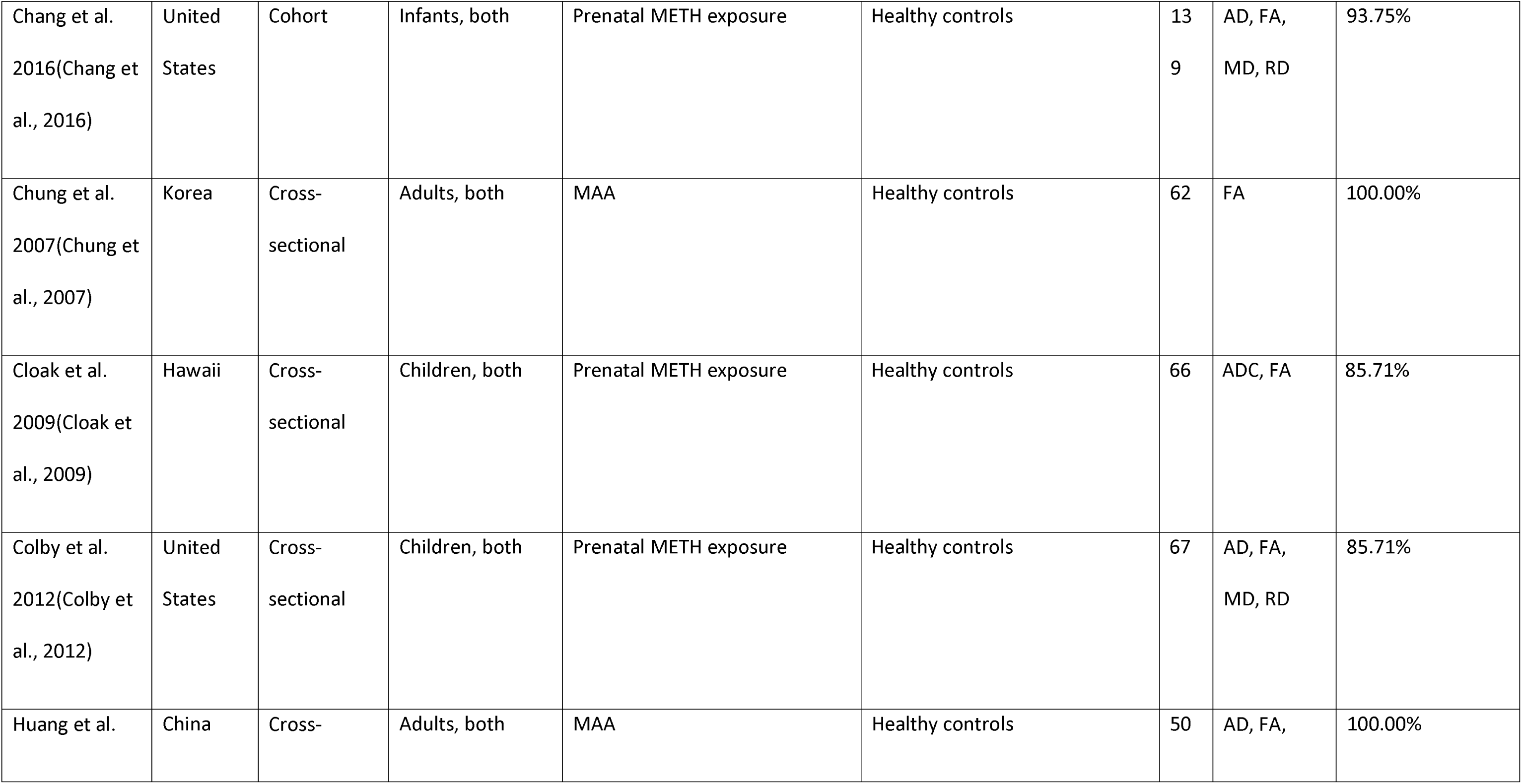
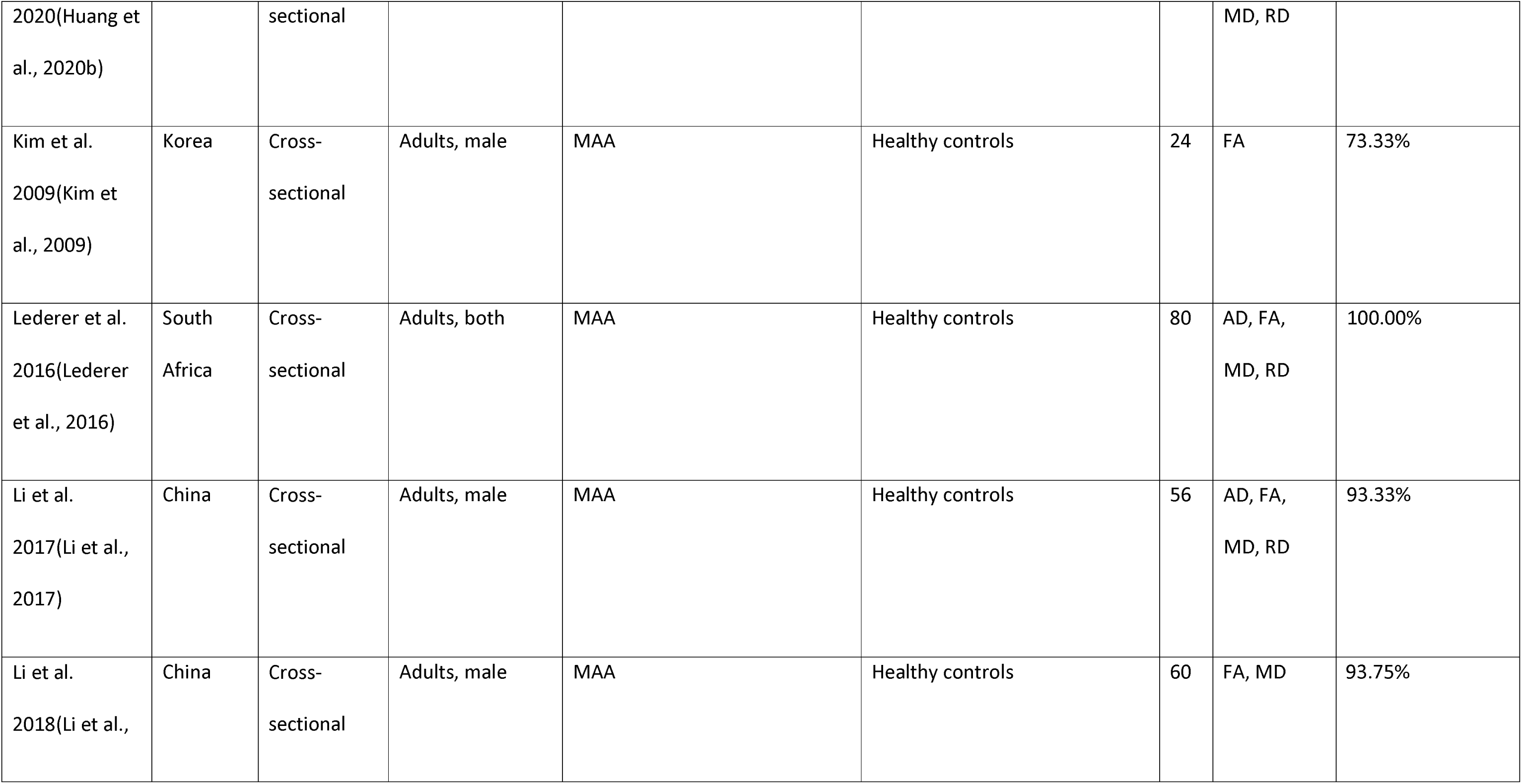
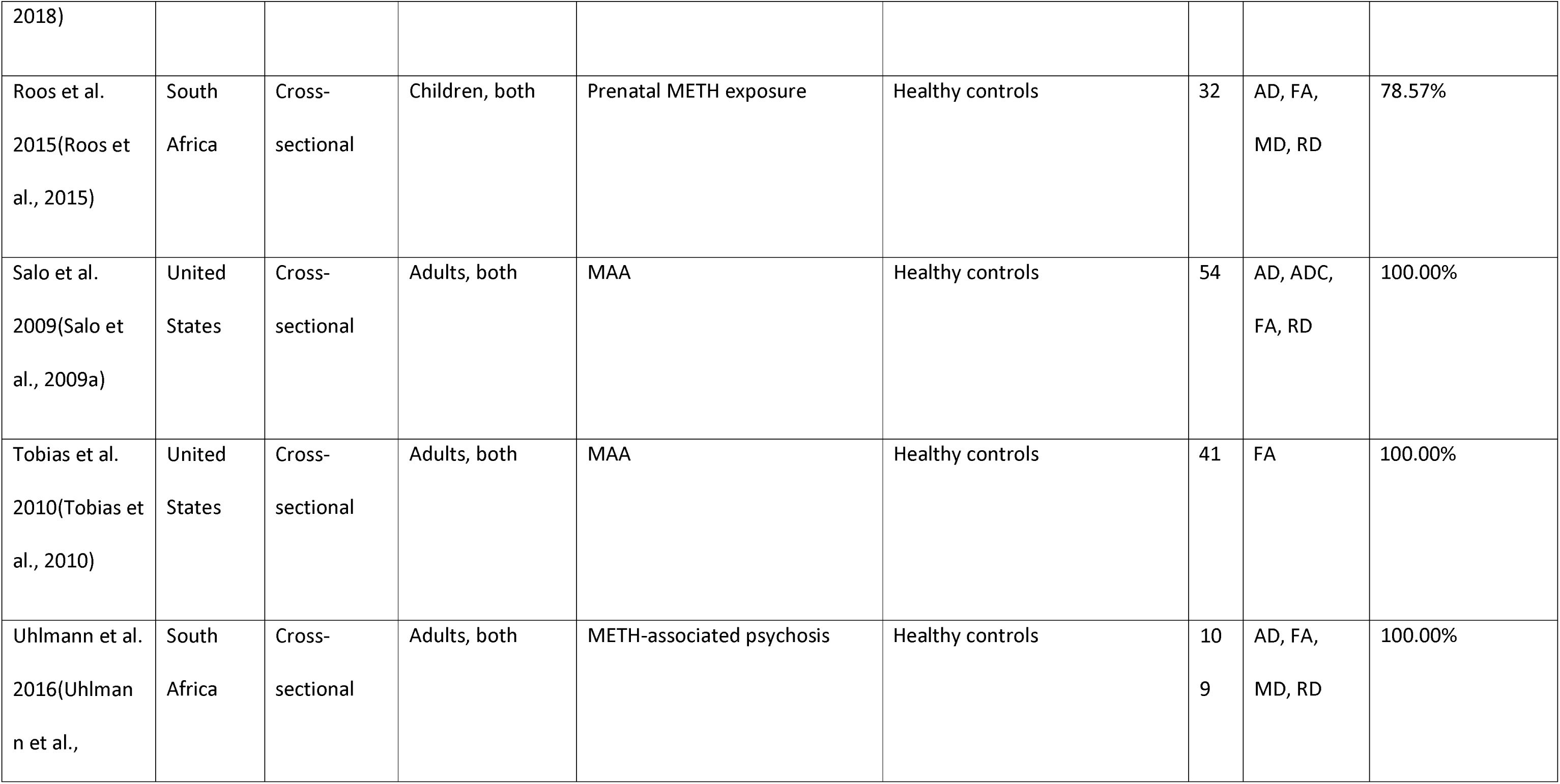
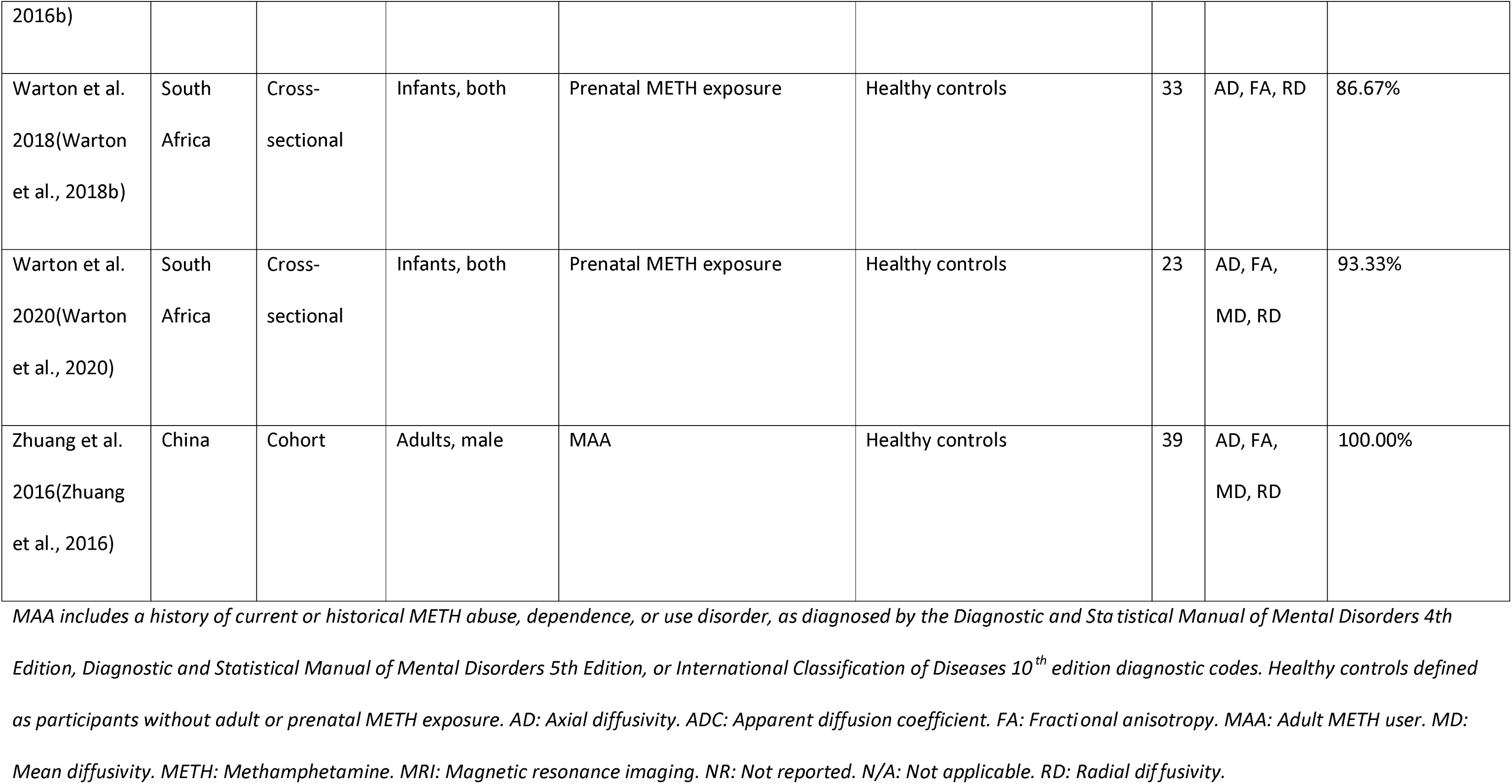
Study demographics and quality appraisal score for diffusion MRI studies.

| Reference | Country | Study design | Age group, sex | Experimental group | Comparison group | N | Diffusivity parameter s | Quality appraisal score |
| --- | --- | --- | --- | --- | --- | --- | --- | --- |
| Alicata et al. 2009(Alicata et al., 2009) | United States | Cross-sectional | Adults, both | MAA | Healthy controls | 60 | AD, ADC, FA, RD | 93.75% |
| Andres et al. 2016(Andres et al., 2016) | United States | Cross-sectional | Adults, both | MAA | Healthy controls | 94 | AD, FA, MD, RD | 100.00% |
| Breen et al. 2017(Breen et al., 2017) | South Africa | Cross-sectional | Adults, both | MAA | Healthy controls | 42 | AD, FA, MD, RD | 93.33% |
| Chang et al.<br>2016(Chang et al., 2016) | United States | Cohort | Infants, both | Prenatal METH exposure | Healthy controls | 13<br>9 | AD, FA,<br>MD, RD | 93.75% |
| Chung et al.<br>2007(Chung et al., 2007) | Korea | Cross-sectional | Adults, both | MAA | Healthy controls | 62 | FA | 100.00% |
| Cloak et al.<br>2009(Cloak et al., 2009) | Hawaii | Cross-sectional | Children, both | Prenatal METH exposure | Healthy controls | 66 | ADC, FA | 85.71% |
| Colby et al.<br>2012(Colby et al., 2012) | United States | Cross-sectional | Children, both | Prenatal METH exposure | Healthy controls | 67 | AD, FA,<br>MD, RD | 85.71% |
| Huang et al. | China | Cross- | Adults, both | MAA | Healthy controls | 50 | AD, FA, | 100.00% |
| 2020(Huang et al., 2020b) |  | sectional |  |  |  |  | MD, RD |  |
| Kim et al. 2009(Kim et al., 2009) | Korea | Cross-sectional | Adults, male | MAA | Healthy controls | 24 | FA | 73.33% |
| Lederer et al. 2016(Lederer et al., 2016) | South Africa | Cross-sectional | Adults, both | MAA | Healthy controls | 80 | AD, FA, MD, RD | 100.00% |
| Li et al. 2017(Li et al., 2017) | China | Cross-sectional | Adults, male | MAA | Healthy controls | 56 | AD, FA, MD, RD | 93.33% |
| Li et al. 2018(Li et al., | China | Cross-sectional | Adults, male | MAA | Healthy controls | 60 | FA, MD | 93.75% |
| 2018) |  |  |  |  |  |  |  |  |
| Roos et al.<br>2015(Roos et al., 2015) | South Africa | Cross-sectional | Children, both | Prenatal METH exposure | Healthy controls | 32 | AD, FA, MD, RD | 78.57% |
| Salo et al.<br>2009(Salo et al., 2009a) | United States | Cross-sectional | Adults, both | MAA | Healthy controls | 54 | AD, ADC, FA, RD | 100.00% |
| Tobias et al.<br>2010(Tobias et al., 2010) | United States | Cross-sectional | Adults, both | MAA | Healthy controls | 41 | FA | 100.00% |
| Uhlmann et al.<br>2016(Uhlman n et al., | South Africa | Cross-sectional | Adults, both | METH-associated psychosis | Healthy controls | 10<br>9 | AD, FA, MD, RD | 100.00% |
| 2016b) |  |  |  |  |  |  |  |  |
| Warton et al.<br>2018(Warton<br>et al., 2018b) | South<br>Africa | Cross-sectional | Infants, both | Prenatal METH exposure | Healthy controls | 33 | AD, FA, RD | 86.67% |
| Warton et al.<br>2020(Warton<br>et al., 2020) | South<br>Africa | Cross-sectional | Infants, both | Prenatal METH exposure | Healthy controls | 23 | AD, FA,<br>MD, RD | 93.33% |
| Zhuang et al.<br>2016(Zhuang<br>et al., 2016) | China | Cohort | Adults, male | MAA | Healthy controls | 39 | AD, FA,<br>MD, RD | 100.00% |

MAA includes a history of current or historical METH abuse, dependence, or use disorder, as diagnosed by the Diagnostic and Statistical Manual of Mental Disorders 4th Edition, Diagnostic and Statistical Manual of Mental Disorders 5th Edition, or International Classification of Diseases 10<sup>th</sup> edition diagnostic codes. Healthy controls defined as participants without adult or prenatal METH exposure. AD: Axial diffusivity. ADC: Apparent diffusion coefficient. FA: Fractional anisotropy. MAA: Adult METH user. MD: Mean diffusivity. METH: Methamphetamine. MRI: Magnetic resonance imaging. NR: Not reported. N/A: Not applicable. RD: Radial diffusivity.

**Table 1c:**
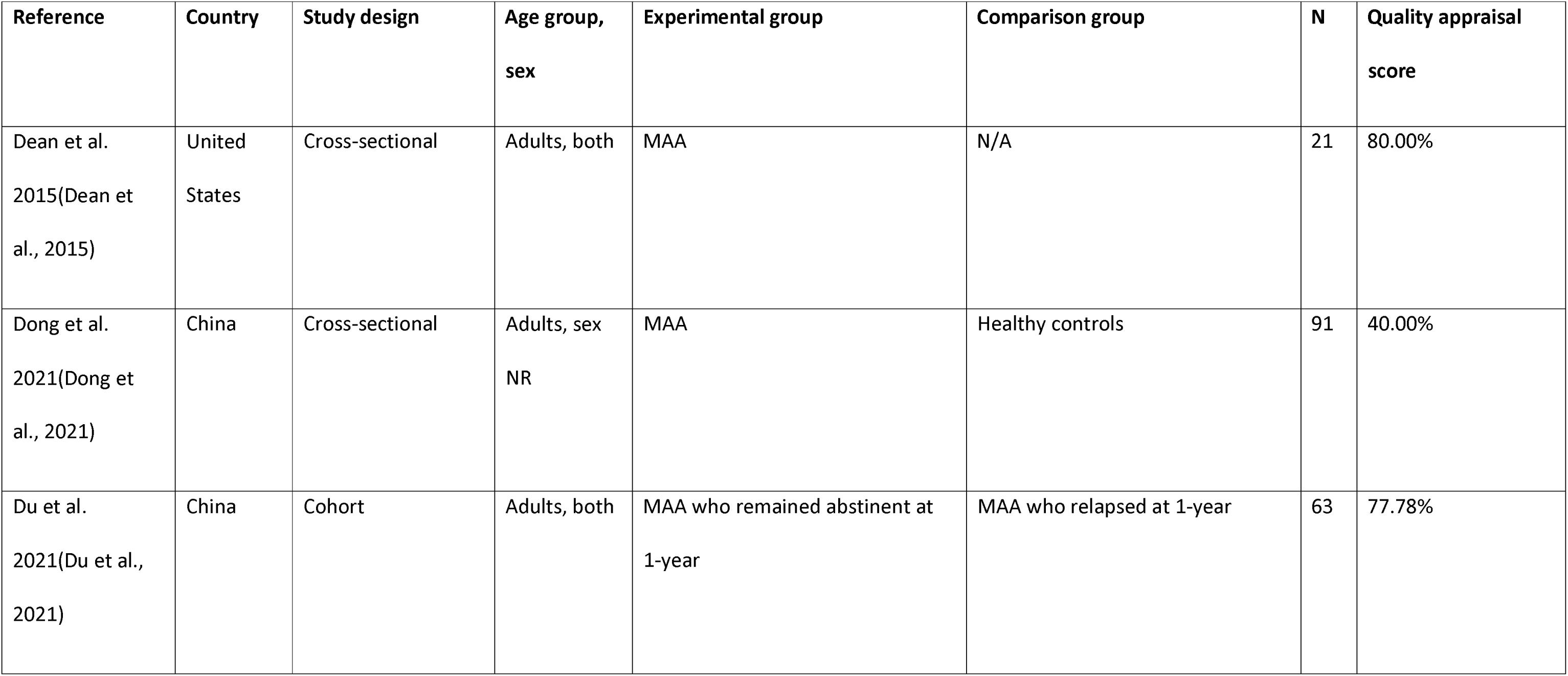
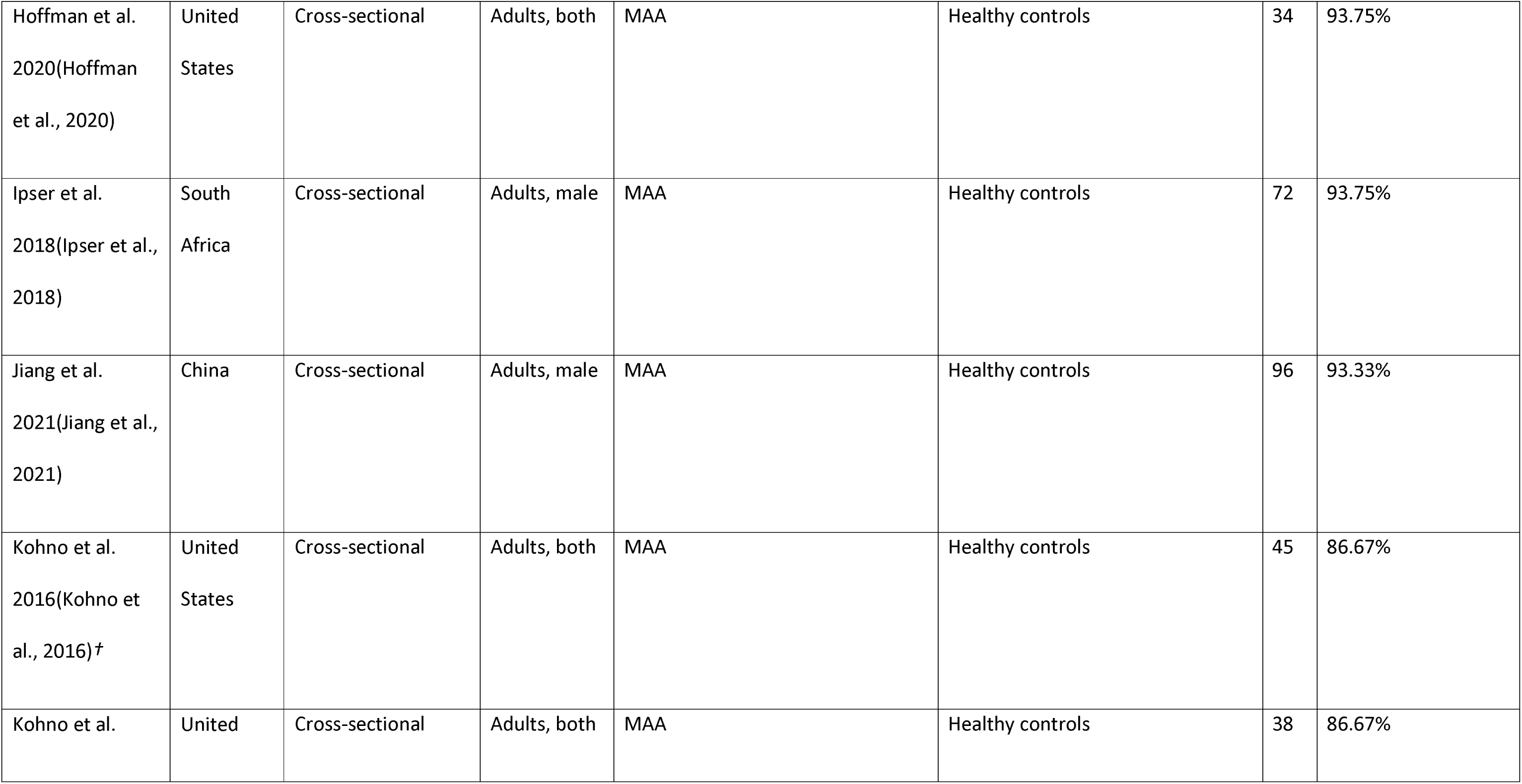
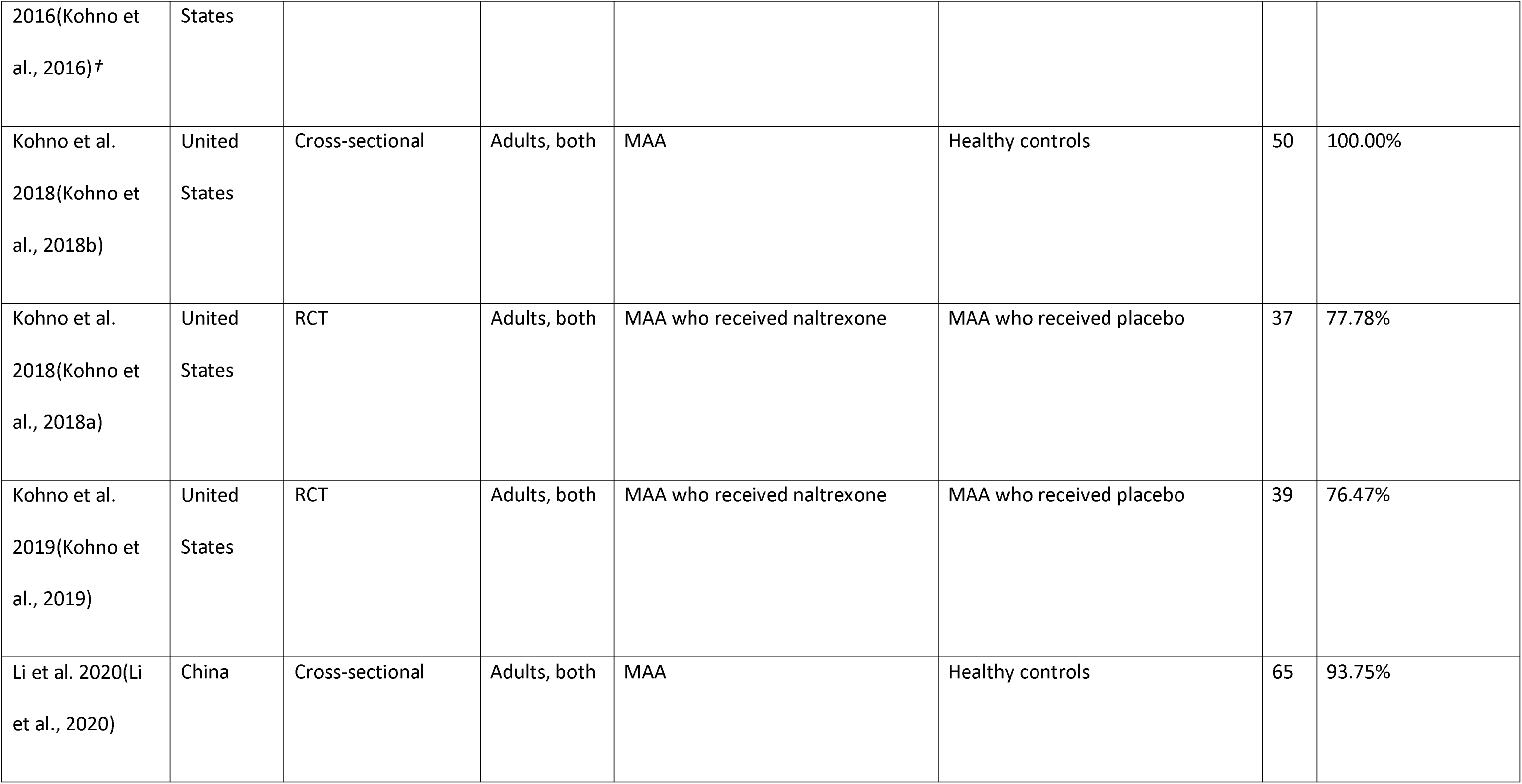
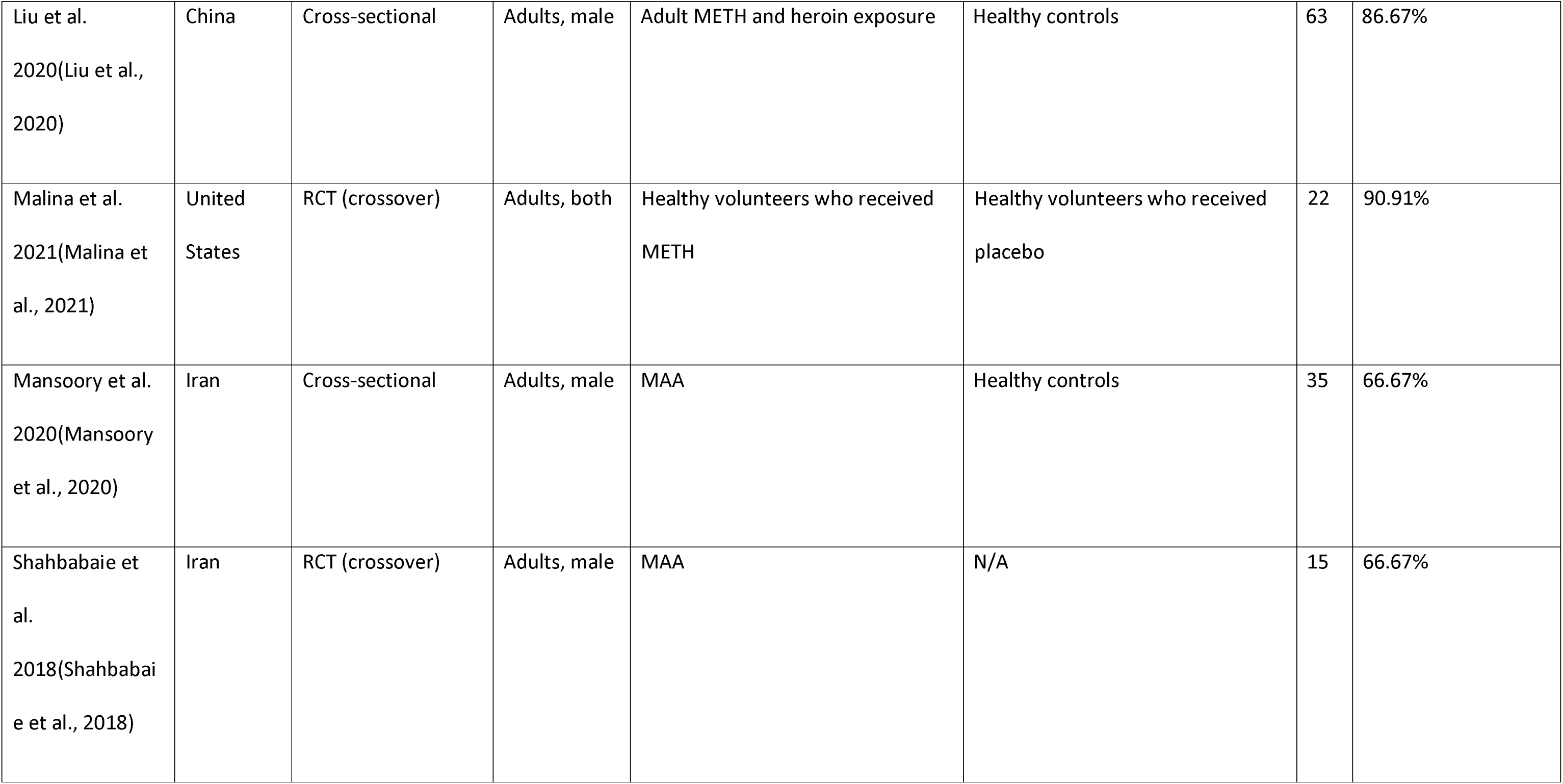
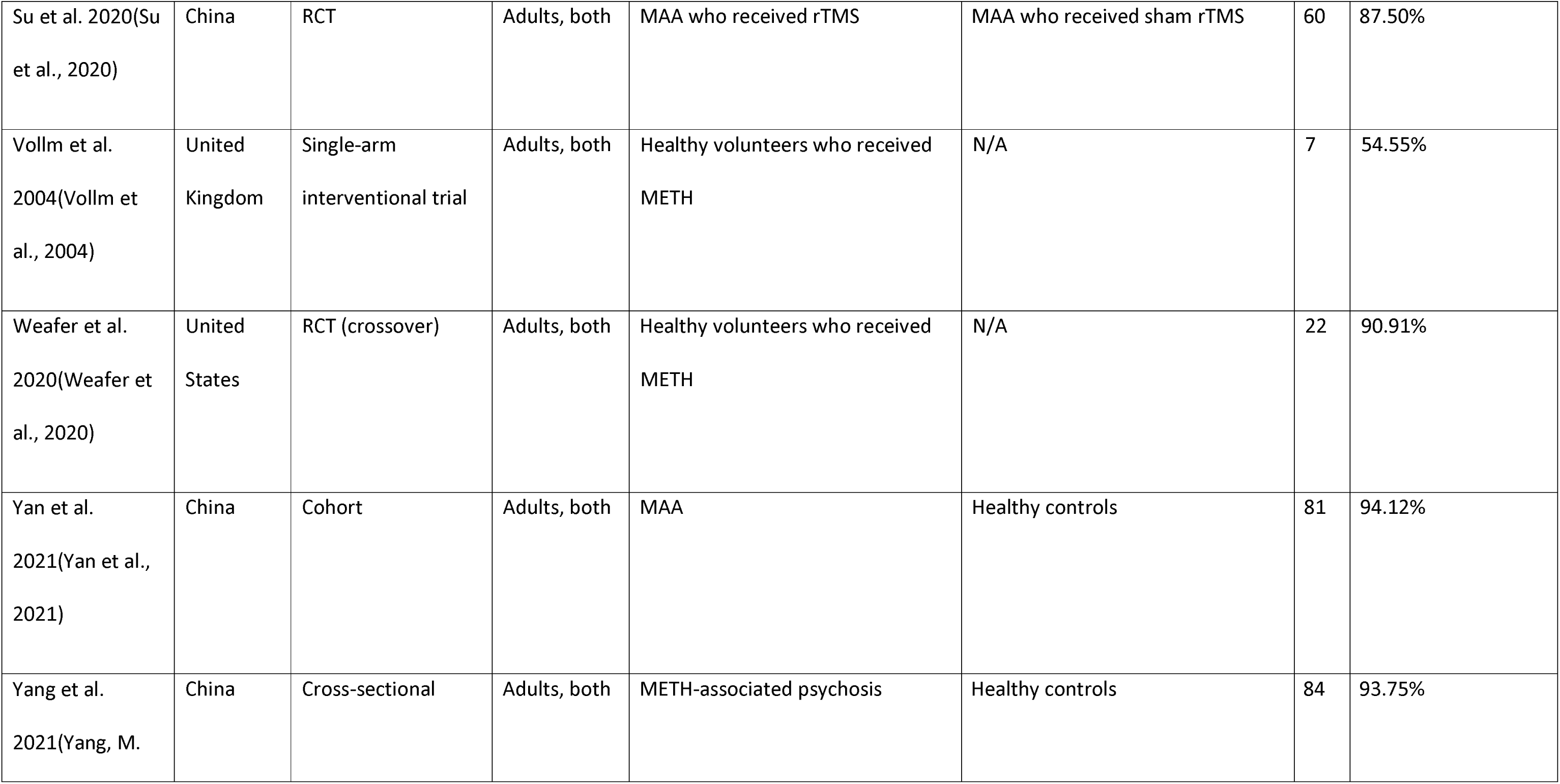
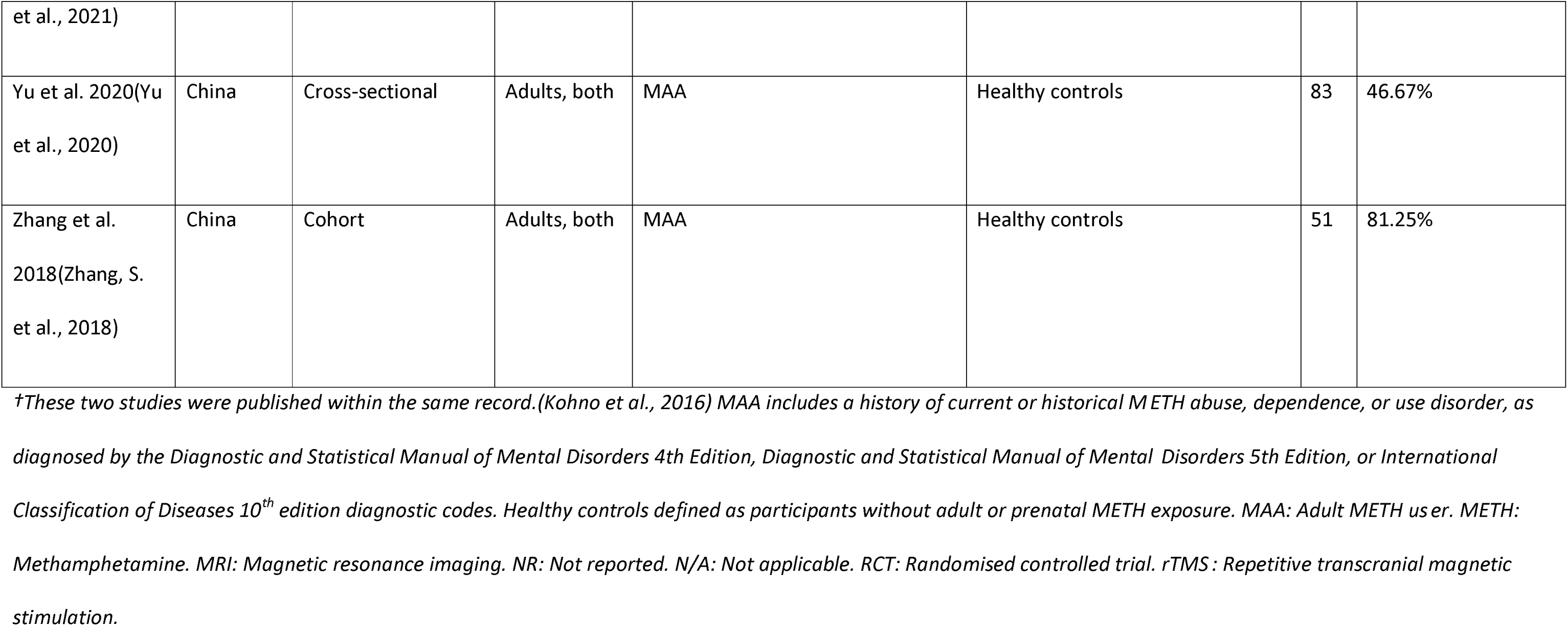
Study demographics and quality appraisal score for resting-state functional MRI studies.

| Reference | Country | Study design | Age group, sex | Experimental group | Comparison group | N | Quality appraisal score |
| --- | --- | --- | --- | --- | --- | --- | --- |
| Dean et al.<br>2015(Dean et al., 2015) | United States | Cross-sectional | Adults, both | MAA | N/A | 21 | 80.00% |
| Dong et al.<br>2021(Dong et al., 2021) | China | Cross-sectional | Adults, sex<br>NR | MAA | Healthy controls | 91 | 40.00% |
| Du et al.<br>2021(Du et al., 2021) | China | Cohort | Adults, both | MAA who remained abstinent at 1-year | MAA who relapsed at 1-year | 63 | 77.78% |
| Hoffman et al.<br>2020(Hoffman<br>et al., 2020) | United<br>States | Cross-sectional | Adults, both | MAA | Healthy controls | 34 | 93.75% |
| Ipser et al.<br>2018(Ipser et al.,<br>2018) | South<br>Africa | Cross-sectional | Adults, male | MAA | Healthy controls | 72 | 93.75% |
| Jiang et al.<br>2021(Jiang et al.,<br>2021) | China | Cross-sectional | Adults, male | MAA | Healthy controls | 96 | 93.33% |
| Kohno et al.<br>2016(Kohno et<br>al., 2016) † | United<br>States | Cross-sectional | Adults, both | MAA | Healthy controls | 45 | 86.67% |
| Kohno et al. | United | Cross-sectional | Adults, both | MAA | Healthy controls | 38 | 86.67% |
| 2016(Kohno et al., 2016)† | States |  |  |  |  |  |  |
| Kohno et al. 2018(Kohno et al., 2018b) | United States | Cross-sectional | Adults, both | MAA | Healthy controls | 50 | 100.00% |
| Kohno et al. 2018(Kohno et al., 2018a) | United States | RCT | Adults, both | MAA who received naltrexone | MAA who received placebo | 37 | 77.78% |
| Kohno et al. 2019(Kohno et al., 2019) | United States | RCT | Adults, both | MAA who received naltrexone | MAA who received placebo | 39 | 76.47% |
| Li et al. 2020(Li et al., 2020) | China | Cross-sectional | Adults, both | MAA | Healthy controls | 65 | 93.75% |
| Liu et al.<br>2020(Liu et al.,<br>2020) | China | Cross-sectional | Adults, male | Adult METH and heroin exposure | Healthy controls | 63 | 86.67% |
| Malina et al.<br>2021(Malina et<br>al., 2021) | United<br>States | RCT (crossover) | Adults, both | Healthy volunteers who received<br>METH | Healthy volunteers who received<br>placebo | 22 | 90.91% |
| Mansoori et al.<br>2020(Mansoori<br>et al., 2020) | Iran | Cross-sectional | Adults, male | MAA | Healthy controls | 35 | 66.67% |
| Shahbabaie et<br>al.<br>2018(Shahbabaie<br>et al., 2018) | Iran | RCT (crossover) | Adults, male | MAA | N/A | 15 | 66.67% |
| Su et al. 2020(Su et al., 2020) | China | RCT | Adults, both | MAA who received rTMS | MAA who received sham rTMS | 60 | 87.50% |
| Vollm et al. 2004(Vollm et al., 2004) | United Kingdom | Single-arm interventional trial | Adults, both | Healthy volunteers who received METH | N/A | 7 | 54.55% |
| Weafer et al. 2020(Weafer et al., 2020) | United States | RCT (crossover) | Adults, both | Healthy volunteers who received METH | N/A | 22 | 90.91% |
| Yan et al. 2021(Yan et al., 2021) | China | Cohort | Adults, both | MAA | Healthy controls | 81 | 94.12% |
| Yang et al. 2021(Yang, M. | China | Cross-sectional | Adults, both | METH-associated psychosis | Healthy controls | 84 | 93.75% |
| et al., 2021) |  |  |  |  |  |  |  |
| Yu et al. 2020(Yu et al., 2020) | China | Cross-sectional | Adults, both | MAA | Healthy controls | 83 | 46.67% |
| Zhang et al. 2018(Zhang, S. et al., 2018) | China | Cohort | Adults, both | MAA | Healthy controls | 51 | 81.25% |

*†These two studies were published within the same record.(Kohno et al., 2016) MAA includes a history of current or historical METH abuse, dependence, or use disorder, as diagnosed by the Diagnostic and Statistical Manual of Mental Disorders 4th Edition, Diagnostic and Statistical Manual of Mental Disorders 5th Edition, or International Classification of Diseases 10<sup>th</sup> edition diagnostic codes. Healthy controls defined as participants without adult or prenatal METH exposure. MAA: Adult METH user. METH: Methamphetamine. MRI: Magnetic resonance imaging. NR: Not reported. N/A: Not applicable. RCT: Randomised controlled trial. rTMS: Repetitive transcranial magnetic stimulation.*

**Table 1d:**
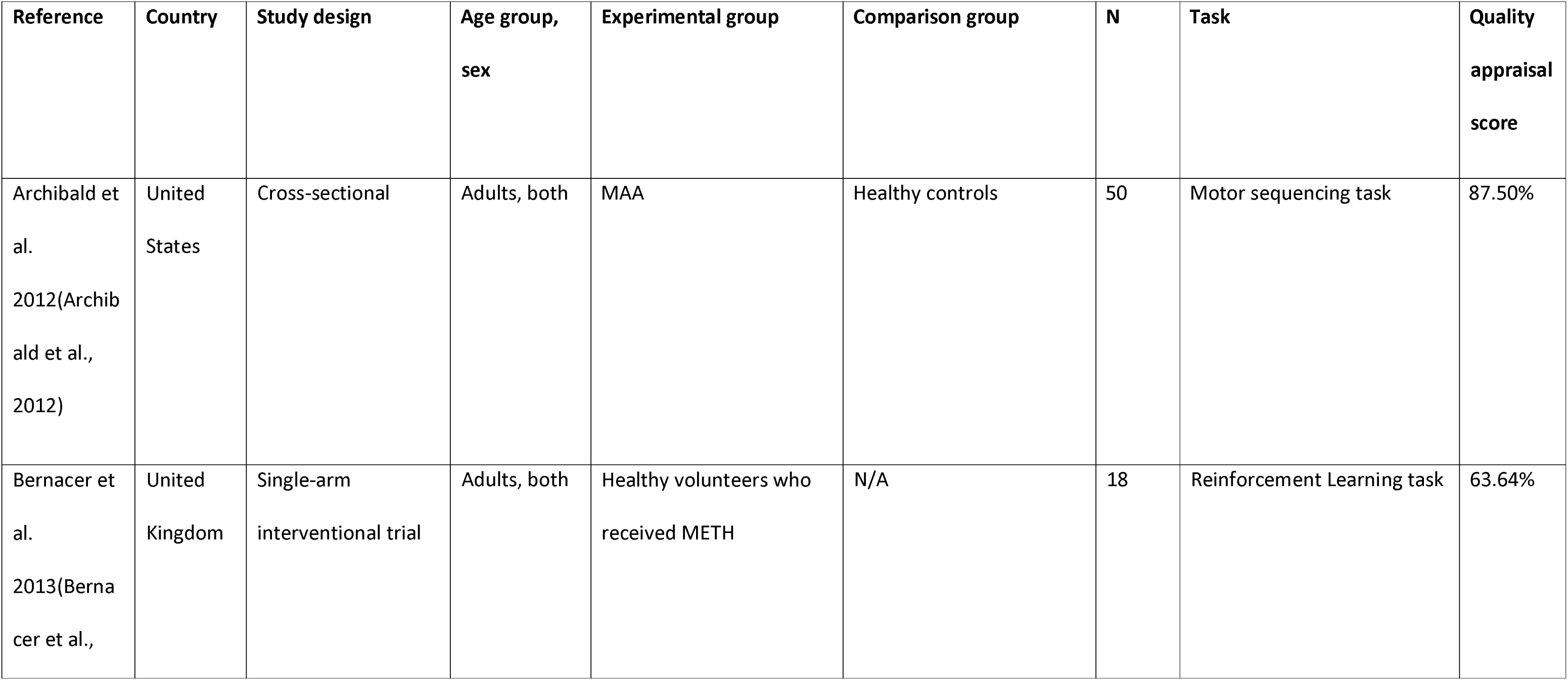
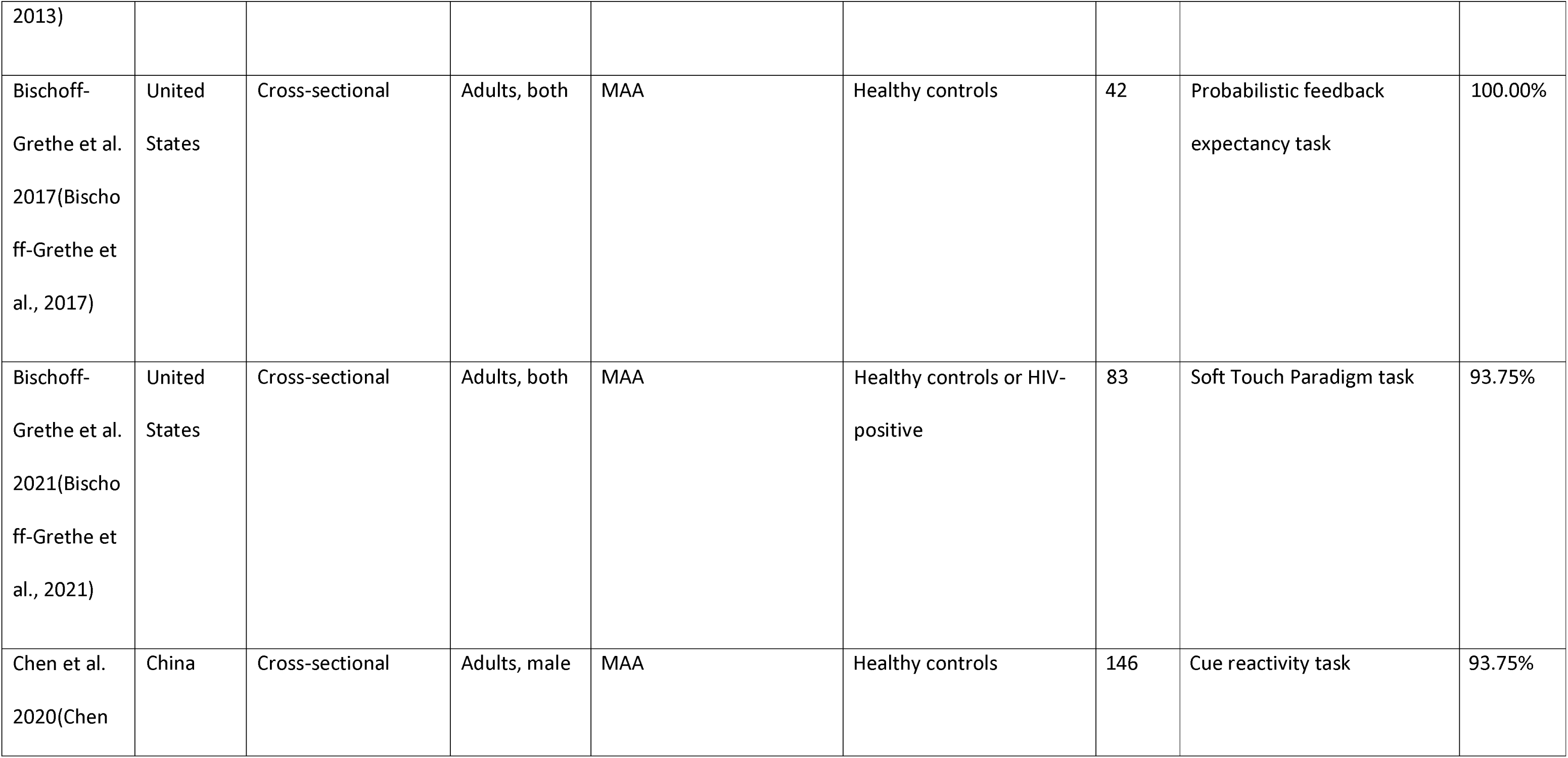
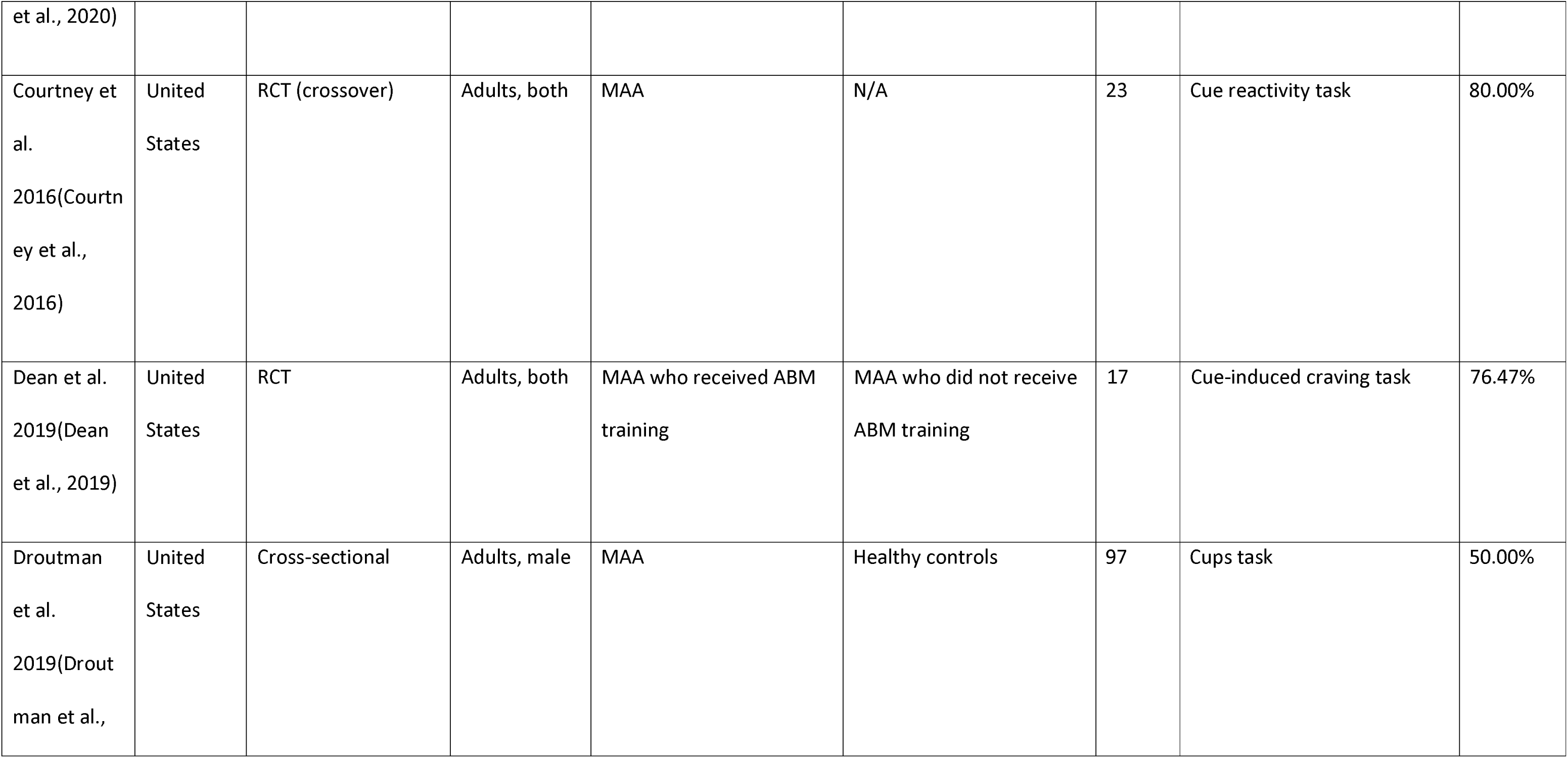
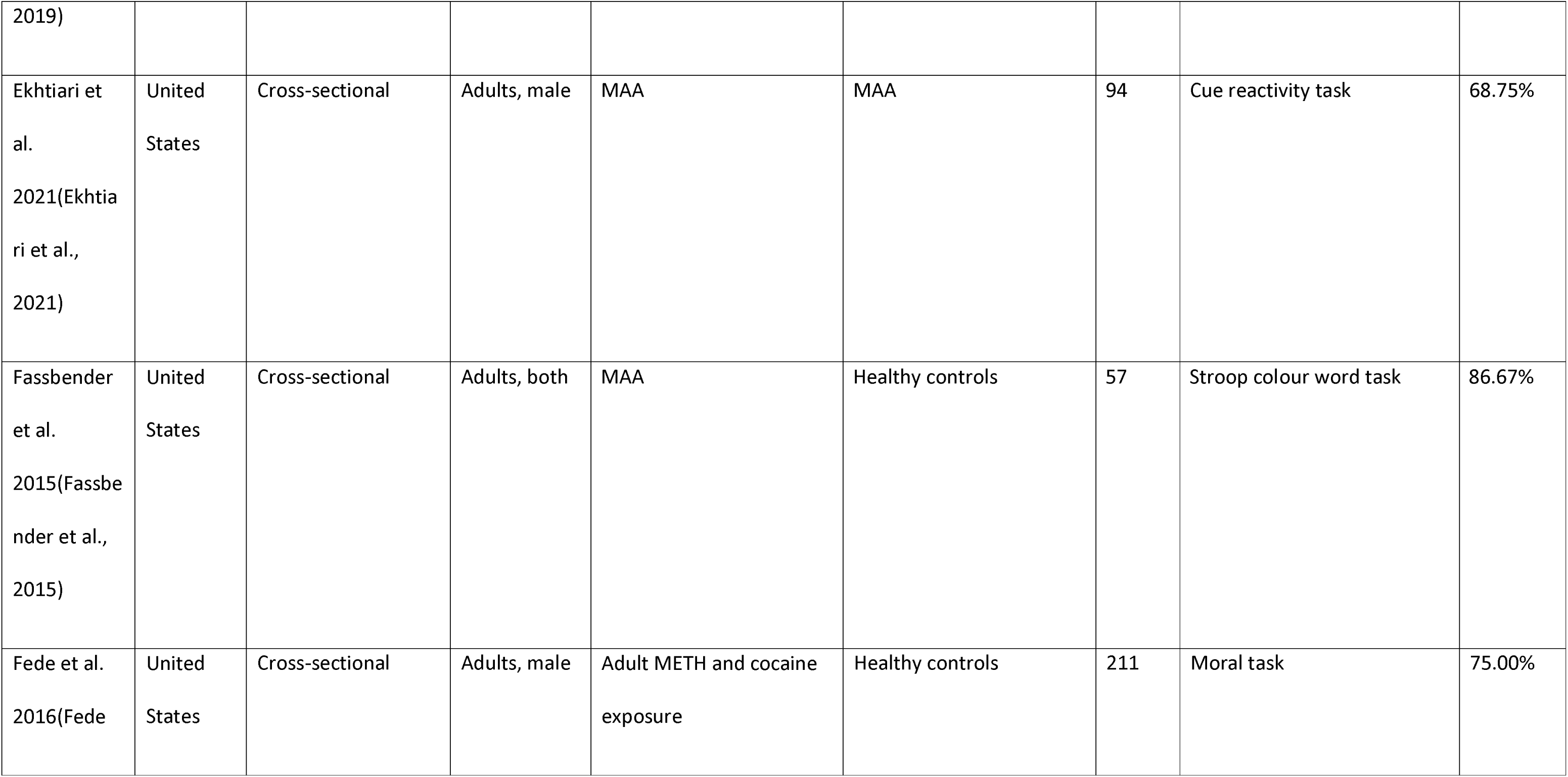
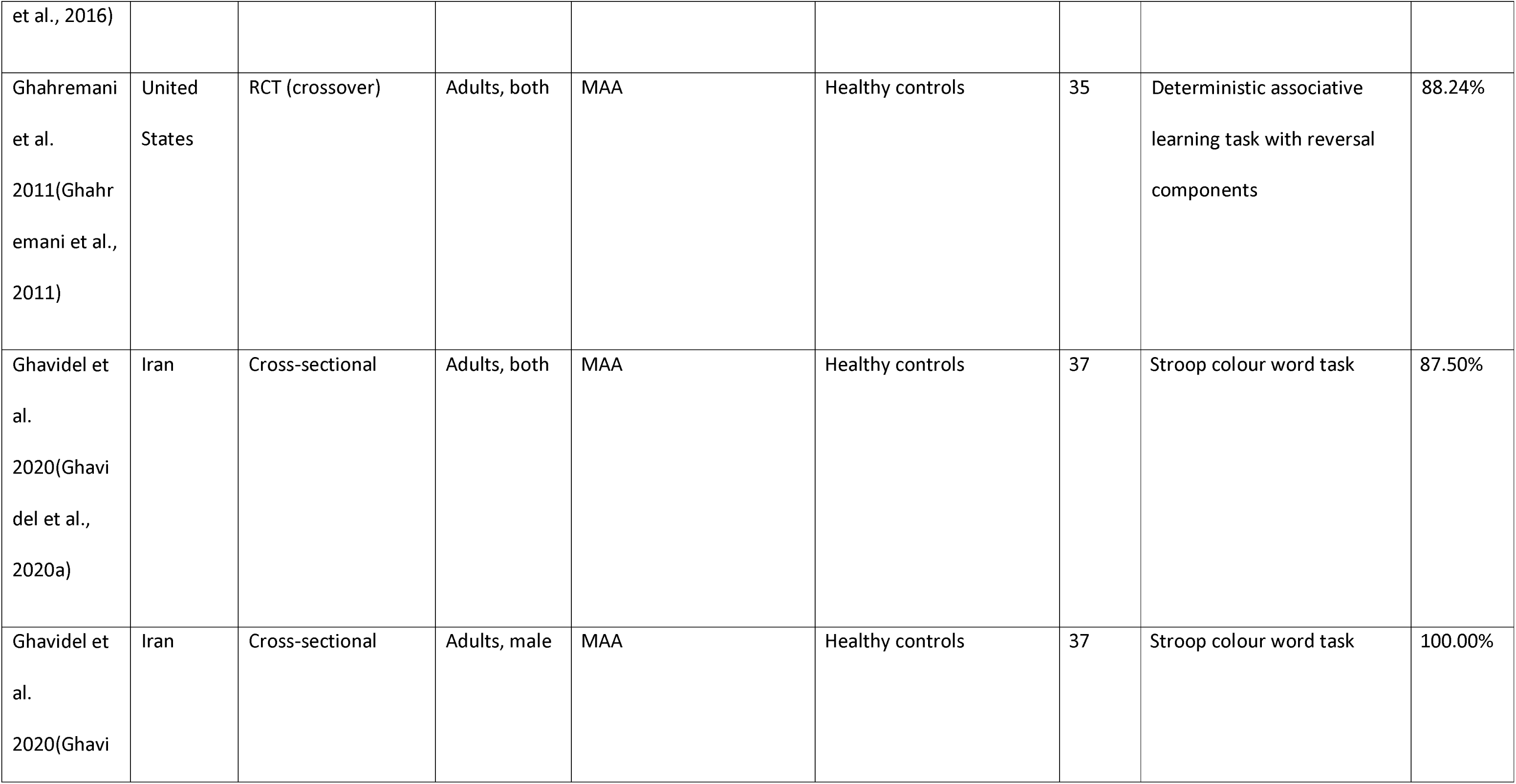
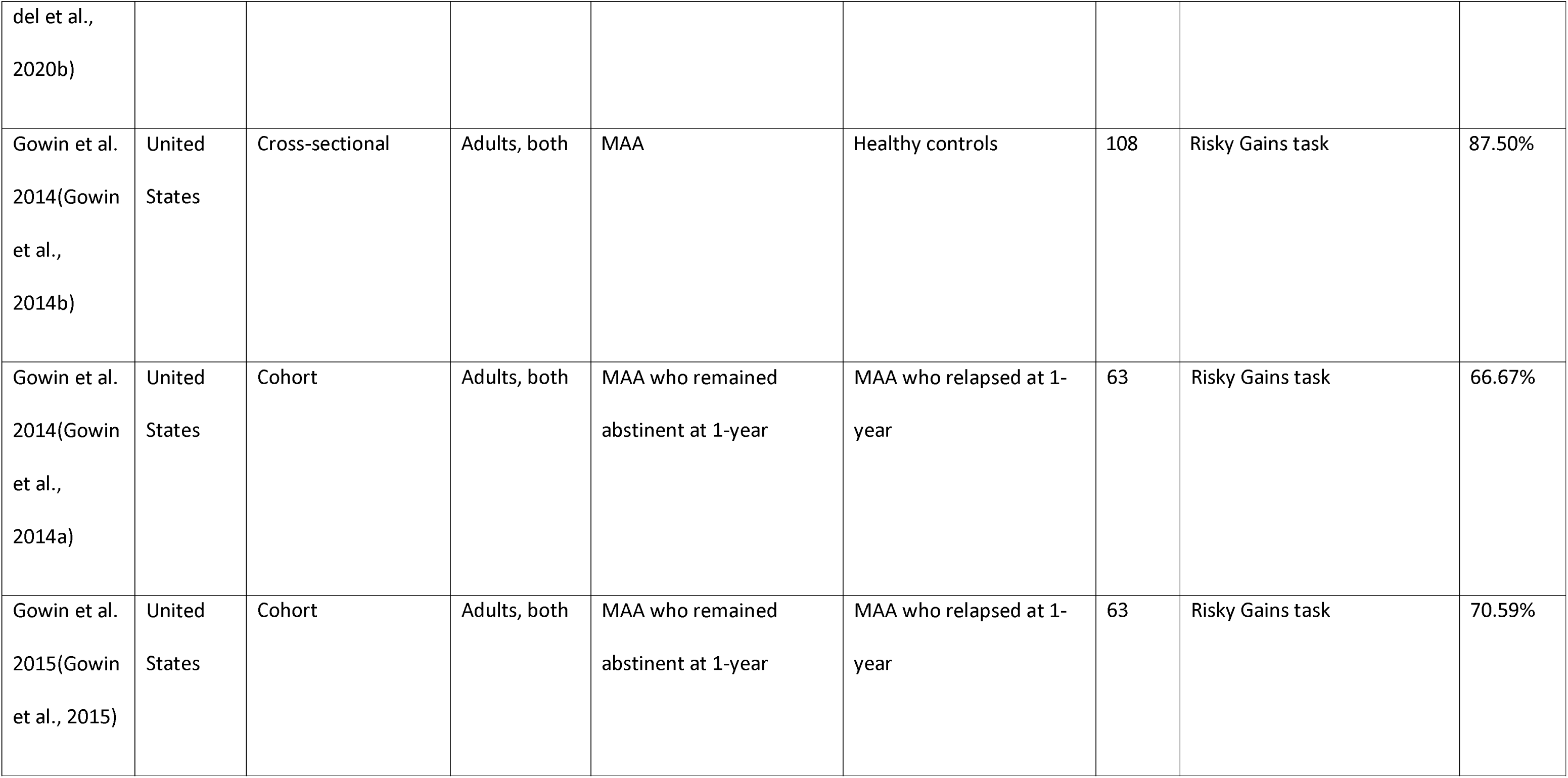
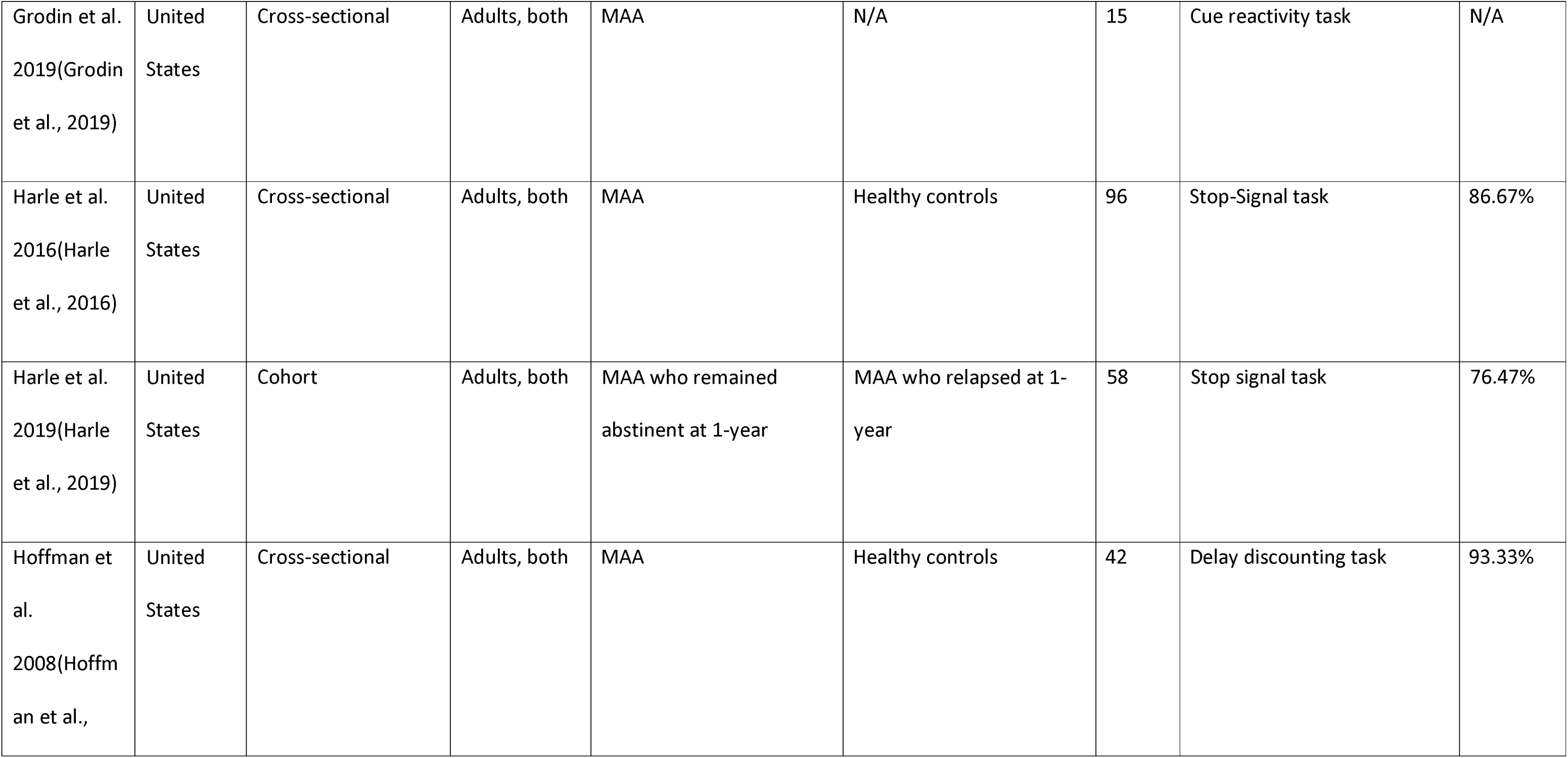
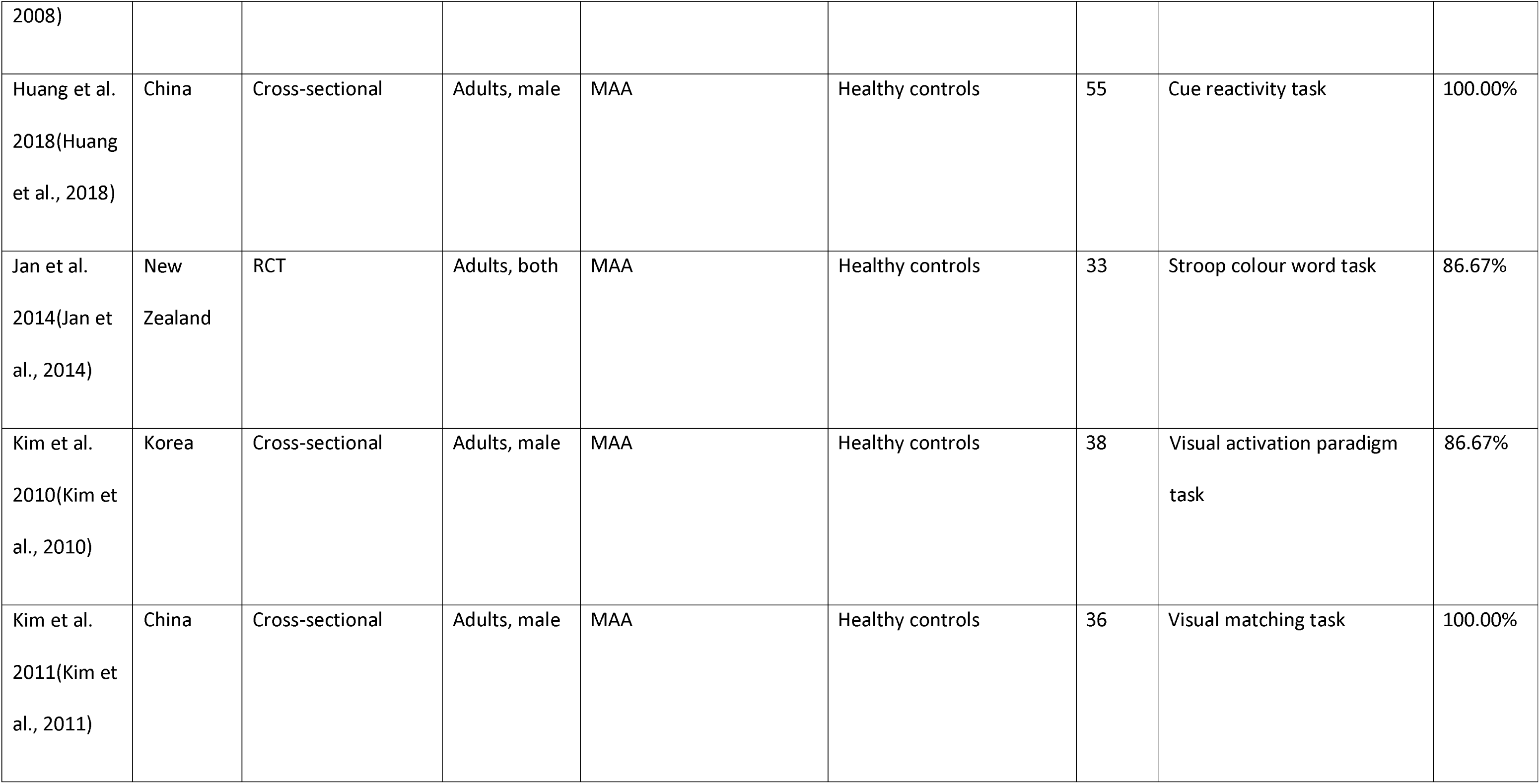
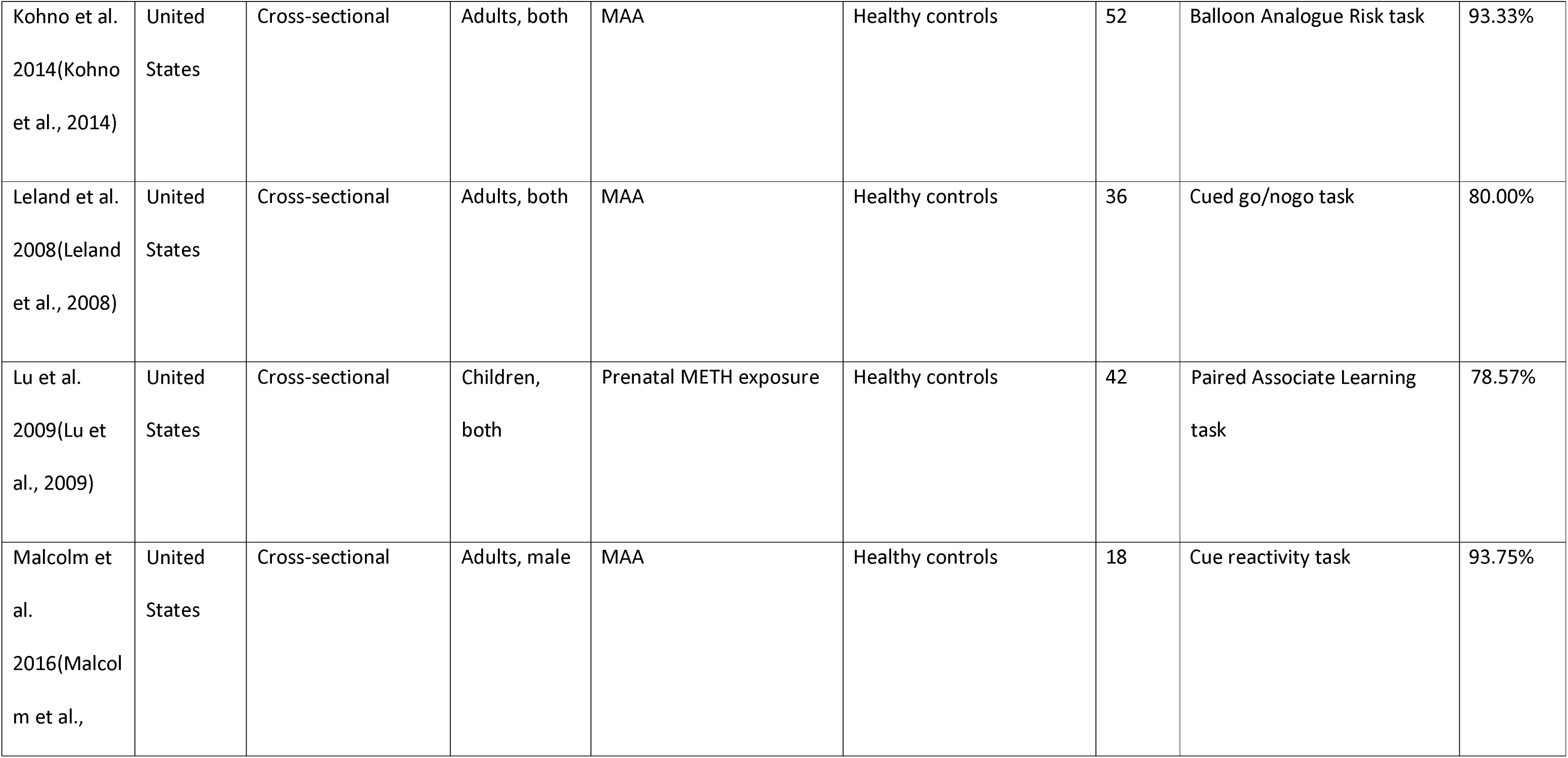
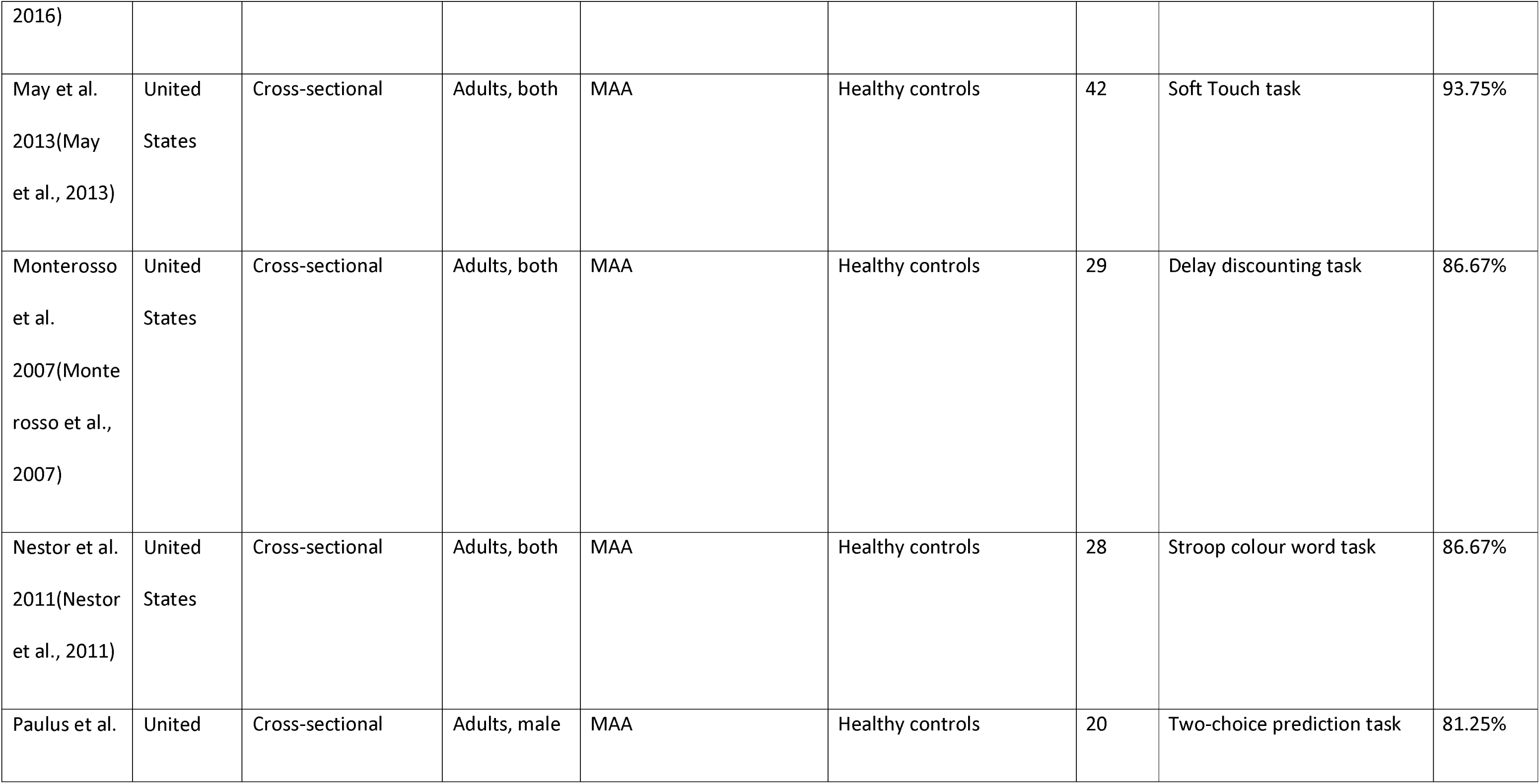
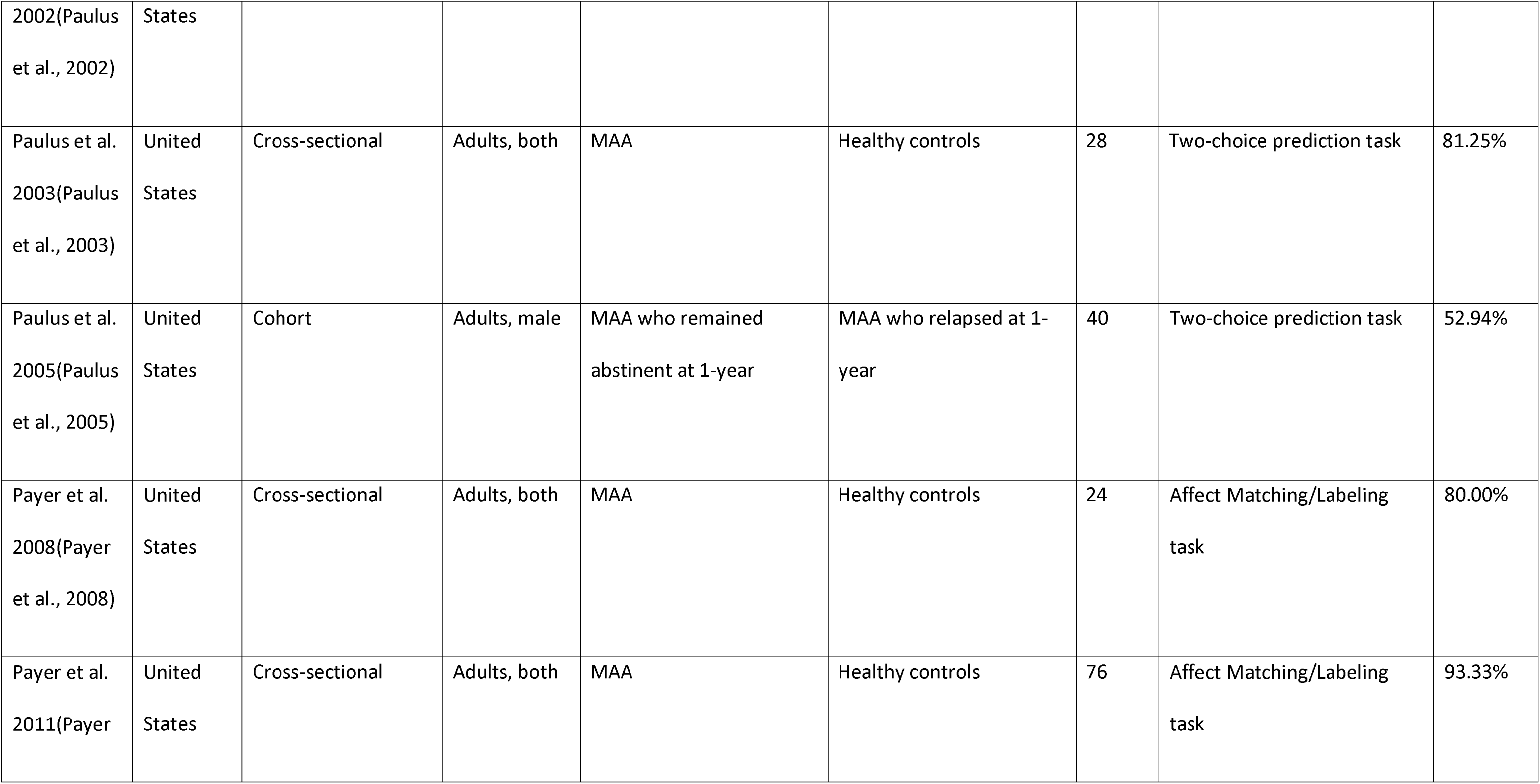
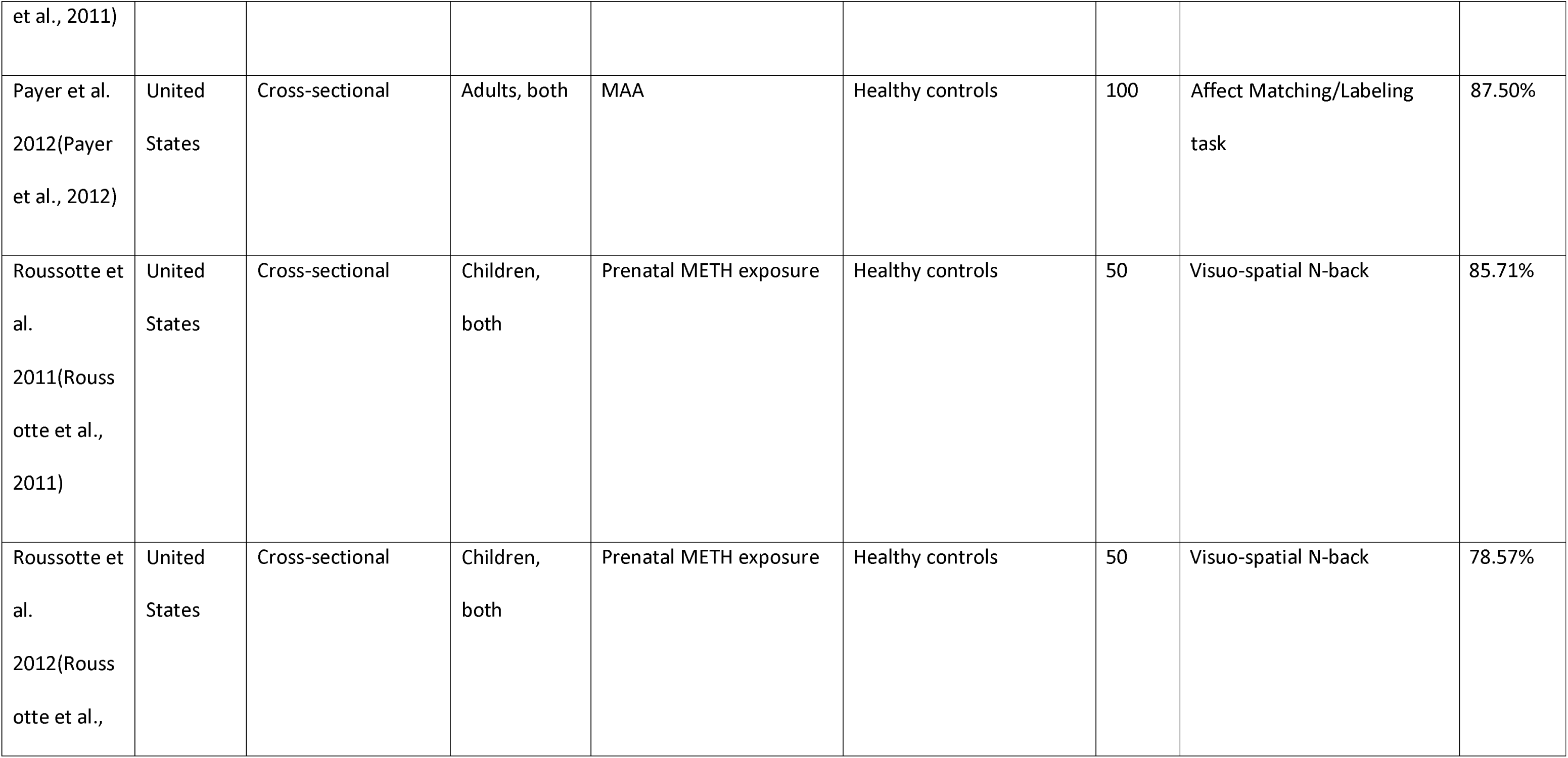
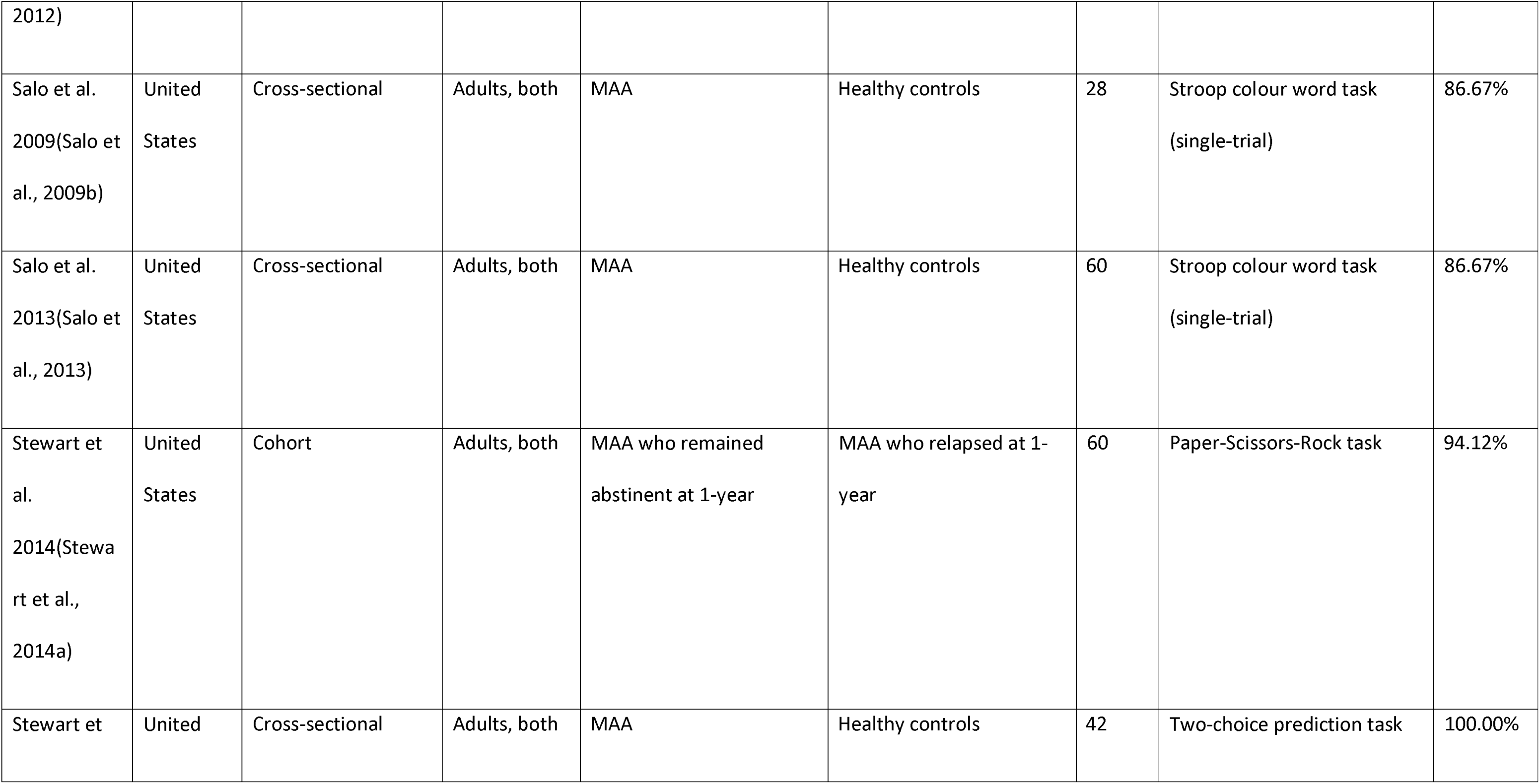
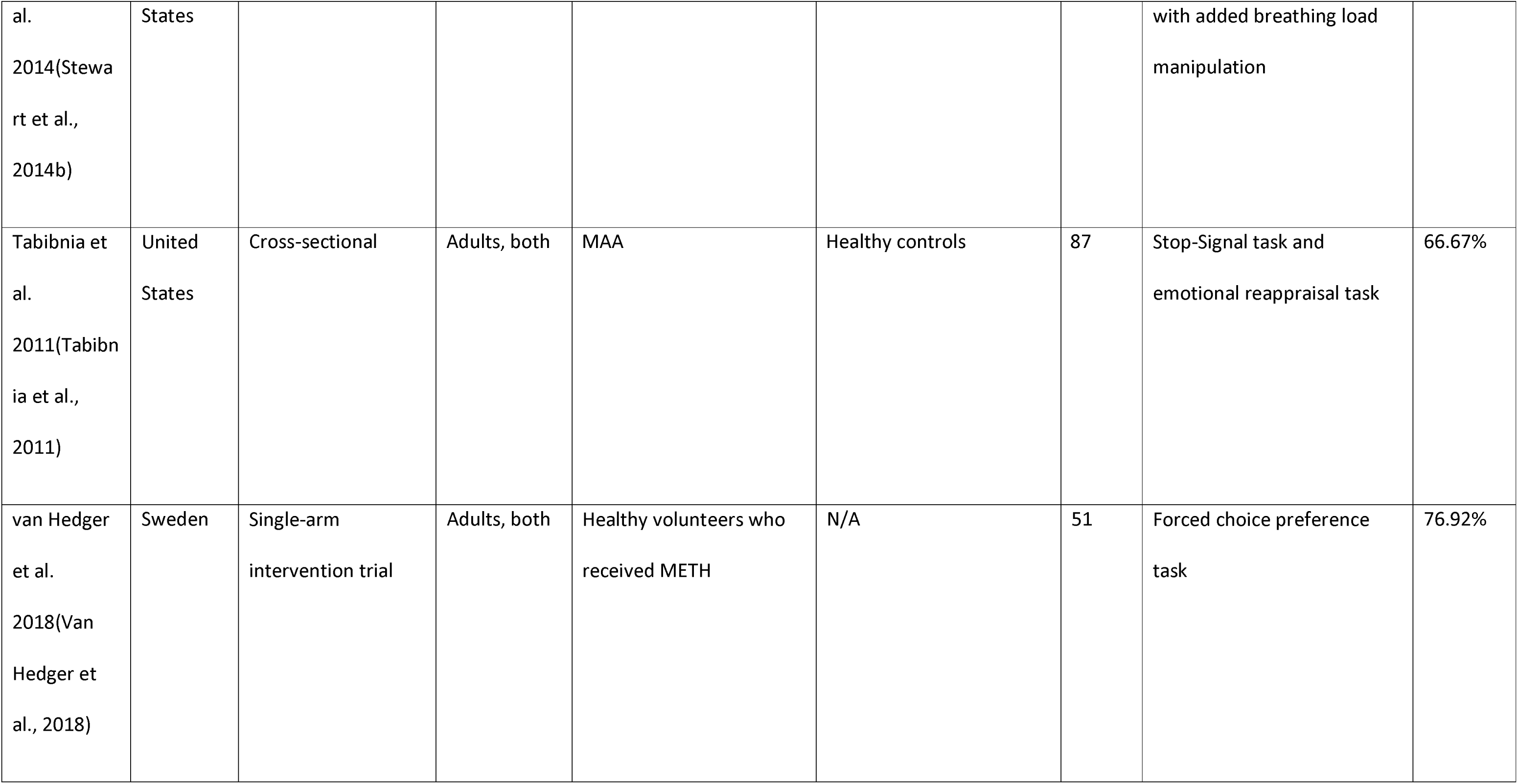
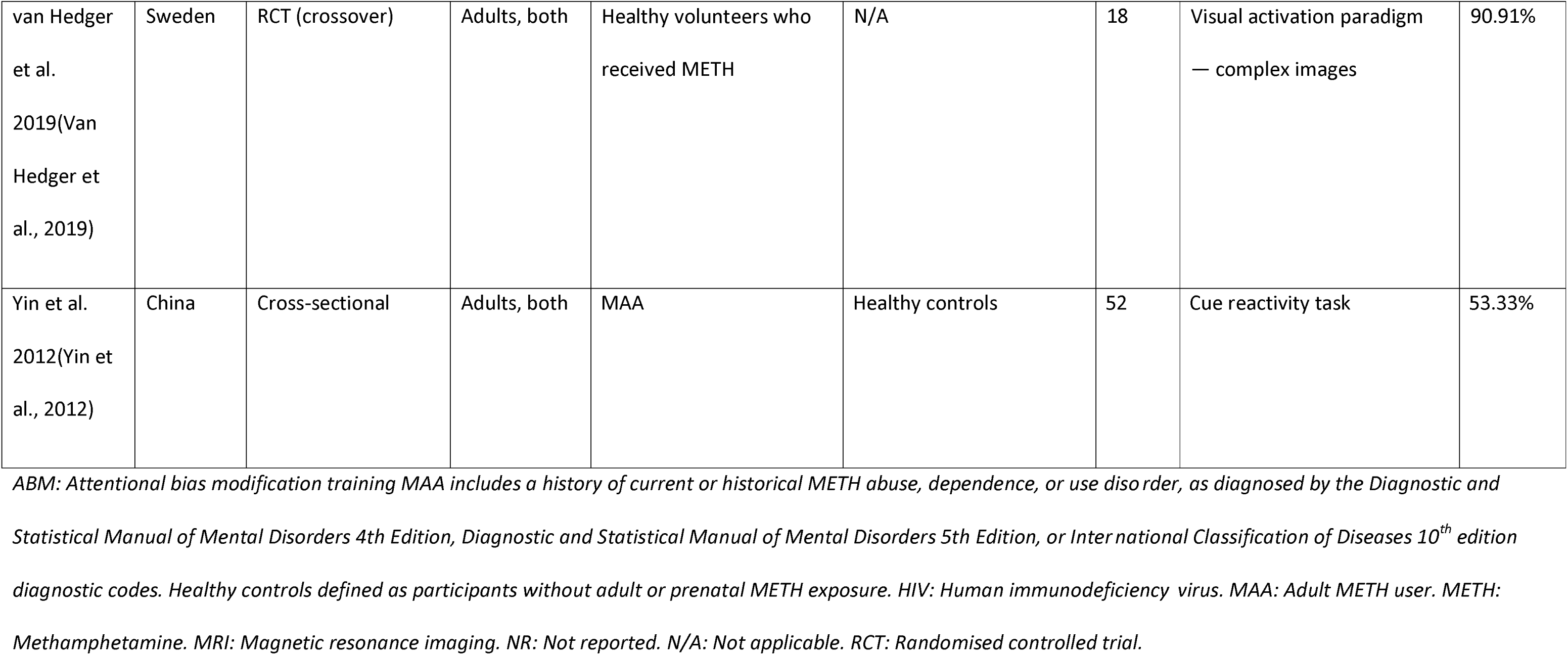
Study demographics and quality appraisal score for task-based functional MRI studies.

| Reference | Country | Study design | Age group, sex | Experimental group | Comparison group | N | Task | Quality appraisal score |
| --- | --- | --- | --- | --- | --- | --- | --- | --- |
| Archibald et al.<br>2012(Archibald et al., 2012) | United States | Cross-sectional | Adults, both | MAA | Healthy controls | 50 | Motor sequencing task | 87.50% |
| Bernacer et al.<br>2013(Bernacer et al., 2013) | United Kingdom | Single-arm interventional trial | Adults, both | Healthy volunteers who received METH | N/A | 18 | Reinforcement Learning task | 63.64% |
| 2013) |  |  |  |  |  |  |  |  |
| Bischoff-Grethe et al. 2017(Bischoff-Grethe et al., 2017) | United States | Cross-sectional | Adults, both | MAA | Healthy controls | 42 | Probabilistic feedback expectancy task | 100.00% |
| Bischoff-Grethe et al. 2021(Bischoff-Grethe et al., 2021) | United States | Cross-sectional | Adults, both | MAA | Healthy controls or HIV-positive | 83 | Soft Touch Paradigm task | 93.75% |
| Chen et al. 2020(Chen | China | Cross-sectional | Adults, male | MAA | Healthy controls | 146 | Cue reactivity task | 93.75% |
| et al., 2020) |  |  |  |  |  |  |  |  |
| Courtney et al.<br>2016(Courtn<br>ey et al.,<br>2016) | United States | RCT (crossover) | Adults, both | MAA | N/A | 23 | Cue reactivity task | 80.00% |
| Dean et al.<br>2019(Dean<br>et al., 2019) | United States | RCT | Adults, both | MAA who received ABM training | MAA who did not receive ABM training | 17 | Cue-induced craving task | 76.47% |
| Droutman et al.<br>2019(Drout<br>man et al., | United States | Cross-sectional | Adults, male | MAA | Healthy controls | 97 | Cups task | 50.00% |
| 2019) |  |  |  |  |  |  |  |  |
| Ekhtiari et al.<br>2021(Ekhtiari et al., 2021) | United States | Cross-sectional | Adults, male | MAA | MAA | 94 | Cue reactivity task | 68.75% |
| Fassbender et al.<br>2015(Fassbender et al., 2015) | United States | Cross-sectional | Adults, both | MAA | Healthy controls | 57 | Stroop colour word task | 86.67% |
| Fede et al.<br>2016(Fede | United States | Cross-sectional | Adults, male | Adult METH and cocaine exposure | Healthy controls | 211 | Moral task | 75.00% |
| et al., 2016) |  |  |  |  |  |  |  |  |
| Ghahremani<br>et al.<br>2011(Ghahremani et al.,<br>2011) | United<br>States | RCT (crossover) | Adults, both | MAA | Healthy controls | 35 | Deterministic associative<br>learning task with reversal<br>components | 88.24% |
| Ghavidel et<br>al.<br>2020(Ghavidel et al.,<br>2020a) | Iran | Cross-sectional | Adults, both | MAA | Healthy controls | 37 | Stroop colour word task | 87.50% |
| Ghavidel et<br>al.<br>2020(Ghavi | Iran | Cross-sectional | Adults, male | MAA | Healthy controls | 37 | Stroop colour word task | 100.00% |
| del et al.,<br>2020b) |  |  |  |  |  |  |  |  |
| Gowin et al.<br>2014(Gowin<br>et al.,<br>2014b) | United<br>States | Cross-sectional | Adults, both | MAA | Healthy controls | 108 | Risky Gains task | 87.50% |
| Gowin et al.<br>2014(Gowin<br>et al.,<br>2014a) | United<br>States | Cohort | Adults, both | MAA who remained<br>abstinent at 1-year | MAA who relapsed at 1-year | 63 | Risky Gains task | 66.67% |
| Gowin et al.<br>2015(Gowin<br>et al., 2015) | United<br>States | Cohort | Adults, both | MAA who remained<br>abstinent at 1-year | MAA who relapsed at 1-year | 63 | Risky Gains task | 70.59% |
| Grodin et al.<br>2019(Grodin<br>et al., 2019) | United<br>States | Cross-sectional | Adults, both | MAA | N/A | 15 | Cue reactivity task | N/A |
| Harle et al.<br>2016(Harle<br>et al., 2016) | United<br>States | Cross-sectional | Adults, both | MAA | Healthy controls | 96 | Stop-Signal task | 86.67% |
| Harle et al.<br>2019(Harle<br>et al., 2019) | United<br>States | Cohort | Adults, both | MAA who remained<br>abstinent at 1-year | MAA who relapsed at 1-year | 58 | Stop signal task | 76.47% |
| Hoffman et<br>al.<br>2008(Hoffm<br>an et al., | United<br>States | Cross-sectional | Adults, both | MAA | Healthy controls | 42 | Delay discounting task | 93.33% |
| 2008) |  |  |  |  |  |  |  |  |
| Huang et al.<br>2018(Huang<br>et al., 2018) | China | Cross-sectional | Adults, male | MAA | Healthy controls | 55 | Cue reactivity task | 100.00% |
| Jan et al.<br>2014(Jan et<br>al., 2014) | New<br>Zealand | RCT | Adults, both | MAA | Healthy controls | 33 | Stroop colour word task | 86.67% |
| Kim et al.<br>2010(Kim et<br>al., 2010) | Korea | Cross-sectional | Adults, male | MAA | Healthy controls | 38 | Visual activation paradigm<br>task | 86.67% |
| Kim et al.<br>2011(Kim et<br>al., 2011) | China | Cross-sectional | Adults, male | MAA | Healthy controls | 36 | Visual matching task | 100.00% |
| Kohno et al.<br>2014(Kohno<br>et al., 2014) | United<br>States | Cross-sectional | Adults, both | MAA | Healthy controls | 52 | Balloon Analogue Risk task | 93.33% |
| Leland et al.<br>2008(Leland<br>et al., 2008) | United<br>States | Cross-sectional | Adults, both | MAA | Healthy controls | 36 | Cued go/nogo task | 80.00% |
| Lu et al.<br>2009(Lu et<br>al., 2009) | United<br>States | Cross-sectional | Children,<br>both | Prenatal METH exposure | Healthy controls | 42 | Paired Associate Learning<br>task | 78.57% |
| Malcolm et<br>al.<br>2016(Malcol<br>m et al., | United<br>States | Cross-sectional | Adults, male | MAA | Healthy controls | 18 | Cue reactivity task | 93.75% |
| 2016) |  |  |  |  |  |  |  |  |
| May et al.<br>2013(May<br>et al., 2013) | United<br>States | Cross-sectional | Adults, both | MAA | Healthy controls | 42 | Soft Touch task | 93.75% |
| Monterosso<br>et al.<br>2007(Monte<br>rosso et al.,<br>2007) | United<br>States | Cross-sectional | Adults, both | MAA | Healthy controls | 29 | Delay discounting task | 86.67% |
| Nestor et al.<br>2011(Nestor<br>et al., 2011) | United<br>States | Cross-sectional | Adults, both | MAA | Healthy controls | 28 | Stroop colour word task | 86.67% |
| Paulus et al. | United | Cross-sectional | Adults, male | MAA | Healthy controls | 20 | Two-choice prediction task | 81.25% |
| 2002(Paulus et al., 2002) | States |  |  |  |  |  |  |  |
| Paulus et al. 2003(Paulus et al., 2003) | United States | Cross-sectional | Adults, both | MAA | Healthy controls | 28 | Two-choice prediction task | 81.25% |
| Paulus et al. 2005(Paulus et al., 2005) | United States | Cohort | Adults, male | MAA who remained abstinent at 1-year | MAA who relapsed at 1-year | 40 | Two-choice prediction task | 52.94% |
| Payer et al. 2008(Payer et al., 2008) | United States | Cross-sectional | Adults, both | MAA | Healthy controls | 24 | Affect Matching/Labeling task | 80.00% |
| Payer et al. 2011(Payer et al., 2011) | United States | Cross-sectional | Adults, both | MAA | Healthy controls | 76 | Affect Matching/Labeling task | 93.33% |
| et al., 2011) |  |  |  |  |  |  |  |  |
| Payer et al.<br>2012(Payer<br>et al., 2012) | United<br>States | Cross-sectional | Adults, both | MAA | Healthy controls | 100 | Affect Matching/Labeling<br>task | 87.50% |
| Roussotte et<br>al.<br>2011(Rouss<br>otte et al.,<br>2011) | United<br>States | Cross-sectional | Children,<br>both | Prenatal METH exposure | Healthy controls | 50 | Visuo-spatial N-back | 85.71% |
| Roussotte et<br>al.<br>2012(Rouss<br>otte et al., | United<br>States | Cross-sectional | Children,<br>both | Prenatal METH exposure | Healthy controls | 50 | Visuo-spatial N-back | 78.57% |
| 2012) |  |  |  |  |  |  |  |  |
| Salo et al.<br>2009(Salo et al., 2009b) | United States | Cross-sectional | Adults, both | MAA | Healthy controls | 28 | Stroop colour word task (single-trial) | 86.67% |
| Salo et al.<br>2013(Salo et al., 2013) | United States | Cross-sectional | Adults, both | MAA | Healthy controls | 60 | Stroop colour word task (single-trial) | 86.67% |
| Stewart et al.<br>2014(Stewart et al., 2014a) | United States | Cohort | Adults, both | MAA who remained abstinent at 1-year | MAA who relapsed at 1-year | 60 | Paper-Scissors-Rock task | 94.12% |
| Stewart et al. | United States | Cross-sectional | Adults, both | MAA | Healthy controls | 42 | Two-choice prediction task | 100.00% |
| al.<br>2014(Stewart et al., 2014b) | States |  |  |  |  |  | with added breathing load manipulation |  |
| Tabibnia et al.<br>2011(Tabibnia et al., 2011) | United States | Cross-sectional | Adults, both | MAA | Healthy controls | 87 | Stop-Signal task and emotional reappraisal task | 66.67% |
| van Hedger et al.<br>2018(Van Hedger et al., 2018) | Sweden | Single-arm intervention trial | Adults, both | Healthy volunteers who received METH | N/A | 51 | Forced choice preference task | 76.92% |
| van Hedger et al.<br>2019(Van Hedger et al., 2019) | Sweden | RCT (crossover) | Adults, both | Healthy volunteers who received METH | N/A | 18 | Visual activation paradigm<br>— complex images | 90.91% |
| Yin et al.<br>2012(Yin et al., 2012) | China | Cross-sectional | Adults, both | MAA | Healthy controls | 52 | Cue reactivity task | 53.33% |

*ABM: Attentional bias modification training MAA includes a history of current or historical METH abuse, dependence, or use disorder, as diagnosed by the Diagnostic and Statistical Manual of Mental Disorders 4th Edition, Diagnostic and Statistical Manual of Mental Disorders 5th Edition, or International Classification of Diseases 10<sup>th</sup> edition diagnostic codes. Healthy controls defined as participants without adult or prenatal METH exposure. HIV: Human immunodeficiency virus. MAA: Adult METH user. METH: Methamphetamine. MRI: Magnetic resonance imaging. NR: Not reported. N/A: Not applicable. RCT: Randomised controlled trial.*

### METH use overall

METH use has been associated with global cortical changes in grey matter volume (GMV). Nakama et al.(Nakama et al., 2011) reported that heavy METH use was associated with smaller GMV in the dorsolateral prefrontal (dlPFC), prefrontal (PFC), orbitofrontal, and superior temporal cortices and also significantly increased the rate of age-related cortical grey matter loss across the frontal, occipital, and temporal cortices. Kogachi et al.(Kogachi et al., 2017) also found smaller frontal GMV and thinner frontal cortices (particularly in the orbitofrontal cortex) in MAA. The frontal lobes are critical for controlling and coordinating a range of cognitive and motor functions, including memory, executive function, personality, behaviour, and movement.(Firat, 2019; Hoffmann, 2013; Niki et al., 2019) In particular, the PFC is a central hub for multiple higher-order cognitive functions, and is anatomically divided into the dorsolateral, medial, and orbitofrontal components (of which the latter two are often functionally combined into the ventromedial PFC (vmPFC)).(Nejati et al., 2021) Put briefly, while the dlPFC is closely linked to maintaining cognitive control and executive function, including memory, problem-solving, and selective attention, the vmPFC is involved with emotion and reward processing and value-based decision-making.(Nejati et al., 2021; Rolls et al., 2020) Further, the PFC is closely linked to various temporal structures, which themselves play key roles in processing social stimuli and enhancing social cognition.(Barbeau et al., 2020; Olson et al., 2013) Thus, these multi-region reductions in cortical GMV observed in MAA are consistent with other studies that have associated METH use with a range of behavioural and cognitive deficits, including greater impulsivity, emotional dysregulation, and impaired executive function and working memory.(Kogachi et al., 2017; Okita et al., 2016; Sabrini et al., 2019) Regarding white matter changes, Heidari et al.(Heidari et al., 2017) and Thompson et al.(Thompson et al., 2004) found evidence of total white matter hypertrophy, particularly within the occipital and temporal regions, which may reflect inflammatory or compensatory neuroplastic processes secondary to METH-induced neurotoxicity.

METH use has also been associated with altered limbic neuroanatomy. Both Thompson et al.(Thompson et al., 2004) and MacDuffie et al.(MacDuffie et al., 2018) identified reduced GMV, thickness, and density of the cingulate cortex in MAA, with Thompson et al.(Thompson et al., 2004) also identifying reduced GMV in the hippocampus and hypertrophic changes in the surrounding white matter. More specifically, Thompson et al.(Thompson et al., 2004) identified marked GMV reduction in the right, but not the left, cingulate gyrus, MacDuffie et al.(MacDuffie et al., 2018) reported subtle cortical thinning in the posterior cingulate gyrus. However, it is unclear whether these region-specific changes are reflective of METH generating a targeted or lateralised pattern of cingulate atrophy, or whether they would still manifest as bilateral functional deficits. The cingulate cortex is closely involved in regulating emotional overlay for action-outcome associations and providing broader emotional and visuospatial processing,(Rolls, 2019; Trés and Brucki, 2014) while the hippocampus plays a critical role in learning, memory storage and retrieval, and modulating social behaviour.(Anand and Dhikav, 2012; Rubin et al., 2014) Put together, these MRI findings provide a possible neural substrate for previous work identifying complex associations between METH use, depression, and maladaptive interpersonal behaviours,(Hellem et al., 2015; Plüddemann et al., 2010; Tyner and Fremouw, 2008) although further research is required to elucidate the strength, directionality, and potential causality of these relationships. Insular dysfunction, which regulates interoception, cognition, and socio- emotional and sensorimotor processing,(Uddin et al., 2017) has also been implicated as a key contributor towards harmful addiction behaviour.(Droutman et al., 2015) In our review, differing findings were reported as to the effect of METH on insula structure, with Nakama et al.(Nakama et al., 2011) identifying greater age-related insular GMV reduction and Uhlmann et al.(Uhlmann et al., 2016a) demonstrating increased insular thickness in MAA compared to CON. However, as with white matter, this increased thickness may reflect compensatory changes secondary to METH-induced ischaemic or cytotoxic injury.(Uhlmann et al., 2016a)

### Abstinent METH use

Previous studies have suggested that certain cortical and limbic regions may undergo some structural recovery following maintained abstinence from METH use. Morales et al.(Morales et al., 2012) conducted a longitudinal analysis of early abstinence (between the first and fourth weeks) and reported that GMV significantly increased across frontal, temporal, occipital, and insular regions. These findings build upon a previous report by Kim et al.(Kim et al., 2006) who found that long-term abstinence from METH (≥six months) was associated with increased grey matter density in the right middle frontal gyrus and that this correlated with improved executive function.

Interestingly, some data suggests that this structural recovery may be sex-specific; Nie et al.(Nie et al., 2021) demonstrated in a cross-sectional analysis that prolonged duration of abstinence was associated with increased GMV in the hippocampus and orbitofrontal and parietal cortices in males specifically but not females. However, they acknowledged that their female participants had been abstinent for significantly longer than their male counterparts, and may have already undergone structural change prior to study recruitment.

Conversely, some data have suggested that METH-induced neural damage may not present until later stages of abstinence. In a longitudinal assessment of 57 patients, Ruan et al.(Ruan et al., 2018) found that cingulate GMV was similar between CON and MAA at six-months of abstinence but lower at 12-months, suggesting that while the cingulate cortex may be able to compensate against neurotoxic damage for short periods in some patients, these deficits may eventually manifest following prolonged abstinence.

### Prenatal exposure to METH

In our review, four structural MRI studies assessed the effect of prenatal exposure to METH in children or infants. Sowell et al.(Sowell et al., 2010) reported that children exposed to METH had reduced prefrontal, occipital, thalamic and striatal GMV and increased limbic GMV (particularly in the cingulate cortices) than CON, and that lower thalamic and medial occipital GMV was associated with lower intelligence quotient. They hypothesised that these GMV reductions may be secondary to METH-induced neurotoxicity specifically targeting dopamine-rich areas or non- aminergic networks that are richly connected to monoaminergic circuits, and that damage to striatal structures may disturb intellectual maturation and future functioning. Further, their finding of increased limbic GMV may reflect either a compensatory hypertrophy secondary to striatal or thalamic dysfunction, or a pathological paucity of synaptic pruning or myelination. These findings are corroborated by Derauf et al.(Derauf et al., 2012) who also reported evidence of reduced caudate GMV that was associated with subtle attentional deficits.

As with adults, some data has suggested that METH may have sex-specific effects on children; Roos et al.(Roos et al., 2014) reported that, when compared to sex-adjusted CON, METH-exposed boys had higher striatal and diencephalic GMV, whereas METH-exposed girls had lower corpus callosum volume and higher cortical thickness in the cuneus and superior central sulcus. They suggested that the harmful effects of METH on the developing brain may, in some areas, manifest as increased GMV or cortical thickness secondary to disorganised or poorly pruned neural architecture, rather than neuroinflammation as in adults, and that the differential impact on sex may relate to partial neuroprotection conferred by oestrogen.

### Diffusion MRI

dMRI involves examining the diffusion of water molecules through white matter tracts to evaluate tissue structure and axonal organisation.(Soares et al., 2013) Common metrics measured in dMRI include mean diffusivity (MD), axonal diffusivity (AD), and radial diffusivity (RD), commonly referred to as “diffusivity indices”, as well as fractional anisotropy (FA). Generally, reduced FA coupled with higher diffusivity indices (MD, AD, and RD) imply disrupted white matter organisation and compromised fibre integrity, which may indicate demyelination, axonal damage, neuronal degeneration, or overall tissue disruption.(Budde et al., 2009; Soares et al., 2013; White et al., 2008)

In total, 20 studies examined the neural impact of METH exposure using dMRI (**Table 3**). Five (25%) evaluated current and nine (45%) assessed abstinent MAA, with the remaining six (30%) studies examining infants or children who were prenatally exposed to METH. Overall, METH use was associated with global and regional white matter structural deficits, including subcortical structures (e.g. amygdala, hippocampus, putamen), specific white matter tracts (e.g. the corpus callosum), and whole-brain fibre networks.

**Table 2a:**
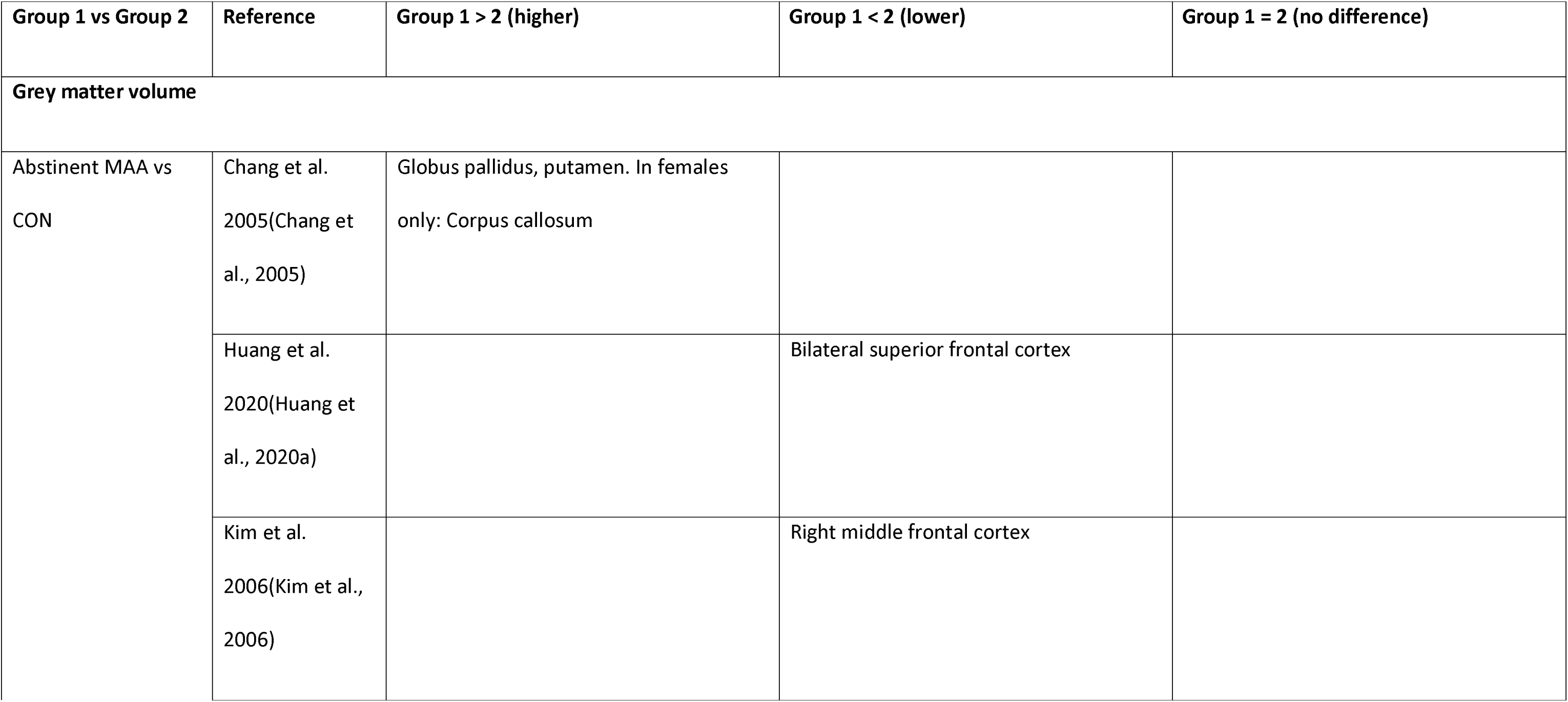
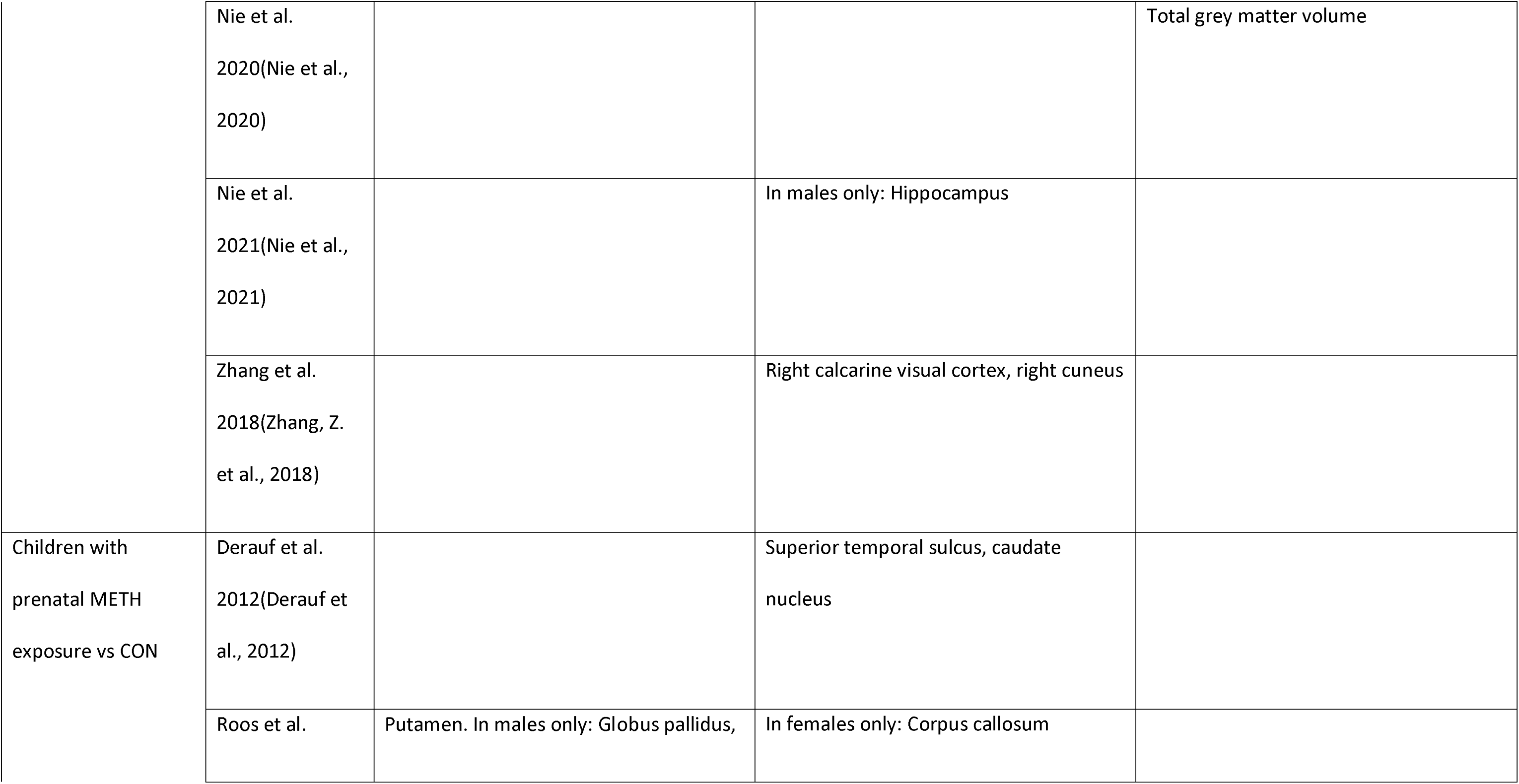
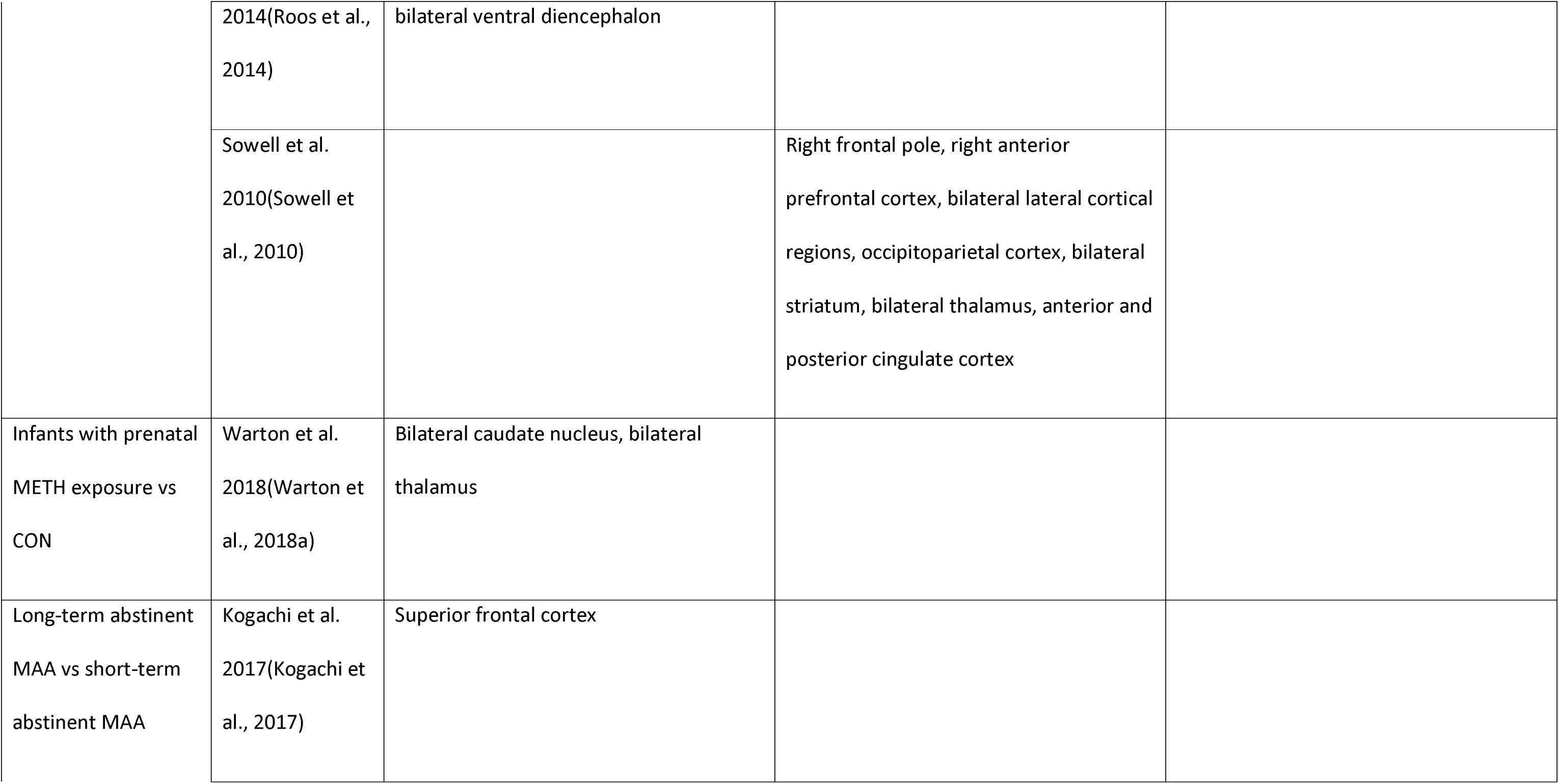
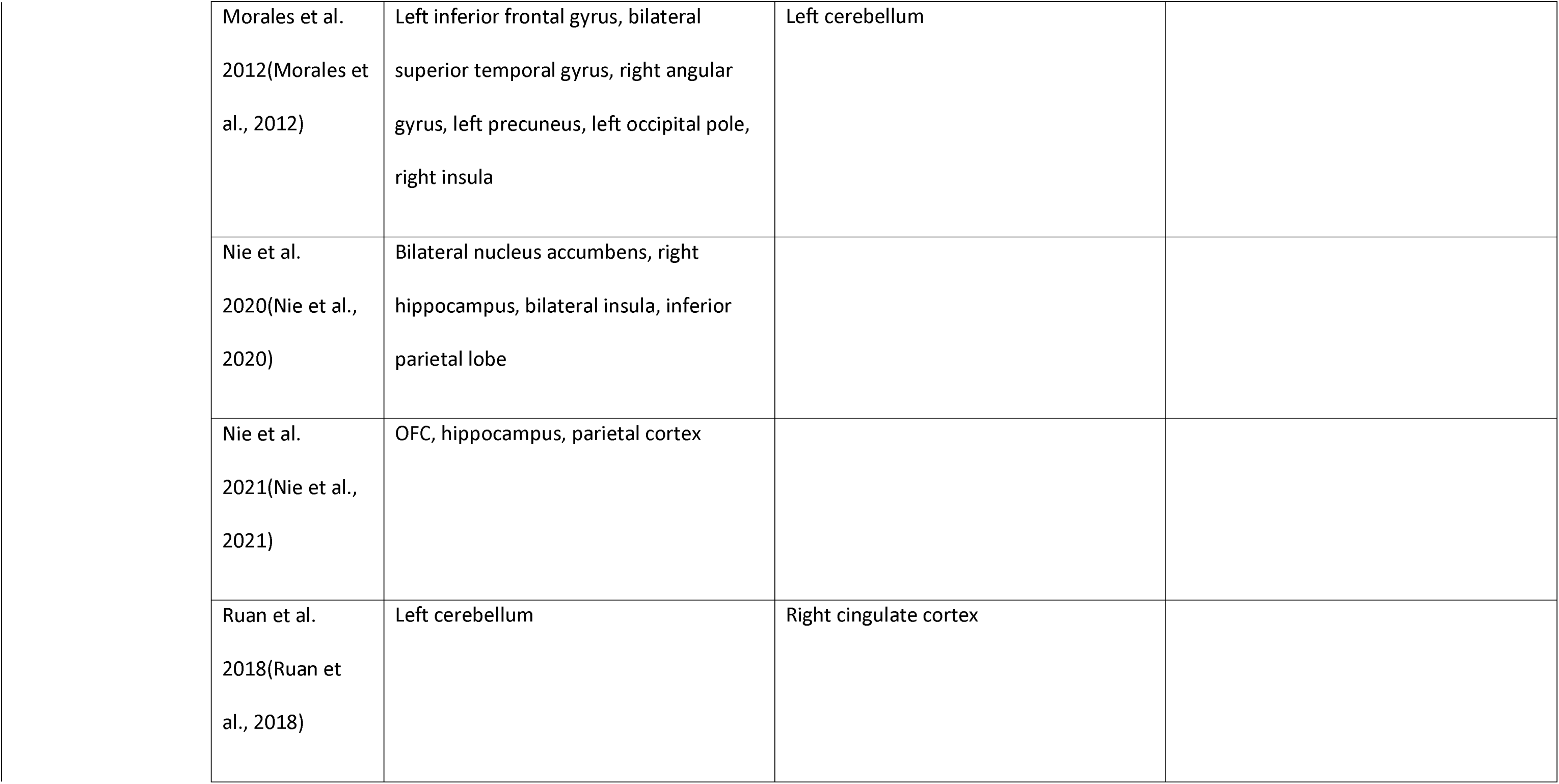
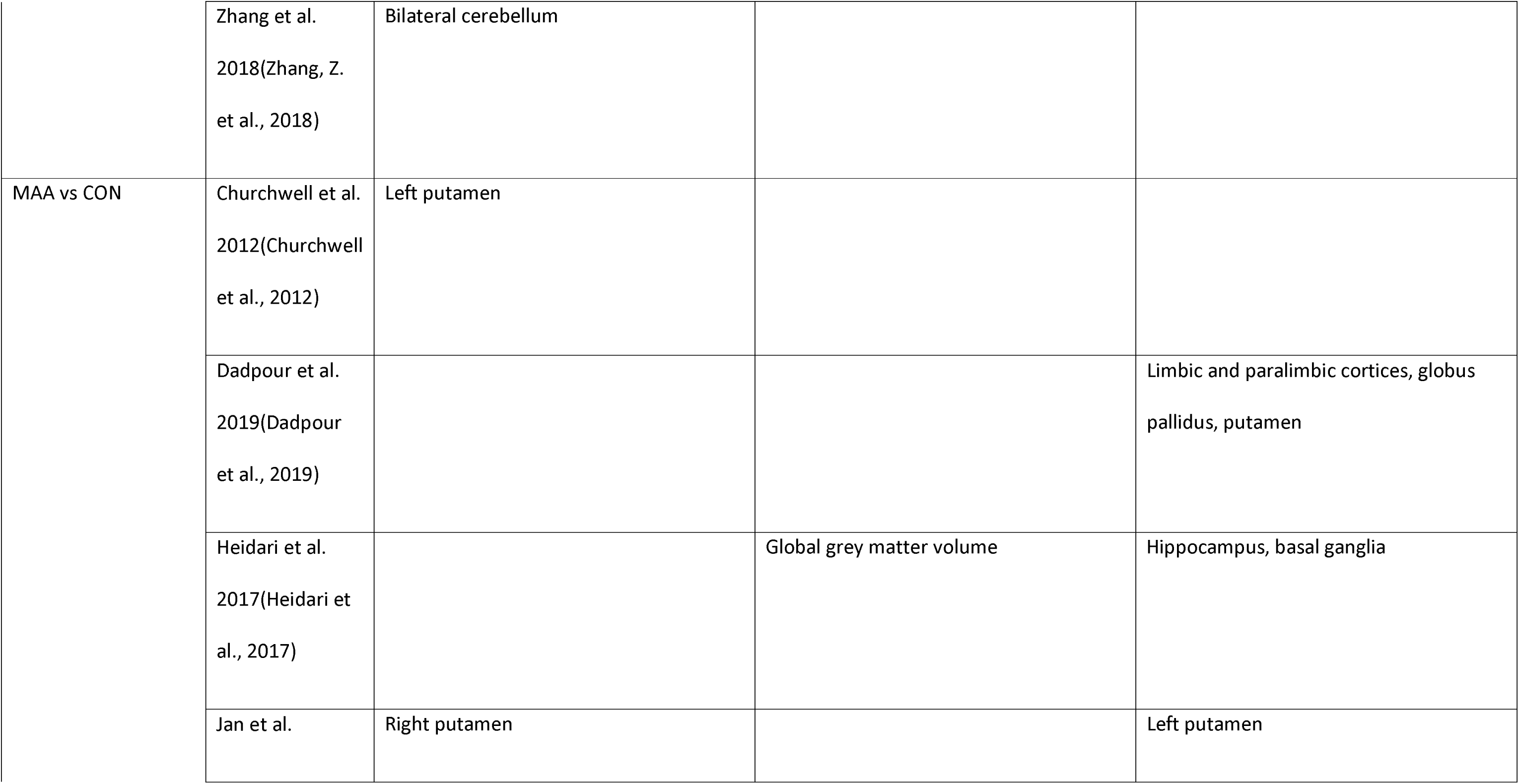
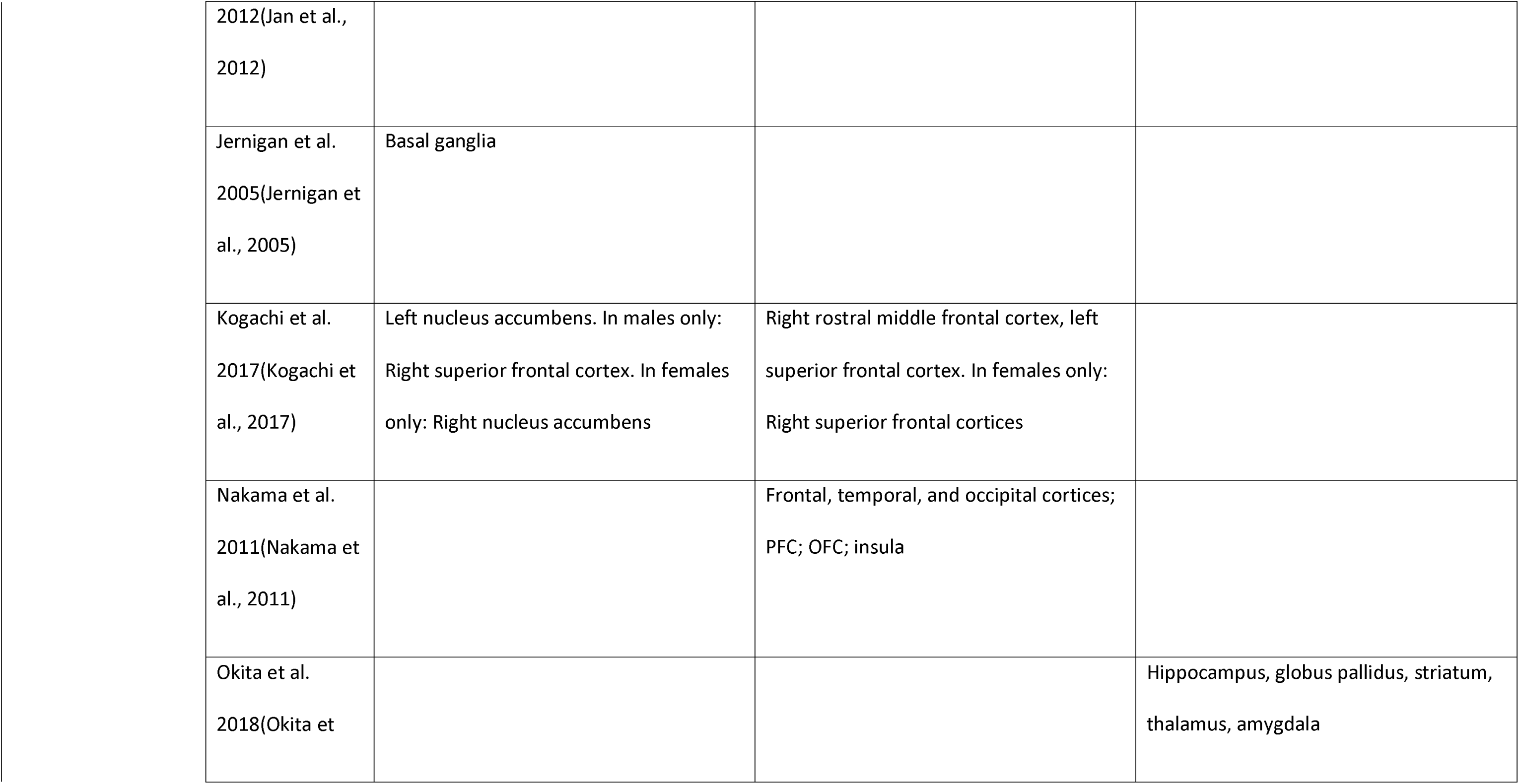
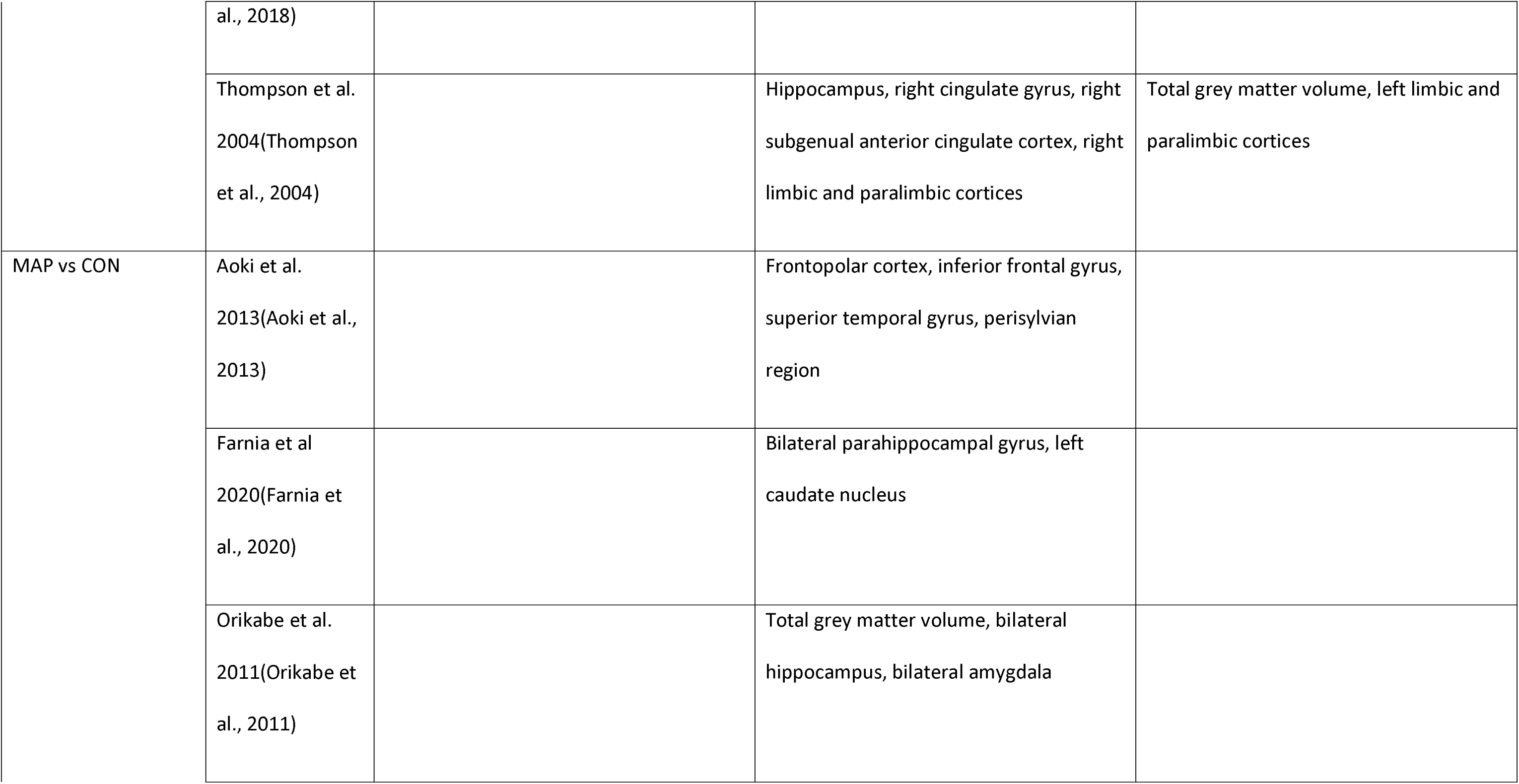
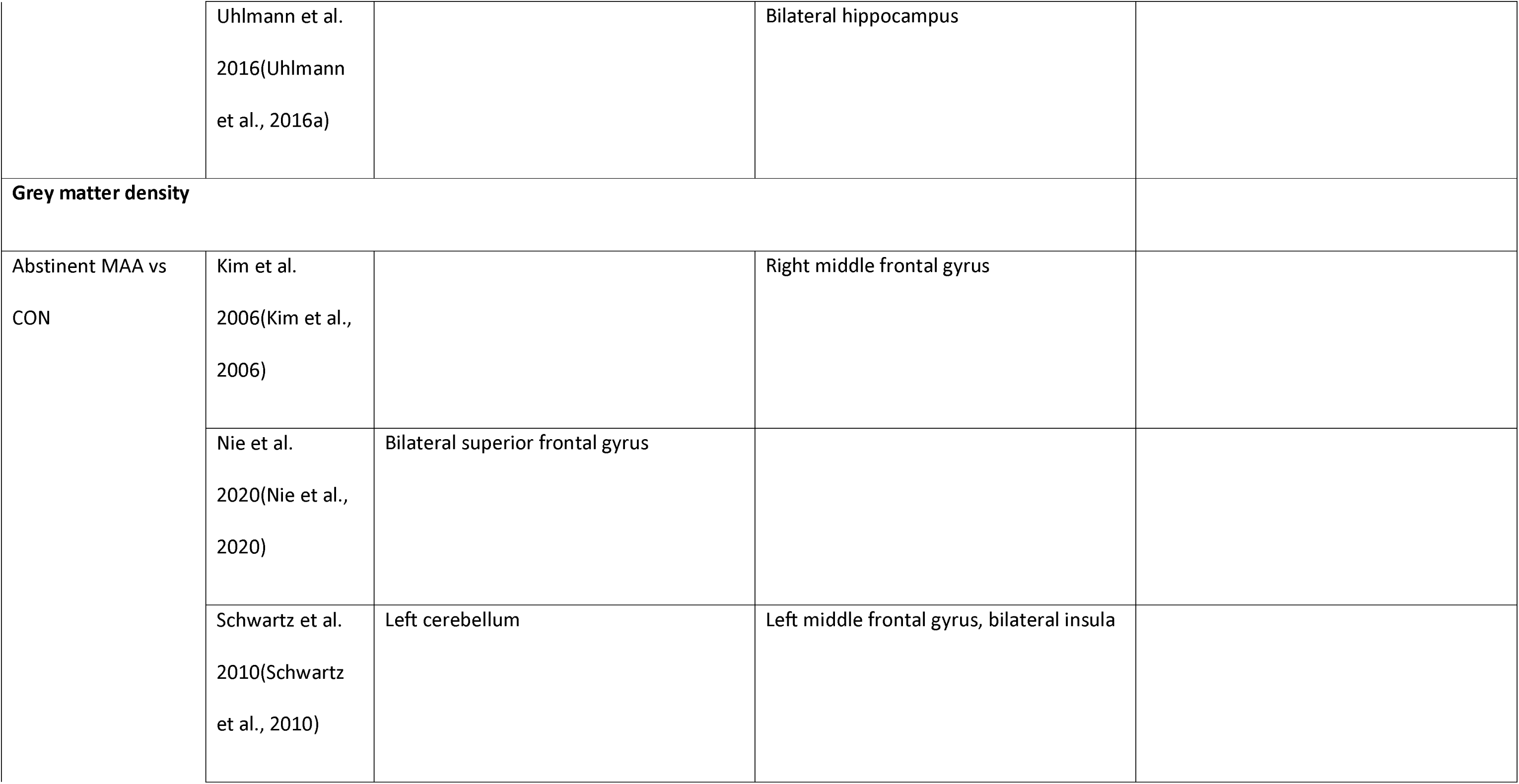
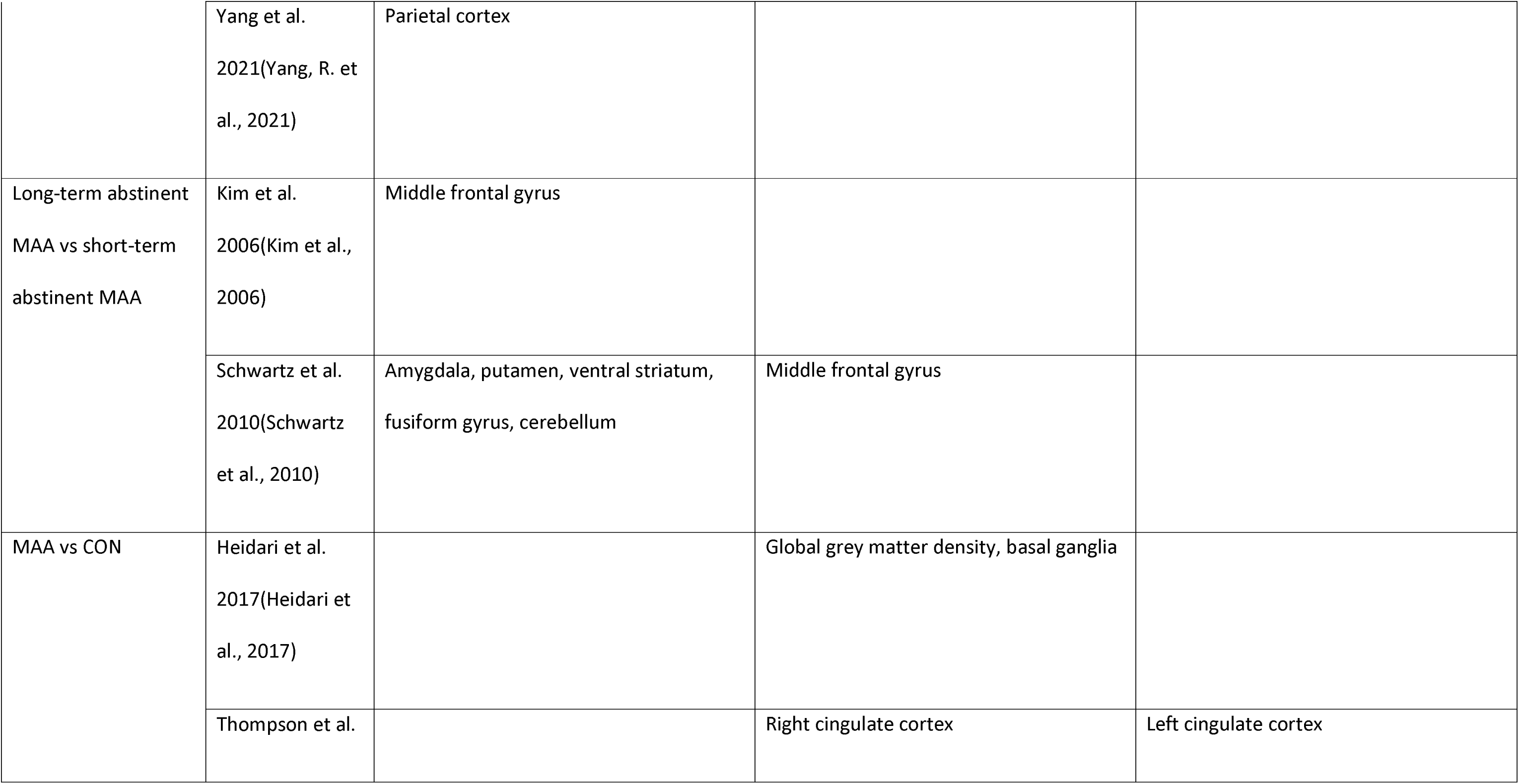
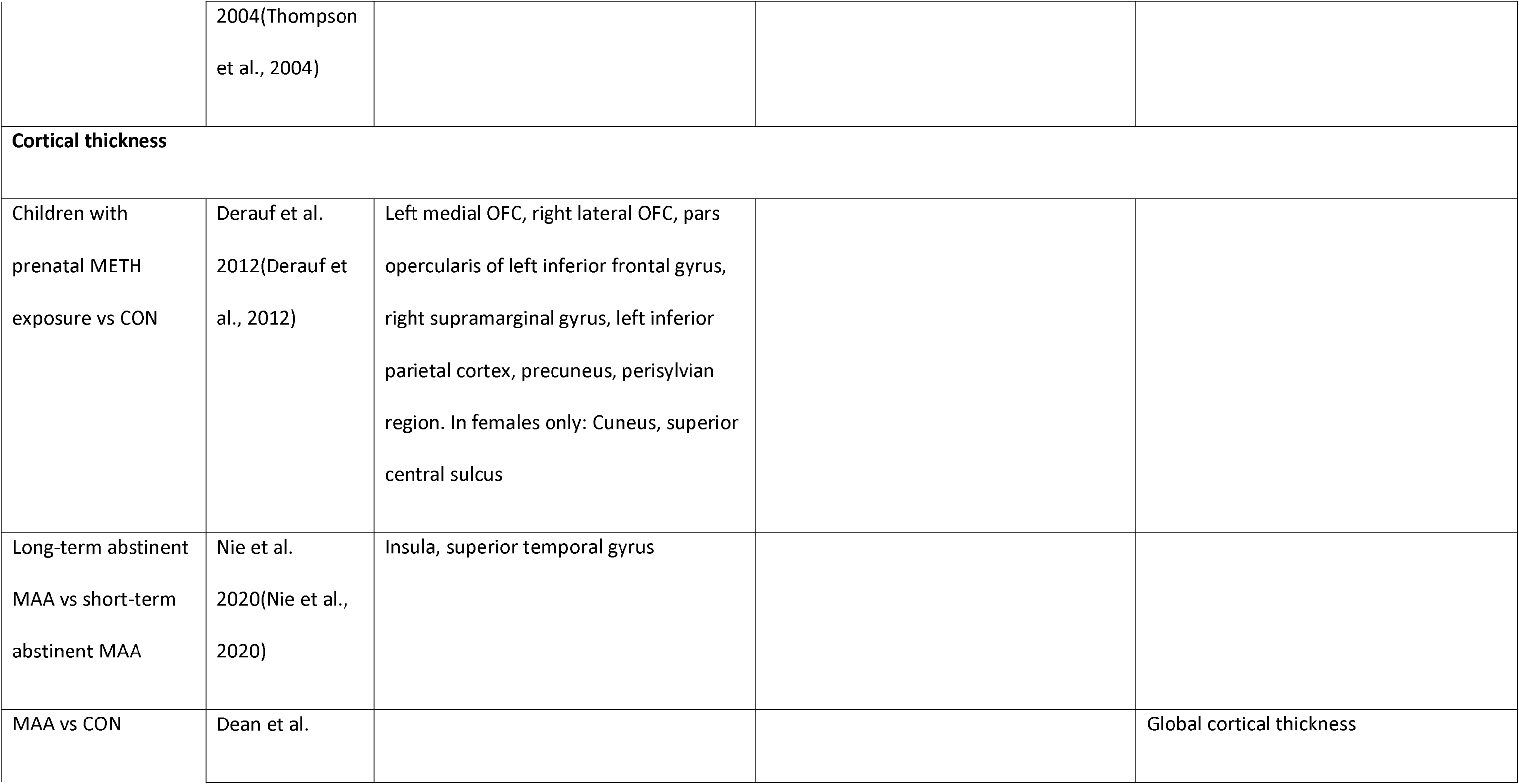
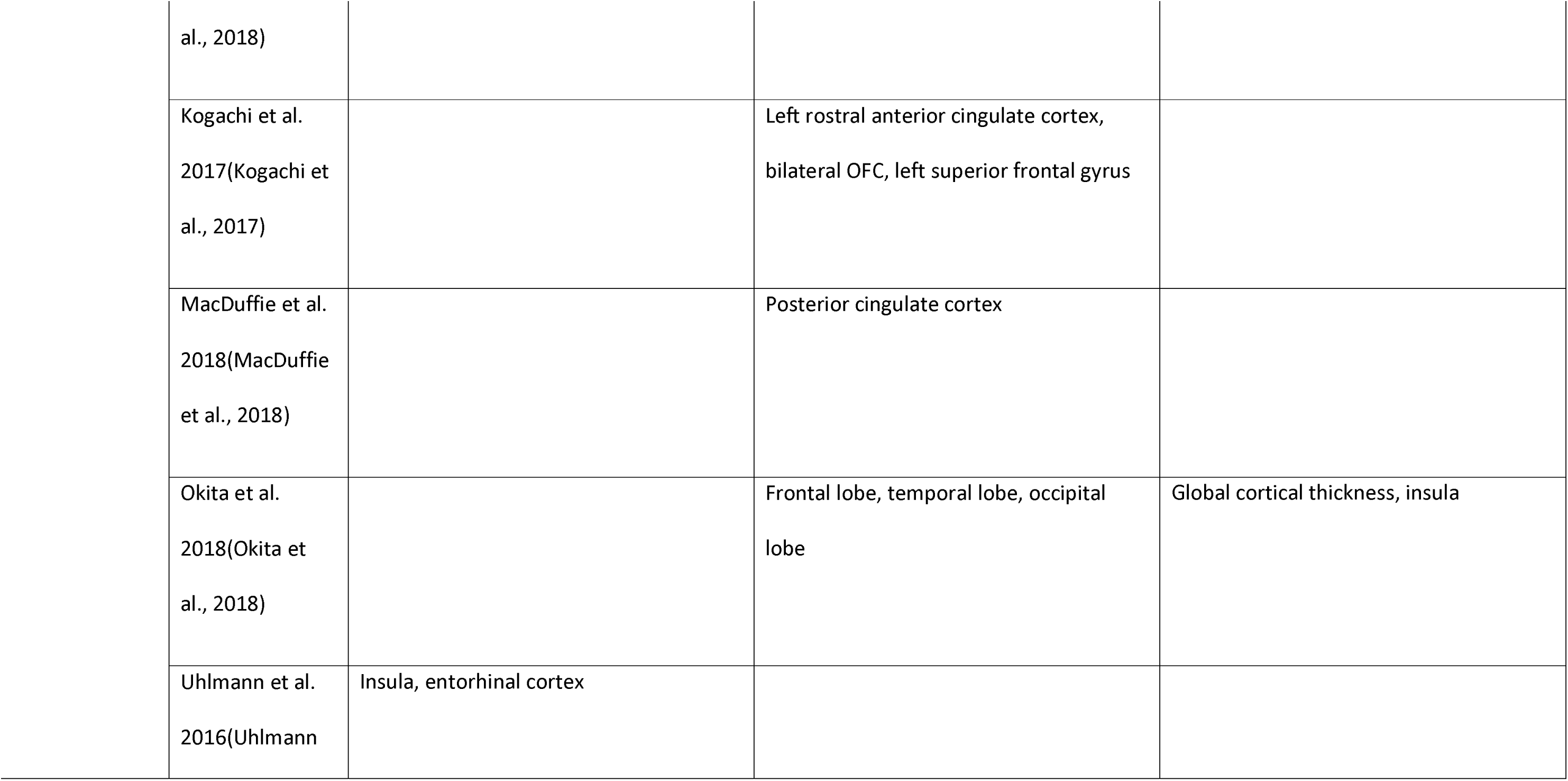
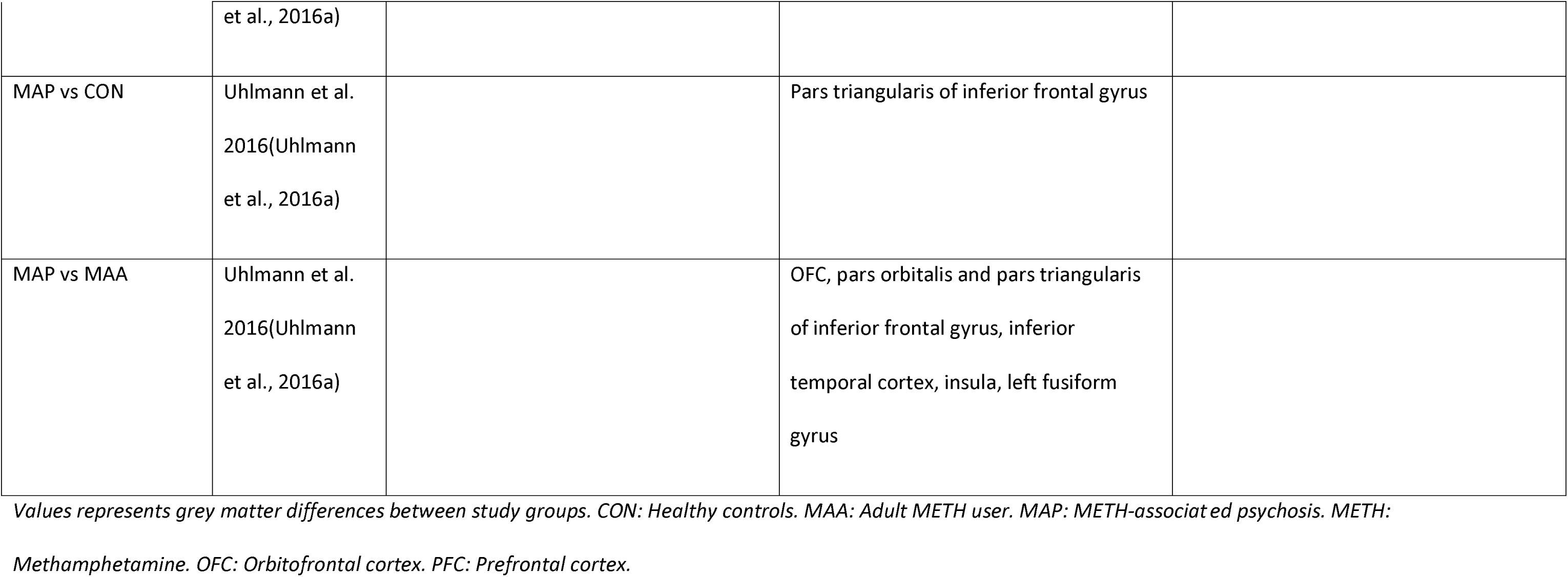
Grey matter changes seen on structural MRI.

**Table 2b:**
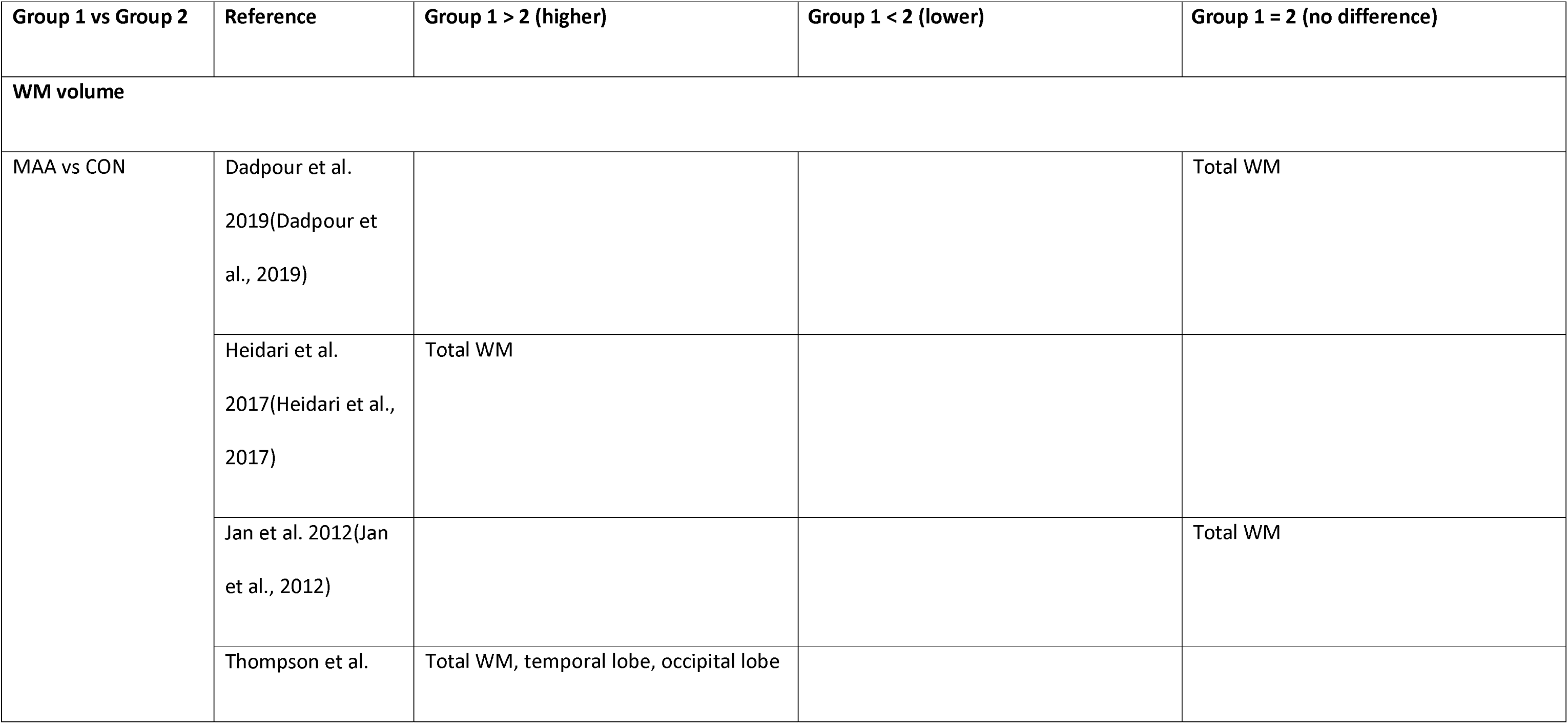
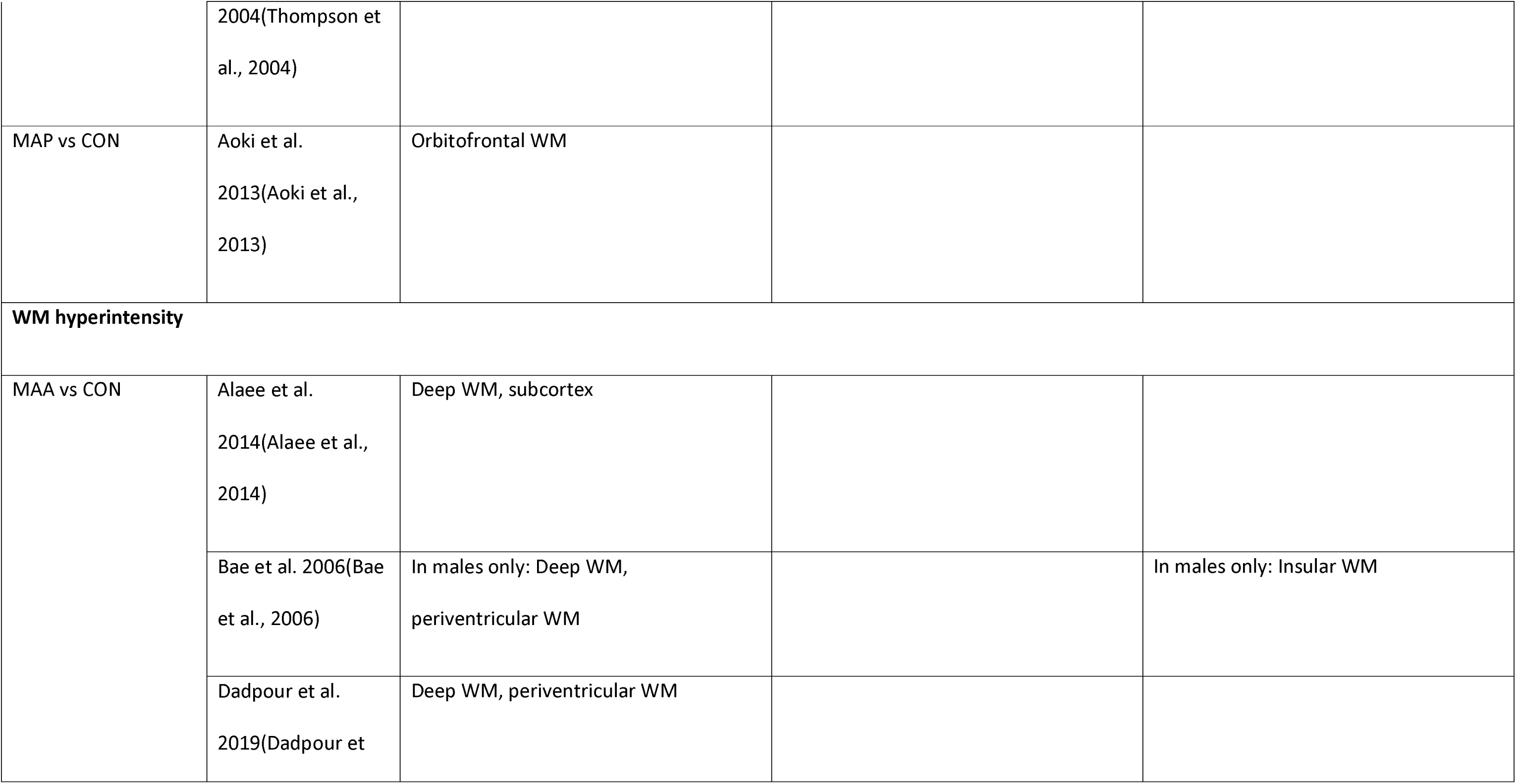
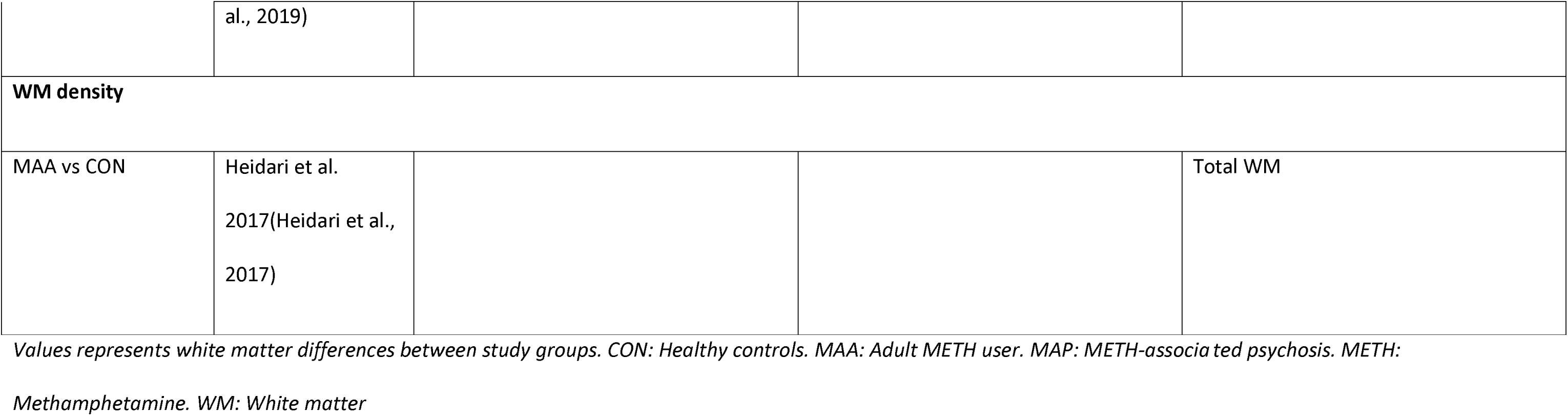
White matter changes as seen on structural MRI.

| Group 1 vs Group 2 | Reference | Group 1 > 2 (higher) | Group 1 < 2 (lower) | Group 1 = 2 (no difference) |
| --- | --- | --- | --- | --- |
| **WM volume** |  |  |  |  |
| MAA vs CON | Dadpour et al.<br>2019(Dadpour et al., 2019) |  |  | Total WM |
|  | Heidari et al.<br>2017(Heidari et al., 2017) | Total WM |  |  |
|  | Jan et al. 2012(Jan et al., 2012) |  |  | Total WM |
|  | Thompson et al. | Total WM, temporal lobe, occipital lobe |  |  |
|  | 2004(Thompson et al., 2004) |  |  |  |
| MAP vs CON | Aoki et al.<br>2013(Aoki et al., 2013) | Orbitofrontal WM |  |  |
| **WM hyperintensity** |  |  |  |  |
| MAA vs CON | Alaee et al.<br>2014(Alaee et al., 2014) | Deep WM, subcortex |  |  |
|  | Bae et al. 2006(Bae et al., 2006) | In males only: Deep WM, periventricular WM |  | In males only: Insular WM |
|  | Dadpour et al.<br>2019(Dadpour et al., 2019) | Deep WM, periventricular WM |  |  |
| **WM density** |  |  |  |  |
| MAA vs CON | Heidari et al.<br>2017(Heidari et al.,<br>2017) |  |  | Total WM |

Values represents white matter differences between study groups. CON: Healthy controls. MAA: Adult METH user. MAP: METH-associated psychosis. METH:
Methamphetamine. WM: White matter

**Table 3:**
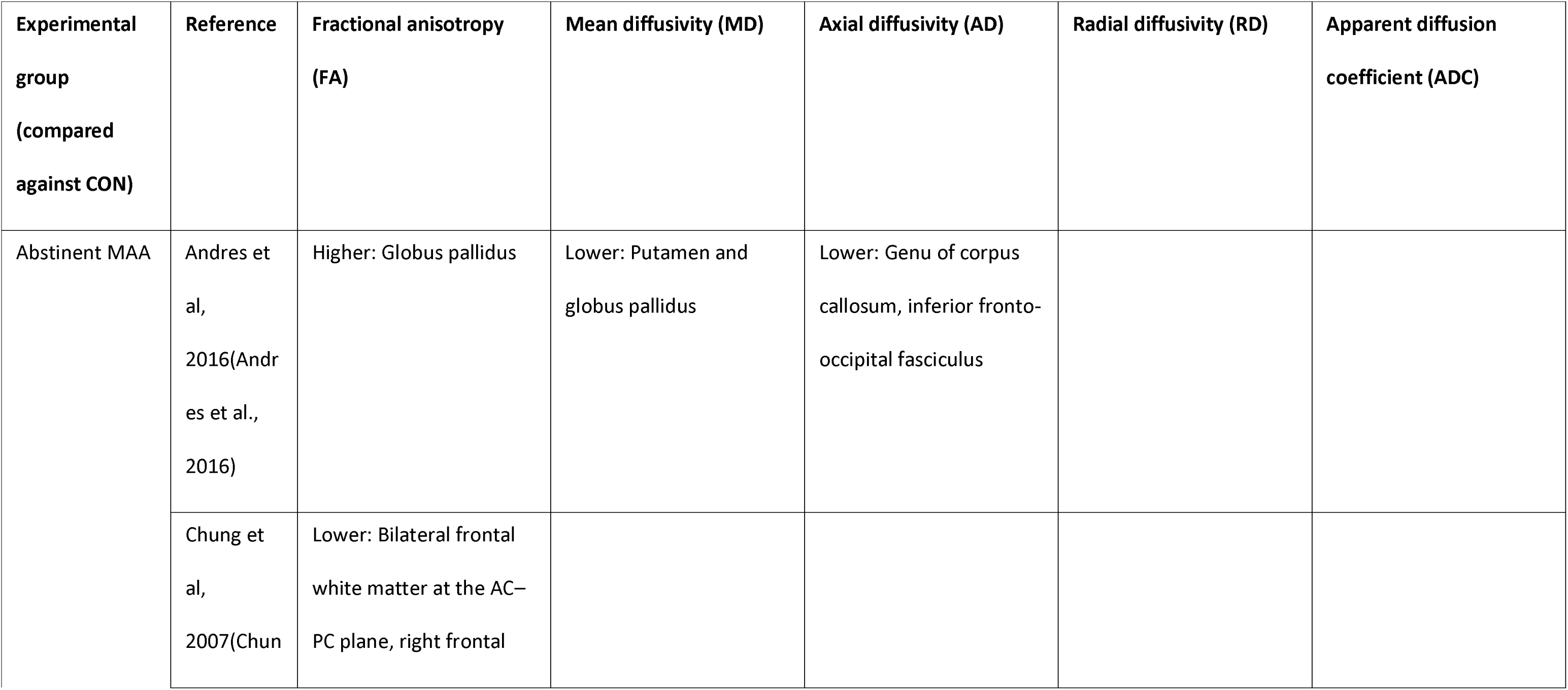
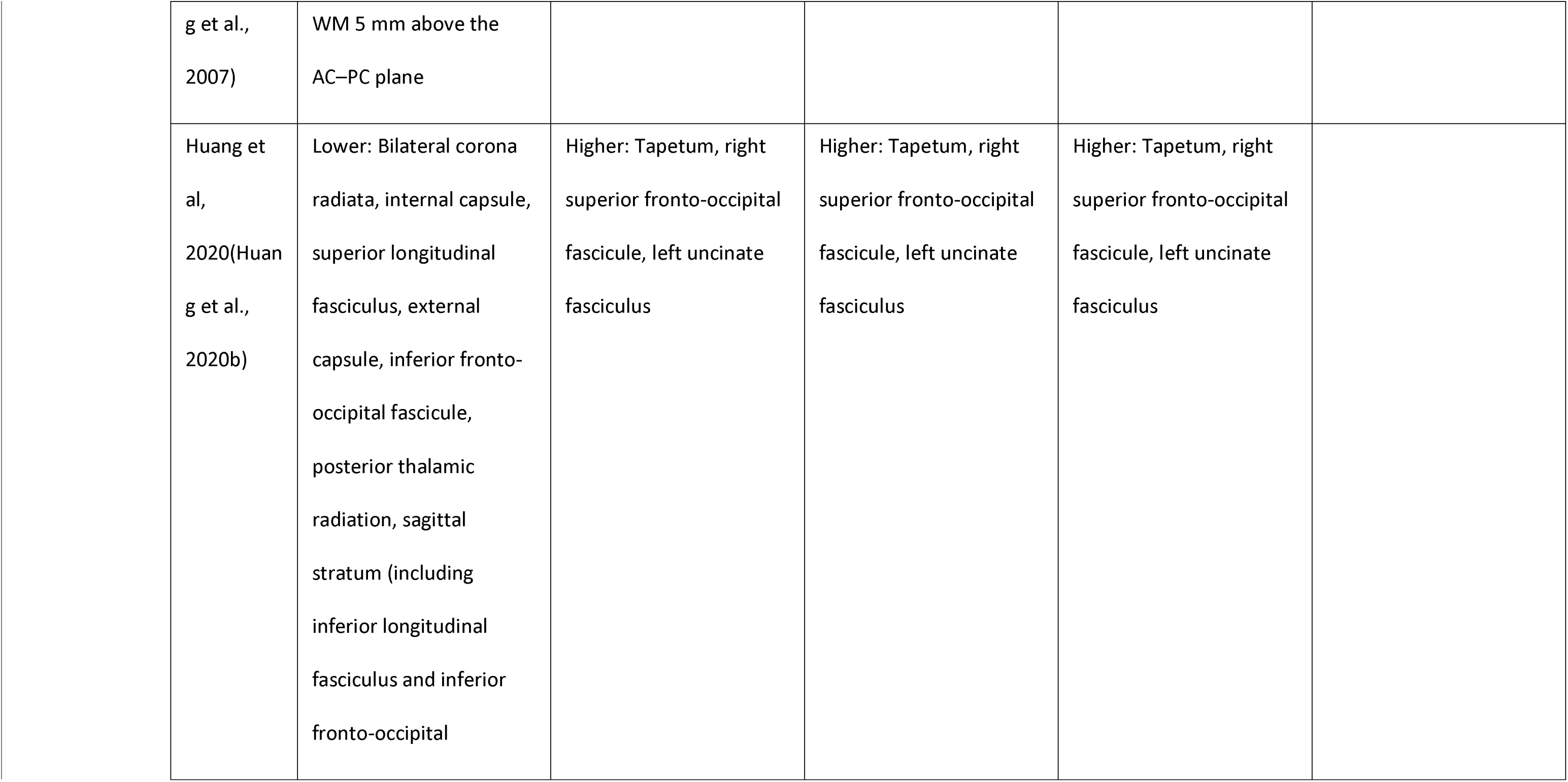
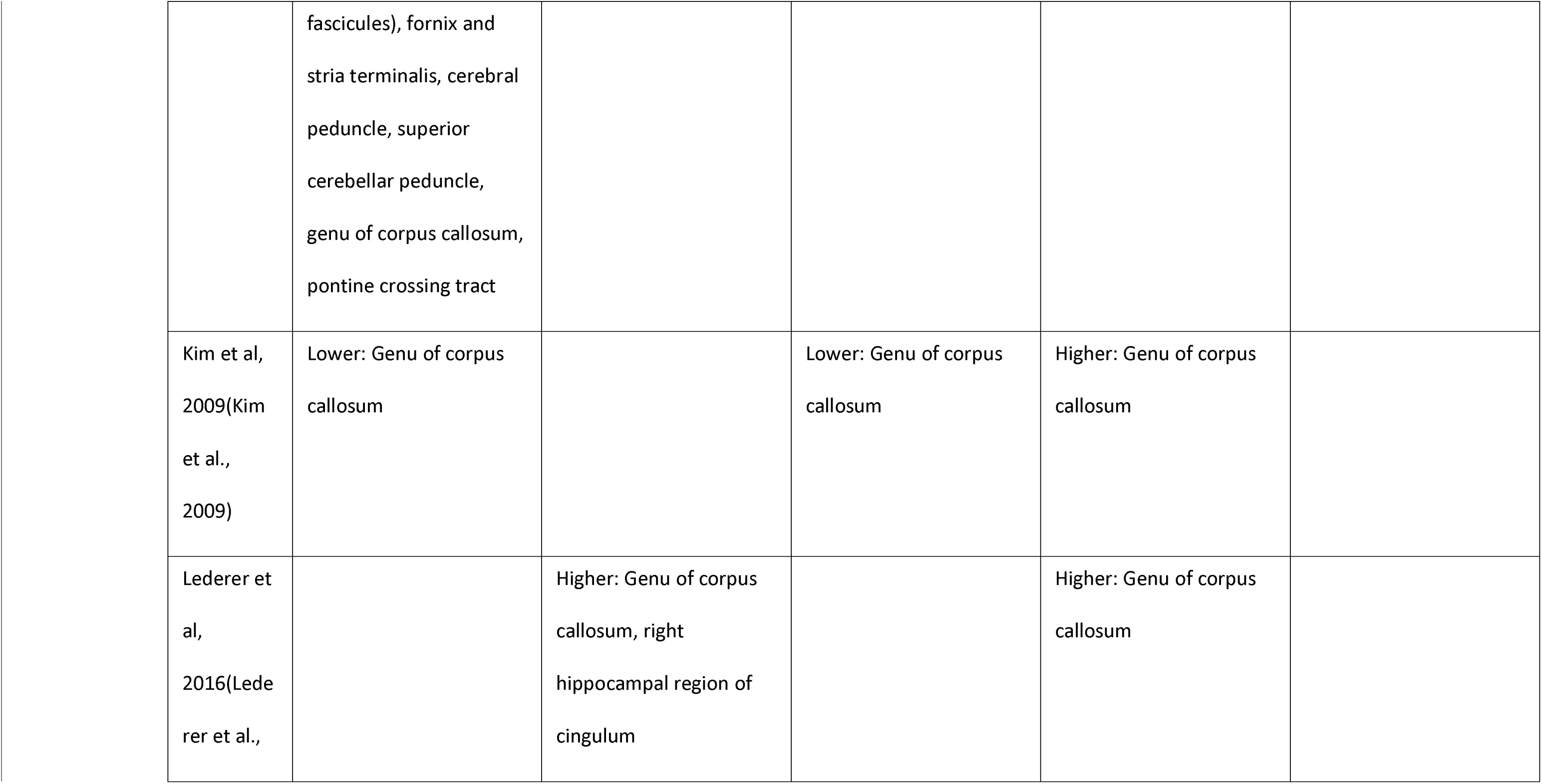
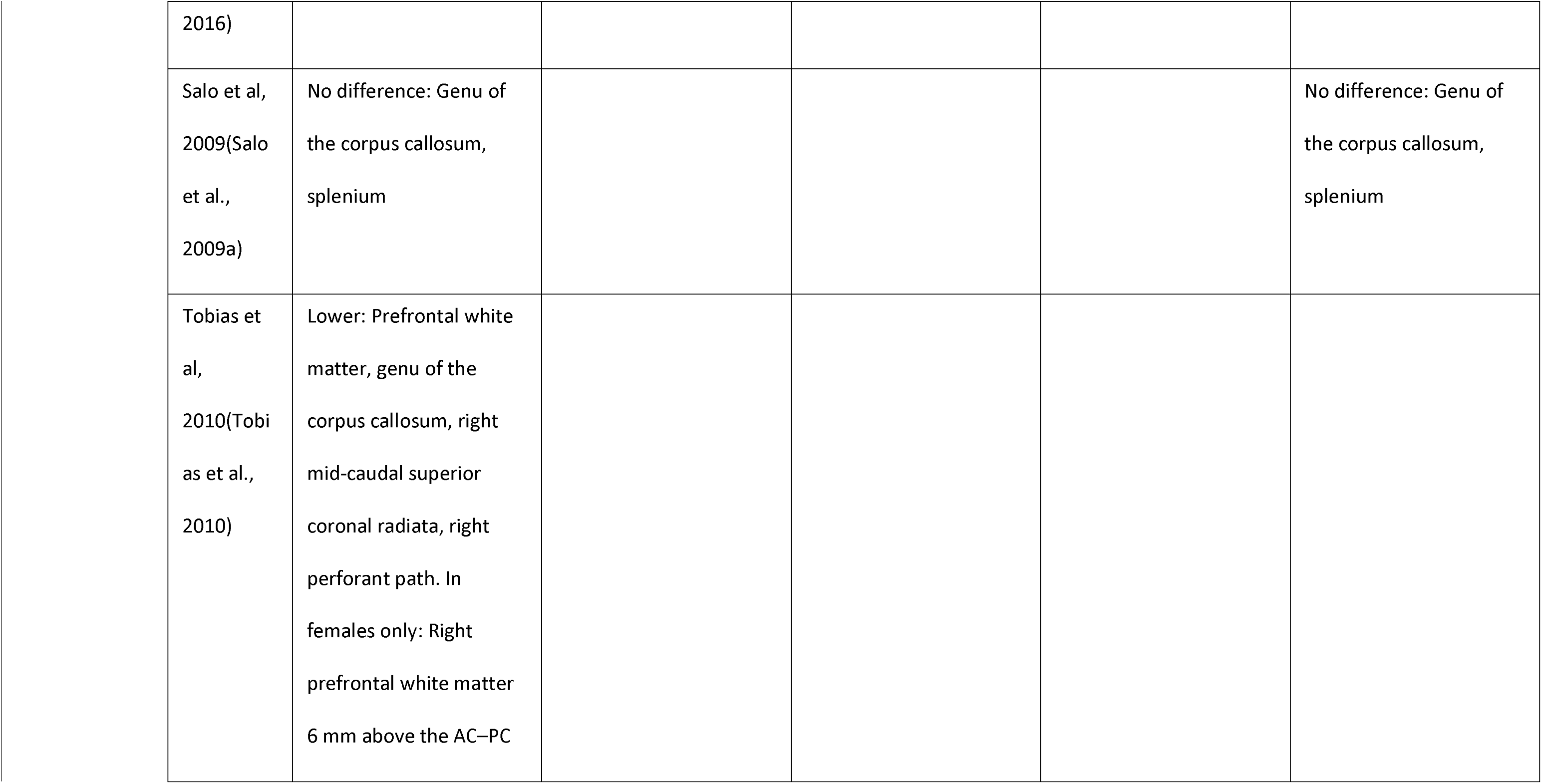
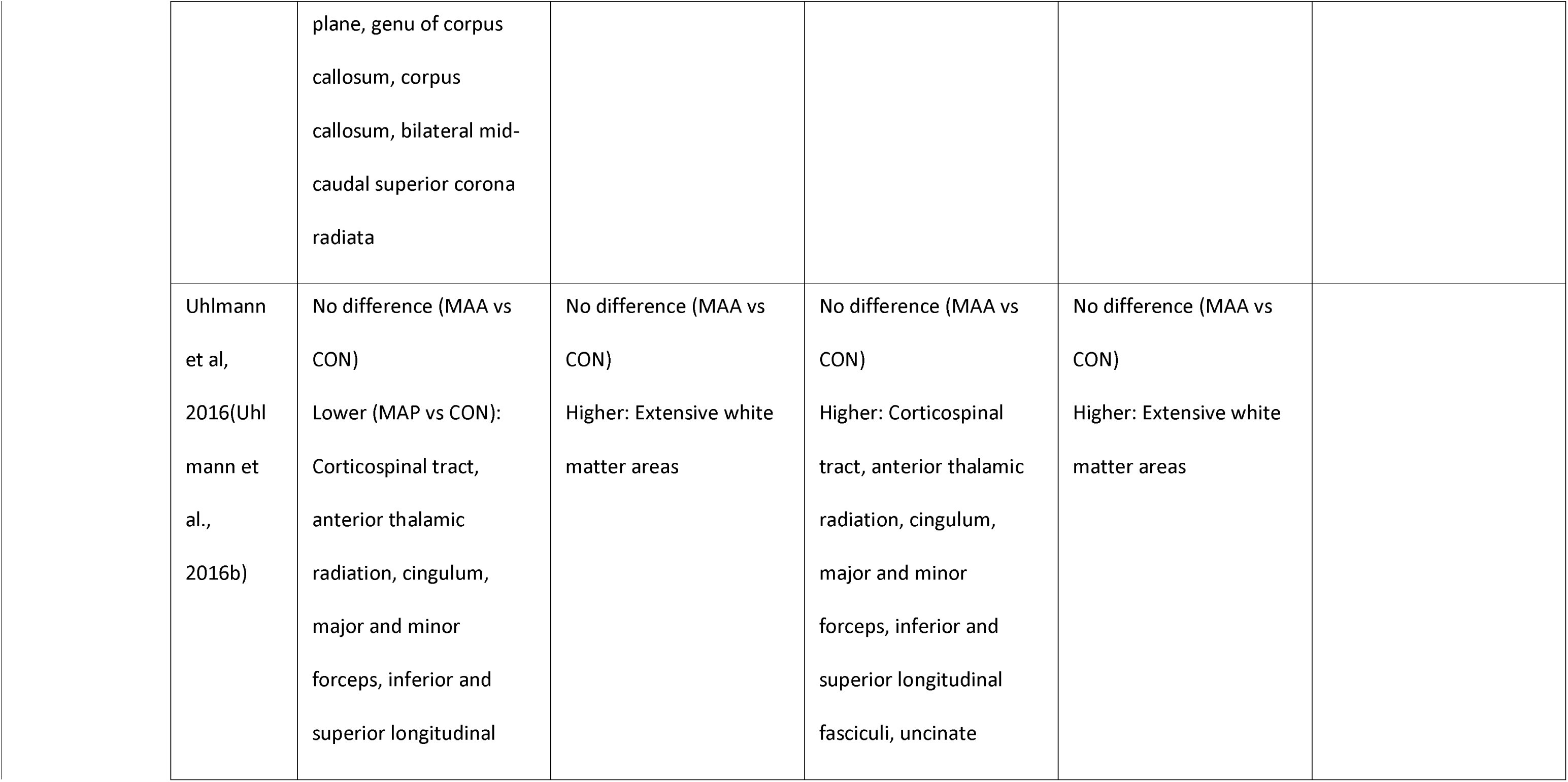
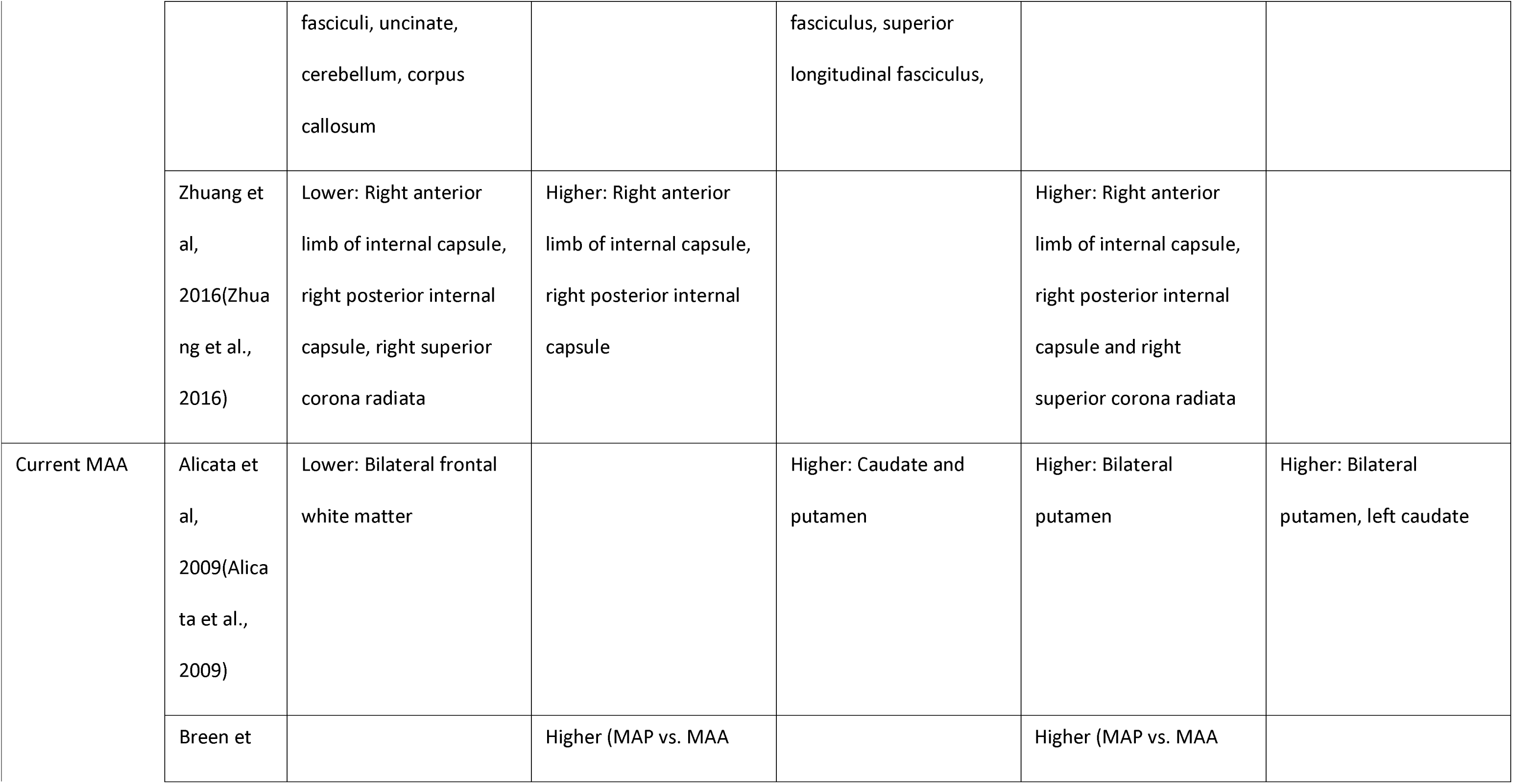
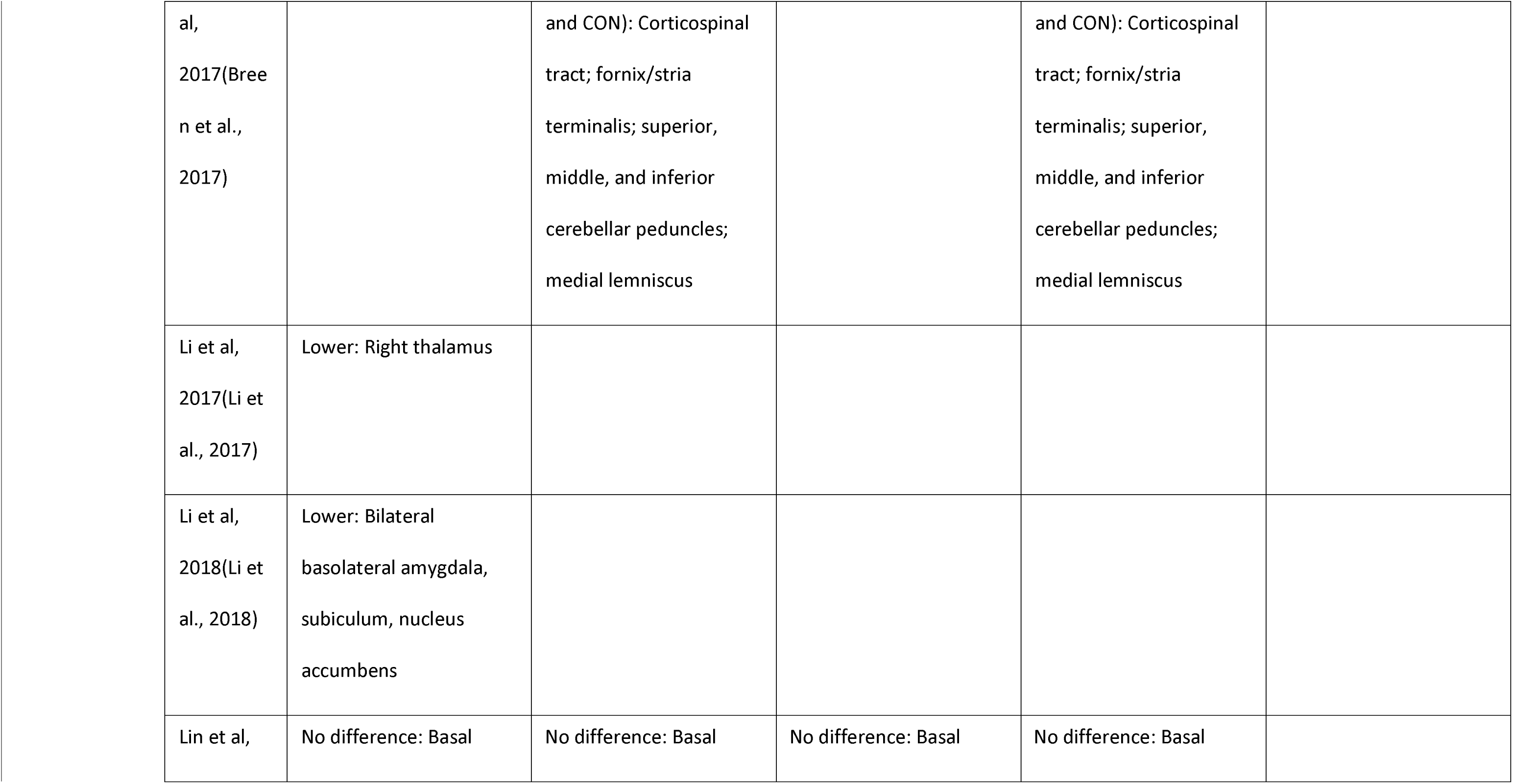
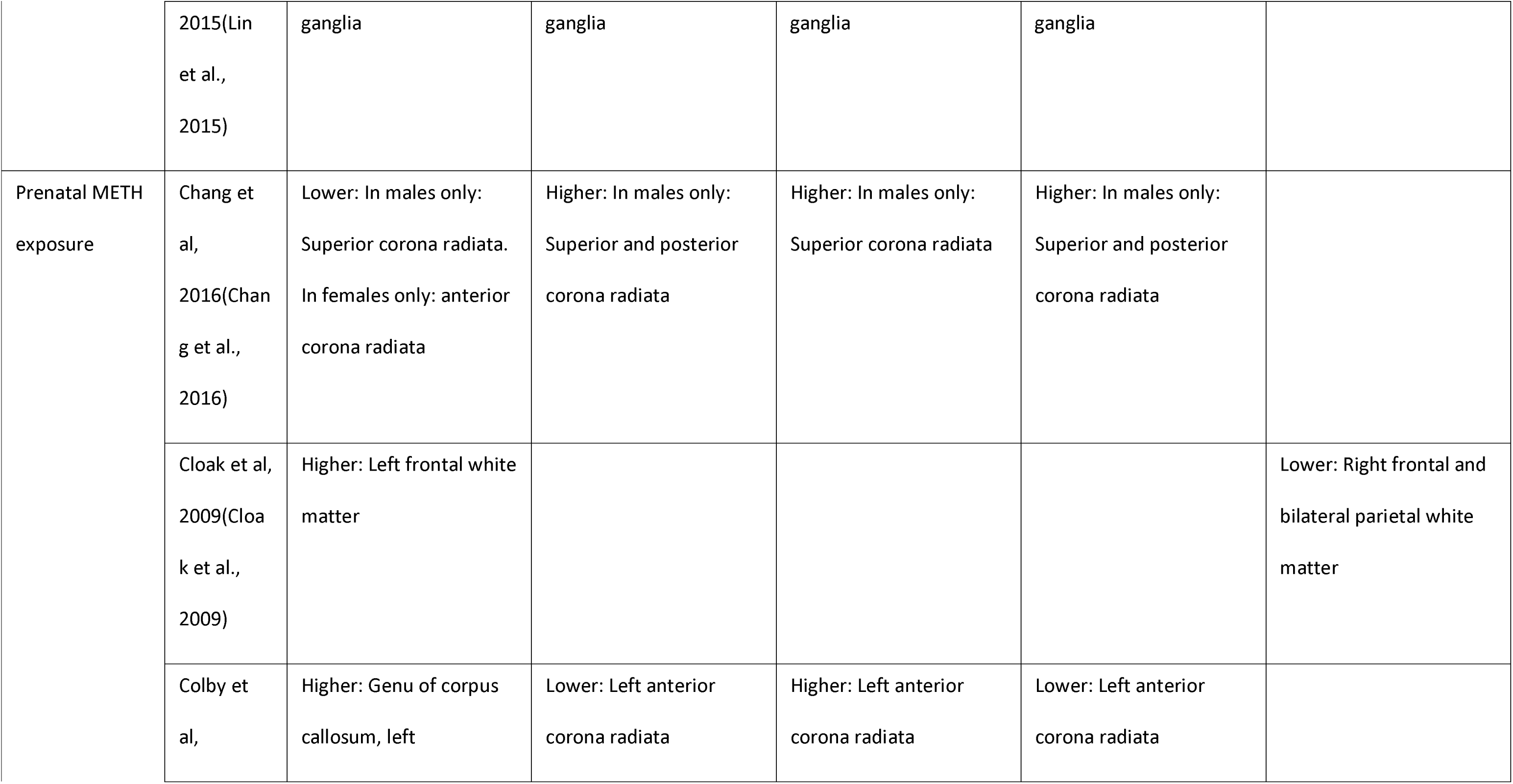
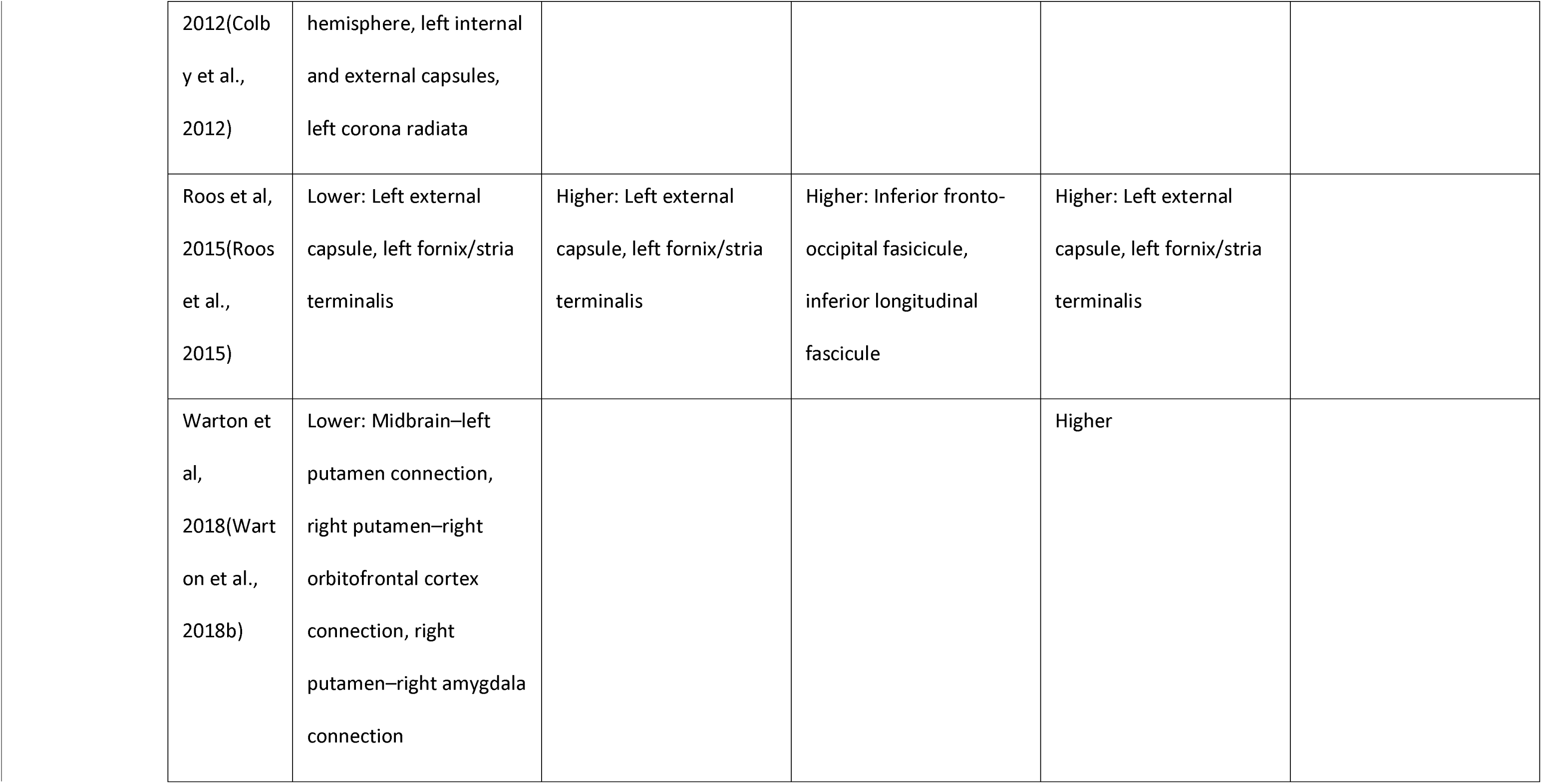
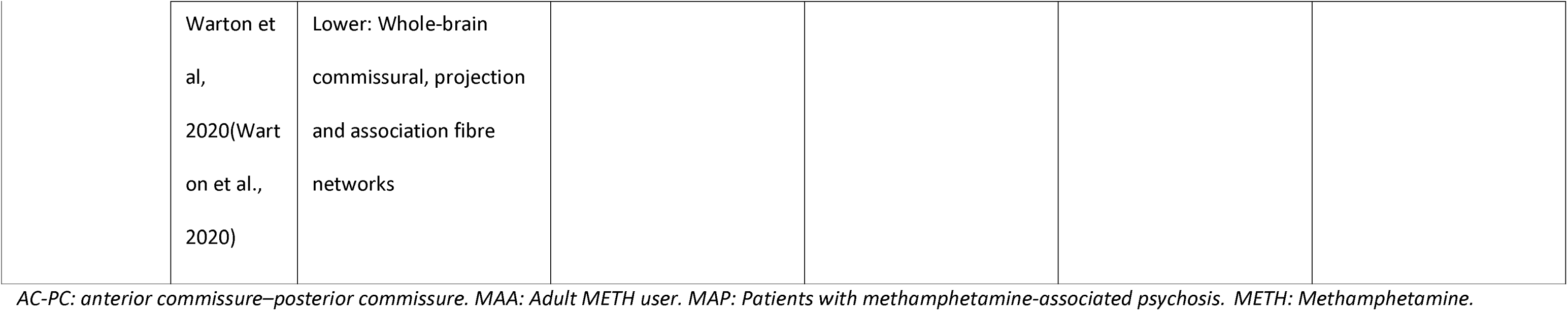
Diffusion MRI findings.

| Experimental group<br>(compared against CON) | Reference | Fractional anisotropy (FA) | Mean diffusivity (MD) | Axial diffusivity (AD) | Radial diffusivity (RD) | Apparent diffusion coefficient (ADC) |
| --- | --- | --- | --- | --- | --- | --- |
| Abstinent MAA | Andres et al, 2016(Andres et al., 2016) | Higher: Globus pallidus | Lower: Putamen and globus pallidus | Lower: Genu of corpus callosum, inferior fronto-occipital fasciculus |  |  |
|  | Chung et al, 2007(Chun | Lower: Bilateral frontal white matter at the AC–PC plane, right frontal |  |  |  |  |

|  |  |  |  |  |
| --- | --- | --- | --- | --- |
| g et al.,<br>2007) | WM 5 mm above the<br>AC–PC plane |  |  |  |
| Huang et<br>al,<br>2020(Huan<br>g et al.,<br>2020b) | Lower: Bilateral corona<br>radiata, internal capsule,<br>superior longitudinal<br>fasciculus, external<br>capsule, inferior fronto-occipital fascicule,<br>posterior thalamic<br>radiation, sagittal<br>stratum (including<br>inferior longitudinal<br>fasciculus and inferior<br>fronto-occipital | Higher: Tapetum, right<br>superior fronto-occipital<br>fascicule, left uncinate<br>fasciculus | Higher: Tapetum, right<br>superior fronto-occipital<br>fascicule, left uncinate<br>fasciculus | Higher: Tapetum, right<br>superior fronto-occipital<br>fascicule, left uncinate<br>fasciculus |

|  |  |  |  |  |
| --- | --- | --- | --- | --- |
|  | fascicules), fornix and<br>stria terminalis, cerebral<br>peduncle, superior<br>cerebellar peduncle,<br>genu of corpus callosum,<br>pontine crossing tract |  |  |  |
| Kim et al,<br>2009(Kim<br>et al.,<br>2009) | Lower: Genu of corpus<br>callosum |  | Lower: Genu of corpus<br>callosum | Higher: Genu of corpus<br>callosum |
| Lederer et<br>al,<br>2016(Lede<br>rer et al., |  | Higher: Genu of corpus<br>callosum, right<br>hippocampal region of<br>cingulum |  | Higher: Genu of corpus<br>callosum |

|  |  |  |  |  |  |
| --- | --- | --- | --- | --- | --- |
| 2016) |  |  |  |  |  |
| Salo et al,<br>2009(Salo<br>et al.,<br>2009a) | No difference: Genu of<br>the corpus callosum,<br>splenium |  |  |  | No difference: Genu of<br>the corpus callosum,<br>splenium |
| Tobias et<br>al,<br>2010(Tobi<br>as et al.,<br>2010) | Lower: Prefrontal white<br>matter, genu of the<br>corpus callosum, right<br>mid-caudal superior<br>coronal radiata, right<br>perforant path. In<br>females only: Right<br>prefrontal white matter<br>6 mm above the AC–PC |  |  |  |  |

|  |  |  |  |  |
| --- | --- | --- | --- | --- |
|  | plane, genu of corpus callosum, corpus callosum, bilateral mid-caudal superior corona radiata |  |  |  |
| Uhlmann et al, 2016(Uhlmann et al., 2016b) | No difference (MAA vs CON)<br>Lower (MAP vs CON):<br>Corticospinal tract, anterior thalamic radiation, cingulum, major and minor forceps, inferior and superior longitudinal | No difference (MAA vs CON)<br>Higher: Extensive white matter areas | No difference (MAA vs CON)<br>Higher: Corticospinal tract, anterior thalamic radiation, cingulum, major and minor forceps, inferior and superior longitudinal fasciculi, uncinate | No difference (MAA vs CON)<br>Higher: Extensive white matter areas |
|  |  | fasciculi, uncinata,<br>cerebellum, corpus<br>callosum |  | fasciculus, superior<br>longitudinal fasciculus, |  |  |
|  | Zhuang et al,<br>2016(Zhuang et al.,<br>2016) | Lower: Right anterior limb of internal capsule, right posterior internal capsule, right superior corona radiata | Higher: Right anterior limb of internal capsule, right posterior internal capsule |  | Higher: Right anterior limb of internal capsule, right posterior internal capsule and right superior corona radiata |  |
| Current MAA | Alicata et al,<br>2009(Alicata et al.,<br>2009) | Lower: Bilateral frontal white matter |  | Higher: Caudate and putamen | Higher: Bilateral putamen | Higher: Bilateral putamen, left caudate |
|  | Breen et |  | Higher (MAP vs. MAA |  | Higher (MAP vs. MAA |  |

|  |  |  |  |  |
| --- | --- | --- | --- | --- |
| al,<br>2017(Bree<br>n et al.,<br>2017) |  | and CON): Corticospinal<br>tract; fornix/stria<br>terminalis; superior,<br>middle, and inferior<br>cerebellar peduncles;<br>medial lemniscus |  | and CON): Corticospinal<br>tract; fornix/stria<br>terminalis; superior,<br>middle, and inferior<br>cerebellar peduncles;<br>medial lemniscus |
| Li et al,<br>2017(Li et<br>al., 2017) | Lower: Right thalamus |  |  |  |
| Li et al,<br>2018(Li et<br>al., 2018) | Lower: Bilateral<br>basolateral amygdala,<br>subiculum, nucleus<br>accumbens |  |  |  |
| Lin et al, | No difference: Basal | No difference: Basal | No difference: Basal | No difference: Basal |
|  | 2015(Lin et al., 2015) | ganglia | ganglia | ganglia | ganglia |  |
| Prenatal METH exposure | Chang et al, 2016(Chang et al., 2016) | Lower: In males only: Superior corona radiata.<br>In females only: anterior corona radiata | Higher: In males only: Superior and posterior corona radiata | Higher: In males only: Superior corona radiata | Higher: In males only: Superior and posterior corona radiata |  |
|  | Cloak et al, 2009(Cloak et al., 2009) | Higher: Left frontal white matter |  |  |  | Lower: Right frontal and bilateral parietal white matter |
|  | Colby et al, | Higher: Genu of corpus callosum, left | Lower: Left anterior corona radiata | Higher: Left anterior corona radiata | Lower: Left anterior corona radiata |  |

|  |  |  |  |  |
| --- | --- | --- | --- | --- |
| 2012(Colby et al., 2012) | hemisphere, left internal and external capsules, left corona radiata |  |  |  |
| Roos et al, 2015(Roos et al., 2015) | Lower: Left external capsule, left fornix/stria terminalis | Higher: Left external capsule, left fornix/stria terminalis | Higher: Inferior fronto-occipital fascicule, inferior longitudinal fascicule | Higher: Left external capsule, left fornix/stria terminalis |
| Warton et al, 2018(Warton et al., 2018b) | Lower: Midbrain–left putamen connection, right putamen–right orbitofrontal cortex connection, right putamen–right amygdala connection |  |  | Higher |

|  |  |  |
| --- | --- | --- |
|  | Warton et al, 2020(Warton et al., 2020) | Lower: Whole-brain commissural, projection and association fibre networks |

AC-PC: anterior commissure–posterior commissure. MAA: Adult METH user. MAP: Patients with methamphetamine-associated psychosis. METH: Methamphetamine.

### METH use overall

Overall, most studies have observed reduced FA and elevated diffusivity values of various subcortical structures in people with current METH use compared with CON (**Table 3**). However, in a smaller case-control analysis of 40 participants (18 METH use and 22 CON), Lin et al.(Lin et al., 2015) also reported no significant differences in diffusion metrics of the basal ganglia between METH use and CON.

With regards to METH consumption metrices and diffusion MRI indices, three studies have found that longer duration, earlier initial age, and greater overall dosage of METH use were associated with decreased FA and AD values(Li et al., 2017; Li et al., 2018) and increased apparent diffusion coefficient and RD(Alicata et al., 2009) across various white and grey matter areas, including the amygdala, hippocampus, nucleus accumbens, orbitofrontal cortex, and putamen. These changes, particularly lower FA, are suggestive of reduced myelination and increased interstitial volumes, which in turn may reflect neuroinflammatory damage and impaired fibre coherence to white matter tracts.(Alicata et al., 2009; Pfefferbaum et al., 2010)

### Abstinent METH users

Studies that have investigated the impact of METH in abstinent MAA using dMRI have typically explored two distinct periods: early abstinence (under three months) and long-term abstinence (over one year). This has predominantly been done using case-control studies. Overall, most studies have observed significantly lower FA(Chung et al., 2007; Huang et al., 2020b; Kim et al., 2009; Tobias et al., 2010; Zhuang et al., 2016) and higher RD and MD(Huang et al., 2020b; Kim et al., 2009; Zhuang et al., 2016) values in various white matter areas among abstinent MAA compared against CON, although conflicting findings have been reported for AD. In particular, lower FA and higher diffusivity in the genu of the corpus callosum has been consistently reported across studies,(Huang et al., 2020b; Lederer et al., 2016; Tobias et al., 2010) which may reflect impairment in axonal structure or myelination. Given the critical role of the corpus callosum in connecting the bilateral prefrontal cortices, structural irregularities in this region may contribute towards patterns of maladaptive behaviour and poor decision-making seen in MAA, with previous research linking corpus callosal damage to reduced cognitive function.(Freeze et al., 2022; Hinkley et al., 2012; Paul, 2011) In contrast, grey matter lesions associated with METH exhibit opposing diffusivity changes to white matter, with Andres et al. reporting increased striatal FA and lower MD in past MAA when compared to current MAA or CON. They proposed that these diffusivity alterations may relate to increased iron deposition, which can heighten oxidative stress and activate toxic inflammatory and ferroptotic processes, similar to changes seen in neurodegenerative conditions.(Ward et al., 2014)

Duration of abstinence from METH appears to exert an inconsistent effect on diffusion indices between METH use and CON. For example, in a longitudinal study that followed abstinent METH use for up to one-year, Zhuang et al.(Zhuang et al., 2016) found that people with METH use had lower FA in the right internal capsule and superior corona radiata that persisted at both six-months and one-year abstinence, although there was no notable correlation between duration of abstinence and FA. In contrast, Uhlmann et al.(Uhlmann et al., 2016b) reported that duration of abstinence (from three-six weeks) was positively correlated with FA in several major white matter tracts, including the corticospinal tract, posterior corona radiata, cingulum, and uncinate and longitudinal fasiculi.

Two studies have evaluated sex-specific differences in females between duration of METH abstinence and diffusion indices. Tobias et al.(Tobias et al., 2010) reported lower FA in multiple white matter regions in female early abstinent METH use only when compared to CON, including the corpus callosum, corona radiata, and genu. In contrast, Chung et al.(Chung et al., 2007) found no variation in FA between female abstinent METH use and CON.

### Prenatal exposure to METH

Six studies have examined diffusion MRI changes associated with prenatal METH exposure; of these, three have focused on infants(Chang et al., 2016; Warton et al., 2018b; Warton et al., 2020) and three on children (aged three to 12 years).(Cloak et al., 2009; Colby et al., 2012; Roos et al., 2015) In infants, prenatal METH exposure has been consistently associated with global changes in various white matter pathways.(Chang et al., 2016; Warton et al., 2018b; Warton et al., 2020) More specifically, compared to CON, infants who are prenatally exposed to METH have lower FA and higher diffusivity values within the superior and posterior corona radiatae; white matter tracts within the striatal-orbitofrontal circuit; and whole-brain commissural, projection, and association fibre networks.

In a longitudinal analysis of 132 infants, Chang et al.(Chang et al., 2016) observed gender-specific white matter changes among infants exposed to METH and nicotine. More specifically, boys who were prenatally exposed to METH exhibited lower FA and/or higher diffusivity in the superior and posterior corona radiata at baseline (mean age 43 weeks) than boys who were not exposed, and girls who were prenatally exposed to METH had lower FA in the anterior corona radiata. In both sexes, prenatal exposure to METH was associated with poorer muscle and neurological function. However, while these functional deficits and structural changes had resolved by 3 months follow-up, the trajectory of FA and diffusivity normalisation was steeper in boys who were prenatally exposed to METH and tobacco than boys who were not (i.e. tobacco-only or non-exposed controls), whereas girls who were prenatally exposed to METH experienced persistently lower FA trajectories. Put together, these findings suggest that infants who were prenatally exposed to METH have less robust neural connections and reduced myelination at birth, leading to delayed maturation of white matter tracts and associated functional deficits, although these changes may normalise in a sex-specific manner following cessation of METH exposure.

In contrast, studies conducted on children have disagreed as to the impact of prenatal METH exposure. While Colby et al.(Colby et al., 2012) and Cloak et al.(Cloak et al., 2009) both reported increased FA values in the frontal white matter, corpus callosum, and internal capsule in children who were prenatally exposed to METH compared to children who were not, Roos et al.(Roos et al., 2015) observed decreased FA values in the internal capsule, fornix, and stria terminalis. Further, while Colby et al.(Colby et al., 2012) reported reduced MD and RD in the left corona radiata, Roos et al.(Roos et al., 2015) found increased MD, AD, and RD in the external capsule, fornix/stria terminalis, and sagittal stratum (specifically, the inferior fronto-occipital and inferior longitudinal fasiculi). However, these conflicting findings may relate to methodological differences in the age range of included participants or the analysis methods by which dMRI diffusivity indices were calculated across studies.(Roos et al., 2015)

Thus, it is possible that prenatal METH exposure may exert its neurotoxic effects by increasing the complexity of white matter neuronal structures and impairing synaptic pruning in children,(Cloak et al., 2009; Colby et al., 2012; Roos et al., 2015) leading to greater axonal density that may manifest in dMRI as decreased diffusivity with or without significant FA alterations.(Cloak et al., 2009; Colby et al., 2012) Indeed, previous research conducted on children with prenatal exposure to cocaine and/or alcohol have reported similar abnormalities in synaptic pruning and organisation which may disrupt neuronal transmission and neuroplasticity and negatively impact future functioning.(Basavarajappa and Subbanna, 2023; Lumeng et al., 2007)

### Resting-state fMRI

In rs-fMRI, the blood oxygen dependent (BOLD) signal is measured in the absence of stimuli as a means of quantifying neural activity at rest, which can then be interrogated to further characterise regional brain function and identify multi-region neural networks.(Lv et al., 2018) A variety of rs-fMRI analysis techniques have been used in the literature, including ICA, seed-based FC, EC, ReHo, and ALFF.

In our review, seed-based FC was the most common analysis technique used (11/26, 42%) followed by ICA (5/26, 19%). Overall, rs-fMRI demonstrated that METH use is associated with impaired function across a variety of local brain regions and global neural networks, most commonly the default mode network (DMN), salience network (SN), and central executive network (CEN). Further details about rs-fMRI findings are available in **Table 4**.

**Table 4:**
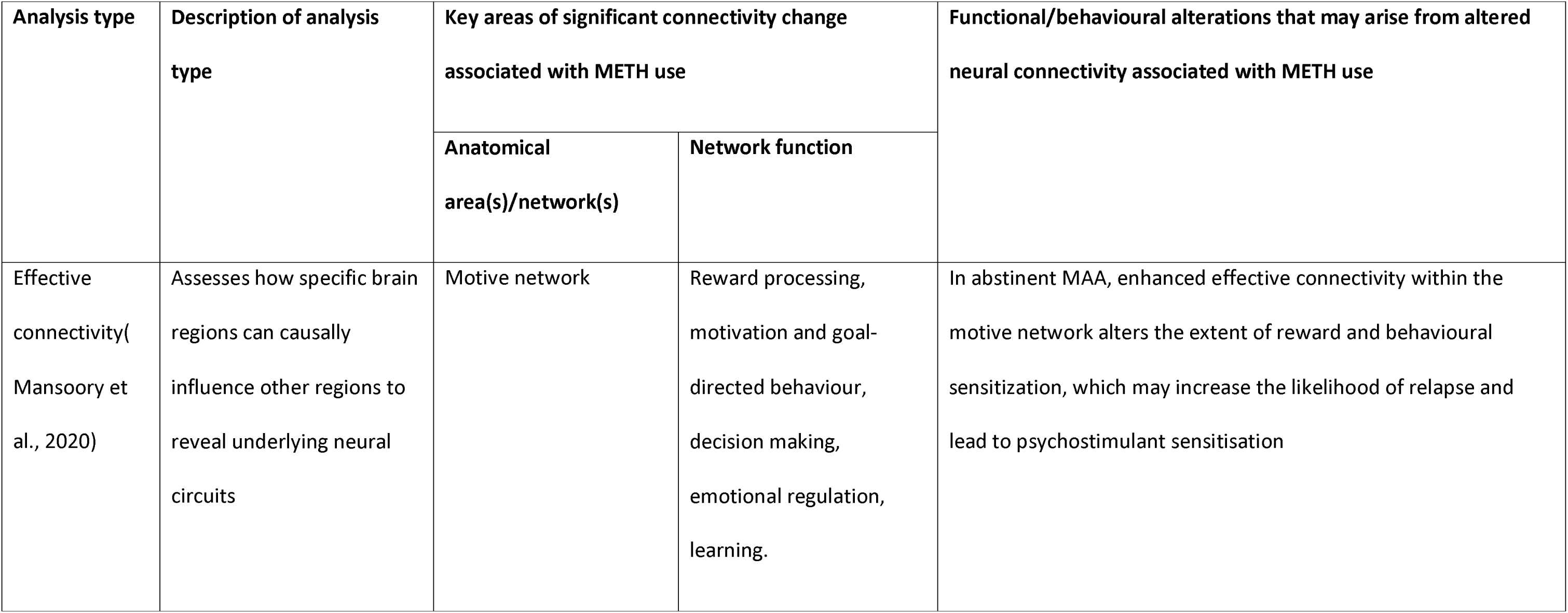
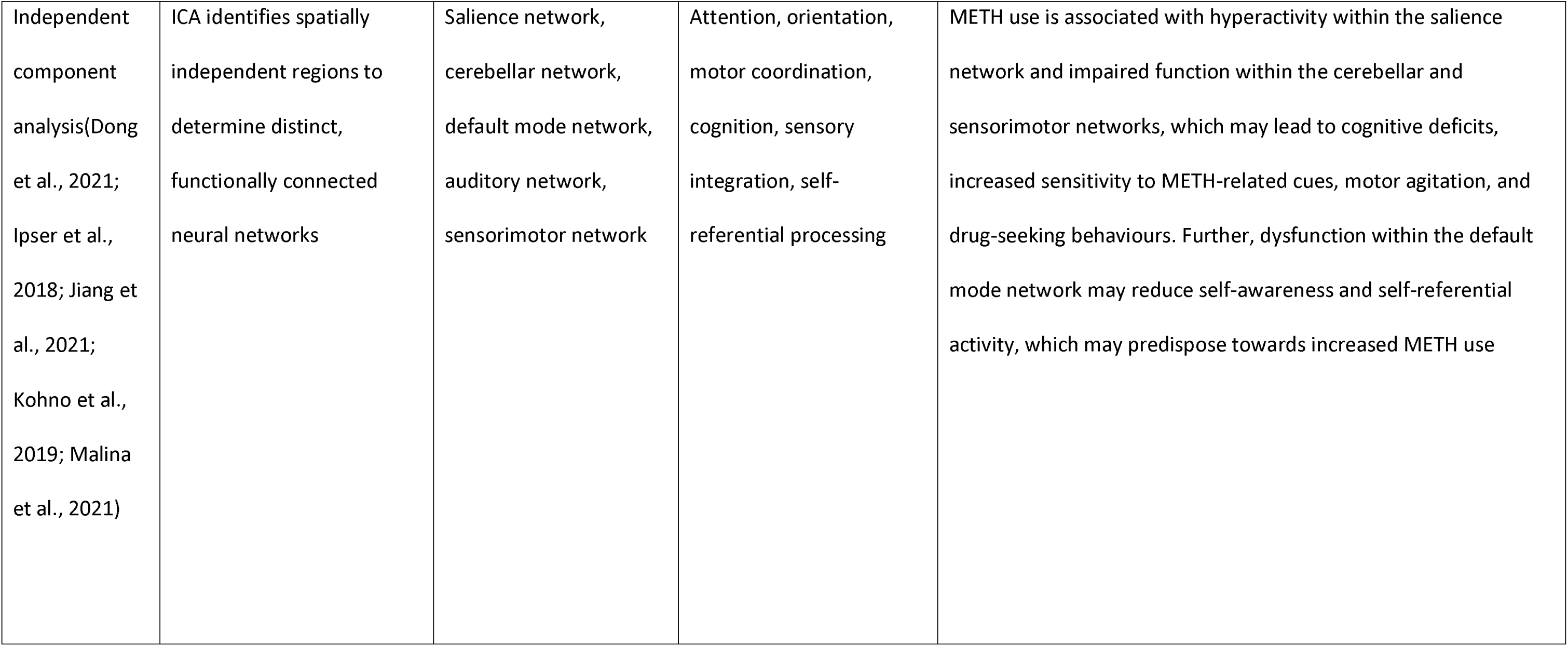
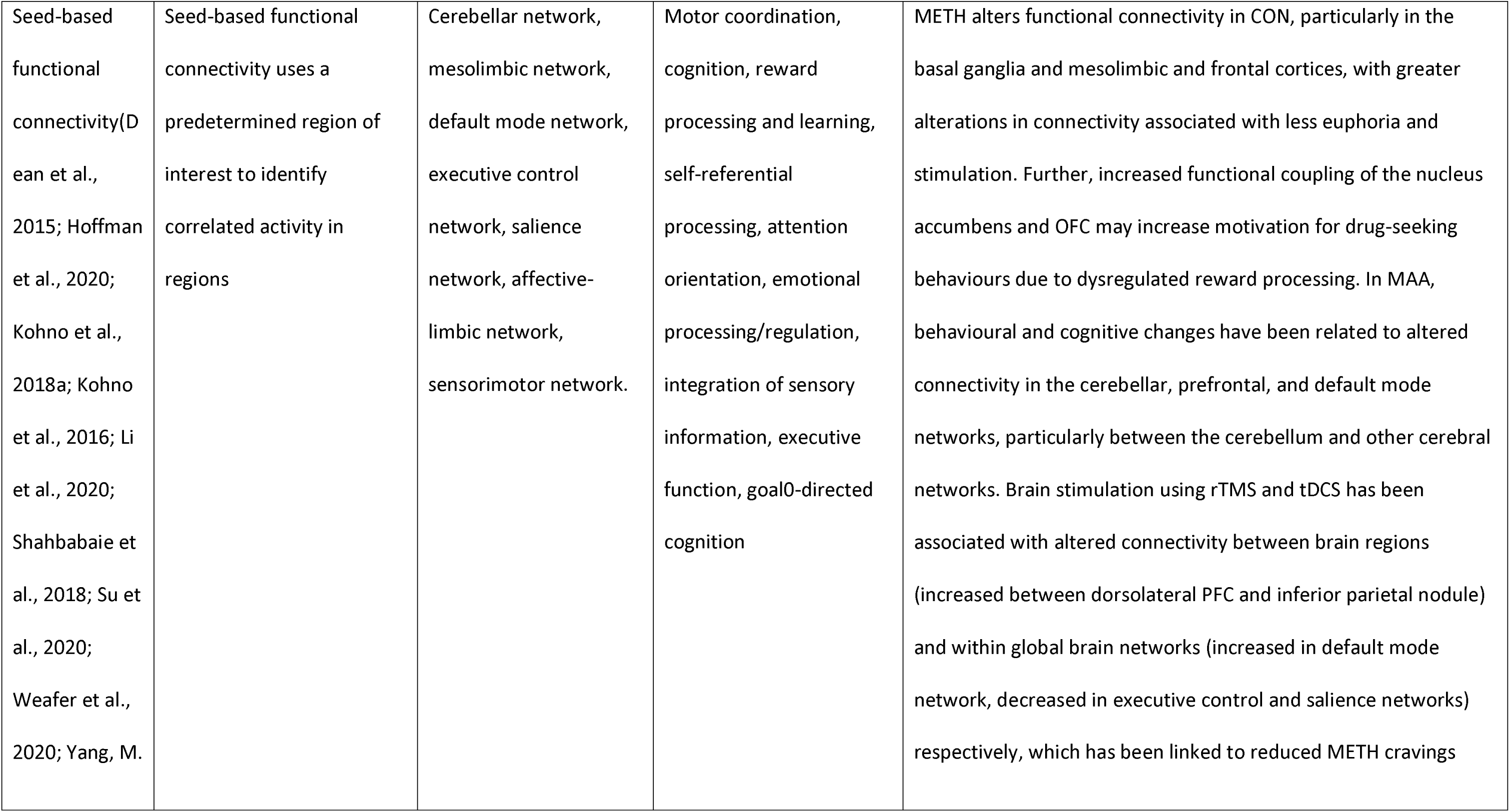
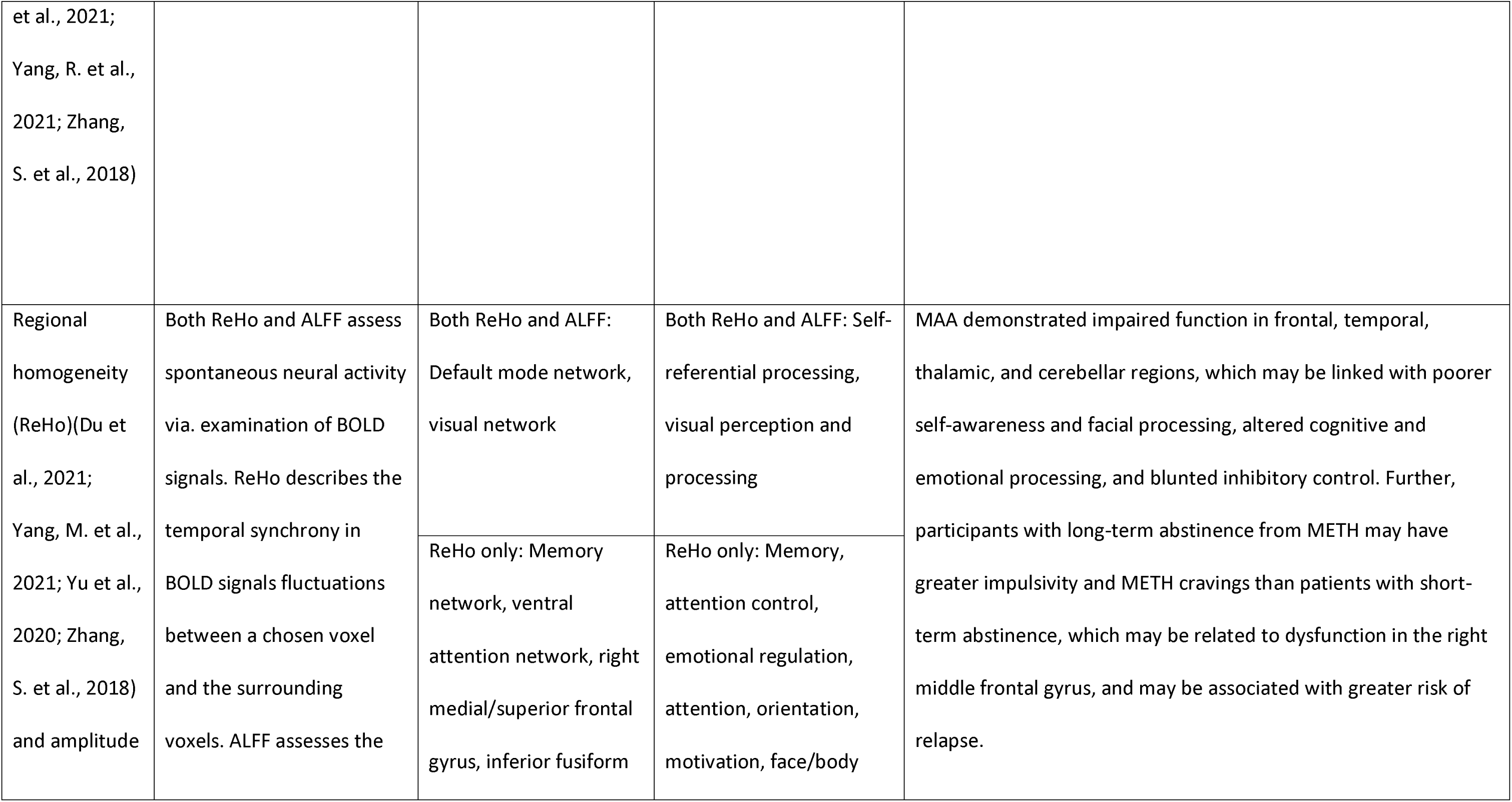
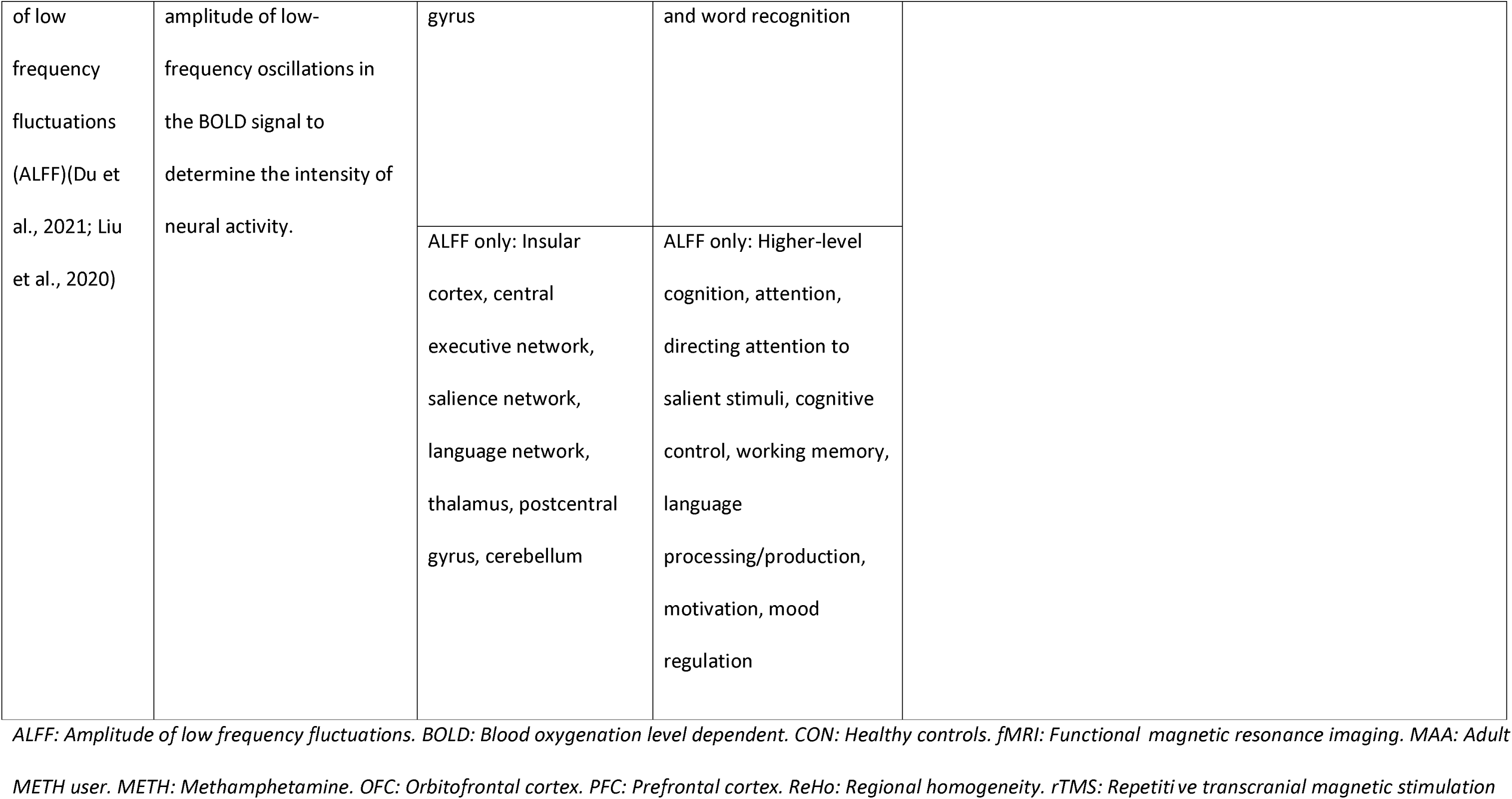
Resting-state fMRI findings.

### Independent Component Analysis

ICA works by separating the global BOLD signal from the entire brain into temporally correlated, functionally connected independent regions, and provides insight into the magnitude of FC between brain regions. Previous research has demonstrated that prolonged METH use is associated with altered FC within multiple neural networks,(Dong et al., 2021; Jiang et al., 2021) including the DMN (which manages self-referential processing and passive cognitive tasks),(Buckner, 2013) SN (which orients attention towards salient stimuli),(Schimmelpfennig et al., 2023) and cerebellar networks (which co-ordinate motor, cognitive, emotional, and autonomic functions),(Schmahmann, 2019) all of which may predispose towards cognitive deficits, maladaptive interpersonal behaviour, and impaired self-awareness in MAA. In particular, Dong et al.(Dong et al., 2021) reported that many of these FC aberrancies centred around the dorsal DMN, and that some of these changes persisted even after sustained METH abstinence. In contrast, Malina et al.(Malina et al., 2021) reported that in METH-naïve individuals, acute METH infusions altered between-network connectivity across the thalamus, cerebellum, and frontal and temporal cortices, but had no effect on FC within networks.

ICA has also been used to investigate potential treatments for METH dependence. Naltrexone, a µ-opioid receptor antagonist, has been posited as a pharmacological treatment for METH cravings.(Karila et al., 2010) In a double-blind RCT, Kohno et al.(Kohno et al., 2019) reported that naltrexone was associated with reduced FC between the DMN and CEN, which correlated with reduced METH use and substance dependence severity. Given the opposing functions of the DMN and CEN, whereby the former is involved in passive cognitive tasks and the latter in coordinating goal-directed responses to external stimuli, Kohno et al. suggested that naltrexone beneficially impacted network dynamics by functionally decoupling the DMN and CEN in MAA, and thereby supported better cognitive and behavioural control.

### Seed-based functional connectivity

Seed-based FC analysis reveals correlations in activity between a pre-specified region of interest (ROI), referred to as a seed ROI, and other brain regions, which can then elucidate functionally connected neural networks. In particular, seed-based FC studies have consistently demonstrated that METH use is associated with altered FC in the mesocorticolimbic network, a dopaminergic neural circuit involved in reward processing and motivated behaviour.(Alcaro et al., 2007) Kohno et al.(Kohno et al., 2016) reported that METH use was associated with increased mesocorticolimbic resting FC that was positively correlated with cognitive impulsivity and negatively correlated with D2-type receptor availability in the ventral striatum. The ventral striatum is a critical component of the mesocorticolimbic pathway that is sensitive to both aberrant dopaminergic and inflammatory processes,(Inagaki et al., 2015; Smith et al., 2018) and ventral striatal dysfunction has been closely implicated in the pathophysiology of METH addiction.(Hoffman et al., 2020; Kohno et al., 2018b; Kohno et al., 2016) Further, Kohno et al.(Kohno et al., 2018b) and Hoffman et al.(Hoffman et al., 2020) found that METH use was associated with increased corticostriatal resting-state FC (specifically between the mesocorticolimbic pathway and dlPFC, involved in cognitive control and goal-directed behaviour), suggesting that METH may impair cognitive control processes and sensitise users towards reward-driven behaviours. Thus, it is possible that METH-associated abnormalities in dopamine release, dopamine receptor availability, and neuroinflammation within the mesocorticolimbic pathway may alter FC between the ventral striatum and other brain regions (e.g. the midbrain), which may subsequently lead to increased cognitive impulsivity and other behavioural deficits.

Following on from these findings, some studies have explored the use of transcranial stimulation of the dlPFC to reduce METH cravings. Specifically, Shahbabaie et al.(Shahbabaie et al., 2018) reported that transcranial direct current stimulation (tDCS) of the dlPFC altered FC across multiple large-scale networks, including the DMN, SN, and CEN, while Su et al.(Su et al., 2020) found that repetitive transcranial magnetic stimulation (rTMS) increased FC between the dlPFC and inferior parietal lobule. Nonetheless, both studies found that tDCS and rTMS were associated with reduced METH cravings, suggesting that neuroplastic modulation of network connectivity using transcranial stimulation may be a promising adjunct for treatment of chronic METH use.

### Effective connectivity

In brief, EC describes the “influence that one neural system exerts over another”.(Friston, 2011) In a cross-sectional analysis of abstinent MAA, Mansoory et al.(Mansoory et al., 2020) reported that METH use was associated with altered EC within the motive circuit, a neural network spanning frontal, thalamic, and basal ganglia structures that modulates reward and learning. Specifically, they found that MAA exhibited increased EC from the PFC to the ventral pallidum (VP) (PFC → VP) and within the mediodorsal thalamus (MD) self-loop (MD → MD) and decreased EC within the VP self-loop (VP → VP), and proposed that these findings may reflect damaged or compensatory activity within the motive circuit, which may translate functionally into behavioural sensitisation, anhedonia, and reduced motivation.

### Regional Homogeneity and Amplitude of Low-Frequency Fluctuations

ReHo and ALFF both measure the intensity and temporal synchrony of local neural activity and are often used simultaneously to identify ROIs for further FC analysis. Du et al.(Du et al., 2021) conducted a longitudinal assessment of 50 individuals with short- and long-term abstinence from METH, and found that long-term abstinent participants demonstrated increased ALFF and ReHo within the right middle frontal gyrus which correlated with increased impulsivity. Given the role of the right middle frontal gyrus in controlling motivation, with increased activity in this region being previously linked to increased desire for substances, they suggested that long-term abstinent MAA may exhibit stronger substance cravings than short-term abstinent counterparts and so be at greater risk for relapse.

However, they acknowledged that their analysis was limited by its short duration of follow-up (1 year). Further, Yu et al.(Yu et al., 2020) constructed a diagnostic model to distinguish between MAA and CON using MRI; they found significant ReHo differences across groups within the right medial superior frontal and temporal inferior fusiform gyri, and reported a model accuracy of 84% when these changes were considered within a machine-learning algorithm with AdaBoost classification.

For MAP specifically, Yang et al.(Yang, M. et al., 2021) demonstrated that patients with MAP had decreased and increased ReHo within cortical and striatal regions respectively when compared to CON; this cortico-striatal imbalance may in turn reflect underlying cortical deficits that can predispose towards reduced top-down control or reactive subcortical compensation. Further, they also found that ReHo within the right superior temporal gyrus was negatively correlated with psychotic symptoms, and that this ReHo alteration mediated the positive correlation between increased frequency of METH use and positive psychotic symptoms. These findings are consistent with previous studies implicating the right superior temporal gyrus in auditory processing, language, and social cognition,(Bigler et al., 2007) and suggests that this region may be a target for future research exploring the neural pathophysiology or treatment of MAP.

### Task-fMRI

Task-fMRI involves measuring changes in regional cerebral blood flow and oxygenation during the performance of certain tasks as an indicator of neural metabolic activity (and therefore, activation).(Ogawa et al., 1990) Of the 48 papers that employed task-fMRI, the most common tasks used were Stroop (7/48, 15%) and cue reactivity (6/48, 13%) tasks.

In brief, METH use is associated with diffuse impairment across a variety of brain regions that may predispose towards functional alterations and maladaptive behaviours. These may include, but are not limited to, emotional dysregulation, prioritisation of small immediate gains over large delayed benefits, impaired learning and feedback processing, and reduced cognitive flexibility and overall cognitive control. **Table 5** summarises the main findings of these papers, categorised according to the task utilised and the overarching cognitive domain that they assessed.

**Table 5:**
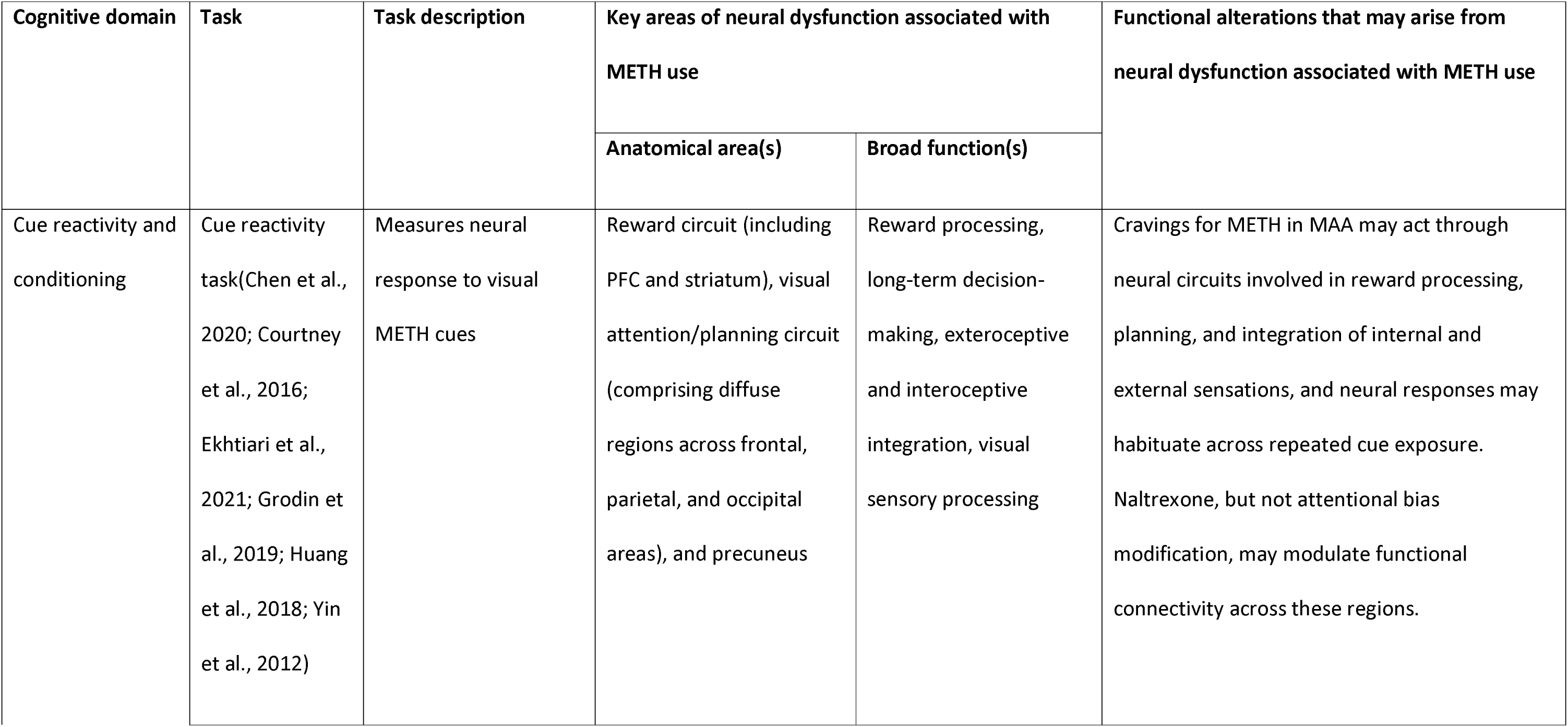
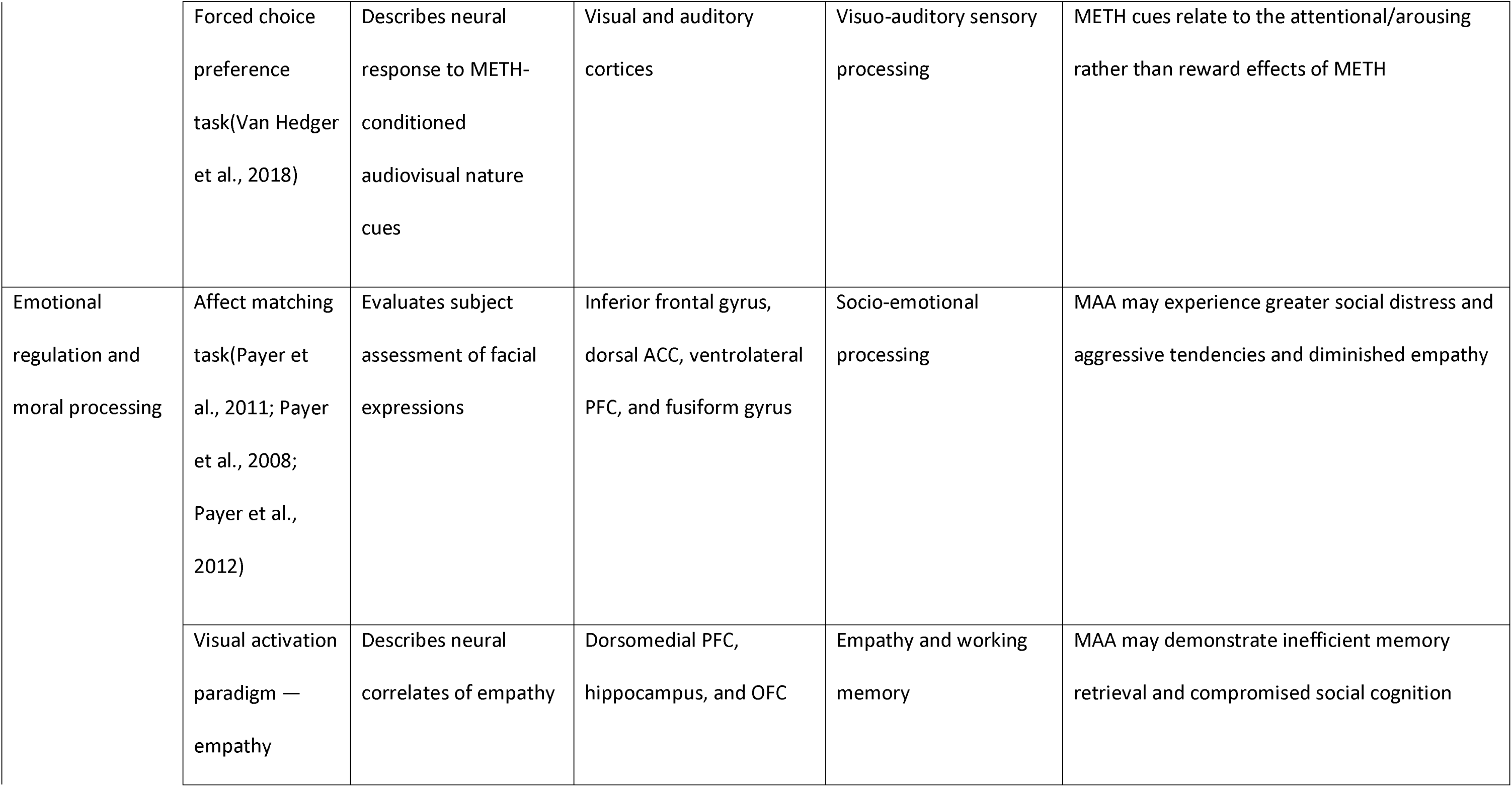
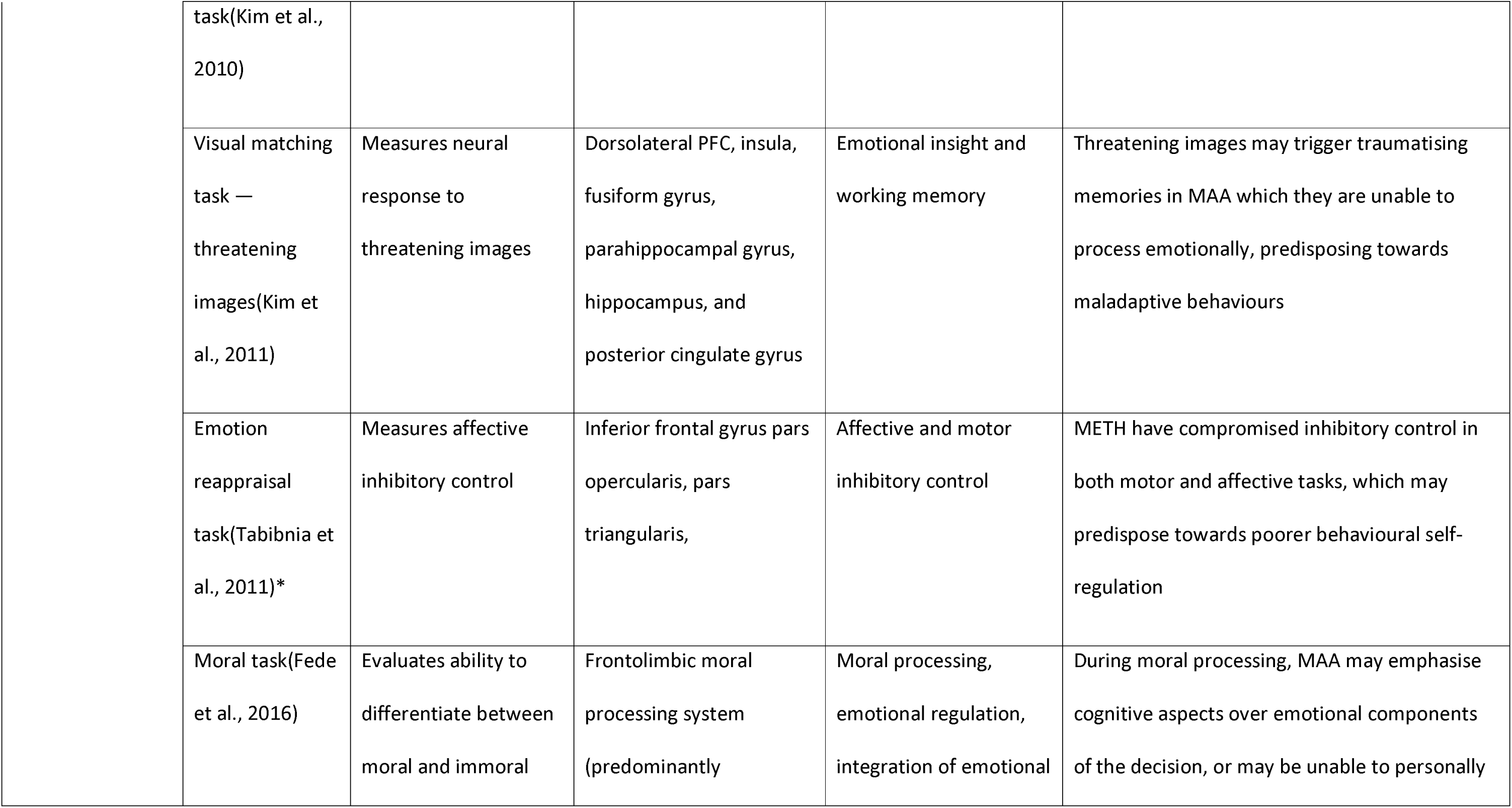
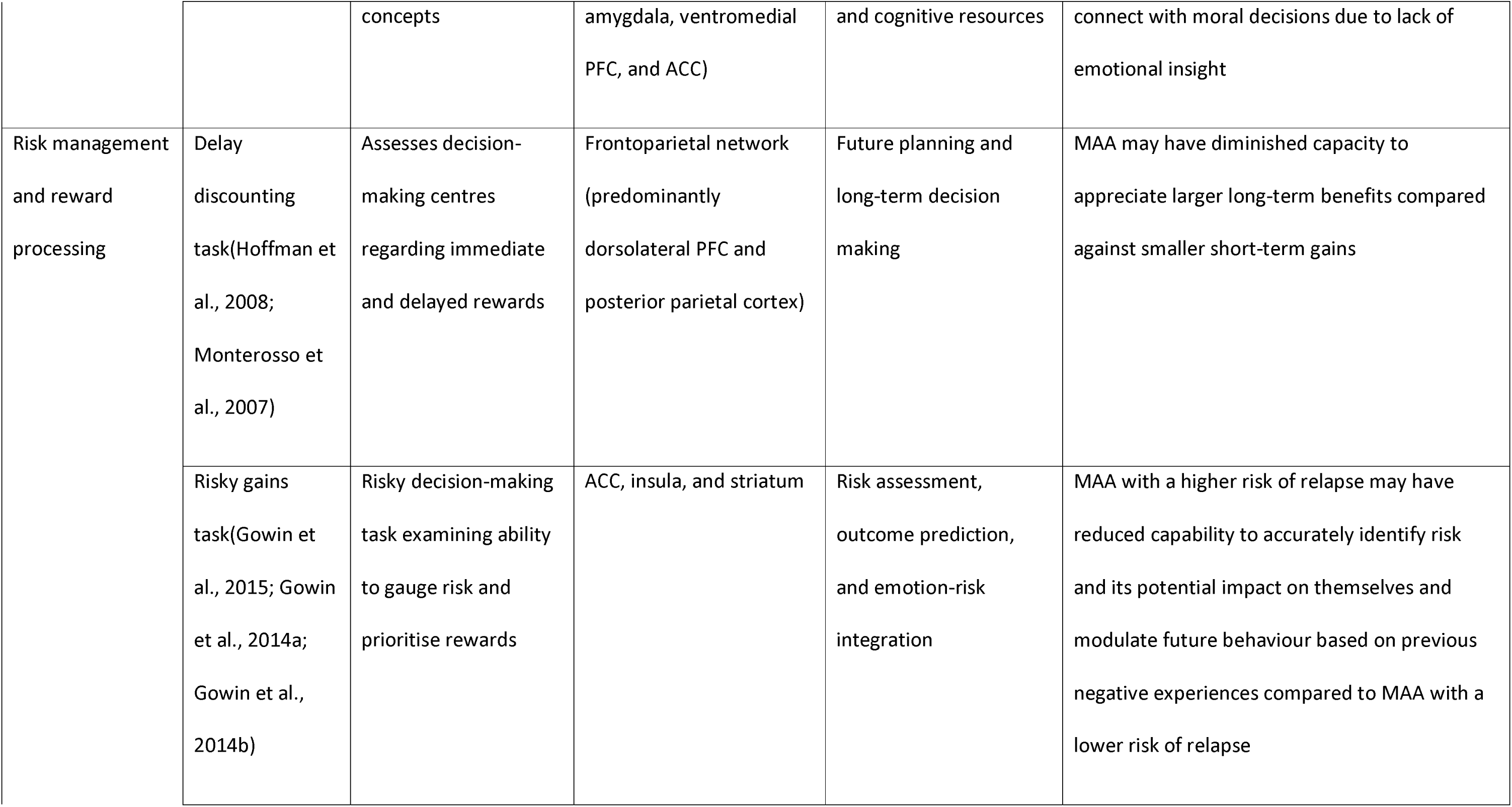
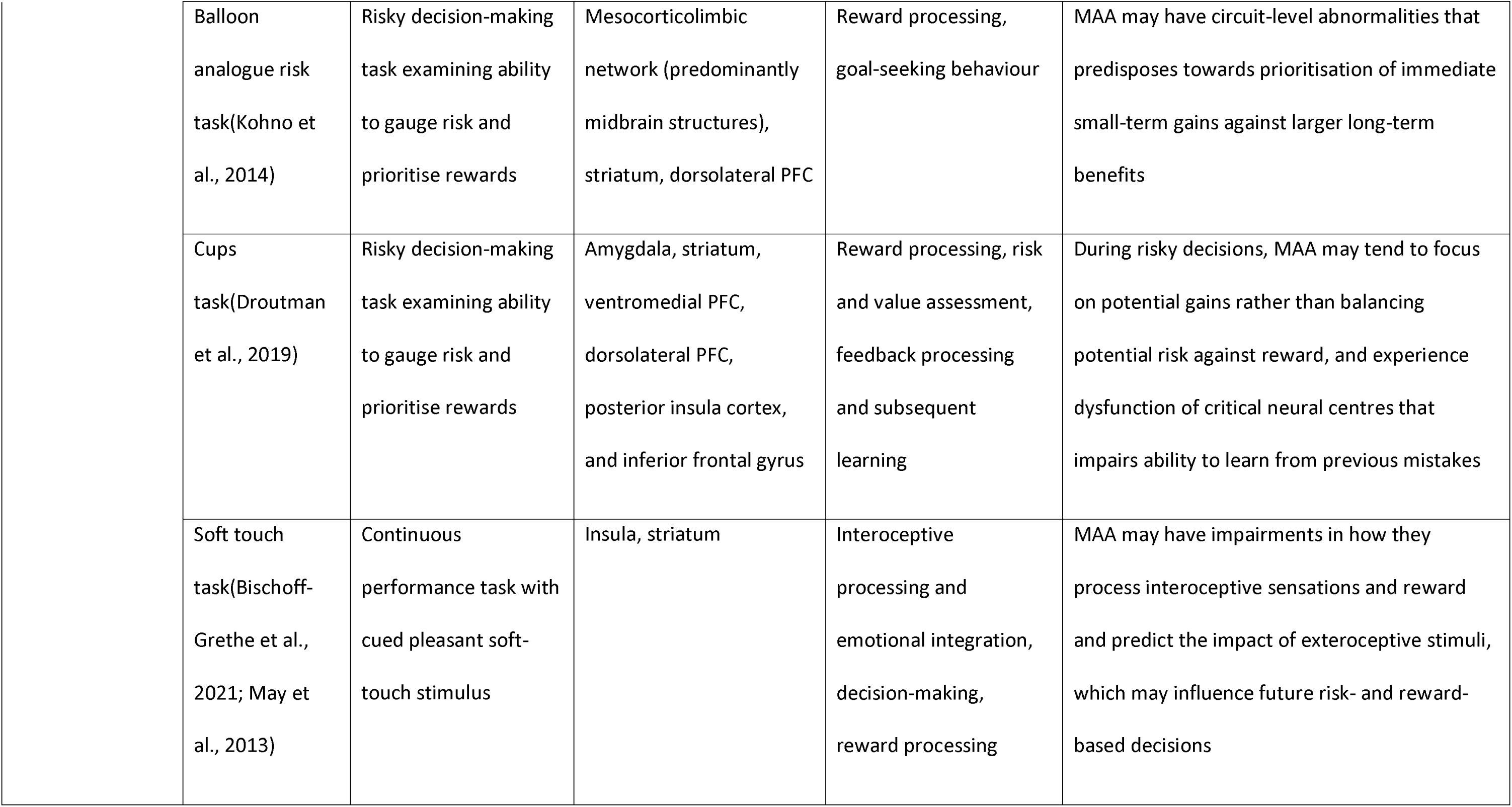
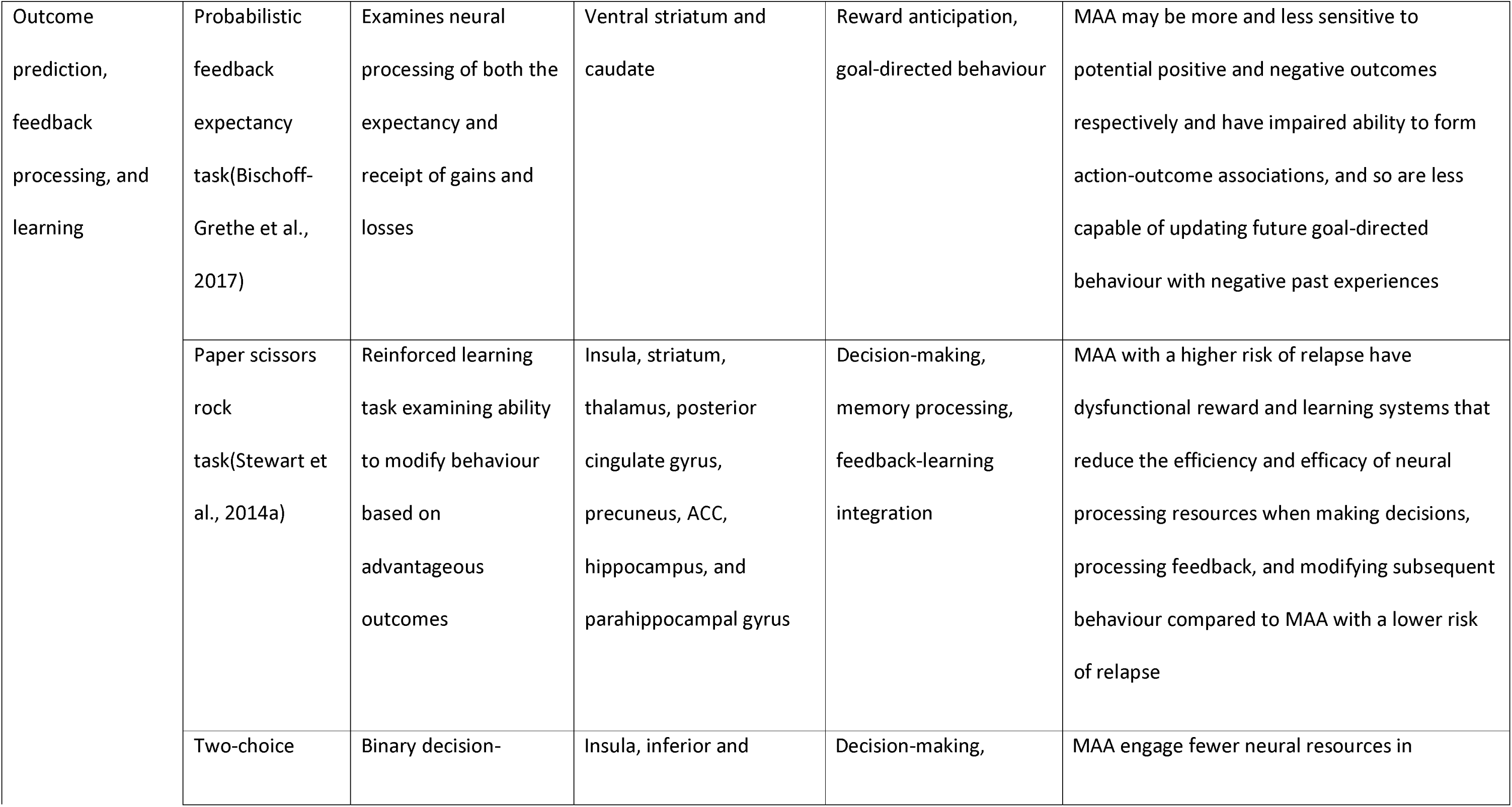
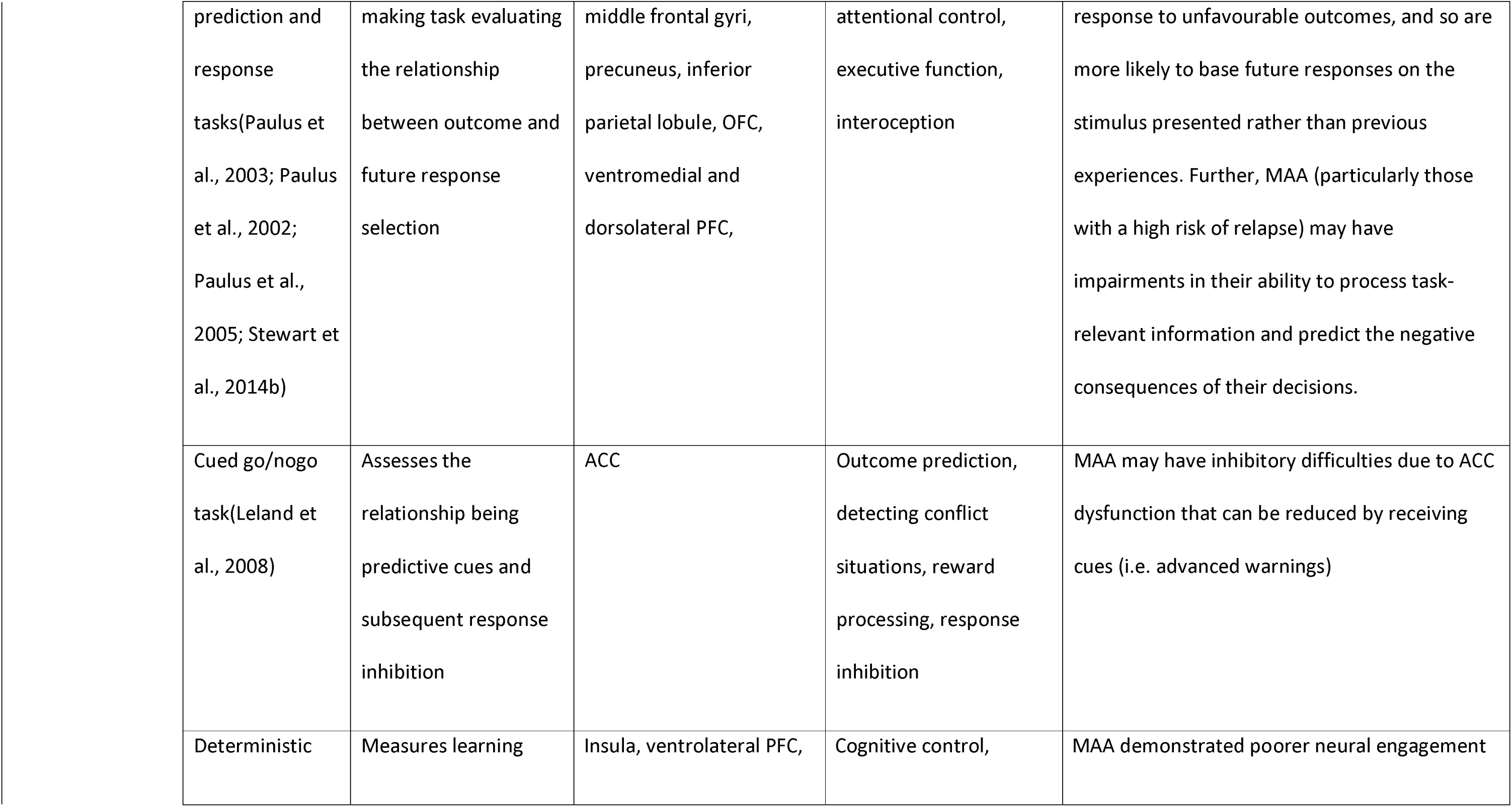
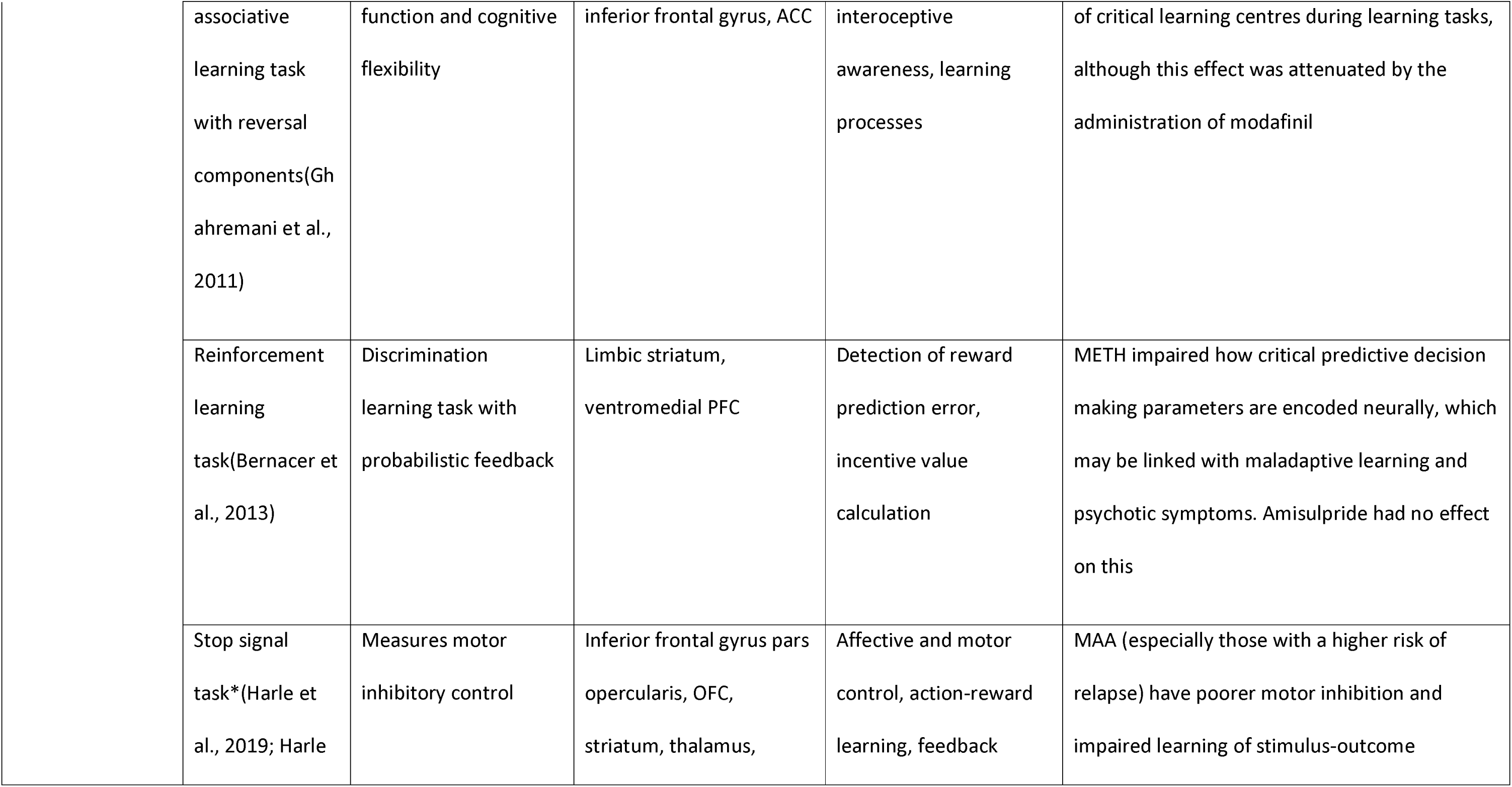
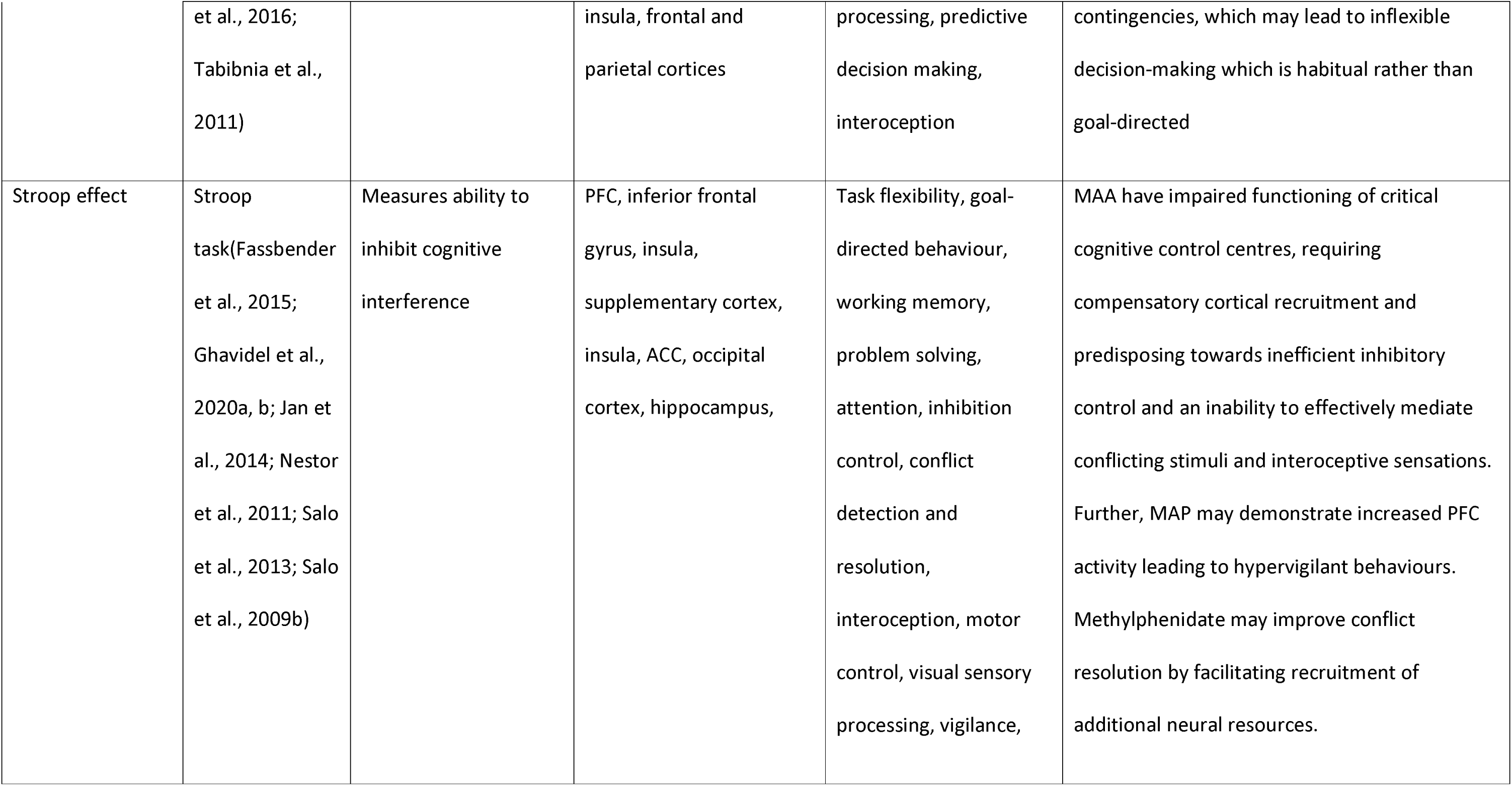
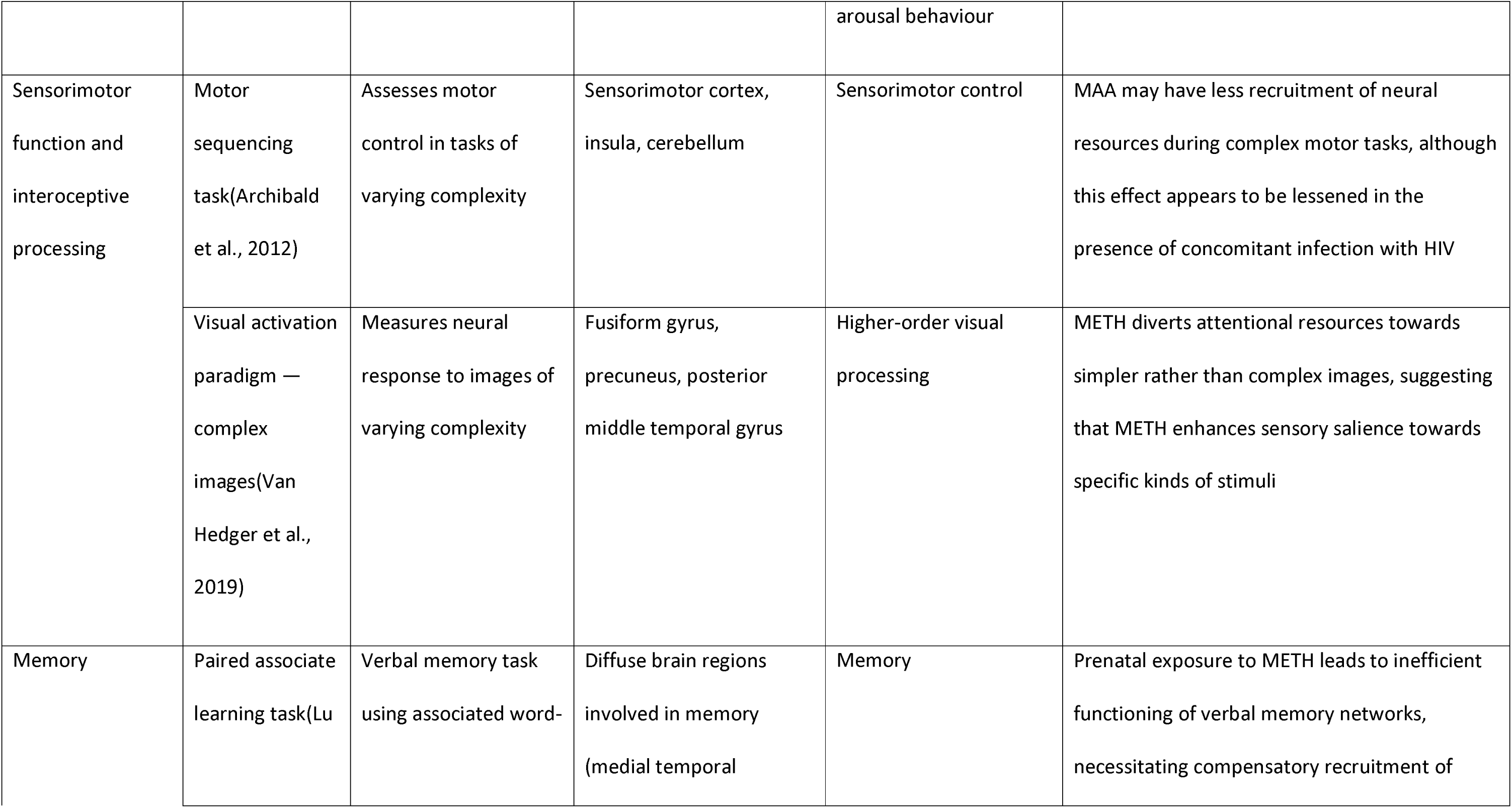
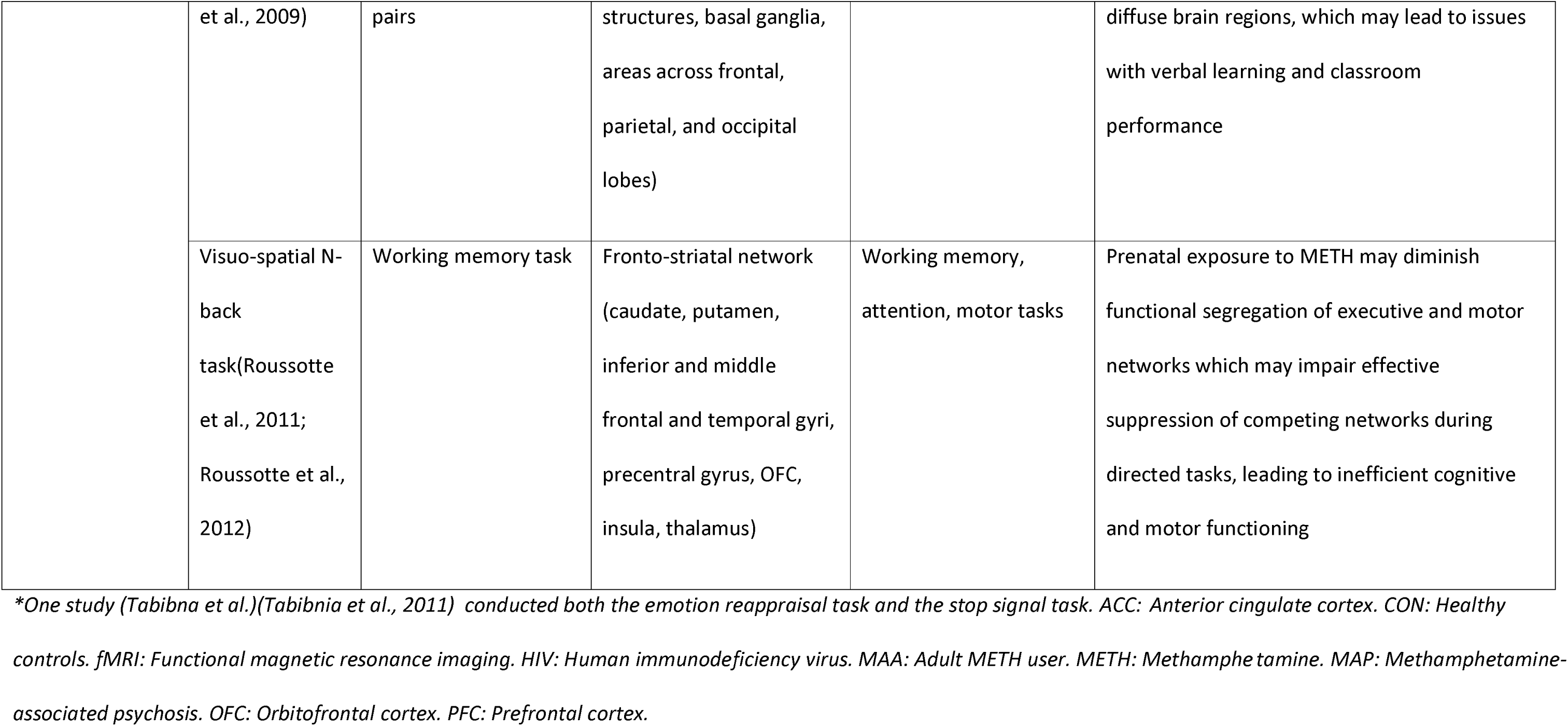
task-fMRI findings.

| Cognitive domain | Task | Task description | Key areas of neural dysfunction associated with METH use |  | Functional alterations that may arise from neural dysfunction associated with METH use |
| --- | --- | --- | --- | --- | --- |
|  |  |  | Anatomical area(s) | Broad function(s) |  |
| Cue reactivity and conditioning | Cue reactivity task(Chen et al., 2020; Courtney et al., 2016; Ekhtiari et al., 2021; Grodin et al., 2019; Huang et al., 2018; Yin et al., 2012) | Measures neural response to visual METH cues | Reward circuit (including PFC and striatum), visual attention/planning circuit (comprising diffuse regions across frontal, parietal, and occipital areas), and precuneus | Reward processing, long-term decision-making, exteroceptive and interoceptive integration, visual sensory processing | Cravings for METH in MAA may act through neural circuits involved in reward processing, planning, and integration of internal and external sensations, and neural responses may habituate across repeated cue exposure. Naltrexone, but not attentional bias modification, may modulate functional connectivity across these regions. |
|  | Forced choice preference task(Van Hedger et al., 2018) | Describes neural response to METH-conditioned audiovisual nature cues | Visual and auditory cortices | Visuo-auditory sensory processing | METH cues relate to the attentional/arousing rather than reward effects of METH |
| Emotional regulation and moral processing | Affect matching task(Payer et al., 2011; Payer et al., 2008; Payer et al., 2012) | Evaluates subject assessment of facial expressions | Inferior frontal gyrus, dorsal ACC, ventrolateral PFC, and fusiform gyrus | Socio-emotional processing | MAA may experience greater social distress and aggressive tendencies and diminished empathy |
|  | Visual activation paradigm — empathy | Describes neural correlates of empathy | Dorsomedial PFC, hippocampus, and OFC | Empathy and working memory | MAA may demonstrate inefficient memory retrieval and compromised social cognition |

|  |  |  |  |  |
| --- | --- | --- | --- | --- |
| task(Kim et al., 2010) |  |  |  |  |
| Visual matching task — threatening images(Kim et al., 2011) | Measures neural response to threatening images | Dorsolateral PFC, insula, fusiform gyrus, parahippocampal gyrus, hippocampus, and posterior cingulate gyrus | Emotional insight and working memory | Threatening images may trigger traumatising memories in MAA which they are unable to process emotionally, predisposing towards maladaptive behaviours |
| Emotion reappraisal task(Tabibnia et al., 2011)* | Measures affective inhibitory control | Inferior frontal gyrus pars opercularis, pars triangularis, | Affective and motor inhibitory control | METH have compromised inhibitory control in both motor and affective tasks, which may predispose towards poorer behavioural self-regulation |
| Moral task(Fede et al., 2016) | Evaluates ability to differentiate between moral and immoral | Frontolimbic moral processing system (predominantly | Moral processing, emotional regulation, integration of emotional | During moral processing, MAA may emphasise cognitive aspects over emotional components of the decision, or may be unable to personally |
|  |  | concepts | amygdala, ventromedial PFC, and ACC) | and cognitive resources | connect with moral decisions due to lack of emotional insight |
| Risk management and reward processing | Delay discounting task(Hoffman et al., 2008; Monterosso et al., 2007) | Assesses decision-making centres regarding immediate and delayed rewards | Frontoparietal network (predominantly dorsolateral PFC and posterior parietal cortex) | Future planning and long-term decision making | MAA may have diminished capacity to appreciate larger long-term benefits compared against smaller short-term gains |
|  | Risky gains task(Gowin et al., 2015; Gowin et al., 2014a; Gowin et al., 2014b) | Risky decision-making task examining ability to gauge risk and prioritise rewards | ACC, insula, and striatum | Risk assessment, outcome prediction, and emotion-risk integration | MAA with a higher risk of relapse may have reduced capability to accurately identify risk and its potential impact on themselves and modulate future behaviour based on previous negative experiences compared to MAA with a lower risk of relapse |

|  |  |  |  |  |
| --- | --- | --- | --- | --- |
| Balloon analogue risk task(Kohno et al., 2014) | Risky decision-making task examining ability to gauge risk and prioritise rewards | Mesocorticolimbic network (predominantly midbrain structures), striatum, dorsolateral PFC | Reward processing, goal-seeking behaviour | MAA may have circuit-level abnormalities that predisposes towards prioritisation of immediate small-term gains against larger long-term benefits |
| Cups task(Droutman et al., 2019) | Risky decision-making task examining ability to gauge risk and prioritise rewards | Amygdala, striatum, ventromedial PFC, dorsolateral PFC, posterior insula cortex, and inferior frontal gyrus | Reward processing, risk and value assessment, feedback processing and subsequent learning | During risky decisions, MAA may tend to focus on potential gains rather than balancing potential risk against reward, and experience dysfunction of critical neural centres that impairs ability to learn from previous mistakes |
| Soft touch task(Bischoff-Grethe et al., 2021; May et al., 2013) | Continuous performance task with cued pleasant soft-touch stimulus | Insula, striatum | Interoceptive processing and emotional integration, decision-making, reward processing | MAA may have impairments in how they process interoceptive sensations and reward and predict the impact of exteroceptive stimuli, which may influence future risk- and reward-based decisions |
| Outcome<br>prediction,<br>feedback<br>processing, and<br>learning | Probabilistic<br>feedback<br>expectancy<br>task(Bischoff-Grethe et al.,<br>2017) | Examines neural<br>processing of both the<br>expectancy and<br>receipt of gains and<br>losses | Ventral striatum and<br>caudate | Reward anticipation,<br>goal-directed behaviour | MAA may be more and less sensitive to<br>potential positive and negative outcomes<br>respectively and have impaired ability to form<br>action-outcome associations, and so are less<br>capable of updating future goal-directed<br>behaviour with negative past experiences |
|  | Paper scissors<br>rock<br>task(Stewart et<br>al., 2014a) | Reinforced learning<br>task examining ability<br>to modify behaviour<br>based on<br>advantageous<br>outcomes | Insula, striatum,<br>thalamus, posterior<br>cingulate gyrus,<br>precuneus, ACC,<br>hippocampus, and<br>parahippocampal gyrus | Decision-making,<br>memory processing,<br>feedback-learning<br>integration | MAA with a higher risk of relapse have<br>dysfunctional reward and learning systems that<br>reduce the efficiency and efficacy of neural<br>processing resources when making decisions,<br>processing feedback, and modifying subsequent<br>behaviour compared to MAA with a lower risk<br>of relapse |
|  | Two-choice | Binary decision- | Insula, inferior and | Decision-making, | MAA engage fewer neural resources in |

|  |  |  |  |  |
| --- | --- | --- | --- | --- |
| prediction and response tasks(Paulus et al., 2003; Paulus et al., 2002; Paulus et al., 2005; Stewart et al., 2014b) | making task evaluating the relationship between outcome and future response selection | middle frontal gyri, precuneus, inferior parietal lobule, OFC, ventromedial and dorsolateral PFC, | attentional control, executive function, interoception | response to unfavourable outcomes, and so are more likely to base future responses on the stimulus presented rather than previous experiences. Further, MAA (particularly those with a high risk of relapse) may have impairments in their ability to process task-relevant information and predict the negative consequences of their decisions. |
| Cued go/nogo task(Leland et al., 2008) | Assesses the relationship being predictive cues and subsequent response inhibition | ACC | Outcome prediction, detecting conflict situations, reward processing, response inhibition | MAA may have inhibitory difficulties due to ACC dysfunction that can be reduced by receiving cues (i.e. advanced warnings) |
| Deterministic | Measures learning | Insula, ventrolateral PFC, | Cognitive control, | MAA demonstrated poorer neural engagement |

|  |  |  |  |  |
| --- | --- | --- | --- | --- |
| associative learning task with reversal components(Ghahremani et al., 2011) | function and cognitive flexibility | inferior frontal gyrus, ACC | interoceptive awareness, learning processes | of critical learning centres during learning tasks, although this effect was attenuated by the administration of modafinil |
| Reinforcement learning task(Bernacer et al., 2013) | Discrimination learning task with probabilistic feedback | Limbic striatum, ventromedial PFC | Detection of reward prediction error, incentive value calculation | METH impaired how critical predictive decision making parameters are encoded neurally, which may be linked with maladaptive learning and psychotic symptoms. Amisulpride had no effect on this |
| Stop signal task*(Harle et al., 2019; Harle | Measures motor inhibitory control | Inferior frontal gyrus pars opercularis, OFC, striatum, thalamus, | Affective and motor control, action-reward learning, feedback | MAA (especially those with a higher risk of relapse) have poorer motor inhibition and impaired learning of stimulus-outcome |
|  | et al., 2016;<br>Tabibnia et al.,<br>2011) |  | insula, frontal and<br>parietal cortices | processing, predictive<br>decision making,<br>interoception | contingencies, which may lead to inflexible<br>decision-making which is habitual rather than<br>goal-directed |
| Stroop effect | Stroop<br>task(Fassbender<br>et al., 2015;<br>Ghavidel et al.,<br>2020a, b; Jan et<br>al., 2014; Nestor<br>et al., 2011; Salo<br>et al., 2013; Salo<br>et al., 2009b) | Measures ability to<br>inhibit cognitive<br>interference | PFC, inferior frontal<br>gyrus, insula,<br>supplementary cortex,<br>insula, ACC, occipital<br>cortex, hippocampus, | Task flexibility, goal-directed behaviour,<br>working memory,<br>problem solving,<br>attention, inhibition<br>control, conflict<br>detection and<br>resolution,<br>interoception, motor<br>control, visual sensory<br>processing, vigilance, arousal behaviour | MAA have impaired functioning of critical<br>cognitive control centres, requiring<br>compensatory cortical recruitment and<br>predisposing towards inefficient inhibitory<br>control and an inability to effectively mediate<br>conflicting stimuli and interoceptive sensations.<br>Further, MAP may demonstrate increased PFC<br>activity leading to hypervigilant behaviours.<br>Methylphenidate may improve conflict<br>resolution by facilitating recruitment of<br>additional neural resources. |
| Sensorimotor function and interoceptive processing | Motor sequencing task(Archibald et al., 2012) | Assesses motor control in tasks of varying complexity | Sensorimotor cortex, insula, cerebellum | Sensorimotor control | MAA may have less recruitment of neural resources during complex motor tasks, although this effect appears to be lessened in the presence of concomitant infection with HIV |
|  | Visual activation paradigm — complex images(Van Hedger et al., 2019) | Measures neural response to images of varying complexity | Fusiform gyrus, precuneus, posterior middle temporal gyrus | Higher-order visual processing | METH diverts attentional resources towards simpler rather than complex images, suggesting that METH enhances sensory salience towards specific kinds of stimuli |
| Memory | Paired associate learning task(Lu | Verbal memory task using associated word- | Diffuse brain regions involved in memory (medial temporal | Memory | Prenatal exposure to METH leads to inefficient functioning of verbal memory networks, necessitating compensatory recruitment of |
|  | et al., 2009) | pairs | structures, basal ganglia, areas across frontal, parietal, and occipital lobes) |  | diffuse brain regions, which may lead to issues with verbal learning and classroom performance |
|  | Visuo-spatial N-back task(Roussotte et al., 2011; Roussotte et al., 2012) | Working memory task | Fronto-striatal network (caudate, putamen, inferior and middle frontal and temporal gyri, precentral gyrus, OFC, insula, thalamus) | Working memory, attention, motor tasks | Prenatal exposure to METH may diminish functional segregation of executive and motor networks which may impair effective suppression of competing networks during directed tasks, leading to inefficient cognitive and motor functioning |

*\*One study (Tabibnia et al.)(Tabibnia et al., 2011) conducted both the emotion reappraisal task and the stop signal task. ACC: Anterior cingulate cortex. CON: Healthy controls. fMRI: Functional magnetic resonance imaging. HIV: Human immunodeficiency virus. MAA: Adult METH user. METH: Methamphetamine. MAP: Methamphetamine-associated psychosis. OFC: Orbitofrontal cortex. PFC: Prefrontal cortex.*

### Cue reactivity and conditioning

Cue-induced cravings are purported to act through two circuits: a reward circuit (comprising cortical and subcortical structures) and a visual attention/planning circuit (comprising frontal, parietal, and occipital cortical regions).(Due et al., 2002) In Huang et al.’s(Huang et al., 2018) analysis of people with long-term abstinence from METH, they proposed that METH cues may trigger cravings through the reward circuit and then draw attention via the visual attention/planning circuit. However, the role of striatal dysfunction (a critical component of the reward circuit) in METH cue-induced cravings remains unclear, although it has been suggested that this may be due to the different studies examining different stages of METH addiction and abstinence.(Chen et al., 2020) This hypothesis is supported by van Hedger et al’s.(Van Hedger et al., 2018) study who found that METH, when paired with visual-auditory cues, activated exclusively sensory processing regions in METH-naïve participants, suggesting that the conditioned neural response relied more heavily on the attentional impact of METH in those people rather than the rewarding effects.

Multiple studies have evaluated potential interventions to modulate METH cravings. Ekhitari et al.(Ekhtiari et al., 2021) reported that repeated METH cue exposure habituated increased activation in the PFC, amygdala, and striatum, and suggested that cue-exposure interventions may be explored as a potential tool for reducing METH cravings. Courtney et al.(Courtney et al., 2016) conducted a randomized controlled trial (RCT) evaluating naltrexone in patients with METH use, and found that naltrexone increased connectivity between the striatum and prefrontal regions during METH cravings, indicating greater frontal recruitment to regulate decision-making. In contrast, Dean et al.(Dean et al., 2019) found that attentional bias modification training did not change brain activation during METH cues or reduce METH cravings.

### Emotional regulation and moral processing

Multiple studies have demonstrated that METH use is associated with reduced or inefficient activation of the inferior frontal gyrus, ACC, insula, and orbitofrontal cortex, which has been linked to deficits in empathy, emotional insight, and social cognition.(Fede et al., 2016; Kim et al., 2010; Kim et al., 2011; Payer et al., 2011; Payer et al., 2008; Payer et al., 2012) Further, METH use is also associated with dysfunction in the PFC, hippocampus, parahippocampal gyrus, and inferior frontal gyrus pars opercularis, which may indicate impairments in working memory,(Kim et al., 2010; Kim et al., 2011) emotional inhibition,(Tabibnia et al., 2011) and moral processing.(Fede et al., 2016) These regions, particularly the PFC and insula/operculum, also act as key hubs in the frontoparietal and cingulo-opercular networks, which are critically involved in coordinating executive function, cognition,(Sheffield et al., 2015) and, in the case of the cingulo-opercular network, the tonic maintenance of attention.(Sadaghiani and D’Esposito, 2015) Put together, these findings suggest that people with METH use may have compromised socio-emotional processing that makes it challenging for them to either correctly interpret the emotional states of themselves and others or draw on previous experiences to perform empathic tasks; this may then lead to feelings of distress and hostility, which can predispose towards inappropriate or aggressive interpersonal behaviours. Previous research has also suggested that the fronto- parietal and cingulo-opercular networks (broadly comprising lateral prefrontal/posterior parietal and cingulate/insular/thalamic regions respectively) are critical in coordinating executive function and other cognitive tasks, and that the cingulo-opercular network may also have further roles in maintaining tonic alertness.

### Risk management and reward processing

Overall, METH use is associated with prioritisation of small immediate gains over larger delayed benefits,(Hoffman et al., 2008; Kohno et al., 2014; Monterosso et al., 2007) reduced ability to fully predict the negative impacts of potentially harmful events,(Gowin et al., 2014a; Gowin et al., 2014b) and impairments in interoceptive and reward processing,(Bischoff-Grethe et al., 2021; Droutman et al., 2019; May et al., 2013) particularly in patients with METH use at a higher risk of relapse.(Gowin et al., 2015) These functional alterations have been related to neural dysfunction in individual regions, particularly the insula (which processes and integrates interoceptive sensations with emotional information)(Bischoff-Grethe et al., 2021; Gowin et al., 2014a; Gowin et al., 2014b; Kohno et al., 2014; May et al., 2013) and striatum (involved in reward processing and risky decision-making),(Bischoff-Grethe et al., 2021; Droutman et al., 2019; Kohno et al., 2014; May et al., 2013) and across broader decision-making neural circuits, predominantly the frontoparietal network (which integrates contextual information, such as prior experiences, interoceptive states, and predicted outcomes, with the intensity/salience of the stimulus to modulate decision-making)(Hoffman et al., 2008; Monterosso et al., 2007) and the mesocorticolimbic pathway (which plays a critical role in reward processing and goal-directed behaviour).(Kohno et al., 2014) Taken together, these findings suggest that risky decision-making that prioritises immediate gain is related to both dysregulated reward processing and impaired cognitive control in MAA, and provide a potential neuropsychological substrate behind why some abstinent people with METH use are at higher risk of relapse than others. Building on this, Gowin et. al(Gowin et al., 2015) developed a neuroimaging predictive model (comprising eight areas from the basal ganglia, nucleus accumbens, and temporal, parietal, and insular cortices) that was able to differentiate between abstinent people with METH use who did or did not relapse with an area under the curve of 0.73 and a sensitivity and specificity of 0.61 and 0.77 respectively.

### Outcome prediction, feedback processing, and learning

Previous studies have suggested that MAA are more likely to use stimulus-driven rather than outcome-driven decision-making approaches (i.e. they tend to make decisions based on habit rather than accurately predicting the potential consequences of their decisions),(Paulus et al., 2003; Paulus et al., 2002; Stewart et al., 2014b) particularly in those at a higher risk of relapse,(Paulus et al., 2005) and that when they do predict decision outcomes, their predictions are skewed towards considering potential positive rather than negative outcomes.(Bischoff-Grethe et al., 2017; Stewart et al., 2014b) This has been linked to dysfunctional activation of critical decision-making centres, including the striatum, inferior and middle frontal gyri, PFC, and orbitofrontal cortex. Once those decisions have been made and the outcomes of those decisions become known, MAA also appear to have difficulties with processing feedback and integrating it into future behaviour change. Harle et al.(Harle et al., 2016) reported that METH use is associated with reduced activation of various frontal, parietal, and striatal regions involved in inhibitory control, managing impulsive urges, and learning stimulus-outcome contingencies. Further, MAA at a higher risk of relapse may also demonstrate reduced function of crucial regions involved in learning (i.e. temporo-parietal junction, inferior frontal gyrus, insula)(Harle et al., 2019) and memory (e.g. hippocampus, parahippocampal gyrus)(Stewart et al., 2014a) than people at a lower risk of relapse, which may limit how effectively they can recall and update internal belief models with new information, subsequently predisposing them towards inflexible learning behaviours. However, greater abstinence from METH in people with METH use has been associated with partial recovery in striatal function.(Bischoff-Grethe et al., 2017)

Some studies have also used pharmacological agents to potentially elucidate or remediate the effects of METH, given previous research indicating that METH increases synaptic dopamine availability.(Hedges et al., 2018) Bernacer et al.(Bernacer et al., 2013) reported that while acute METH infusions given to people with METH use led to disrupted encoding of incentive value signals in the PFC (i.e. how the value of actions and stimuli are represented neurally) and reward prediction error in the striatum (i.e. how differences between predicted and actual decision outcomes are detected and reconciled), amisulpride (a dopamine D2 antagonist) did not attenuate the effects of METH, suggesting that METH may also operate either via D1 dopamine receptors and/or using other monoamines (e.g. serotonin, noradrenaline). Meanwhile, Ghahremani et al.(Ghahremani et al., 2011) reported that modafinil, an analeptic stimulant, enhanced learning performance in people with METH use, possibly through increasing activation in important learning and cognitive control centres, such as the insula, ACC, ventrolateral PFC, and inferior frontal gyrus.

### Stroop effect

The Stroop test measures how effectively a person can inhibit cognitive interference.(Scarpina and Tagini, 2017) The most common variant (the Stroop Colour Word Test) involves asking subjects to identify the font colours of words which may be consistent with the font (i.e. congruent conditions, such as the word “blue” displayed in blue font) or inconsistent (i.e. incongruent conditions, such as the word “red” displayed in blue font). The incongruent condition thus generates cognitive conflict, where subjects must suppress a more automated process (i.e. reading the word) in order to conduct a less automated task (i.e. naming the font colour), and can be used to measure various cognitive functions, including inhibitory control, cognitive flexibility, and attention. Overall, METH use is associated with poorer performance on the Stroop task. This has been linked to widespread neural dysfunction, including areas managing cognitive control (e.g. PFC, frontal cortex),(Nestor et al., 2011; Salo et al., 2013; Salo et al., 2009a) vision (e.g. occipital cortex),(Nestor et al., 2011) conflict resolution (e.g. parietal and temporal cortices),(Jan et al., 2014; Nestor et al., 2011) interoception (e.g. insula),(Nestor et al., 2011), and complex motor planning (e.g. supplementary motor cortex),(Nestor et al., 2011) which has, in some cases, led to compensatory recruitment of additional neural resources.(Fassbender et al., 2015; Ghavidel et al., 2020b; Jan et al., 2014) People with both METH use and MAP have demonstrated poorer task performance and increased activation of the right PFC, which has been associated with vigilance and arousal behaviour.(Fassbender et al., 2015) Importantly, conflicting findings have been reported as to whether METH use is associated with changes in ACC function –which is involved in reward processing, incentive salience, task performance during risky decisions, and connects with the dlPFC via the executive network to regulate attention, inhibition control, and other cognitive functions(Ghavidel et al., 2020a) – with some data suggesting that METH use is linked to reduced ACC activation(Ghavidel et al., 2020a) and other studies reporting that there is no relationship.(Jan et al., 2014; Salo et al., 2009b) Methylphenidate has been shown to improve Stroop task performance and correct brain activation patterns in people with METH use to be comparable to CON, specifically by increasing activity in the dlPFC and decreasing activity in the parietal and occipital cortices).(Jan et al., 2014)

### Sensorimotor function

Only two studies formally evaluated the relationship on task-fMRI between METH exposure and sensorimotor tasks. Archibald et al.(Archibald et al., 2012) reported that during complex motor tasks, people with METH use had reduced activation of the cerebellar vermis and sensorimotor and parietal cortices but greater activation of the right temporal lobe and cerebellum. Meanwhile, van Hedger et al.(Van Hedger et al., 2019) reported that in people without METH use, acute administration of METH enhanced sensory-attentional integration for simple, but not complex, images by increasing activity in heteromodal association cortices involved in higher-order tertiary processing. Overall, these findings suggest a complex relationship between both acute and chronic METH exposure and sensorimotor neural activation.

### Memory

Our search returned three studies that examined the impact of METH on memory tasks, all of which specifically evaluated children who had been prenatally exposed to METH, and suggested that METH exposure in-utero was associated with impairments in both verbal and working memory. Lu et al.(Lu et al., 2009) reported that children who were prenatally exposed to METH had more diffuse brain activation during verbal memory tasks (involving the superior temporal gyrus and inferior and medial temporal structures) than children who were not, suggesting both an immature and ineffective verbal memory system necessitating generalised neural recruitment. Moreover, prenatal METH exposure has also been associated with modulation of frontostriatal network connectivity, which is typically partitioned into frontal-caudate (which engages working memory) and frontal-putamen (which engages motor tasks), such that the frontal-putamen loop becomes more active during working memory tasks.(Roussotte et al., 2011; Roussotte et al., 2012) Roussotte et al.(Roussotte et al., 2012) theorised that while these changes may reflect neural compensation for METH-induced damage of the frontal-caudate loop during gestation, they may also lead towards maladaptive overlap of motor and cognitive networks, and raised concerns that this loss of functional segregation may impede effective suppression of interfering or competing networks during directed tasks.

## Summary

In summary, METH is a highly neurotoxic agent that is associated with significant short- and long-term harm on brain structure and function in both MAA and children who were prenatally exposed to METH. Structural MRI studies have reported that chronic exposure to METH is linked to multi-region reductions in grey matter volume and thickness, particularly in the frontal and limbic areas, along with temporal and occipital white matter hypertrophy, and that some of these changes may not manifest until later stages of use or abstinence. dMRI studies have noted abnormal diffusivity indices, particularly reduced FA, in numerous subcortical white matter regions among MAA, including the amygdala, striatum, hippocampus, and corpus callosum, and prenatal exposure to METH in children can damage both specific white matter tracts as well as whole-brain fibre networks. Regarding fMRI, rs-fMRI studies have reported METH-associated changes across multiple neural networks, including the DMN, SN, and cerebellar, mesocorticolimbic, and motive networks, which may be broadly linked to deficits in cognition, motivation, self-referential and reward processing, and maladaptive interpersonal behaviours. Task-fMRI studies have similarly shown various functional deficiencies in MAA when compared to healthy controls, including emotional dysregulation and impaired socio-emotional cognition, risky decision-making with limited consideration of negative potential consequences, inflexible feedback processing and learning patterns, and cognitive deficits more broadly.

With regards to sex, it remains challenging to confidently assert whether METH-related neural effects differ across males and females. For instance, in dMRI studies, Chung et al.(Chung et al., 2007) reported that METH use was associated with greater damage in frontal white matter for males than females, while Tobias et al.(Tobias et al., 2010) found that females exhibited more diffuse METH-related white matter deficits across prefrontal, corpus callosal, and corona radiatae regions than males. In contrast, Salo et al.(Salo et al., 2009a) found no significant difference between sexes when assessing METH-related corpus callosal damage. The reason behind these inconsistent findings remains unclear, but may relate to the presence of other confounding factors (e.g. clinical parameters of METH use, duration of abstinence prior to imaging) that may conflate, dampen, or mask any potential sex-specific effects. Further, since most studies comprise predominantly or exclusively male participants and/or often neglect to conduct sex-specific analyses, it remains challenging to further explore any differential impact of METH across sexes within the current literature.

Although most studies focus on exploring the neurotoxic effects of METH, some evidence suggests that maintained abstinence from METH may lead to structural recovery in some cortical and limbic regions. Further, previous studies have proposed that cue-exposure interventions, naltrexone, and transcranial PFC stimulation may reduce METH cravings and that modafinil may enhance learning performance.

## Limitations

Several limitations restrict the generalisability of these findings. Firstly, the inconsistent inclusion or exclusion of participants with historical or ongoing dependencies on other substances aside from METH can make it challenging to distinguish METH-specific effects from other potentially neurotoxic agents. Polydrug use is common amongst METH users(Crummy et al., 2020) and has been frequently identified as a limitation of the literature in previous reviews investigating METH-related neural changes.(Moghaddam et al., 2021; Sabrini et al., 2019) While various methods can be employed to reduce the confounding impact of polydrug use, such as excluding participants who use substances other than METH, recruiting participants who only use METH as a separate analysis group, multivariable adjustment, or post-hoc analyses based on polydrug status, the inclusion or extent of use of these techniques varies considerably across studies. Secondly, there is wide variation between studies regarding the duration or magnitude of METH use or duration of abstinence, which can make it challenging to differentiate between the acute, chronic, and acute-on-chronic neurotoxic effects of METH. Most of the study populations included in our review comprised long-term current or historical users of METH, and while some studies investigated the immediate effects of METH infusions on otherwise drug-naïve participants, these findings may not fully reflect the acute changes seen in chronic MAA following drug use. Thirdly, the vast majority of studies were cross-sectional analyses with no follow-up, and there are few cohort studies to assess the long-term effects of METH use or abstinence. Of the longitudinal data available, most are limited to 1-year follow-up, which may not be sufficient time for the neural effects of METH to be fully realised. Further, the relative paucity of gold-standard RCT data limits the identification of causal relationships, with much of the current literature being purely correlational analyses. Finally, wide heterogeneity in the MRI analysis techniques used between studies can lead to disparate findings across brain regions within the same imaging modality, which can limit the generalisability and comparability of results.

## Future directions

Despite the limitations described above, it is clear that MRI is a powerful imaging technique that can sensitively detect METH-associated neural damage across a range of modalities. Most of the literature currently available has been conducted using conventional MRI modalities, which can be beneficial in that they are often compatible with clinical MRI scanners and can be relatively simple to interpret. However, higher-order MRI sequences, such as neurite orientation dispersion and density imaging or diffusion kurtosis imaging, can provide greater detail about microstructural cellular architecture and organisation and are emerging as new tools in the MRI toolbox,(Hansen, 2020; Lakhani et al., 2020; Zhang et al., 2012) although they have yet to become standard imaging techniques in METH research. Targeted MRI sequences, such as Multiplied, Added, Subtracted and/or Divided Inversion Recovery (MASDIR), may also be useful in enhancing interpretability of images and detecting subtle pathological changes (e.g. neuroinflammation associated with METH toxicity).(Ma et al., 2022; Ma et al., 2023) Finally, multimodal MRI can provide comprehensive and complementary neuroimaging data that minimises inter-subject variability and enables deeper analysis between brain structure and function.(Hao et al., 2013; Meade et al., 2021)

## Conclusion

In conclusion, we conducted a comprehensive systematic review of the literature examining the neural effects of METH as detected by MRI. We found that METH is associated with a range of deleterious macro- and microstructural and functional abnormalities across both individual brain regions and functionally connected neural networks, which can subsequently translate into cognitive, behavioural, socio-emotional, and motor deficits. Future studies should aim to extend current findings by conducting methodologically rigorous studies with adequate duration of follow-up and consider the use of multimodal, targeted, or higher-order MRI modalities which may provide greater neuroimaging detail.

## Data Availability

All data produced in the present study are available upon reasonable request to the authors

## Acknowledgements

Nil.

