## Supplementary file 1 for "How does methamphetamine affect the brain? A systematic review of magnetic resonance imaging studies"

### Supplementary file 1: Search strategy

#### Methamphetamine (METH) AND magnetic resonance imaging (MRI)

*PubMed*

1. "Methamphetamine"[Title/Abstract] OR "Desoxyephedrine"[Title/Abstract] OR "Metamfetamine"[Title/Abstract] OR "Methylamphetamine"[Title/Abstract] OR "n methylamphetamine"[Title/Abstract] OR "n methylamphetamine"[Title/Abstract] OR "Deoxyephedrine"[Title/Abstract] OR "Desoxyn"[Title/Abstract] OR "Methamphetamine Hydrochloride"[Title/Abstract]
2. "Quantitative imaging"[Title/Abstract] OR "Quantitative MR imaging"[Title/Abstract] OR "Quantitative MRI imaging"[Title/Abstract] OR "MRI"[Title/Abstract] OR "Magnetic resonance image"[Title/Abstract] OR "Magnetic resonance images"[Title/Abstract] OR "Magnetic resonance imaging"[Title/Abstract]
3. "english"[Language]
4. "pubmed books"[Filter] OR "case reports"[Publication Type] OR "meta analysis"[Publication Type] OR "review"[Publication Type] OR "systematic review"[Filter]
5. 1 AND 2 AND 3 NOT 4

*Scopus*

1. TITLE-ABS ( methamphetamine OR desoxyephedrine OR metamfetamine OR methylamphetamine OR n-methylamphetamine OR {N Methylamphetamine} OR deoxyephedrine OR desoxyn OR {Methamphetamine Hydrochloride} )
2. TITLE-ABS ( {Quantitative imaging} OR {Quantitative MR imaging} OR {Quantitative MRI imaging} OR mri OR {Magnetic resonance image} OR {Magnetic resonance images} OR {Magnetic resonance imaging} )
3. LANGUAGE , "English"
4. EXCLUDE ( ( DOCTYPE , "re" ) OR ( DOCTYPE , "ch" ) OR ( DOCTYPE , "bk" ) OR ( DOCTYPE , "cr" ) )
5. 1 AND 2 AND 3 AND 4

*Web of Science Core Collection*

1. TI=(Methamphetamine) OR TI=(Desoxyephedrine) OR TI=(Metamfetamine) OR TI=(Methylamphetamine) OR TI=("N-Methylamphetamine") OR TI=("N Methylamphetamine") OR TI=(Deoxyephedrine) OR TI=(Desoxyn) OR TI=("Methamphetamine Hydrochloride") OR AB=(Methamphetamine)) OR AB=(Desoxyephedrine)) OR AB=(Metamfetamine) OR AB=(Methylamphetamine) OR AB=("N-Methylamphetamine") OR AB=("N Methylamphetamine") OR AB=(Deoxyephedrine) OR AB=(Desoxyn) OR AB=("Methamphetamine Hydrochloride")
2. TI=("Quantitative imaging") OR TI=("Quantitative MR imaging") OR TI=("Quantitative MRI imaging") OR TI=(MRI) OR TI=("Magnetic resonance image") OR TI=("Magnetic resonance images") OR TI=("Magnetic resonance imaging") OR AB=("Quantitative imaging") OR AB=("Quantitative MR imaging") OR AB=("Quantitative MRI imaging") OR AB=(MRI) OR AB=("Magnetic resonance image") OR AB=("Magnetic resonance images") OR AB=("Magnetic resonance imaging")
3. LA==("ENGLISH")
4. DT==("REVIEW" OR "MEETING ABSTRACT" OR "EDITORIAL MATERIAL")
5. 1 AND 2 AND 3 NOT 4

#### Methamphetamine (METH) AND diffusion magnetic resonance imaging (dMRI)

*PubMed*

1. "Methamphetamine"[Title/Abstract] OR "Desoxyephedrine"[Title/Abstract] OR "Metamfetamine"[Title/Abstract] OR "Methylamphetamine"[Title/Abstract] OR "n methylamphetamine"[Title/Abstract] OR "n methylamphetamine"[Title/Abstract] OR "Deoxyephedrine"[Title/Abstract] OR "Desoxyn"[Title/Abstract] OR "Methamphetamine Hydrochloride"[Title/Abstract]
2. "Diffusion MRI"[Title/Abstract] OR "Diffusion Weighted MRI"[Title/Abstract] OR "Diffusion Tensor Imaging"[Title/Abstract] OR "Diffusion Tensor Magnetic Resonance Imaging"[Title/Abstract] OR "Diffusion Tensor MRI"[Title/Abstract] OR "DTI"[Title/Abstract] OR “DWI”[Title/Abstract] OR "Tractography"[Title/Abstract])

3. "english"[Language]
4. "pubmed books"[Filter] OR "case reports"[Publication Type] OR "meta analysis"[Publication Type] OR "review"[Publication Type] OR "systematic review"[Filter]
5. 1 AND 2 AND 3 NOT 4

*Scopus*

1. TITLE-ABS ( methamphetamine OR desoxyephedrine OR metamfetamine OR methylamphetamine OR n-methylamphetamine OR {N Methylamphetamine} OR deoxyephedrine OR desoxyn OR {Methamphetamine Hydrochloride} )
2. TITLE-ABS ( {Diffusion MRI} OR {Diffusion Weighted MRI} OR {Diffusion Tensor Imaging} OR {Diffusion Tensor Magnetic Resonance Imaging} OR {Diffusion Tensor MRI} OR dti OR dwi OR tractography)
3. LIMIT-TO ( LANGUAGE , "English" )
4. EXCLUDE ( ( DOCTYPE , "re" ) OR ( DOCTYPE , "ch" ) OR ( DOCTYPE , "bk" ) OR ( DOCTYPE , "cr" ) )
5. 1 AND 2 AND 3 AND 4

*Web of Science Core Collection*

1. TI=(Methamphetamine) OR TI=(Desoxyephedrine) OR TI=(Metamfetamine) OR TI=(Methylamphetamine) OR TI=("N-Methylamphetamine") OR TI=("N Methylamphetamine") OR TI=(Deoxyephedrine) OR TI=(Desoxyn) OR TI=("Methamphetamine Hydrochloride") OR AB=(Methamphetamine) OR AB=(Desoxyephedrine) OR AB=(Metamfetamine) OR AB=(Methylamphetamine) OR AB=("N-Methylamphetamine") OR AB=("N Methylamphetamine") OR AB=(Deoxyephedrine) OR AB=(Desoxyn) OR AB=("Methamphetamine Hydrochloride")
2. TI=("Diffusion MRI") OR TI=("Diffusion Weighted MRI") OR TI=("Diffusion Tensor Imaging") OR TI=(“Diffusion Tensor Magnetic Resonance Imaging”) OR TI=("Diffusion Tensor MRI") OR TI=(DTI) OR TI=(DWI) OR TI=(Tractography) OR AB=("Diffusion MRI") OR AB=("Diffusion Weighted MRI") OR AB=("Diffusion Tensor Imaging") OR AB=(“Diffusion Tensor Magnetic Resonance Imaging”) OR AB=("Diffusion Tensor MRI") OR AB=(DTI) OR AB=(DWI) OR AB=(Tractography)

3. LA==("ENGLISH")
4. DT==("REVIEW" OR "MEETING ABSTRACT" OR "EDITORIAL MATERIAL")
5. 1 AND 2 AND 3 NOT 4

#### Methamphetamine (METH) AND functional magnetic resonance imaging (fMRI)

*PubMed*

1. "Methamphetamine"[Title/Abstract] OR "Desoxyephedrine"[Title/Abstract] OR "Metamfetamine"[Title/Abstract] OR "Methylamphetamine"[Title/Abstract] OR "n methylamphetamine"[Title/Abstract] OR "n methylamphetamine"[Title/Abstract] OR "Deoxyephedrine"[Title/Abstract] OR "Desoxyn"[Title/Abstract] OR "Methamphetamine Hydrochloride"[Title/Abstract]
2. "fMRI"[Title/Abstract] OR "Functional MRI"[Title/Abstract] OR "Functional Magnetic Resonance Imaging"[Title/Abstract] OR "Functional Brain Imaging"[Title/Abstract] OR "N-back task"[Title/Abstract]
3. "english"[Language]
4. "pubmed books"[Filter] OR "case reports"[Publication Type] OR "meta analysis"[Publication Type] OR "review"[Publication Type] OR "systematic review"[Filter]
5. 1 AND 2 AND 3 NOT 4

*Scopus*

1. TITLE-ABS ( methamphetamine OR desoxyephedrine OR metamfetamine OR methylamphetamine OR n-methylamphetamine OR {N Methylamphetamine} OR deoxyephedrine OR desoxyn OR {Methamphetamine Hydrochloride} )
2. TITLE-ABS ( fMRI OR {Functional MRI} OR {Functional Magnetic Resonance Imaging} OR {Functional Brain Imaging} OR {N-back task} )
3. LIMIT-TO ( LANGUAGE , "English" )
4. EXCLUDE ( ( DOCTYPE , "re" ) OR ( DOCTYPE , "ch" ) OR ( DOCTYPE , "bk" ) OR ( DOCTYPE , "cr" ) )

5. 1 AND 2 AND 3 AND 4

*Web of Science Core Collection*

1. TI=(Methamphetamine) OR TI=(Desoxyephedrine) OR TI=(Metamfetamine) OR TI=(Methylamphetamine) OR TI=("N-Methylamphetamine") OR TI=("N Methylamphetamine") OR TI=(Deoxyephedrine) OR TI=(Desoxyn) OR TI=("Methamphetamine Hydrochloride") OR AB=(Methamphetamine) OR AB=(Desoxyephedrine) OR AB=(Metamfetamine) OR AB=(Methylamphetamine) OR AB=("N-Methylamphetamine") OR AB=("N Methylamphetamine") OR AB=(Deoxyephedrine) OR AB=(Desoxyn) OR AB=("Methamphetamine Hydrochloride")
2. TI=("fMRI") OR TI=("Functional MRI") OR TI=("Functional Magnetic Resonance Imaging") OR TI=("Functional Brain Imaging") OR TI=("N-back task") OR AB=("fMRI") OR AB=("Functional MRI") OR AB=("Functional Magnetic Resonance Imaging") OR AB=("Functional Brain Imaging") OR AB=("N-back task")
3. LA==("ENGLISH")
4. DT==("REVIEW" OR "MEETING ABSTRACT" OR "EDITORIAL MATERIAL")
5. 1 AND 2 AND 3 NOT 4

#### Methamphetamine (METH) and structural magnetic resonance imaging (structural MRI)

*PubMed*

1. "Methamphetamine"[Title/Abstract] OR "Desoxyephedrine"[Title/Abstract] OR "Metamfetamine"[Title/Abstract] OR "Methylamphetamine"[Title/Abstract] OR "n methylamphetamine"[Title/Abstract] OR "n methylamphetamine"[Title/Abstract] OR "Deoxyephedrine"[Title/Abstract] OR "Desoxyn"[Title/Abstract] OR "Methamphetamine Hydrochloride"[Title/Abstract]
2. "Structural imaging"[Title/Abstract] OR "Anatomic imaging"[Title/Abstract] OR "Anatomical imaging"[Title/Abstract] OR volumetric[Title/Abstract] OR "MR Imaging"[Title/Abstract] OR "T1 weighted imaging"[Title/Abstract] OR "T1 imaging"[Title/Abstract] OR morphomet*[Title/Abstract]

3. "english"[Language]
4. "pubmed books"[Filter] OR "case reports"[Publication Type] OR "meta analysis"[Publication Type] OR "review"[Publication Type] OR "systematic review"[Filter]
5. 1 AND 2 AND 3 NOT 4

*Scopus*

1. TITLE-ABS ( methamphetamine OR desoxyephedrine OR metamfetamine OR methylamphetamine OR n-methylamphetamine OR {N Methylamphetamine} OR deoxyephedrine OR desoxyn OR {Methamphetamine Hydrochloride} )
2. TITLE-ABS ( {Structural Imaging} OR {Anatomic Imaging} OR {Anatomical Imaging} OR volumetric OR {MR Imaging} OR {T1 weighted imaging} OR {T1 imaging} OR {morphomet*} )
3. LIMIT-TO ( LANGUAGE , "English" )
4. EXCLUDE ( ( DOCTYPE , "re" ) OR ( DOCTYPE , "ch" ) OR ( DOCTYPE , "bk" ) OR ( DOCTYPE , "cr" ) )
5. 1 AND 2 AND 3 AND 4

*Web of Science Core Collection*

1. TI=(Methamphetamine) OR TI=(Desoxyephedrine) OR TI=(Metamfetamine) OR TI=(Methylamphetamine) OR TI=("N-Methylamphetamine") OR TI=("N Methylamphetamine") OR TI=(Deoxyephedrine) OR TI=(Desoxyn) OR TI=("Methamphetamine Hydrochloride") OR AB=(Methamphetamine) OR AB=(Desoxyephedrine) OR AB=(Metamfetamine) OR AB=(Methylamphetamine) OR AB=("N-Methylamphetamine") OR AB=("N Methylamphetamine") OR AB=(Deoxyephedrine) OR AB=(Desoxyn) OR AB=("Methamphetamine Hydrochloride")
2. TI=("Structural Imaging") OR TI=("Anatomic Imaging") OR TI=("Anatomical Imaging") OR TI=(Volumetric) OR TI=("MR Imaging") OR TI=(“T1 weighted imaging”) OR TI=(“T1 imaging”) OR TI=(morphomet*) OR AB=("Structural Imaging") OR AB=("Anatomic Imaging") OR AB=("Anatomical Imaging") OR AB=(“Volumetric”) OR AB=("MR Imaging”) OR AB=(“T1 weighted imaging”) OR AB=(“T1 imaging”) OR AB=(morphomet*)

3. LA==("ENGLISH")

4. DT==("REVIEW" OR "MEETING ABSTRACT" OR "EDITORIAL MATERIAL")

5. 1 AND 2 AND 3 NOT 4
