## Supplementary file 2 for "How does methamphetamine affect the brain? A systematic review of magnetic resonance imaging studies"

### Supplementary file 2: Data extraction proforma

| **Domain** | **Data field** |
|---|---|
| Study | Title |
| | Record number |
| | 1st author |
| | Year |
| | County |
| | Aims + hypothesis |
| Design | Study design |
| | Retrospective/prospective |
| | Follow-up (Y/N) |
| | Follow-up duration |
| Population (METH) | Age group |
| | Age range (years) |
| | Sex |
| | Description |
| | Duration of abstinence |
| | Sample size |

| | |
|---|---|
| | Location of recruitment |
| | Inclusion criteria |
| | Exclusion criteria |
| | Duration of METH use |
| | Other drug use (Y/N) |
| | Other drug use (list) |
| | Other drug use (extent) |
| Population (comparison) | Age group |
| | Age range (years) |
| | Sex |
| | Description |
| | Duration of abstinence |
| | Sample size |
| | Location of recruitment |
| | Inclusion criteria |
| | Exclusion criteria |
| | Duration of METH use |
| | Other drug use (Y/N) |

| | |
|---|---|
| | Other drug use (list) |
| | Other drug use (extent) |
| Population (total) | Total sample size |
| MRI | Magnet |
| | Modality |
| | Image processing technique |
| | Task (if task-fMRI) |
| Findings (grouped by modality) | Structural |
| | Diffusion |
| | Resting-state functional |
| | Task-based functional |
| | Other |
| Notes | Notes |
| Data collector | Data collector |
