## Supplementary file 3 for "How does methamphetamine affect the brain? A systematic review of magnetic resonance imaging studies"

### Supplementary file 3: Quality appraisal questionnaire

| Criteria | Number of points that could be scored | Scoring guide |
|---|---|---|
| **1. Was the study aim clearly stated?** | 1 | 1 point was allocated if the study had a clearly stated research aim. |
| **2. Was the study population clearly defined?** | 2 or 3 | 1 point was allocated if both age and sex of participants were reported.<br><br>1 point was allocated if information on polydrug (e.g. nicotine, alcohol, cocaine, stimulants) use (e.g. presence/absence, duration of use, quantity consumed, etc.) OR history of mental health conditions was reported.<br><br>1 point was allocated if either the route or duration of METH use by participants (or participants' mothers if study was conducted on children exposed to METH in-utero) was reported. This would be classed as "N/A" if study participants had no prior history of METH exposure (e.g. healthy volunteers). |
| **3. Were control participants clearly defined and had no prior history of METH use?** | 2 | 1 point was allocated if a control group was included.<br><br>1 point was allocated if the control group had no history of previous METH use. |
| **4. Were inclusion and exclusion criteria for study participants thoroughly defined and applied?** | 2 | 1 point was allocated if inclusion/exclusion criteria were reported. |

| | | |
|---|---|---|
| | | 1 point was allocated if inclusion/exclusion criteria were reported exhaustively and applied uniformly |
| **5. Was the duration of abstinence from METH use provided?** | 0 or 1 | 1 point was allocated if the study reported duration of abstinence from last time of METH use to time of study recruitment/MRI scan.<br><br>Classed as "N/A" if study participants did not use METH (e.g. children who were exposed to METH prenatally, healthy volunteers, etc.) |
| **6. Did the study examine different levels of METH exposure as related to differential brain changes on MRI?** | 0 or 1 | 1 point was allocated if the study investigated differential brain MRI changes as related to different levels of METH exposure.<br><br>Classed as “N/A” if the study aim did not pre-specify this as a research objective. |
| **7. Was severity of METH exposure quantified using clearly defined and valid study measures?** | 0 or 2 | 1 point was allocated if severity of METH exposure was categorised in study participants (e.g. METH dependence, METH abuse).<br><br>1 point was allocated if severity of METH consumption was categorised in study participants using standardised guidelines and/or diagnostic criteria (e.g. DSM-4, DSM-5, ICD-10).<br><br>Classed as "N/A" if study participants had no prior history of METH exposure (e.g. healthy volunteers). |
| **8. Were multiple MRI scans conducted over time?** | 0 or 1 | 1 point was allocated if MRI scans were obtained over multiple timepoints. |

| | | |
|---|---|---|
| | | Classed as "N/A" if the study was cross-sectional. |
| **9. Was loss to follow-up after baseline 20% or less?** | 0 or 1 | 1 point was allocated when loss to follow-up after baseline was 20% or less for the entire study population<br><br>Classified as 'N/A' if MRI assessment was cross-sectional. |
| **10. Were the MRI scan and pre-processing protocols, and findings clearly and consistently reported?** | 3 | 1 point was allocated if the MRI scan protocol was clearly reported.[†]<br><br>1 point was allocated if the MRI pre-processing protocol was clearly reported.[‡]<br><br>1 point was allocated if MRI protocols were consistently applied and results were appropriately adjusted for multiple comparisons.[§] |
| **11. Were key potential confounding variables adjusted for during statistical analysis?** | 1 | 1 point was allocated if at least one of the following confounding variables were statistically adjusted for:<br><br>• Age<br>• Sex<br>• History of head injury<br>• Smoking<br>• Alcohol<br>• Recreational drug use except for METH<br>• History of family addiction<br>• History of mental health issues |

*Minimum and maximum total scores of 11 and 18 respectively. DSM-4: Diagnostic and Statistical Manual of Mental Disorders, 4th Edition. DSM-5: Diagnostic and Statistical Manual of Mental Disorders, 5th Edition. ICD-10: International Statistical Classification of Diseases and Related Health Problems, 10th revision. METH: Methamphetamine. MRI: Magnetic resonance imaging. N/A: Not applicable.*

†*A study must report the following parameters to receive 1 point for having a clear MRI scan protocol:*

- *Type of scan*
- *Repetition time (TR)*
- *Echo time (TE)*
- *Flip angle*
- *Number of slices OR slice gap OR slice thickness*
- *Analysis type*
- *≥2 of the following: field of view (FOV), matrix, voxel size*

‡*A study must report the following parameters to receive 1 point for having a clear MRI pre-processing protocol:*

- *Computer program used for pre-processing*
- *Motion correction*
- *Slice-time correction (or eddy correction if diffusion MRI scan)*
- *Smoothing*

§*A study must fulfil the following criteria to receive 1 point for having a consistently applied MRI protocol and appropriate adjustment for multiple comparisons:*

- *Protocol consistently applied to all participants, or reasons are clearly stated for participants where the protocol was not consistently applied*
- *Corrections of multiple comparisons or analytic pipelines that do not reduce the total number of comparisons were done*
