## Supplementary table 1 for "How does methamphetamine affect the brain? A systematic review of magnetic resonance imaging studies"

### Supplementary table 1: Study demographics and quality appraisal score for studies with MRI modalities not covered in tables 1a-d or with combinations of different MRI modalities

| Reference | Country | Study design | Age group, sex | Experimental group | Comparison group | N | MRI modality | Quality appraisal score |
|---|---|---|---|---|---|---|---|---|
| Pang et al. 2021(Pang and Peng, 2021) | China | Cross-sectional | Adults, both | MAA | Healthy controls | 54 | MRS | 86.67% |
| Salo et al. 2011(Salo et al., 2011) | United States | Cross-sectional | Adults, both | MAA | Healthy controls | 71 | MRS | 100.00% |
| Lin et al. 2015(Lin et al., 2015) | New Zealand | Cross-sectional | Adults, both | MAA | Healthy controls | 40 | MRS and dMRI† | 93.33% |

| | | | | | | | | |
|---|---|---|---|---|---|---|---|---|
| Smith et al. 2001(Smith et al., 2001) | United States | Cross-sectional | Children, sex NR | Prenatal METH exposure | Healthy controls | 26 | MRS and Structural | 85.71% |
| Wu et al. 2018(Wu et al., 2018) | China | Cross-sectional | Adults, both | MAA | Healthy controls | 126 | MRS and Structural | 93.75% |
| Vuletic et al. 2018(Vuletic et al., 2018) | South Africa | Cross-sectional | Adults, both | MAA | Healthy controls | 31 | pCASL | 86.67% |
| Li et al. 2019(Li et al., 2019) | China | Cross-sectional | Adults, male | MAA | Healthy controls | 126 | pCASL | 93.33% |
| Chang et al. 2002(Chang et al., 2002) | United States | Cross-sectional | Adults, both | MAA | Healthy controls | 40 | Perfusion MRI and Structural | 100.00% |

| | | | | | | | | |
|---|---|---|---|---|---|---|---|---|
| Yang et al. 2021(Yang et al., 2021) | China | Cross-sectional | Adults, male | MAA | Healthy controls | 70 | rs-fMRI and Structural | 100.00% |

*†Diffusivity parameters were AD, FA, MD, RD. MAA includes a history of current or historical METH abuse, dependence, or use disorder, as diagnosed by the Diagnostic and Statistical Manual of Mental Disorders 4th Edition, Diagnostic and Statistical Manual of Mental Disorders 5th Edition, or International Classification of Diseases 10$^{th}$ edition diagnostic codes. Healthy controls defined as participants without adult or prenatal METH exposure. AD: Axial diffusivity. ADC: Apparent diffusion coefficient. dMRI: Diffusion magnetic resonance imaging. FA: Fractional anisotropy. MD: Mean diffusivity. MAA: Adult METH user. METH: Methamphetamine. MRI: Magnetic resonance imaging. NR: Not reported. N/A: Not applicable. pCASL: Pseudo continuous arterial spin labelling. RD: Radial diffusivity. rs-fMRI: Resting-state functional magnetic resonance imaging.*
