## Supplementary table 2 for "How does methamphetamine affect the brain? A systematic review of magnetic resonance imaging studies"

### Supplementary table 2: Study characteristics and quality appraisal score calculations

| Reference | Study design | Age group, sex | Experimental group | Comparison group | MRI sequence | Quality appraisal: Total score | Quality appraisal: Points scored | Quality appraisal: Total potential score |
|---|---|---|---|---|---|---|---|---|
| Alaee et al. 2014(Alaee et al., 2014) | Cross-sectional | Adults, both | MAA | Healthy controls | Structural | 80.00% | 12 | 15 |
| Alicata et al. 2009(Alicata et al., 2009) | Cross-sectional | Adults, both | MAA | Healthy controls | dMRI | 93.75% | 15 | 16 |
| Andres et al. 2016(Andres et al., 2016) | Cross-sectional | Adults, both | MAA | Healthy controls | dMRI | 100.00% | 15 | 15 |
| Aoki et al. 2013(Aoki et al., 2013) | Cross-sectional | Adults, both | METH-associated psychosis | Healthy controls | Structural | 86.67% | 13 | 15 |

| | | | | | | | | |
|---|---|---|---|---|---|---|---|---|
| Archibald et al. 2012(Archibald et al., 2012) | Cross-sectional | Adults, both | MAA | Healthy controls | task-fMRI | 87.50% | 14 | 16 |
| Bae et al. 2006(Bae et al., 2006) | Cross-sectional | Adults, both | MAA | Healthy controls | Structural | 100.00% | 15 | 15 |
| Bernacer et al. 2013(Bernacer et al., 2013) | Single-arm interventional trial | Adults, both | Healthy volunteers who received METH | N/A | task-fMRI | 63.64% | 7 | 11 |
| Bischoff-Grethe et al. 2021(Bischoff-Grethe et al., 2021) | Cross-sectional | Adults, both | MAA | Healthy controls or HIV-positive | task-fMRI | 93.75% | 16 | 16 |
| Bischoff-Grethe et al. 2017(Bischoff- | Cross-sectional | Adults, both | MAA | Healthy controls | task-fMRI | 100.00% | 15 | 16 |

| | | | | | | | | |
|---|---|---|---|---|---|---|---|---|
| Grethe et al., 2017) | | | | | | | | |
| Breen et al. 2017(Breen et al., 2017) | Cross-sectional | Adults, both | MAA | Healthy controls | dMRI | 93.33% | 14 | 15 |
| Brooks et al. 2016(Brooks et al., 2016) | RCT (quasi) | Adults, male | MAA | Healthy controls | Structural | 100.00% | 17 | 17 |
| Chang et al. 2002(Chang et al., 2002) | Cross-sectional | Adults, both | MAA | Healthy controls | Structural and Perfusion MRI | 100.00% | 15 | 15 |
| Chang et al. 2004(Chang et al., 2004) | Cross-sectional | Children, both | Prenatal METH exposure | Healthy controls | Structural | 92.86% | 13 | 14 |

| | | | | | | | | |
|---|---|---|---|---|---|---|---|---|
| Chang et al. 2005(Chang et al., 2005) | Cross-sectional | Adults, both | MAA | Healthy controls | Structural | 100.00% | 15 | 15 |
| Chang et al. 2016(Chang et al., 2016) | Cohort | Infants, both | Prenatal METH exposure | Healthy controls | dMRI | 93.75% | 15 | 16 |
| Chen et al. 2020(Chen et al., 2020) | Cross-sectional | Adults, male | MAA | Healthy controls | task-fMRI | 93.75% | 15 | 16 |
| Chung et al. 2007(Chung et al., 2007) | Cross-sectional | Adults, both | MAA | Healthy controls | dMRI | 100.00% | 15 | 15 |
| Churchwell et al. 2012(Churchwell et al., 2012) | Cross-sectional | Adolescents, both | MAA | Healthy controls | Structural | 68.75% | 11 | 16 |

| | | | | | | | | |
|---|---|---|---|---|---|---|---|---|
| Cloak et al. 2009(Cloak et al., 2009) | Cross-sectional | Children, both | Prenatal METH exposure | Healthy controls | dMRI | 85.71% | 12 | 14 |
| Colby et al. 2012(Colby et al., 2012) | Cross-sectional | Children, both | Prenatal METH exposure | Healthy controls | dMRI | 85.71% | 12 | 14 |
| Courtney et al. 2016(Courtney et al., 2016) | RCT (crossover) | Adults, both | MAA | N/A | task-fMRI | 80.00% | 12 | 15 |
| Dadpour et al. 2019(Dadpour et al., 2019) | Cohort | Adults/adolescents, male | MAA | Healthy controls | Structural | 80.00% | 12 | 15 |
| Dean et al. 2015(Dean et al., 2015) | Cross-sectional | Adults, both | MAA | N/A | rs-fMRI | 80.00% | 12 | 16 |

| | | | | | | | | |
|---|---|---|---|---|---|---|---|---|
| Dean et al. 2018(Dean et al., 2018) | Cross-sectional | Adults, both | MAA | Healthy controls | Structural | 100.00% | 15 | 15 |
| Dean et al. 2019(Dean et al., 2019) | RCT | Adults, both | MAA who received ABM training | MAA who did not receive ABM training | task-fMRI | 76.47% | 13 | 17 |
| Dean et al. 2021(Dean et al., 2021) | Cross-sectional | Adults, both | MAA | Healthy controls | Structural | 93.33% | 14 | 15 |
| Derauf et al. 2012(Derauf et al., 2012) | Cross-sectional | Children, both | Prenatal METH exposure | Healthy controls | Structural | 92.86% | 13 | 14 |
| Dong et al. 2021(Dong et al., 2021) | Cross-sectional | Adults, sex NR | MAA | Healthy controls | rs-fMRI | 40.00% | 6 | 15 |

| | | | | | | | | |
|---|---|---|---|---|---|---|---|---|
| Droutman et al. 2019(Droutman et al., 2019) | Cross-sectional | Adults, male | MAA | Healthy controls | task-fMRI | 50.00% | 8 | 16 |
| Du et al. 2021(Du et al., 2021) | Cohort | Adults, both | MAA who remained abstinent at 1-year | MAA who relapsed at 1-year | rs-fMRI | 77.78% | 14 | 18 |
| Ekhtiari et al. 2021(Ekhtiari et al., 2021) | Cross-sectional | Adults, male | MAA | MAA | task-fMRI | 68.75% | 11 | 16 |
| Farnia et al. 2018(Farnia et al., 2018) | Cross-sectional | Adults, both | MAA | Healthy controls | Structural | 60.00% | 9 | 15 |
| Farnia et al. 2020(Farnia et al., 2020) | Cross-sectional | Adults, both | MAA | Healthy controls | Structural | 93.33% | 14 | 15 |

| | | | | | | | | |
|---|---|---|---|---|---|---|---|---|
| Fassbender et al. 2015(Fassbender et al., 2015) | Cross-sectional | Adults, both | MAA | Healthy controls | task-fMRI | 86.67% | 13 | 15 |
| Fede et al. 2016(Fede et al., 2016) | Cross-sectional | Adults, male | Adult METH and cocaine exposure | Healthy controls | task-fMRI | 75.00% | 12 | 16 |
| Ghahremani et al. 2011(Ghahremani et al., 2011) | RCT (crossover) | Adults, both | MAA | Healthy controls | task-fMRI | 88.24% | 15 | 17 |
| Ghavidel et al. 2020(Ghavidel et al., 2020a) | Cross-sectional | Adults, both | MAA | Healthy controls | task-fMRI | 87.50% | 14 | 16 |
| Ghavidel et al. 2020(Ghavidel et al., 2020b) | Cross-sectional | Adults, male | MAA | Healthy controls | task-fMRI | 100.00% | 15 | 15 |

| | | | | | | | | |
|---|---|---|---|---|---|---|---|---|
| Gowin et al. 2014(Gowin et al., 2014b) | Cross-sectional | Adults, both | MAA | Healthy controls | task-fMRI | 87.50% | 14 | 16 |
| Gowin et al. 2014(Gowin et al., 2014a) | Cohort | Adults, both | MAA who remained abstinent at 1-year | MAA who relapsed at 1-year | task-fMRI | 66.67% | 12 | 18 |
| Gowin et al. 2015(Gowin et al., 2015) | Cohort | Adults, both | MAA who remained abstinent at 1-year | MAA who relapsed at 1-year | task-fMRI | 70.59% | 12 | 17 |
| Grodin et al. 2019(Grodin et al., 2019) | Cross-sectional | Adults, both | MAA | N/A | task-fMRI | N/A | 0 | 0 |
| Harle et al. 2016(Harle et al., 2016) | Cross-sectional | Adults, both | MAA | Healthy controls | task-fMRI | 86.67% | 13 | 15 |

| | | | | | | | | |
|---|---|---|---|---|---|---|---|---|
| Harle et al. 2019(Harle et al., 2019) | Cohort | Adults, both | MAA who remained abstinent at 1-year | MAA who relapsed at 1-year | task-fMRI | 76.47% | 13 | 17 |
| Heidari et al. 2017(Heidari et al., 2017) | Cross-sectional | Adults, male | MAA | Healthy controls | Structural | 73.33% | 11 | 15 |
| Hoffman et al. 2008(Hoffman et al., 2008) | Cross-sectional | Adults, both | MAA | Healthy controls | task-fMRI | 93.33% | 14 | 15 |
| Hoffman et al. 2020(Hoffman et al., 2020) | Cross-sectional | Adults, both | MAA | Healthy controls | rs-fMRI | 93.75% | 15 | 16 |
| Huang et al. 2018(Huang et al., 2018) | Cross-sectional | Adults, male | MAA | Healthy controls | task-fMRI | 100.00% | 16 | 16 |

| | | | | | | | | |
|---|---|---|---|---|---|---|---|---|
| Huang et al. 2020(Huang et al., 2020b) | Cross-sectional | Adults, both | MAA | Healthy controls | dMRI | 100.00% | 15 | 15 |
| Huang et al. 2020(Huang et al., 2020a) | Cross-sectional | Adults, male | MAA | Healthy controls | Structural | 100.00% | 15 | 15 |
| Ipser et al. 2018(Ipser et al., 2018) | Cross-sectional | Adults, male | MAA | Healthy controls | rs-fMRI | 93.75% | 15 | 16 |
| Jan et al. 2012(Jan et al., 2012) | Cross-sectional | Adults, both | MAA | Healthy controls | Structural | 93.33% | 14 | 15 |
| Jan et al. 2014(Jan et al., 2014) | RCT | Adults, both | MAA | Healthy controls | task-fMRI | 86.67% | 13 | 15 |
| Jeong et al. 2013(Jeong et al., 2013) | Cross-sectional | Adults, both | MAA | Healthy controls | Structural | 100.00% | 15 | 15 |

| | | | | | | | | |
|---|---|---|---|---|---|---|---|---|
| Jeong et al. 2021(Jeong et al., 2021) | RCT | Adults, both | MAA | Healthy controls | Structural | 100.00% | 17 | 17 |
| Jernigan et al. 2005(Jernigan et al., 2005) | Cross-sectional | Adults, both | MAA and HIV-positive | Healthy controls | Structural | 100.00% | 16 | 16 |
| Jiang et al. 2021(Jiang et al., 2021) | Cross-sectional | Adults, male | MAA | Healthy controls | rs-fMRI | 93.33% | 14 | 15 |
| Kim et al. 2006(Kim et al., 2006) | Cross-sectional | Adults, both | MAA | Healthy controls | Structural | 100.00% | 16 | 16 |
| Kim et al. 2009(Kim et al., 2009) | Cross-sectional | Adults, male | MAA | Healthy controls | dMRI | 73.33% | 11 | 15 |

| | | | | | | | | |
|---|---|---|---|---|---|---|---|---|
| Kim et al. 2010(Kim et al., 2010) | Cross-sectional | Adults, male | MAA | Healthy controls | task-fMRI | 86.67% | 13 | 15 |
| Kim et al. 2011(Kim et al., 2011) | Cross-sectional | Adults, male | MAA | Healthy controls | task-fMRI | 100.00% | 15 | 15 |
| Kogachi et al. 2017(Kogachi et al., 2017) | Cross-sectional | Adults, both | MAA | Healthy controls | Structural | 100.00% | 15 | 15 |
| Kohno et al. 2014(Kohno et al., 2014) | Cross-sectional | Adults, both | MAA | Healthy controls | task-fMRI | 93.33% | 14 | 15 |
| Kohno et al. 2016(Kohno et al., 2016)* | Cross-sectional | Adults, both | MAA | Healthy controls | rs-fMRI | 86.67% | 13 | 15 |

| | | | | | | | | |
|---|---|---|---|---|---|---|---|---|
| Kohno et al. 2016(Kohno et al., 2016)* | Cross-sectional | Adults, both | MAA | Healthy controls | rs-fMRI | 86.67% | 13 | 15 |
| Kohno et al. 2018(Kohno et al., 2018b) | Cross-sectional | Adults, both | MAA | Healthy controls | rs-fMRI | 100.00% | 15 | 15 |
| Kohno et al. 2018(Kohno et al., 2018a) | RCT | Adults, both | MAA who received naltrexone | MAA who received placebo | rs-fMRI | 77.78% | 14 | 18 |
| Kohno et al. 2019(Kohno et al., 2019) | RCT | Adults, both | MAA who received naltrexone | MAA who received placebo | rs-fMRI | 76.47% | 13 | 17 |
| Lederer et al. 2016(Lederer et al., 2016) | Cross-sectional | Adults, both | MAA | Healthy controls | dMRI | 100.00% | 15 | 15 |

| | | | | | | | | |
|---|---|---|---|---|---|---|---|---|
| Leland et al. 2008(Leland et al., 2008) | Cross-sectional | Adults, both | MAA | Healthy controls | task-fMRI | 80.00% | 12 | 15 |
| Li et al. 2017(Li et al., 2017) | Cross-sectional | Adults, male | MAA | Healthy controls | dMRI | 93.33% | 14 | 15 |
| Li et al. 2018(Li et al., 2018) | Cross-sectional | Adults, male | MAA | Healthy controls | dMRI | 93.75% | 15 | 16 |
| Li et al. 2019(Li et al., 2019) | Cross-sectional | Adults, male | MAA | Healthy controls | pCASL | 93.33% | 14 | 15 |
| Li et al. 2020(Li et al., 2020) | Cross-sectional | Adults, both | MAA | Healthy controls | rs-fMRI | 93.75% | 15 | 16 |
| Lin et al. 2015(Lin et al., 2015) | Cross-sectional | Adults, both | MAA | Healthy controls | dMRI and MRS | 93.33% | 14 | 15 |
| Liu et al. 2020(Liu et al., 2020) | Cross-sectional | Adults, male | Adult METH and heroin exposure | Healthy controls | rs-fMRI | 86.67% | 13 | 15 |

| | | | | | | | | |
|---|---|---|---|---|---|---|---|---|
| Lu et al. 2009(Lu et al., 2009) | Cross-sectional | Children, both | Prenatal METH exposure | Healthy controls | task-fMRI | 78.57% | 11 | 14 |
| MacDuffie et al. 2018(MacDuffie et al., 2018) | Cross-sectional | Adults, both | MAA | Healthy controls | Structural | 100.00% | 15 | 15 |
| Malcolm et al. 2016(Malcolm et al., 2016) | Cross-sectional | Adults, male | MAA | Healthy controls | task-fMRI | 93.75% | 15 | 16 |
| Malina et al. 2021(Malina et al., 2021) | RCT (crossover) | Adults, both | Healthy volunteers who received METH | Healthy volunteers who received placebo | rs-fMRI | 90.91% | 10 | 11 |
| Mansoory et al. 2020(Mansoory et al., 2020) | Cross-sectional | Adults, male | MAA | Healthy controls | rs-fMRI | 66.67% | 10 | 15 |

| | | | | | | | | |
|---|---|---|---|---|---|---|---|---|
| May et al. 2013(May et al., 2013) | Cross-sectional | Adults, both | MAA | Healthy controls | task-fMRI | 93.75% | 15 | 16 |
| Monterosso et al. 2007(Monterosso et al., 2007) | Cross-sectional | Adults, both | MAA | Healthy controls | task-fMRI | 86.67% | 13 | 15 |
| Morales et al. 2012(Morales et al., 2012) | Cohort | Adults, both | MAA | Healthy controls | Structural | 100.00% | 17 | 17 |
| Morales et al. 2015(Morales et al., 2015) | Cross-sectional | Adults, both | MAA | Healthy controls | Structural | 100.00% | 15 | 15 |
| Nakama et al. 2011(Nakama et al., 2011) | Cross-sectional | Adults, both | MAA | Healthy controls | Structural | 100.00% | 15 | 15 |

| | | | | | | | | |
|---|---|---|---|---|---|---|---|---|
| Nestor et al. 2011(Nestor et al., 2011) | Cross-sectional | Adults, both | MAA | Healthy controls | task-fMRI | 86.67% | 13 | 15 |
| Nie et al. 2020(Nie et al., 2020) | Cross-sectional | Adults, both | MAA | Healthy controls | Structural | 100.00% | 16 | 16 |
| Nie et al. 2021(Nie et al., 2021) | Cross-sectional | Adults, both | MAA | Healthy controls | Structural | 100.00% | 15 | 15 |
| Okita et al. 2018(Okita et al., 2018) | Cross-sectional | Adults, both | MAA | Healthy controls | Structural | 100.00% | 16 | 16 |
| Orikabe et al. 2011(Orikabe et al., 2011) | Cross-sectional | Adults, both | METH-associated psychosis | Healthy controls | Structural | 80.00% | 12 | 15 |

| | | | | | | | | |
|---|---|---|---|---|---|---|---|---|
| Pang et al. 2021(Pang and Peng, 2021) | Cross-sectional | Adults, both | MAA | Healthy controls | MRS | 86.67% | 13 | 15 |
| Paulus et al. 2002(Paulus et al., 2002) | Cross-sectional | Adults, male | MAA | Healthy controls | task-fMRI | 81.25% | 13 | 16 |
| Paulus et al. 2003(Paulus et al., 2003) | Cross-sectional | Adults, both | MAA | Healthy controls | task-fMRI | 81.25% | 13 | 16 |
| Paulus et al. 2005(Paulus et al., 2005) | Cohort | Adults, male | MAA who remained abstinent at 1-year | MAA who relapsed at 1-year | task-fMRI | 52.94% | 9 | 17 |
| Payer et al. 2008(Payer et al., 2008) | Cross-sectional | Adults, both | MAA | Healthy controls | task-fMRI | 80.00% | 12 | 15 |

| | | | | | | | | |
|---|---|---|---|---|---|---|---|---|
| Payer et al. 2011(Payer et al., 2011) | Cross-sectional | Adults, both | MAA | Healthy controls | task-fMRI | 93.33% | 14 | 15 |
| Payer et al. 2012(Payer et al., 2012) | Cross-sectional | Adults, both | MAA | Healthy controls | task-fMRI | 87.50% | 14 | 16 |
| Qi et al. 2020(Qi et al., 2020) | Cohort | Adults, both | MAA with baseline cravings | MAA without baseline cravings | Structural | 94.12% | 16 | 17 |
| Roos et al. 2014(Roos et al., 2014) | Cross-sectional | Children, both | Prenatal METH exposure | Healthy controls | Structural | 78.57% | 11 | 14 |
| Roos et al. 2015(Roos et al., 2015) | Cross-sectional | Children, both | Prenatal METH exposure | Healthy controls | dMRI | 78.57% | 11 | 14 |

| | | | | | | | | |
|---|---|---|---|---|---|---|---|---|
| Roussotte et al. 2011(Roussotte et al., 2011) | Cross-sectional | Children, both | Prenatal METH exposure | Healthy controls | task-fMRI | 85.71% | 12 | 14 |
| Roussotte et al. 2012(Roussotte et al., 2012) | Cross-sectional | Children, both | Prenatal METH exposure | Healthy controls | task-fMRI | 78.57% | 11 | 14 |
| Ruan et al. 2018(Ruan et al., 2018) | Cohort | Adults, both | MAA | Healthy controls | Structural | 100.00% | 18 | 18 |
| Salo et al. 2009(Salo et al., 2009a) | Cross-sectional | Adults, both | MAA | Healthy controls | dMRI | 100.00% | 15 | 15 |
| Salo et al. 2009(Salo et al., 2009b) | Cross-sectional | Adults, both | MAA | Healthy controls | task-fMRI | 86.67% | 13 | 15 |

| | | | | | | | | |
|---|---|---|---|---|---|---|---|---|
| Salo et al. 2011(Salo et al., 2011) | Cross-sectional | Adults, both | MAA | Healthy controls | MRS | 100.00% | 16 | 16 |
| Salo et al. 2013(Salo et al., 2013) | Cross-sectional | Adults, both | MAA | Healthy controls | task-fMRI | 86.67% | 13 | 15 |
| Schwartz et al. 2010(Schwartz et al., 2010) | Cross-sectional | Adults, both | MAA | Healthy controls | Structural | 100.00% | 15 | 15 |
| Shahbabaie et al. 2018(Shahbabaie et al., 2018) | RCT (crossover) | Adults, male | MAA | N/A | rs-fMRI | 66.67% | 12 | 18 |
| Smith et al. 2001(Smith et al., 2001) | Cross-sectional | Children, sex NR | Prenatal METH exposure | Healthy controls | Structural and MRS | 85.71% | 12 | 14 |

| | | | | | | | | |
|---|---|---|---|---|---|---|---|---|
| Sowell et al. 2010(Sowell et al., 2010) | Cross-sectional | Children, both | Prenatal METH exposure | Healthy controls | Structural | 92.86% | 13 | 14 |
| Stewart et al. 2014(Stewart et al., 2014a) | Cohort | Adults, both | MAA who remained abstinent at 1-year | MAA who relapsed at 1-year | task-fMRI | 94.12% | 16 | 17 |
| Stewart et al. 2014(Stewart et al., 2014b) | Cross-sectional | Adults, both | MAA | Healthy controls | task-fMRI | 100.00% | 15 | 15 |
| Su et al. 2020(Su et al., 2020) | RCT | Adults, both | MAA who received rTMS | MAA who received sham rTMS | rs-fMRI | 87.50% | 14 | 16 |
| Tabibnia et al. 2011(Tabibnia et al., 2011) | Cross-sectional | Adults, both | MAA | Healthy controls | task-fMRI | 66.67% | 10 | 15 |

| | | | | | | | | |
|---|---|---|---|---|---|---|---|---|
| Thompson et al. 2004(Thompson et al., 2004) | Cross-sectional | Adults, both | MAA | Healthy controls | Structural | 100.00% | 15 | 15 |
| Tobias et al. 2010(Tobias et al., 2010) | Cross-sectional | Adults, both | MAA | Healthy controls | dMRI | 100.00% | 15 | 15 |
| Uhlmann et al. 2016(Uhlmann et al., 2016a) | Cross-sectional | Adults, both | METH-associated psychosis | Healthy controls | Structural | 100.00% | 15 | 15 |
| Uhlmann et al. 2016(Uhlmann et al., 2016b) | Cross-sectional | Adults, both | METH-associated psychosis | Healthy controls | dMRI | 100.00% | 15 | 15 |
| van Hedger et al. 2018(Van Hedger et al., 2018) | Single-arm intervention trial | Adults, both | Healthy volunteers who received METH | N/A | task-fMRI | 76.92% | 10 | 13 |

| | | | | | | | | |
|---|---|---|---|---|---|---|---|---|
| van Hedger et al. 2019(Van Hedger et al., 2019) | RCT (crossover) | Adults, both | Healthy volunteers who received METH | N/A | task-fMRI | 90.91% | 10 | 11 |
| Vollm et al. 2004(Vollm et al., 2004) | Single-arm interventional trial | Adults, both | Healthy volunteers who received METH | N/A | rs-fMRI | 54.55% | 6 | 11 |
| Vuletic et al. 2018(Vuletic et al., 2018) | Cross-sectional | Adults, both | MAA | Healthy controls | pCASL | 86.67% | 13 | 15 |
| Warton et al. 2018(Warton et al., 2018a) | Cross-sectional | Infants, both | Prenatal METH exposure | Healthy controls | Structural | 93.33% | 14 | 15 |
| Warton et al. 2018(Warton et al., 2018b) | Cross-sectional | Infants, both | Prenatal METH exposure | Healthy controls | dMRI | 86.67% | 13 | 15 |

| | | | | | | | | |
|---|---|---|---|---|---|---|---|---|
| Warton et al. 2020(Warton et al., 2020) | Cross-sectional | Infants, both | Prenatal METH exposure | Healthy controls | dMRI | 93.33% | 14 | 15 |
| Weafer et al. 2020(Weafer et al., 2020) | RCT (crossover) | Adults, both | Healthy volunteers who received METH | N/A | rs-fMRI | 90.91% | 10 | 11 |
| Wu et al. 2018(Wu et al., 2018) | Cross-sectional | Adults, both | MAA | Healthy controls | MRS and Structural | 93.75% | 15 | 16 |
| Yan et al. 2021(Yan et al., 2021) | Cohort | Adults, both | MAA | Healthy controls | rs-fMRI | 94.12% | 16 | 17 |
| Yang et al. 2021(Yang, R. et al., 2021) | Cross-sectional | Adults, male | MAA | Healthy controls | Structural and rs-fMRI | 100.00% | 15 | 16 |

| | | | | | | | | |
|---|---|---|---|---|---|---|---|---|
| Yang et al. 2021(Yang, M. et al., 2021) | Cross-sectional | Adults, both | METH-associated psychosis | Healthy controls | rs-fMRI | 93.75% | 15 | 15 |
| Yin et al. 2012(Yin et al., 2012) | Cross-sectional | Adults, both | MAA | Healthy controls | task-fMRI | 53.33% | 8 | 15 |
| Yu et al. 2020(Yu et al., 2020) | Cross-sectional | Adults, both | MAA | Healthy controls | rs-fMRI | 46.67% | 7 | 15 |
| Zhang et al. 2018(Zhang, S. et al., 2018) | Cohort | Adults, both | MAA | Healthy controls | rs-fMRI | 81.25% | 13 | 16 |
| Zhang et al. 2018(Zhang, Z. et al., 2018) | Cross-sectional | Adults, male | MAA | Healthy controls | Structural | 100.00% | 18 | 18 |
| Zhuang et al. 2016(Zhuang et al., 2016) | Cohort | Adults, male | MAA | Healthy controls | dMRI | 100.00% | 16 | 16 |

*Adult METH user includes a history of current or historical METH abuse, dependence, or use disorder, as diagnosed by the Diagnostic and Statistical Manual of Mental Disorders 4th Edition, Diagnostic and Statistical Manual of Mental Disorders 5th Edition, or International Classification of Diseases 10$^{th}$ edition diagnostic* *codes. Healthy controls defined as participants without adult or prenatal METH exposure. dMRI: Diffusion magnetic resonance imaging. MAA: Adult METH user. METH: Methamphetamine. MRI: Magnetic resonance imaging. N/A: Not applicable. rs-fMRI: Resting-state functional magnetic resonance imaging. Task-fMRI: Task-based functional magnetic resonance imaging.*
